# Investigating the structure of Schizotypy through the “Multidimensional Schizotypy Scale” and “Oxford-Liverpool Inventory”: an Exploratory network analysis approach in the healthy population

**DOI:** 10.1101/2024.07.13.24310316

**Authors:** Pierfrancesco Sarti, Werner Surbeck, Giacomo Cecere, Noemi Dannecker, Rahel Horisberger, Nils Kallen, Wolfgang Omlor, Anna Steiner, Dario Palpella, Nicolas Langer, Johanna M.C. Blom, Philipp Homan

## Abstract

**Background:** Schizophrenia spectrum disorders and schizotypy share traits across positive, negative, and disorganized domains. Instruments like MSS and O-LIFE provide insights into these dimensions. Despite challenges posed by cultural variations and measurement methodologies, these instruments offer nuanced perspectives on the spectrum of schizotypy, spanning from its expression as a personality trait to its potential implications in clinical settings.

**Methods:** Through an Exploratory Graph Analysis (EGA) applied to a sample of 1059 healthy subjects, we compare the resulting networks with the original factorial structure of the two questionnaires and explore how each one conceptualizes schizotypy. Comparing both models quantitatively and qualitatively these models, we seek to elucidate the unique insights provided by each instrument regarding the spectrum of schizotypy as a personality trait.

**Results:** EGA analyses unveiled a three-dimensional structure plus an additional one consisting of items related to the concept of “Disconnection”. Confirmatory factor analysis showed this four-dimensional model outperformed others. High reliability and strong factor saturation were observed for all four factors.

O-LIFE’s EGA revealed a complex structure, refined to four factors removing items with low fitting indices from the scale. Confirmatory analysis validated a final model with robust reliability and well-defined factor structures across the new “Cognitive and Behavioural Disorganization”, “Introversion”, “Unusual Experiences”, and “Environmental Pressure” factors.

**Conclusions:** This study improves the understanding of schizotypy by proposing new MSS and O-LIFE factor structures via EGA. Both scales uniquely contribute to schizotypy assessment in healthy populations, warranting further research to validate and refine these new domains across diverse populations.

## 1. Introduction

Schizophrenia spectrum disorders (SSD) are among the most severe mental illnesses globally, characterized by a high burden of disease and a significant negative effect on the individual’s occupational, educational, social, and economic life (Crespo-Facorro et al., 2021). These disorders are broadly defined by altered and abnormal mental states over certain periods of time, characterised by symptom clusters including delusions and hallucinations, disorganised thinking and motor behaviour, negative symptoms such as apathy, anhedonia and blunted affect. The presentation of these symptoms varies from person to person and often co-occurs with other disorders, hindering a prompt and precise diagnosis (Tandon et al., 2013; Schultze-Lutter et al., 2019; Sarraf et al., 2022).

The notion of schizotypy, distinct from clinical SSD, has garnered significant attention in psychiatric research. Both schizotypy and SSD share traits and symptoms across the same domains — positive, negative, and disorganized. Unlike the episodic nature of SSD symptoms, these dimensions represent enduring characteristics in individuals’ personal lives (Grant et al., 2018; Kwapil, Gross, Burgin, et al., 2018; Nelson et al., 2013).

Due to its combination of behavioural personality traits that can manifest also as pathological, it has been proposed as a potential endophenotype or “schizophrenia liability” (Grant, 2015; Lenzenweger, 2006, 2015) and has provided valuable insights into various schizophrenia-spectrum disorders and their underlying aetiologies (Barrantes-Vidal et al., 2015). By studying individuals expressing high schizotypy without clinical SSD, researchers have been able to identify both risk and resilience factors. This research has contributed to an enhanced understanding of SSD and the development of more effective intervention strategies(Barrantes-Vidal et al., 2015; Grant & Hennig, 2020).

However, debates surrounding the nature of Schizotypy persist; some argue for a taxonomic model that sees schizotypy as one end of the psychosis spectrum and therefore inherently pathological(Morton et al., 2017), while others propose a broader dimensional model that suggests that schizotypy is not necessarily associated with pathology (Grant et al., 2018). Claridge’s fully dimensional model, for instance, proposes that schizotypy traits can be adaptive or maladaptive, implying a continuum between schizotypy as a personality trait having no psychopathological value up to a pathological manifestation of the symptoms (Mohr & Claridge, 2015). This view has paved the way for the inclusion of non-clinical populations in schizotypy research, shedding light on the dynamic nature of these traits(Debbané & Barrantes-Vidal, 2015).

Despite these challenges and differences, schizotypy research continues to provide invaluable insights into the complex interplay between personality and psychopathology. In addition, the current clinical classification systems of DSM-5 and ICD-11 have some major limitations because they fail to integrate new models of psychopathology with advanced research methodologies (Kendell & Jablensky, 2003; Wright et al., 2013; Cohen et al., 2018; Hengartner & Lehmann, 2017; Kotov et al., 2018) leaving the definition of these disorders obsolete or often misdiagnosed as schizophrenia(Schultze-Lutter et al., 2019).

To validly measure schizotypy as a stable, non-pathologic personality structure, psychometric instruments are preferred; in particular self-reports that are cost-effective and non-invasive(Barrantes-Vidal et al., 2015; O. J. Mason, 2015, p. 2).

However, the theoretical framework behind these instruments significantly influences their content and the validity of their assessments (Fonseca-Pedrero et al., 2021; Grant et al., 2018; Kwapil, Gross, Burgin, et al., 2018): some instruments target clinical populations with a focus on pathological aspects, while others aim to measure schizotypy as a broader, dimensional construct.

For instance, the Wisconsin Schizotypy Scales, based on Meehl’s taxonomic framework, consists of four scales (J. P. Chapman et al., 1995; L. J. Chapman et al., 1976, 1978, 1980; Eckblad & Chapman, 1983). While it targets clinical populations, it may not fully cover the disorganized dimension or negative aspects schizotypy, such as alogia or avolition.

Similarly, the Schizotypal Personality Questionnaire developed by Adrian Raine (1991) (Raine, 1991) and measures aspects of SPD through nine subscales but inadequately addresses the disorganized dimension.

Two further scales, the Multidimensional Schizotypy scale (MSS) and the Oxford-Liverpool inventory for feelings and experiences (O-LIFE), offer comprehensive coverage of the three established dimensions of Schizotypy and demonstrate robust psychometric properties (Grant et al., 2013; Oezgen & Grant, 2018; Christensen et al., 2019; Kemp et al., 2020; Polner et al., 2021). These scales offer researchers comprehensive tools to explore the nuanced interplay between personality traits and psychopathology.

The MSS, aligned with the taxonomic view of schizotypy, explores experiences akin to symptoms of SSD but in milder form (Kwapil, Gross, Silvia, et al., 2018). In contrast, the O-LIFE adopts a personality-oriented approach, where high manifestations of Schizotypy extend beyond the clinical spectrum (O. Mason & Claridge, 2006).

These instruments thus provide diverse viewpoints on schizotypy, offering new insights into its dimensional structure and potentially shedding light on SSD (Grant et al., 2018).

Given that cultural influences can significantly impact schizotypal traits and experiences, it is crucial to assess the structural equivalence of these measures across diverse populations. This enhances our understanding of the universal aspects of Schizotypy (Fonseca-Pedrero et al., 2014; Cohen et al., 2015; Fonseca-Pedrero et al., 2015; Barron, 2017). Authors emphasize the importance of replicating the factorial structure of these instruments across different cultural and linguistic contexts, using both clinical and non-clinical samples (Kwapil, Gross, Silvia, et al., 2018; Kwapil, Gross, Burgin, et al., 2018). Issues related to translation may alter the interpretation of questionnaire items, potentially affecting response patterns and the reliability of the instrument (Grant et al., 2013). Moreover, as psychopathologies and personality are non-crystallised constructs that evolve with the passage of time and the ability to adapt to an increasingly complex environment, there is a constant need to incorporate new statistical methodologies that better capture this fluidity (Achterhof et al., 2022; Onchev, 2021).

Network psychometrics is a swiftly advancing field applied to various psychopathological constructs, including Schizotypy (Christensen et al., 2018; Fonseca-Pedrero et al., 2018). This approach views psychopathological constructs as complex systems arising from interactions between their constituent elements (symptoms, internal and external factors) (Borsboom & Cramer, 2013; Schmittmann et al., 2013).

The network theory of psychopathology proposes that symptoms can reinforce each other, can be influenced by factors such as biological, environmental, and social mechanisms, and can lead to self-sustaining states that are difficult to modify (Borsboom, 2017). This theory supports current views on Schizotypy as a latent liability for schizophrenia spectrum disorders, where interactions with several factors may facilitate the disorder transition (A. Isvoranu et al., 2016; A.-M. Isvoranu et al., 2017; Lenzenweger, 2018).

In the field Network analysis and modelling, the Exploratory Graph Analysis (EGA) offers a novel approach to uncovering dimensions within psychometric instruments (van Borkulo et al., 2014; Borsboom, 2017; H. F. Golino & Epskamp, 2017). EGA employs a Gaussian Graphical Model, computed using the graphical least absolute shrinkage and selection operator (glasso) (Friedman et al., 2008; Strobl et al., 2012; Epskamp et al., 2018). The walktrap community detection algorithm then identifies network dimensions by performing “random walks” from one node to another, forming communities based on densely connected edges (Pons & Latapy, 2005). Because of the implementation of this algorithm, EGA’s dimensions are determined without researcher bias, offering an advantage over other exploratory dimension reduction methods by providing immediately interpretable content and dimensions without needing to interpret component loadings. EGA has shown comparable or superior accuracy in identifying dimensions compared to other methods such as principal component analysis and factor analysis (H. Golino et al., 2020; H. F. Golino & Demetriou, 2017; H. F. Golino & Epskamp, 2017) and has effectively replicated factor analytic findings and discovered new construct dimensions(Bell & O’Driscoll, 2018) .

To our knowledge, no study has yet conducted an Exploratory Graph Analysis (EGA) comparing the factorial structures of two schizotypal instruments within the same sample, contrasting their similarities and differences with the original models. This represents critical area for further research.

Therefore, the main objective of this study is to validate the factorial structures of the Multidimensional Schizotypy Scale (MSS) and the Oxford-Liverpool Inventory of Feelings and Experiences (O-LIFE) in a large German-speaking non-clinical sample. We aim to compare this structure with those identified by Exploratory Factor Analysis (EFA).

Furthermore, through interpreting these resulting models, our study will facilitate a qualitative comparison of how each questionnaire conceptualizes schizotypy and the unique insights each instrument offers.

## 2. Methods

### 2.1 Participants

The participants were recruited via an online survey aimed at identifying potential candidates for a comprehensive study at the University Hospital of Psychiatry Zurich (PUK). For further details on the study, please visit the VELAS website (https://homanlab.github.io/velas/). The survey included questions about demographics, MRI safety, the German versions of the MSS and O-LIFE, and the German version of the Childhood Trauma Questionnaire(Wingenfeld et al., 2010; *[The German version of the Childhood Trauma Questionnaire (CTQ): preliminary psychometric properties] - PubMed*, s.d.). Completing the survey took approximately 20 to 30 minutes. The current study analysis focused exclusively on the MSS and O-LIFE results.

Due to the main study’s specific research requirements, participants had to meet several criteria to be considered eligible:

- they needed to be healthy without any psychiatric or neurological conditions,
- aged between 18 and 40 years,
- right-handed,
- proficient in German.

The online survey was widely distributed across various platforms, including Swiss university and college marketplaces, public forums like RonOrp, the “PSYCH-Pool” of the University of Zurich (server with registered persons who want to participate in studies), the psychology students’ association (FAPS) mailing list, tweets from the research team, and direct outreach to various educational institutions. This approach aimed to attract individuals with diverse educational and social backgrounds.

Participants who were selected and participated in the main study received a compensation of 150 Swiss francs, while those not selected after completing the survey received no compensation.

A total of 1059 participants completed the entire questionnaire between April 2021 and March 2024. The questionnaire was converted into an online format using the Enterprise Feedback Suite and since the demographic, MSS, and O-LIFE items were programmed as forced-choice questions, no missing values were present in the dataset. The consent of the healthy subjects to the data processing was provided at the beginning of the compilation of the test battery. They had to read the protocol and subsequently tick the items of understanding the study and analysing the collected data.

Without this, the questionnaires could not be accessed.

Participants were instructed before each instrument (MSS or O-LIFE) that the following statements or questions cover a broad range of attitudes, thoughts, experiences, preferences, and beliefs, and they should respond as honestly as possible. They were also informed that there were no right or wrong answers and that they should answer in a way that best reflects themselves.

### 2.2 Materials

The MSS and O-LIFE were used in their German versions, translated by bilingual researchers from the International Consortium of Schizotypy Research (ICSR) (https://srconsortium.org), and have been utilized in other studies(Grant et al., 2013; Nenadić et al., 2021).

The MSS consists of 77 Yes-or-No items, divided into subscales: 26 items in both the “Negative” and “Positive” subscales, and 25 items in the “Disorganised” subscale. Previous studies with larger samples have shown the internal consistency of these subscales to be good to excellent, with binary adjusted alpha coefficients ranging from 0.87 to 0.95 in English-speaking samples, and from 0.78 to 0.89 in German-speaking samples (Kwapil, Gross, Burgin, et al., 2018; Kwapil, Gross, Silvia, et al., 2018; Nenadić et al., 2021).

The O-LIFE comprises 104 Yes-or-No items, with 30 items in the “Unusual Experiences” (UnEx) subscale describing perceptual aberrations, magical thinking, and hallucinations, 27 in the “Introvertive Anhedonia” (IntA), a subscale that describe a lack of enjoyment from social and physical sources of pleasure, as well as avoidance of intimacy, 24 in the “Cognitive Dysregulation” (CogDis) subscale that taps aspects of poor attention, concentration, social anxiety and poor decision-making, and 23 in the “Impulsive nonconformity” (ImpNon) subscale which contains items describing anti-social, eccentric and impulsive forms of behaviour, often suggesting a lack of self-control .

Alpha coefficients for these subscales were found to be between 0.77 and 0.89 in studies by Mason and Claridge (2006) (O. Mason & Claridge, 2006), and between 0.68 and 0.88 in a German-speaking sample by Grant et al. (2013)(Grant et al., 2013).

Notably, the lowest alpha coefficient was in the ImpNon subscale, whereas the other three subscales (UnEx, IntA, CogDis) had coefficients above 0.8, indicating good to excellent internal consistency.

### 2.3 Statistical Analysis

The data were analysed quantitatively to extrapolate descriptive information from the sample using mean, standard deviation and percentages.

In order to choose the best correlation method for constructing the matrices then used for the Exploratory Graph Analysis, we conducted the Jenrich’s Test (Atiany & Sharif, 2018) comparing the Fiml (Full information Maximum Likelihood) correlations used in the article by Christensen et al. 2019 (Christensen et al., 2019), Spearman’s correlations, since these of the MSS and O-LIFE are categorical variables and, finally, the Phi (mean square contingency coefficient) coefficients used in the specific case in which the variables are categorical and dichotomous. The latter is interpreted as a Pearson’s ‘p’.

We utilised the EGA package(H. F. Golino & Epskamp, 2017) in R to perform the Exploratory Graph Analysis, employing first the qgraph to apply the glasso algorithm (Epskamp et al., 2012) and then igraph(Csardi & Nepusz, 2005) to compute the walktrap clustering methods (Pons & Latapy, 2005). The glasso method was estimated using a penalised maximum likelihood solution based on the Extended Bayesian Information Criterion – EBIC (Chen & Chen, 2008; Epskamp et al., 2018).

The representation of the networks was done using the ‘qgraph’ package and analyses were also carried out to calculate the centrality indices. Betweenness centrality was used as a value for the size of the individual nodes(Bringmann et al., 2019).

If the number of factors found was excessively large or if issues emerged in the correlations of individual items from the representation, an Exploratory Factor Analysis was conducted. First, a Kaiser-Meyer-Olkin test (Kaiser, 1974) and Bartlett’s test of sphericity(Bartlett, 1950) were performed. Then, if items had inadequate values of Measure of Sample Adequacy (MSA), the EGA would be conducted again without those items.

A Confirmatory Factor Analysis was then conducted to compare the resulting models with the original ones. The correlation matrices of the individual factors of each model were estimated using the WLSMV (diagonally weighted least squares estimator)(Muthén, 1984). Each model was fitted using the ‘lavaan’ package(Rosseel, 2012) in R.

The fitting of the models obtained were compared to each other using the Satorra-Bentler chi-square test(Satorra & Bentler, 2010) and qualitatively using: comparative fit index (CFI), standardised root mean residual (SRMR), root mean square error of approximation (RMSEA) and Tucker-Lewis Index (TLI).

Based on the results, the best model was chosen, and the individual factors were taken and reliability measures such as McDonalds’ Omega(McDonald, 1999) and Cronbach’s Alpha(Cronbach, 1951) were calculated.

### 2.4 R code and materials sharing

All the R code with results and graph is available as Supplementary Material.

## 3. Results

The demographic information concerning the sample of 1059 healthy subjects who participated in the study is in Table 1. Frequencies, means, standard deviations and percentages are indicated.

**Table 1:** demographic variables of the population. The breakdown of the variable Education was made according to the Swiss educational system.

| Demographic table | Total |  | Males |  | Females |  | Others |  |
| --- | --- | --- | --- | --- | --- | --- | --- | --- |
|  | n° | % | n° | % | n° | % | n° | % |
| **N sample** | 1059 | 100 | 300 | 28.33 | 757 | 71.48 | 2 | 0.19 |
| **Age: mean and sd** | 24.78 ± 4.48 |  | 25.52 ± 4.59 |  | 24.46 ± 4.37 |  | 25.44 ± 4.83 |  |
| **Education** |  |  |  |  |  |  |  |  |
| No grade | 3 | 0.28 | 1 | 0.33 | 2 | 2.80 | 0 | 0.00 |
| Secondary School | 40 | 3.78 | 13 | 4.33 | 26 | 36.37 | 1 | 50.00 |
| Apprenticeship | 224 | 21.15 | 71 | 23.67 | 153 | 214.05 | 0 | 0.00 |
| Technical School | 20 | 1.89 | 5 | 1.67 | 15 | 20.98 | 0 | 0.00 |
| Matura | 362 | 34.18 | 99 | 33.00 | 263 | 367.94 | 0 | 0.00 |
| Higher Technical Institute | 43 | 4.06 | 9 | 3.00 | 34 | 47.57 | 0 | 0.00 |
| University of applied sciences | 74 | 6.99 | 28 | 9.33 | 46 | 64.35 | 0 | 0.00 |
| University | 289 | 27.29 | 72 | 24.00 | 216 | 302.18 | 1 | 50.00 |
| Other | 4 | 0.38 | 2 | 0.67 | 2 | 2.80 | 0 | 0.00 |

Information on it can be found at https://www.berufsberatung.ch/dyn/show/2800. We also specify that some categories include more than one type of studies: secondary school (type A, B, C); Apprenticeship (Federal Vocational Certificate (EBA), Federal Certificate of Competence (EFZ) with/without matura, HPF); Technical school (with/without Matura); Matura (grammar school, adult matura, passerelle); both types of University include Bachelor, Master, Promotion).

For both questionnaires, the correlation matrices used for the EGA contained Spearman correlations. Jennrich’s tests were not significant, so there no difference can be assumed between the correlation matrices using Phi, Spearman or Fiml. Spearman was preferred as the resulting matrix provided one more significant digit.

### 3.1 MSS model

EGA analyses revealed a three-dimensional structure in the sample reflecting the division of positive, negative and disorganisation symptoms. The positive and disorganisation dimensions have a higher density of connections between them than the negative domain. Of all the items, eight of them were wrongly included in the positive domain by the EGA: seven of them were part of the disorganisation factor and one of the negative factor. The specific items are listed below:

- NEG1: I have always preferred to be disconnected from the world
- DIS06: When people ask me a question, I often do not understand what they are asking
- DIS30: I often find that when I talk to people, I do not make any sense to them
- DIS42: I often feel so disconnected from the world that I am not able to do things
- DIS45: I am very often confused about what is going on around me
- DIS51: People find my conversations to be confusing or hard to follow
- DIS69: I have trouble following conversations with others
- DIS75: It is usually easy for me to follow conversations

All these items share a common theme related to “disconnection” from the world or difficulties in understanding or being understood by others. In Figure 1, these items are visually separated from the others, forming a distinct fourth domain we named “Disconnection”. The size of each node in the figure also reflects its “Betweenness” centrality value. This index indicates the importance of these nodes as “gatekeepers” within the network, as they frequently lie on the shortest paths between other nodes.

Subsequently, a CFA analysis was conducted to evaluate the fit of several models, the original scale model, the three-factor model identified by the EGA, and a third model that includes a separate factor for the items classified as misfits. The indices of interest are shown below:

- Original model: Chi-square: 4899.17; df: 2846; SRMR: 0.123; CFI: 0.979; RMSEA: 0.026; TLI: 0.978
- EGA model with 3 dimensions: Chi-square: 5082.16; df: 2846; SRMR: 0.124; CFI: 0.977; RMSEA: 0.027; TLI: 0.976
- EGA model with 4 dimensions: Chi-square: 4680.91; df: 2843; SRMR: 0.120; CFI: 0.981; RMSEA: 0.025; TLI: 0.981

The Satorra-Bentler tests performed between the original, three-dimensional EGA model (with misclassified items) and the four-dimensional EGA model were not significant, indicating that there is no difference between the three results.

However, the four-dimensional model obtains better fitting values than the other two; furthermore: the Negative factor exhibits high internal consistency (Alpha = 0.85) and strong general factor saturation (Omega Total = 0.87). Similar to the negative factor, the Positive factor shows high reliability (Alpha = 0.85) and strong general factor saturation (Omega Total = 0.87).

The Disorganised factor demonstrates excellent reliability (Alpha = 0.92) and a strong general factor saturation (Omega Total = 0.93).

Finally. the Disconnected factor exhibits good reliability (Alpha = 0.74), a lower general factor saturation (Omega Total = 0.81) than the others but still considered good as a substantial portion of the variance is due to common factors, suggesting good internal consistency.

**Figure 1:**
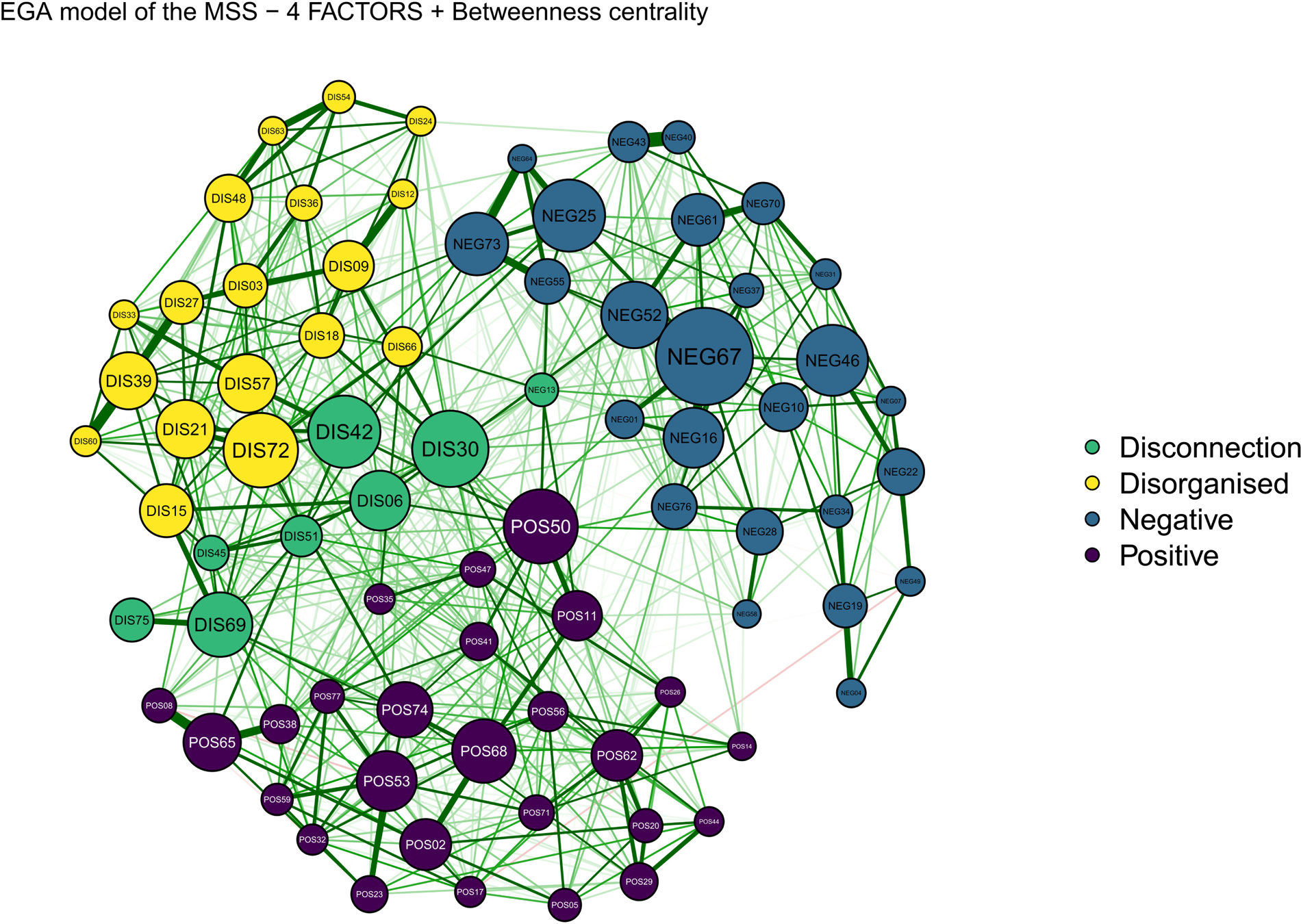
MSS network representing the three factors identified by the EGA and the green group/factor “Disconnection”, called such following the analysis of the individual items. The size of the nodes reflects the Betweenness Centrality value. The green edges represent positive Spearman correlations.

### 3.2 O-LIFE model

The first EGA analysis on the O-LIFE questionnaire showed a 13-factor structure whose representation, using the four subscales of the test (Figure 2), revealed multiple nodes that were either uncorrelated or connected two by two. Moreover, the factor ‘Impulsive Nonconformity’ is the one that least maintains a unitary and separate structure, spreading out among the others.

For possible confounding effects and to reduce the number of factors, an Exploratory Factor Analysis was conducted.

The Kaiser-Meyer-Olkin test, although giving an Overall MSA of 0.9, revealed that seven items had values below 0.7 and ten others between 0.7 and 0.8.

The Parallel Analysis performed with tetrachoric correlations identified four main factors. looking at the individual loadings several items had high positive and/or negative correlations with more than one factor at the same time.

**Figure 2:**
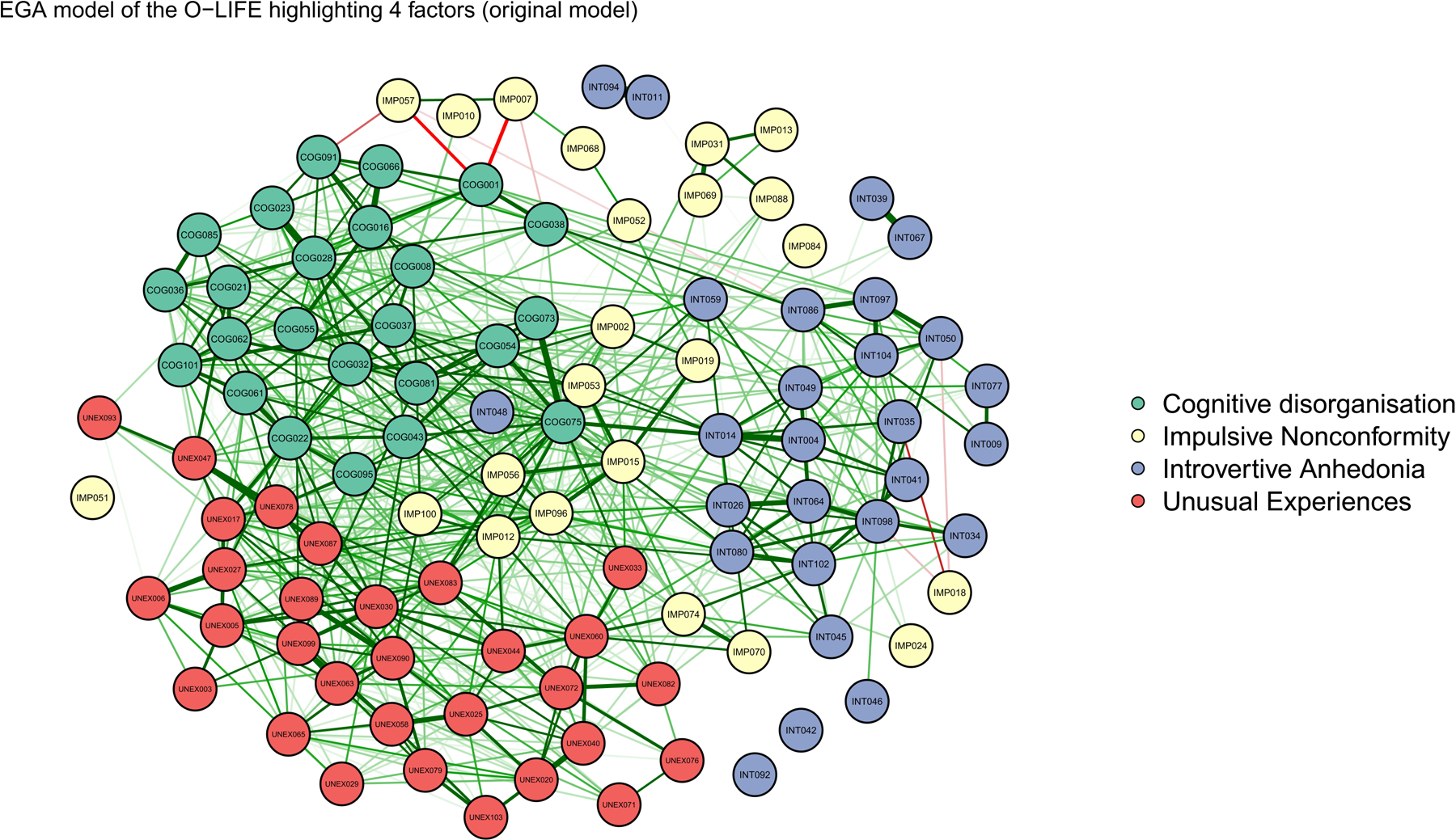
O-LIFE network identified by the EGA separating the nodes with the four subscales proposed by the model of Mason and Claridge (2006).

Thus, items with a Measure of Sampling Adequacy of less than 0.7 (middling) were first removed from the dataset, resulting in a reduction in the number of factors from 13 to 9, and then also those with a value between 0.7 and 0.8 (middling), resulting in a total of 4 factors. The resulting network can be observed in Figure 3. Confirmatory Factor Analysis was performed for the three models: original model with all items, model obtained with Exploratory Factor Analysis and the 4-dimensional model with 87 items (after removing those with low MSA).

The results of the fitting of each model are shown in Table 2. The model obtained after removing 17 items (final model) from the questionnaire turns out to be the one with better indices. Furthermore, the Satorra-Bentler test was significant when crossing the final model with the other two in both cases:

- EGA model vs. Original: Chi-squared difference = 2637; Df difference: 1496; p = 2.2x10−16
- EGA model vs. EFA model: Chi-squared difference = 2454; Df difference: 1598; p = 2.2x10−16

After analysing the new item groups, two remained stable and similar to the original model (Introvertive Anhedonia, now Introversion, and Unusual Experiences) while the other two were renamed to (Cognitive and Behavioural Disorganization and Environmental Pressure).

The reliability analysis for the four variables — Cognitive and Behavioural Disorganization, Introversion, Unusual Experiences, and Environmental Pressure — indicates robust internal consistency and well-defined factor structures. The first one shows high reliability with an alpha of 0.88 and an omega total of 0.89, alongside significant contributions from both general and group factors. Introversion is also reliable, evidenced by an alpha of 0.82 and an omega total of 0.83, with the general factor explaining a substantial portion of the variance. Unusual Experiences has strong reliability, with an alpha of 0.86 and an omega total of 0.87, indicating that both the general factor and group factors are important. Environmental Pressure, while slightly lower in reliability, still demonstrates good consistency with an alpha of 0.70 and an omega total of 0.75, dominated by the general factor. The model fits, as indicated by RMSEA values below 0.05 for all variables, suggest that the models are appropriate and explain a considerable amount of the variance. Correlations of scores with factors and multiple R square values are high for the general factor, confirming the adequacy of the factor score estimates. Despite some negative values in the minimum correlations for group factors, the overall reliability and factor structures are solid, ensuring the scales are reliable and effectively capture the intended constructs.

**Figure 3:**
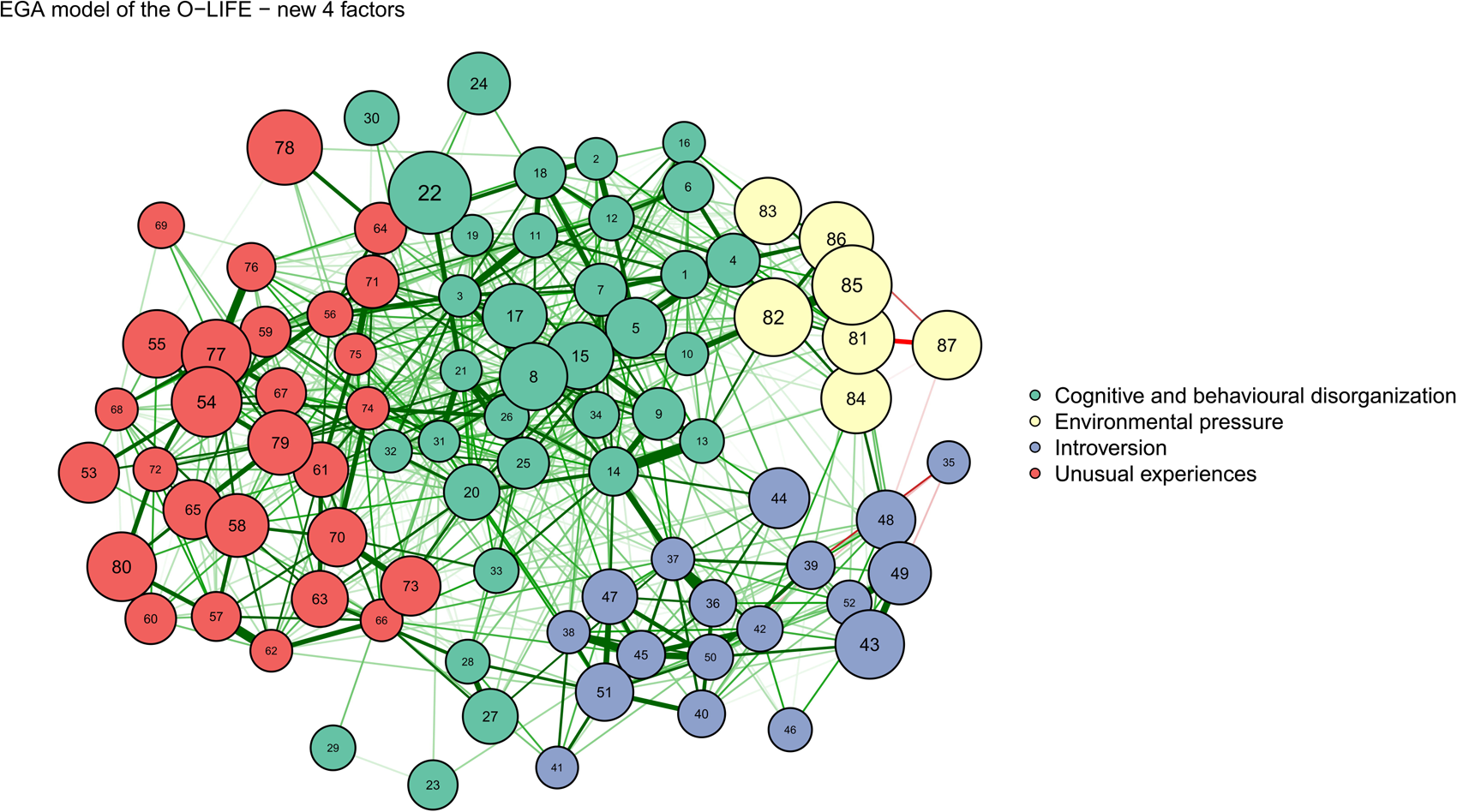
O-LIFE network representing the three factors identified by the EGA. The size of the nodes reflects the Betweenness Centrality value. The green edges represent positive Spearman correlations, red edges are negative.

**Table 2:**
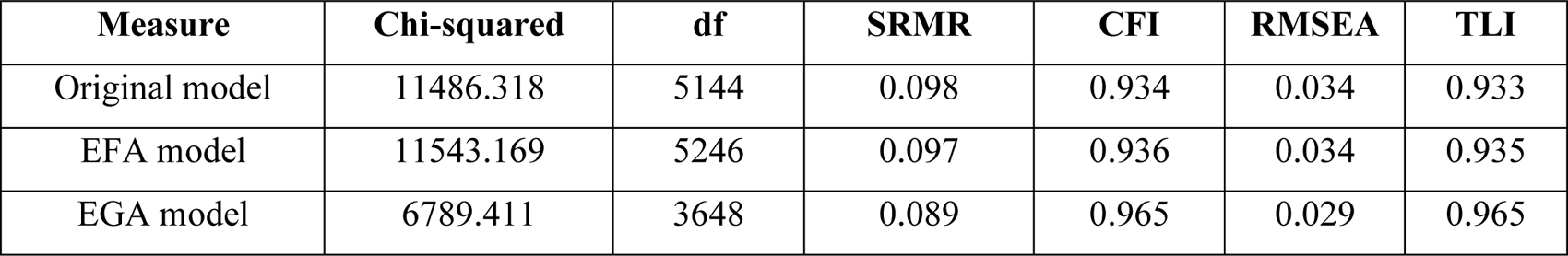
This summarizes the fit measures for each model evaluated (Original, Exploratory Factor Analysis, and EGA done with 87 items), including chi-square, degrees of freedom (df), standardized root mean square residual (SRMR), comparative fit index (CFI), root mean square error of approximation (RMSEA), and the Tucker-Lewis index (TLI).

| Measure | Chi-squared | df | SRMR | CFI | RMSEA | TLI |
| --- | --- | --- | --- | --- | --- | --- |
| Original model | 11486.318 | 5144 | 0.098 | 0.934 | 0.034 | 0.933 |
| EFA model | 11543.169 | 5246 | 0.097 | 0.936 | 0.034 | 0.935 |
| EGA model | 6789.411 | 3648 | 0.089 | 0.965 | 0.029 | 0.965 |

## 4. Discussion

This study was the first to test the validity of the three- and four-factor structures, respectively, in the German versions of the MSS and O-LIFE, using a single sample of healthy subjects and applying EGA. The resulting models were compared with the original calibrations, and our results demonstrate that EGA, in addition to providing more information on the relationships between individual items, offers fittings that provide new insights into the structure of schizotypy.

Regarding the Multidimensional Schizotypy Scale, the EGA results revealed a three-dimensional structure (positive, negative, and disorganized schizotypy), consistent with the theoretical dimensions. Eight items were misclassified into the positive dimension. An analysis of these individual items indicated that they referred to aspects related to the person’s disconnection from the world and/or difficulty in understanding conversations with others or not being understood during conversations, as well as confusion about what is happening around them. These items, identified in the new domain “disconnection,” reflect symptoms of derealization/depersonalization, often found in schizotypy and/or the early stages of schizophrenia(Hamilton & Simeon, 2019). Interestingly, these items/nodes are located exactly between the Positive and Disorganized domains and, with their Betweenness values, serve a “mediating” function between the two (DIS06: 0.46; DIS30: 0.7; DIS42: 0.64; DIS69: 0.52). The separation of these items into a new domain did not lead to a decrease in the validity of the other three domains, which maintained excellent Alpha and Omega values; instead, it provided a new index to consider, which in turn can be expanded.

As found in previous studies (Christensen et al., 2018), the positive and negative domains are more distinct, with the disorganized domain acting as a link between the two. In addition, we did not observe any subdivision within the negative domain or the positive whereas in the study by Christensen et al., 2019, the ‘Negative’ domain was divided by Exploratory Graph Analysis into two distinct groups that were identified as affective anhedonia and social anhedonia (Christensen et al., 2019).

Although statistically there are no differences between the three models analysed for the MSS, the four-dimension model shows better indices and provides more detailed insights into a symptom that may serve as an early behavioural warning sign for schizotypal disorder and schizophrenia.

The Oxford-Liverpool Inventory of Feelings and Experiences required adjustments primarily due to its larger number of items and the lesser stability of certain factors already observed in the calibration of the German version. Notably, Figure 2 shows that Impulsive Nonconformity is an extremely disseminated domain, which is reflected in its unacceptable internal validity found in other studies (Grant et al., 2013).

The re-analysis of the individual MSAs of each item made it possible to exclude those that could lead to greater problems in modelling and play a role as confounding variables.

The final questionnaire comprised 87 items compared to the original 104, and the 4-factor model, made explicit in the analyses, was significantly better than both the original theoretical model and that identified by means of Exploratory Factor Analysis.

The domains “Unusual Experiences” and “Introvertive Anhedonia”, now renamed “Introversion”, correspond to those of the theoretical model. Two new domains were identified: one defined as “Cognitive and behavioural disorganisation”, which was created by merging the items of “Impulsive nonconformity” and “Cognitive disorganisation”, showing that rather than divided they can be considered in a symbiotic relationship.

The main point identified thanks to the Exploratory Graph Analysis is the domain of what we have called “Environmental pressure” which, although it remains graphically at a marginal point in the network, its nodes all have high Betweenness centrality values.

These items (six from the Cognitive disorganisation scale and one from the Impulsive nonconformity scale), all refer to the fear of being judged/wounded by others for the decisions or actions made. It clearly emerges how the environment and the opinion/judgment of others has an effect and acts as a conduit between Introversion and Disorganisation.

Carrying out a proper assessment of environmental aspects such as relationships and the degree of pressure felt by the person on his or her behaviour, can make it possible to discriminate whether such thinking/tracking may also be due to a misinterpretation by the individual, highlighting emotional deficits in the schizotypal personality (Martin et al., 2019).

Both tests demonstrated a stable four-factor structure and, while some domains can be compared with each other, they provide two different interpretations of schizotypy. The MSS clearly shows the dimensionality of the construct since the Positive and Negative domains remain well-separated, and Disorganization is more correlated with Positivity through the newly identified domain called Disconnection. Furthermore, this test presents a more robust and differentiated factor structure compared to the O-LIFE. The latter, referring to a “full-dimensional” model of Schizotypy(Goulding, 2004; O. Mason & Claridge, 2006; Pfarr et al., 2023) and aiming to include many more characteristics, like behavioural ones, is more complex and more easily influenced by demographic and cultural aspects. This characteristic must be considered in possible future calibrations of the questionnaire in other populations and cultures.

This study suffers from some limitations that should be mentioned. First, all the model analysed in this article presented out of range SRMR while all the other measures are considered good or optimal. Since the goodness of a model should not be based on a single fitting index, the models were presented as valid while considering the presence of some residuals responsible for an SRMR value greater than 0.8.

Secondly, it is conceivable that due to the extensive number of items in both instruments, participants could experience content overload, leading to decreased attention and potentially random responses. Finally, because this is a self-report assessment, certain inherent issues cannot be anticipated or prevented. For instance, it is uncertain whether subjects respond truthfully to all items. Additionally, some responses may be influenced by social desirability bias, where subjects answer in a way they believe aligns with societal expectations.

## 5. Conclusion

This study advances the understanding of schizotypy by proposing possible new factor structures of the MSS and O-LIFE using Exploratory Graph Analysis. MSS’s clear dimensionality and the addition of the disconnection domain offer a precise framework for identifying schizotypal traits. O-LIFE’s revised model, with its focus on environmental influences, provides a comprehensive, though complex, interpretation of schizotypal characteristics. Both scales contribute uniquely to the assessment and understanding of schizotypy in healthy populations.

Future research should explore more in depth and rearrange the number of items of the new domains to assess and increase validity and stability of the new ones considering the possible differences in language and culture.

## Supporting information

Supplementary R Code for analysis and results

## Data Availability

All data produced in the present study are available upon reasonable request to the corresponding Authors

## Acknowledgements

This research was supported by grants from the Brain & Behavior Research Foundation and the OPO Foundation. We gratefully acknowledge their financial assistance, which made this work possible.

## Ethical standards

The authors assert that the protocol was approved by the local ethics committee (KEK-ZH 2020/01049) and that all procedures contributing to this work comply with the ethical standards of the relevant national and institutional committees on human experimentation and with the Helsinki Declaration of 1975, as revised in 2008.

