## Supplementary R Code for analysis and results for "Investigating the structure of Schizotypy through the “Multidimensional Schizotypy Scale” and “Oxford-Liverpool Inventory”: an Exploratory network analysis approach in the healthy population"

### R\_code and Results

Pierfrancesco Sarti

Uploading the 2 datasets (MSS and OLIFE) and extracted only the test ITEMS

```
library(readxl)

load(file = "MSS_OLIFE_environment.RData")

data.MSS <- read_xlsx("1_MSS.xlsx")
data.OLIFE <- read_xlsx("1_OLIFE.xlsx")

# MSS
colnames(data.MSS)
```

```
## [1] "Age"      "Group"    "Gender"   "Education" "01NEG"    "04NEG"
## [7] "07NEG"    "10NEG"    "13NEG"    "16NEG"     "19NEG"    "22NEG"
## [13] "25NEG"    "28NEG"    "31NEG"    "34NEG"     "37NEG"    "40NEG"
## [19] "43NEG"    "46NEG"    "49NEG"    "52NEG"     "55NEG"    "58NEG"
## [25] "61NEG"    "64NEG"    "67NEG"    "70NEG"     "73NEG"    "76NEG"
## [31] "02POS"    "05POS"    "08POS"    "11POS"     "14POS"    "17POS"
## [37] "20POS"    "23POS"    "26POS"    "29POS"     "32POS"    "35POS"
## [43] "38POS"    "41POS"    "44POS"    "47POS"     "50POS"    "53POS"
## [49] "56POS"    "59POS"    "62POS"    "65POS"     "68POS"    "71POS"
## [55] "74POS"    "77POS"    "03DIS"    "06DIS"     "09DIS"    "12DIS"
## [61] "15DIS"    "18DIS"    "21DIS"    "24DIS"     "27DIS"    "30DIS"
## [67] "33DIS"    "36DIS"    "39DIS"    "42DIS"     "45DIS"    "48DIS"
## [73] "51DIS"    "54DIS"    "57DIS"    "60DIS"     "63DIS"    "66DIS"
## [79] "69DIS"    "72DIS"    "75DIS"
```

```
MSS <- data.MSS[,c(5:81)]

colnames(MSS) <- c("NEG01", "NEG04", "NEG07", "NEG10", "NEG13", "NEG16",
                  "NEG19", "NEG22", "NEG25", "NEG28", "NEG31", "NEG34",
                  "NEG37", "NEG40", "NEG43", "NEG46", "NEG49", "NEG52",
                  "NEG55", "NEG58", "NEG61", "NEG64", "NEG67", "NEG70",
                  "NEG73", "NEG76", "POS02", "POS05", "POS08", "POS11",
                  "POS14", "POS17", "POS20", "POS23", "POS26", "POS29",
                  "POS32", "POS35", "POS38", "POS41", "POS44", "POS47",
                  "POS50", "POS53", "POS56", "POS59", "POS62", "POS65",
                  "POS68", "POS71", "POS74", "POS77", "DIS03", "DIS06",
                  "DIS09", "DIS12", "DIS15", "DIS18", "DIS21", "DIS24",
                  "DIS27", "DIS30", "DIS33", "DIS36", "DIS39", "DIS42",
                  "DIS45", "DIS48", "DIS51", "DIS54", "DIS57", "DIS60",
                  "DIS63", "DIS66", "DIS69", "DIS72", "DIS75")

sum(is.na(data.MSS))
```

```
## [1] 0
```

```
# OLIFE
colnames(data.OLIFE)
```

```
## [1] "Age"      "Group"    "Gender"   "Education" "COG001"    "COG008"
## [7] "COG016"    "COG021"    "COG022"    "COG023"     "COG028"    "COG032"
```

```
## [13] "COG036"      "COG037"      "COG038"      "COG043"      "COG054"      "COG055"
## [19] "COG061"      "COG062"      "COG066"      "COG073"      "COG075"      "COG081"
## [25] "COG085"      "COG091"      "COG095"      "COG101"      "IMP002"      "IMP007"
## [31] "IMP010"      "IMP012"      "IMP013"      "IMP015"      "IMP018"      "IMP019"
## [37] "IMP024"      "IMP031"      "IMP051"      "IMP052"      "IMP053"      "IMP056"
## [43] "IMP057"      "IMP068"      "IMP069"      "IMP070"      "IMP074"      "IMP084"
## [49] "IMP088"      "IMP096"      "IMP100"      "UNEX003"      "UNEX005"      "UNEX006"
## [55] "UNEX017"      "UNEX020"      "UNEX025"      "UNEX027"      "UNEX029"      "UNEX030"
## [61] "UNEX033"      "UNEX040"      "UNEX044"      "UNEX047"      "UNEX058"      "UNEX060"
## [67] "UNEX063"      "UNEX065"      "UNEX071"      "UNEX072"      "UNEX076"      "UNEX078"
## [73] "UNEX079"      "UNEX082"      "UNEX083"      "UNEX087"      "UNEX089"      "UNEX090"
## [79] "UNEX093"      "UNEX099"      "UNEX103"      "INT004"      "INT009"      "INT011"
## [85] "INT014"      "INT026"      "INT034"      "INT035"      "INT039"      "INT041"
## [91] "INT042"      "INT045"      "INT046"      "INT048"      "INT049"      "INT050"
## [97] "INT059"      "INT064"      "INT067"      "INT077"      "INT080"      "INT086"
## [103] "INT092"      "INT094"      "INT097"      "INT098"      "INT102"      "INT104"
```

```
OLIFE <- data.OLIFE[,c(5:108)]
sum(is.na(data.OLIFE))
```

```
## [1] 0
```

Libraries needed for the Exploratory Graph Analysis - CFA - EFA

```
library(psych)
library(NetworkToolbox)
library(igraph)
library(qgraph)
library(ega)
library(lavaan)
library(EGAnet)
library(IsingFit)
library(polycor)
```

#### MULTIDIMENSIONAL SCHIZOTYPY SCALE

##### 1. CHOOSE THE BEST CORRELATIONAL METHOD:

```
# mat.MSS.fiml <- corFiml(MSS) # Full Information Maximum Likelihood (FIML) is a
# robust method to compute correlations but used mostly when there are NAs

mat.MSS <- cor(MSS, method = "spearman")
# spearman correlations are the best option for categorical data!

# Phi Coefficient is the recommended for DiCHOTOMIC variables
# Function to calculate Phi coefficient for a pair of variables
calculate_phi <- function(x, y) {
  table <- table(x, y)
  phi(table)
}

# Get all pairs of columns
pairsMSS <- combn(ncol(MSS), 2)

# Calculate Phi coefficient for each pair
phi_matrix_MSS <- matrix(NA, ncol(MSS), ncol(MSS))
colnames(phi_matrix_MSS) <- colnames(MSS)
rownames(phi_matrix_MSS) <- colnames(MSS)

for (i in 1:ncol(pairsMSS)) {
  var1 <- pairsMSS[1, i]
  var2 <- pairsMSS[2, i]
  phi_valueMSS <- calculate_phi(MSS[[var1]], MSS[[var2]])
  phi_matrix_MSS[var1, var2] <- phi_valueMSS
  phi_matrix_MSS[var2, var1] <- phi_valueMSS
}

# Set diagonal to 1 as the Phi coefficient of a variable with itself is 1
diag(phi_matrix_MSS) <- 1

# Display the Phi coefficient matrix

head(print(phi_matrix_MSS))
```

```
##          NEG01 NEG04 NEG07 NEG10 NEG13 NEG16 NEG19 NEG22 NEG25 NEG28 NEG31 NEG34
## NEG01   1.00  0.09  0.14  0.23  0.15  0.37  0.14  0.14  0.17  0.18  0.21  0.11
## NEG04   0.09  1.00  0.18  0.13  0.05  0.05  0.34  0.14  0.08  0.07  0.09  0.15
## NEG07   0.14  0.18  1.00  0.26  0.10  0.18  0.24  0.27  0.14  0.08  0.25  0.22
## NEG10   0.23  0.13  0.26  1.00  0.21  0.17  0.26  0.24  0.20  0.18  0.17  0.24
## NEG13   0.15  0.05  0.10  0.21  1.00  0.10  0.10  0.11  0.18  0.10  0.15  0.11
## NEG16   0.37  0.05  0.18  0.17  0.10  1.00  0.11  0.14  0.28  0.26  0.09  0.00
## NEG19   0.14  0.34  0.24  0.26  0.10  0.11  1.00  0.21  0.07  0.12  0.15  0.28
## NEG22   0.14  0.14  0.27  0.24  0.11  0.14  0.21  1.00  0.14  0.21  0.19  0.19
## NEG25   0.17  0.08  0.14  0.20  0.18  0.28  0.07  0.14  1.00  0.21  0.13  0.05
```

|  |  |  |  |  |  |  |  |  |  |  |  |  |  |
| --- | --- | --- | --- | --- | --- | --- | --- | --- | --- | --- | --- | --- | --- |
| ## | NEG28 | 0.18 | 0.07 | 0.08 | 0.18 | 0.10 | 0.26 | 0.12 | 0.21 | 0.21 | 1.00 | 0.15 | 0.14 |
| ## | NEG31 | 0.21 | 0.09 | 0.25 | 0.17 | 0.15 | 0.09 | 0.15 | 0.19 | 0.13 | 0.15 | 1.00 | 0.21 |
| ## | NEG34 | 0.11 | 0.15 | 0.22 | 0.24 | 0.11 | 0.00 | 0.28 | 0.19 | 0.05 | 0.14 | 0.21 | 1.00 |
| ## | NEG37 | 0.24 | 0.03 | 0.14 | 0.10 | 0.13 | 0.35 | 0.03 | 0.11 | 0.29 | 0.20 | 0.17 | 0.08 |
| ## | NEG40 | 0.15 | 0.07 | 0.07 | 0.25 | 0.22 | 0.20 | 0.15 | 0.11 | 0.17 | 0.17 | 0.10 | 0.10 |
| ## | NEG43 | 0.12 | 0.08 | 0.13 | 0.20 | 0.21 | 0.24 | 0.15 | 0.09 | 0.25 | 0.16 | 0.19 | 0.14 |
| ## | NEG46 | 0.16 | 0.17 | 0.17 | 0.31 | 0.19 | 0.16 | 0.24 | 0.33 | 0.25 | 0.17 | 0.12 | 0.17 |
| ## | NEG49 | 0.10 | 0.23 | 0.23 | 0.13 | 0.01 | 0.05 | 0.22 | 0.28 | 0.03 | 0.10 | 0.06 | 0.14 |
| ## | NEG52 | 0.19 | 0.09 | 0.21 | 0.29 | 0.19 | 0.27 | 0.12 | 0.14 | 0.19 | 0.10 | 0.22 | 0.08 |
| ## | NEG55 | 0.22 | 0.10 | 0.14 | 0.24 | 0.23 | 0.28 | 0.06 | 0.12 | 0.47 | 0.20 | 0.24 | 0.11 |
| ## | NEG58 | 0.12 | 0.09 | 0.06 | 0.15 | 0.09 | 0.10 | 0.09 | 0.15 | 0.18 | 0.27 | 0.13 | 0.10 |
| ## | NEG61 | 0.19 | 0.04 | 0.18 | 0.28 | 0.18 | 0.31 | 0.12 | 0.18 | 0.29 | 0.14 | 0.24 | 0.12 |
| ## | NEG64 | 0.15 | 0.07 | 0.10 | 0.21 | 0.16 | 0.17 | 0.07 | 0.13 | 0.54 | 0.21 | 0.14 | 0.09 |
| ## | NEG67 | 0.41 | 0.10 | 0.24 | 0.25 | 0.18 | 0.58 | 0.13 | 0.20 | 0.40 | 0.30 | 0.15 | 0.10 |
| ## | NEG70 | 0.23 | 0.05 | 0.23 | 0.26 | 0.13 | 0.24 | 0.09 | 0.14 | 0.15 | 0.14 | 0.34 | 0.20 |
| ## | NEG73 | 0.23 | 0.06 | 0.11 | 0.25 | 0.25 | 0.27 | 0.07 | 0.12 | 0.54 | 0.17 | 0.23 | 0.12 |
| ## | NEG76 | 0.10 | 0.09 | 0.16 | 0.24 | 0.06 | 0.06 | 0.12 | 0.11 | 0.21 | 0.21 | 0.22 | 0.25 |
| ## | POS02 | 0.11 | 0.01 | 0.05 | 0.10 | 0.10 | 0.07 | 0.07 | 0.06 | 0.05 | 0.03 | 0.00 | 0.05 |
| ## | POS05 | 0.03 | -0.01 | 0.04 | 0.07 | 0.08 | 0.18 | 0.03 | 0.02 | 0.06 | 0.07 | -0.04 | 0.01 |
| ## | POS08 | 0.11 | 0.01 | 0.08 | 0.09 | 0.12 | 0.07 | 0.07 | 0.03 | 0.16 | -0.02 | -0.01 | 0.03 |
| ## | POS11 | 0.05 | 0.04 | 0.02 | 0.14 | 0.24 | 0.08 | 0.00 | 0.06 | 0.19 | 0.13 | 0.04 | 0.06 |
| ## | POS14 | 0.03 | 0.01 | 0.09 | 0.09 | -0.02 | 0.17 | 0.03 | 0.03 | 0.10 | 0.09 | 0.03 | 0.04 |
| ## | POS17 | 0.11 | -0.06 | 0.07 | 0.04 | 0.06 | 0.18 | -0.02 | -0.02 | 0.17 | 0.04 | 0.02 | 0.00 |
| ## | POS20 | 0.05 | 0.02 | 0.03 | 0.01 | 0.10 | 0.07 | 0.09 | 0.03 | 0.06 | -0.02 | -0.01 | 0.04 |
| ## | POS23 | 0.11 | 0.00 | -0.05 | 0.01 | 0.10 | 0.12 | 0.05 | -0.02 | 0.01 | 0.13 | 0.07 | 0.00 |
| ## | POS26 | 0.07 | -0.01 | -0.01 | 0.01 | 0.09 | 0.17 | 0.03 | 0.06 | 0.09 | 0.10 | -0.01 | 0.01 |
| ## | POS29 | 0.04 | -0.02 | 0.01 | 0.04 | 0.07 | 0.09 | 0.05 | 0.01 | 0.10 | 0.01 | 0.00 | 0.01 |
| ## | POS32 | 0.03 | 0.00 | 0.01 | 0.01 | 0.08 | 0.11 | 0.02 | 0.01 | 0.07 | 0.09 | 0.03 | 0.04 |
| ## | POS35 | 0.02 | 0.08 | 0.00 | 0.07 | 0.10 | 0.05 | 0.06 | 0.04 | 0.16 | 0.09 | 0.06 | 0.03 |
| ## | POS38 | 0.19 | 0.03 | 0.04 | 0.09 | 0.16 | 0.10 | 0.03 | 0.05 | 0.10 | 0.06 | 0.08 | 0.07 |
| ## | POS41 | 0.09 | 0.03 | 0.00 | 0.11 | 0.10 | 0.08 | 0.07 | 0.03 | 0.20 | 0.09 | 0.04 | 0.05 |
| ## | POS44 | 0.05 | 0.02 | 0.00 | 0.02 | 0.12 | 0.08 | 0.05 | 0.06 | 0.12 | 0.04 | 0.02 | 0.01 |
| ## | POS47 | 0.09 | 0.00 | 0.00 | 0.06 | 0.15 | 0.19 | 0.07 | 0.03 | 0.16 | 0.04 | 0.00 | 0.09 |
| ## | POS50 | 0.07 | 0.03 | 0.02 | 0.11 | 0.14 | 0.14 | 0.03 | 0.01 | 0.21 | 0.22 | 0.06 | 0.06 |
| ## | POS53 | 0.04 | 0.03 | -0.01 | 0.10 | 0.16 | 0.24 | 0.06 | -0.06 | 0.14 | 0.16 | 0.00 | 0.01 |
| ## | POS56 | 0.06 | 0.01 | 0.03 | 0.09 | 0.13 | 0.12 | 0.09 | 0.06 | 0.14 | 0.12 | 0.04 | 0.05 |
| ## | POS59 | 0.09 | 0.00 | 0.00 | 0.06 | 0.17 | 0.14 | 0.08 | 0.01 | 0.08 | 0.13 | -0.01 | 0.03 |
| ## | POS62 | 0.13 | 0.02 | 0.01 | 0.07 | 0.16 | 0.19 | 0.08 | 0.06 | 0.17 | 0.03 | 0.00 | 0.03 |
| ## | POS65 | 0.14 | 0.04 | 0.04 | 0.13 | 0.17 | 0.02 | 0.11 | 0.10 | 0.09 | 0.05 | 0.11 | 0.11 |
| ## | POS68 | 0.03 | 0.04 | 0.04 | 0.10 | 0.12 | 0.07 | 0.00 | 0.03 | 0.11 | 0.03 | -0.02 | 0.01 |
| ## | POS71 | 0.11 | 0.04 | 0.03 | 0.06 | 0.08 | 0.20 | 0.04 | 0.02 | 0.17 | 0.03 | -0.03 | 0.05 |
| ## | POS74 | 0.08 | 0.04 | -0.02 | 0.08 | 0.11 | 0.06 | 0.07 | 0.01 | 0.07 | 0.09 | 0.01 | 0.07 |
| ## | POS77 | 0.05 | 0.03 | -0.02 | 0.08 | 0.10 | 0.15 | 0.05 | 0.03 | 0.09 | 0.13 | 0.02 | 0.00 |
| ## | DIS03 | 0.10 | 0.01 | 0.04 | 0.03 | 0.15 | 0.10 | 0.07 | 0.00 | 0.19 | 0.07 | 0.11 | 0.01 |
| ## | DIS06 | 0.16 | 0.06 | 0.09 | 0.13 | 0.20 | 0.24 | 0.09 | 0.04 | 0.23 | 0.17 | 0.08 | 0.03 |
| ## | DIS09 | 0.07 | 0.00 | 0.05 | 0.03 | 0.18 | 0.14 | 0.05 | -0.01 | 0.29 | 0.08 | 0.03 | -0.02 |
| ## | DIS12 | 0.11 | 0.01 | 0.04 | 0.10 | 0.12 | 0.13 | 0.08 | -0.02 | 0.17 | 0.15 | 0.01 | 0.05 |
| ## | DIS15 | 0.09 | 0.03 | 0.02 | 0.07 | 0.21 | 0.16 | 0.04 | -0.02 | 0.26 | 0.05 | 0.02 | 0.05 |
| ## | DIS18 | 0.12 | 0.02 | 0.00 | 0.03 | 0.15 | 0.12 | 0.08 | 0.01 | 0.23 | 0.05 | 0.07 | 0.05 |
| ## | DIS21 | 0.10 | -0.03 | 0.02 | 0.09 | 0.21 | 0.13 | 0.01 | 0.01 | 0.21 | 0.13 | 0.10 | 0.04 |
| ## | DIS24 | 0.08 | -0.02 | -0.01 | 0.05 | 0.10 | 0.09 | 0.07 | -0.01 | 0.16 | 0.07 | 0.03 | 0.05 |
| ## | DIS27 | 0.12 | 0.01 | 0.06 | 0.03 | 0.10 | 0.11 | 0.05 | 0.01 | 0.19 | 0.03 | 0.08 | -0.01 |
| ## | DIS30 | 0.15 | 0.03 | 0.04 | 0.11 | 0.18 | 0.26 | 0.04 | 0.09 | 0.20 | 0.15 | 0.09 | 0.05 |
| ## | DIS33 | 0.09 | -0.01 | 0.03 | 0.07 | 0.17 | 0.05 | 0.03 | 0.01 | 0.21 | 0.04 | 0.11 | 0.01 |

|  |  |  |  |  |  |  |  |  |  |  |  |  |  |
| --- | --- | --- | --- | --- | --- | --- | --- | --- | --- | --- | --- | --- | --- |
| ## | DIS36 | 0.11 | -0.01 | 0.03 | 0.08 | 0.16 | 0.11 | 0.09 | 0.03 | 0.19 | 0.03 | 0.04 | 0.04 |
| ## | DIS39 | 0.13 | 0.05 | 0.07 | 0.09 | 0.19 | 0.17 | 0.04 | 0.08 | 0.27 | 0.05 | 0.13 | 0.02 |
| ## | DIS42 | 0.13 | 0.04 | 0.04 | 0.14 | 0.21 | 0.11 | 0.05 | 0.07 | 0.37 | 0.12 | 0.18 | 0.11 |
| ## | DIS45 | 0.05 | -0.02 | 0.03 | 0.09 | 0.20 | 0.16 | -0.01 | 0.02 | 0.28 | 0.10 | 0.05 | -0.03 |
| ## | DIS48 | 0.10 | 0.00 | 0.03 | 0.04 | 0.10 | 0.11 | 0.08 | 0.01 | 0.19 | 0.15 | 0.06 | 0.06 |
| ## | DIS51 | 0.13 | -0.03 | 0.03 | 0.11 | 0.23 | 0.17 | 0.04 | 0.01 | 0.26 | 0.04 | 0.00 | 0.06 |
| ## | DIS54 | 0.10 | -0.01 | -0.02 | 0.03 | 0.11 | 0.04 | 0.00 | -0.04 | 0.18 | 0.11 | 0.02 | 0.00 |
| ## | DIS57 | 0.10 | 0.02 | 0.02 | 0.08 | 0.18 | 0.09 | 0.04 | 0.03 | 0.29 | 0.07 | 0.12 | 0.04 |
| ## | DIS60 | 0.12 | 0.01 | 0.08 | 0.11 | 0.18 | 0.08 | 0.02 | 0.05 | 0.22 | 0.09 | 0.11 | 0.01 |
| ## | DIS63 | 0.09 | 0.03 | -0.02 | 0.06 | 0.09 | 0.07 | 0.06 | 0.01 | 0.19 | 0.10 | 0.04 | 0.05 |
| ## | DIS66 | 0.16 | -0.02 | 0.09 | 0.06 | 0.23 | 0.20 | 0.02 | 0.01 | 0.28 | 0.08 | 0.05 | 0.02 |
| ## | DIS69 | 0.09 | -0.02 | 0.01 | 0.07 | 0.17 | 0.12 | 0.03 | 0.00 | 0.24 | 0.06 | 0.04 | 0.01 |
| ## | DIS72 | 0.13 | 0.03 | 0.02 | 0.08 | 0.18 | 0.14 | 0.05 | 0.01 | 0.29 | 0.07 | 0.12 | 0.04 |
| ## | DIS75 | 0.01 | 0.01 | 0.00 | 0.02 | 0.09 | 0.08 | 0.02 | -0.03 | 0.10 | 0.12 | 0.05 | 0.01 |
| ## |  | NEG37 | NEG40 | NEG43 | NEG46 | NEG49 | NEG52 | NEG55 | NEG58 | NEG61 | NEG64 | NEG67 | NEG70 |
| ## | NEG01 | 0.24 | 0.15 | 0.12 | 0.16 | 0.10 | 0.19 | 0.22 | 0.12 | 0.19 | 0.15 | 0.41 | 0.23 |
| ## | NEG04 | 0.03 | 0.07 | 0.08 | 0.17 | 0.23 | 0.09 | 0.10 | 0.09 | 0.04 | 0.07 | 0.10 | 0.05 |
| ## | NEG07 | 0.14 | 0.07 | 0.13 | 0.17 | 0.23 | 0.21 | 0.14 | 0.06 | 0.18 | 0.10 | 0.24 | 0.23 |
| ## | NEG10 | 0.10 | 0.25 | 0.20 | 0.31 | 0.13 | 0.29 | 0.24 | 0.15 | 0.28 | 0.21 | 0.25 | 0.26 |
| ## | NEG13 | 0.13 | 0.22 | 0.21 | 0.19 | 0.01 | 0.19 | 0.23 | 0.09 | 0.18 | 0.16 | 0.18 | 0.13 |
| ## | NEG16 | 0.35 | 0.20 | 0.24 | 0.16 | 0.05 | 0.27 | 0.28 | 0.10 | 0.31 | 0.17 | 0.58 | 0.24 |
| ## | NEG19 | 0.03 | 0.15 | 0.15 | 0.24 | 0.22 | 0.12 | 0.06 | 0.09 | 0.12 | 0.07 | 0.13 | 0.09 |
| ## | NEG22 | 0.11 | 0.11 | 0.09 | 0.33 | 0.28 | 0.14 | 0.12 | 0.15 | 0.18 | 0.13 | 0.20 | 0.14 |
| ## | NEG25 | 0.29 | 0.17 | 0.25 | 0.25 | 0.03 | 0.19 | 0.47 | 0.18 | 0.29 | 0.54 | 0.40 | 0.15 |
| ## | NEG28 | 0.20 | 0.17 | 0.16 | 0.17 | 0.10 | 0.10 | 0.20 | 0.27 | 0.14 | 0.21 | 0.30 | 0.14 |
| ## | NEG31 | 0.17 | 0.10 | 0.19 | 0.12 | 0.06 | 0.22 | 0.24 | 0.13 | 0.24 | 0.14 | 0.15 | 0.34 |
| ## | NEG34 | 0.08 | 0.10 | 0.14 | 0.17 | 0.14 | 0.08 | 0.11 | 0.10 | 0.12 | 0.09 | 0.10 | 0.20 |
| ## | NEG37 | 1.00 | 0.13 | 0.21 | 0.11 | 0.05 | 0.26 | 0.17 | 0.02 | 0.26 | 0.15 | 0.39 | 0.27 |
| ## | NEG40 | 0.13 | 1.00 | 0.66 | 0.28 | 0.08 | 0.23 | 0.16 | 0.12 | 0.21 | 0.25 | 0.25 | 0.24 |
| ## | NEG43 | 0.21 | 0.66 | 1.00 | 0.24 | 0.09 | 0.26 | 0.22 | 0.17 | 0.23 | 0.19 | 0.25 | 0.29 |
| ## | NEG46 | 0.11 | 0.28 | 0.24 | 1.00 | 0.12 | 0.16 | 0.23 | 0.19 | 0.19 | 0.22 | 0.30 | 0.25 |
| ## | NEG49 | 0.05 | 0.08 | 0.09 | 0.12 | 1.00 | 0.08 | 0.01 | 0.05 | 0.08 | 0.05 | 0.08 | 0.11 |
| ## | NEG52 | 0.26 | 0.23 | 0.26 | 0.16 | 0.08 | 1.00 | 0.31 | 0.01 | 0.39 | 0.17 | 0.29 | 0.30 |
| ## | NEG55 | 0.17 | 0.16 | 0.22 | 0.23 | 0.01 | 0.31 | 1.00 | 0.17 | 0.29 | 0.54 | 0.36 | 0.29 |
| ## | NEG58 | 0.02 | 0.12 | 0.17 | 0.19 | 0.05 | 0.01 | 0.17 | 1.00 | 0.06 | 0.18 | 0.17 | 0.04 |
| ## | NEG61 | 0.26 | 0.21 | 0.23 | 0.19 | 0.08 | 0.39 | 0.29 | 0.06 | 1.00 | 0.18 | 0.37 | 0.45 |
| ## | NEG64 | 0.15 | 0.25 | 0.19 | 0.22 | 0.05 | 0.17 | 0.54 | 0.18 | 0.18 | 1.00 | 0.26 | 0.19 |
| ## | NEG67 | 0.39 | 0.25 | 0.25 | 0.30 | 0.08 | 0.29 | 0.36 | 0.17 | 0.37 | 0.26 | 1.00 | 0.29 |
| ## | NEG70 | 0.27 | 0.24 | 0.29 | 0.25 | 0.11 | 0.30 | 0.29 | 0.04 | 0.45 | 0.19 | 0.29 | 1.00 |
| ## | NEG73 | 0.18 | 0.21 | 0.30 | 0.24 | 0.02 | 0.25 | 0.62 | 0.19 | 0.27 | 0.64 | 0.33 | 0.26 |
| ## | NEG76 | 0.24 | 0.09 | 0.19 | 0.12 | 0.06 | 0.22 | 0.24 | 0.13 | 0.11 | 0.22 | 0.14 | 0.13 |
| ## | POS02 | 0.01 | 0.10 | 0.07 | 0.05 | -0.05 | 0.09 | 0.08 | 0.08 | 0.07 | 0.06 | 0.11 | 0.06 |
| ## | POS05 | 0.04 | 0.12 | 0.11 | 0.05 | 0.00 | 0.13 | 0.10 | 0.04 | 0.07 | 0.04 | 0.14 | 0.03 |
| ## | POS08 | 0.06 | 0.10 | 0.14 | 0.06 | 0.01 | 0.13 | 0.04 | 0.04 | 0.14 | 0.11 | 0.12 | 0.16 |
| ## | POS11 | 0.01 | 0.17 | 0.16 | 0.15 | -0.04 | 0.10 | 0.25 | 0.15 | 0.08 | 0.16 | 0.13 | 0.07 |
| ## | POS14 | 0.13 | 0.10 | 0.13 | 0.05 | 0.03 | 0.15 | 0.10 | 0.02 | 0.09 | 0.06 | 0.13 | 0.11 |
| ## | POS17 | 0.07 | 0.05 | 0.01 | 0.01 | -0.10 | 0.13 | 0.20 | 0.05 | 0.11 | 0.09 | 0.14 | 0.04 |
| ## | POS20 | -0.01 | 0.08 | 0.10 | 0.00 | -0.02 | 0.12 | 0.12 | 0.03 | 0.09 | 0.04 | 0.02 | 0.04 |
| ## | POS23 | 0.02 | 0.05 | 0.04 | 0.01 | -0.04 | 0.07 | 0.01 | 0.05 | 0.04 | 0.05 | 0.08 | 0.04 |
| ## | POS26 | 0.04 | 0.07 | 0.07 | 0.09 | -0.05 | 0.13 | 0.10 | 0.02 | 0.11 | 0.03 | 0.12 | 0.07 |
| ## | POS29 | 0.02 | 0.03 | 0.01 | 0.00 | -0.07 | 0.10 | 0.08 | 0.02 | 0.13 | 0.08 | 0.07 | 0.02 |
| ## | POS32 | 0.03 | 0.10 | 0.10 | 0.09 | -0.06 | 0.05 | 0.06 | 0.06 | 0.01 | 0.11 | 0.07 | -0.02 |
| ## | POS35 | -0.02 | 0.08 | 0.07 | 0.06 | -0.06 | 0.10 | 0.12 | 0.01 | 0.11 | 0.13 | 0.02 | 0.04 |
| ## | POS38 | 0.08 | 0.10 | 0.17 | 0.11 | 0.02 | 0.08 | 0.06 | 0.06 | 0.13 | 0.11 | 0.16 | 0.17 |

|  |  |  |  |  |  |  |  |  |  |  |  |  |
| --- | --- | --- | --- | --- | --- | --- | --- | --- | --- | --- | --- | --- |
| ## POS41 | -0.01 | 0.18 | 0.15 | 0.10 | -0.06 | 0.10 | 0.22 | 0.10 | 0.11 | 0.20 | 0.14 | 0.03 |
| ## POS44 | 0.03 | 0.08 | 0.10 | 0.04 | 0.00 | 0.11 | 0.11 | 0.04 | 0.10 | 0.10 | 0.07 | 0.02 |
| ## POS47 | 0.07 | 0.12 | 0.11 | 0.11 | 0.02 | 0.05 | 0.15 | 0.04 | 0.03 | 0.10 | 0.14 | 0.05 |
| ## POS50 | 0.00 | 0.17 | 0.13 | 0.09 | -0.07 | 0.09 | 0.34 | 0.15 | 0.15 | 0.24 | 0.14 | 0.09 |
| ## POS53 | 0.08 | 0.10 | 0.10 | 0.03 | -0.04 | 0.09 | 0.17 | 0.08 | 0.12 | 0.12 | 0.18 | -0.02 |
| ## POS56 | 0.08 | 0.18 | 0.20 | 0.07 | 0.00 | 0.13 | 0.11 | 0.14 | 0.12 | 0.09 | 0.18 | 0.06 |
| ## POS59 | 0.02 | 0.12 | 0.09 | 0.09 | -0.04 | 0.14 | 0.17 | 0.05 | 0.08 | 0.08 | 0.17 | 0.04 |
| ## POS62 | 0.15 | 0.11 | 0.11 | 0.11 | -0.05 | 0.15 | 0.15 | 0.03 | 0.16 | 0.11 | 0.18 | 0.10 |
| ## POS65 | 0.05 | 0.14 | 0.13 | 0.13 | 0.02 | 0.17 | 0.04 | 0.09 | 0.13 | 0.10 | 0.15 | 0.15 |
| ## POS68 | 0.03 | 0.14 | 0.14 | 0.06 | -0.03 | 0.20 | 0.15 | 0.11 | 0.09 | 0.07 | 0.11 | 0.11 |
| ## POS71 | 0.02 | 0.03 | 0.03 | 0.05 | -0.05 | 0.08 | 0.14 | 0.04 | 0.09 | 0.08 | 0.12 | 0.03 |
| ## POS74 | -0.03 | 0.09 | 0.08 | 0.07 | -0.04 | 0.05 | 0.07 | 0.06 | 0.08 | 0.04 | 0.11 | 0.07 |
| ## POS77 | -0.02 | 0.05 | 0.08 | 0.04 | -0.08 | 0.04 | 0.09 | 0.12 | 0.08 | 0.09 | 0.11 | -0.02 |
| ## DIS03 | 0.04 | 0.08 | 0.09 | 0.11 | -0.05 | 0.10 | 0.19 | 0.08 | 0.13 | 0.18 | 0.11 | 0.03 |
| ## DIS06 | 0.08 | 0.16 | 0.15 | 0.07 | -0.02 | 0.27 | 0.27 | 0.15 | 0.15 | 0.21 | 0.17 | 0.04 |
| ## DIS09 | 0.02 | 0.11 | 0.12 | 0.09 | -0.03 | 0.09 | 0.21 | 0.10 | 0.10 | 0.22 | 0.16 | 0.01 |
| ## DIS12 | 0.07 | 0.18 | 0.12 | 0.08 | 0.00 | 0.04 | 0.15 | 0.08 | 0.04 | 0.16 | 0.13 | 0.03 |
| ## DIS15 | 0.11 | 0.14 | 0.14 | 0.11 | -0.06 | 0.14 | 0.25 | 0.05 | 0.09 | 0.21 | 0.17 | 0.01 |
| ## DIS18 | 0.03 | 0.12 | 0.11 | 0.11 | -0.07 | 0.09 | 0.20 | 0.06 | 0.12 | 0.17 | 0.15 | 0.02 |
| ## DIS21 | 0.07 | 0.15 | 0.14 | 0.12 | -0.05 | 0.10 | 0.32 | 0.07 | 0.06 | 0.28 | 0.14 | 0.05 |
| ## DIS24 | 0.01 | 0.16 | 0.12 | 0.08 | -0.03 | 0.07 | 0.15 | 0.04 | 0.09 | 0.18 | 0.10 | 0.02 |
| ## DIS27 | 0.00 | 0.01 | 0.07 | 0.05 | -0.03 | 0.13 | 0.26 | 0.06 | 0.07 | 0.22 | 0.07 | 0.01 |
| ## DIS30 | 0.09 | 0.18 | 0.13 | 0.21 | -0.03 | 0.15 | 0.28 | 0.08 | 0.21 | 0.18 | 0.19 | 0.19 |
| ## DIS33 | 0.04 | 0.08 | 0.13 | 0.07 | -0.04 | 0.09 | 0.23 | 0.14 | 0.10 | 0.21 | 0.12 | 0.02 |
| ## DIS36 | 0.00 | 0.09 | 0.11 | 0.10 | -0.02 | 0.08 | 0.17 | 0.09 | 0.08 | 0.20 | 0.12 | 0.04 |
| ## DIS39 | 0.01 | 0.09 | 0.18 | 0.12 | -0.06 | 0.16 | 0.33 | 0.10 | 0.09 | 0.31 | 0.14 | 0.03 |
| ## DIS42 | 0.15 | 0.14 | 0.17 | 0.16 | -0.01 | 0.19 | 0.33 | 0.08 | 0.17 | 0.35 | 0.16 | 0.14 |
| ## DIS45 | 0.05 | 0.16 | 0.18 | 0.17 | -0.07 | 0.10 | 0.18 | 0.10 | 0.11 | 0.21 | 0.19 | 0.01 |
| ## DIS48 | 0.00 | 0.12 | 0.08 | 0.10 | -0.06 | 0.08 | 0.19 | 0.06 | 0.08 | 0.24 | 0.13 | 0.02 |
| ## DIS51 | 0.03 | 0.14 | 0.16 | 0.15 | -0.07 | 0.09 | 0.28 | 0.09 | 0.10 | 0.24 | 0.20 | 0.09 |
| ## DIS54 | 0.01 | 0.12 | 0.07 | 0.09 | -0.04 | 0.10 | 0.13 | 0.01 | 0.04 | 0.13 | 0.12 | 0.02 |
| ## DIS57 | 0.01 | 0.12 | 0.14 | 0.13 | -0.08 | 0.10 | 0.32 | 0.19 | 0.11 | 0.34 | 0.16 | 0.03 |
| ## DIS60 | 0.05 | 0.17 | 0.19 | 0.12 | -0.04 | 0.18 | 0.28 | 0.09 | 0.08 | 0.29 | 0.15 | 0.02 |
| ## DIS63 | 0.04 | 0.14 | 0.11 | 0.11 | -0.05 | 0.01 | 0.15 | 0.05 | 0.06 | 0.18 | 0.14 | -0.01 |
| ## DIS66 | 0.14 | 0.07 | 0.12 | 0.05 | -0.04 | 0.05 | 0.20 | 0.14 | 0.09 | 0.14 | 0.17 | 0.03 |
| ## DIS69 | 0.04 | 0.16 | 0.16 | 0.12 | -0.04 | 0.12 | 0.23 | 0.08 | 0.10 | 0.15 | 0.16 | 0.06 |
| ## DIS72 | 0.05 | 0.15 | 0.17 | 0.13 | -0.07 | 0.13 | 0.32 | 0.16 | 0.11 | 0.30 | 0.18 | 0.07 |
| ## DIS75 | 0.02 | 0.06 | 0.08 | 0.03 | -0.04 | 0.06 | 0.11 | 0.07 | 0.02 | 0.04 | 0.07 | 0.00 |
| ## | NEG73 | NEG76 | POS02 | POS05 | POS08 | POS11 | POS14 | POS17 | POS20 | POS23 | POS26 | POS29 |
| ## NEG01 | 0.23 | 0.10 | 0.11 | 0.03 | 0.11 | 0.05 | 0.03 | 0.11 | 0.05 | 0.11 | 0.07 | 0.04 |
| ## NEG04 | 0.06 | 0.09 | 0.01 | -0.01 | 0.01 | 0.04 | 0.01 | -0.06 | 0.02 | 0.00 | -0.01 | -0.02 |
| ## NEG07 | 0.11 | 0.16 | 0.05 | 0.04 | 0.08 | 0.02 | 0.09 | 0.07 | 0.03 | -0.05 | -0.01 | 0.01 |
| ## NEG10 | 0.25 | 0.24 | 0.10 | 0.07 | 0.09 | 0.14 | 0.09 | 0.04 | 0.01 | 0.01 | 0.01 | 0.04 |
| ## NEG13 | 0.25 | 0.06 | 0.10 | 0.08 | 0.12 | 0.24 | -0.02 | 0.06 | 0.10 | 0.10 | 0.09 | 0.07 |
| ## NEG16 | 0.27 | 0.06 | 0.07 | 0.18 | 0.07 | 0.08 | 0.17 | 0.18 | 0.07 | 0.12 | 0.17 | 0.09 |
| ## NEG19 | 0.07 | 0.12 | 0.07 | 0.03 | 0.07 | 0.00 | 0.03 | -0.02 | 0.09 | 0.05 | 0.03 | 0.05 |
| ## NEG22 | 0.12 | 0.11 | 0.06 | 0.02 | 0.03 | 0.06 | 0.03 | -0.02 | 0.03 | -0.02 | 0.06 | 0.01 |
| ## NEG25 | 0.54 | 0.21 | 0.05 | 0.06 | 0.16 | 0.19 | 0.10 | 0.17 | 0.06 | 0.01 | 0.09 | 0.10 |
| ## NEG28 | 0.17 | 0.21 | 0.03 | 0.07 | -0.02 | 0.13 | 0.09 | 0.04 | -0.02 | 0.13 | 0.10 | 0.01 |
| ## NEG31 | 0.23 | 0.22 | 0.00 | -0.04 | -0.01 | 0.04 | 0.03 | 0.02 | -0.01 | 0.07 | -0.01 | 0.00 |
| ## NEG34 | 0.12 | 0.25 | 0.05 | 0.01 | 0.03 | 0.06 | 0.04 | 0.00 | 0.04 | 0.00 | 0.01 | 0.01 |
| ## NEG37 | 0.18 | 0.24 | 0.01 | 0.04 | 0.06 | 0.01 | 0.13 | 0.07 | -0.01 | 0.02 | 0.04 | 0.02 |
| ## NEG40 | 0.21 | 0.09 | 0.10 | 0.12 | 0.10 | 0.17 | 0.10 | 0.05 | 0.08 | 0.05 | 0.07 | 0.03 |
| ## NEG43 | 0.30 | 0.19 | 0.07 | 0.11 | 0.14 | 0.16 | 0.13 | 0.01 | 0.10 | 0.04 | 0.07 | 0.01 |

|  |  |  |  |  |  |  |  |  |  |  |  |  |  |
| --- | --- | --- | --- | --- | --- | --- | --- | --- | --- | --- | --- | --- | --- |
| ## | NEG46 | 0.24 | 0.12 | 0.05 | 0.05 | 0.06 | 0.15 | 0.05 | 0.01 | 0.00 | 0.01 | 0.09 | 0.00 |
| ## | NEG49 | 0.02 | 0.06 | -0.05 | 0.00 | 0.01 | -0.04 | 0.03 | -0.10 | -0.02 | -0.04 | -0.05 | -0.07 |
| ## | NEG52 | 0.25 | 0.22 | 0.09 | 0.13 | 0.13 | 0.10 | 0.15 | 0.13 | 0.12 | 0.07 | 0.13 | 0.10 |
| ## | NEG55 | 0.62 | 0.24 | 0.08 | 0.10 | 0.04 | 0.25 | 0.10 | 0.20 | 0.12 | 0.01 | 0.10 | 0.08 |
| ## | NEG58 | 0.19 | 0.13 | 0.08 | 0.04 | 0.04 | 0.15 | 0.02 | 0.05 | 0.03 | 0.05 | 0.02 | 0.02 |
| ## | NEG61 | 0.27 | 0.11 | 0.07 | 0.07 | 0.14 | 0.08 | 0.09 | 0.11 | 0.09 | 0.04 | 0.11 | 0.13 |
| ## | NEG64 | 0.64 | 0.22 | 0.06 | 0.04 | 0.11 | 0.16 | 0.06 | 0.09 | 0.04 | 0.05 | 0.03 | 0.08 |
| ## | NEG67 | 0.33 | 0.14 | 0.11 | 0.14 | 0.12 | 0.13 | 0.13 | 0.14 | 0.02 | 0.08 | 0.12 | 0.07 |
| ## | NEG70 | 0.26 | 0.13 | 0.06 | 0.03 | 0.16 | 0.07 | 0.11 | 0.04 | 0.04 | 0.04 | 0.07 | 0.02 |
| ## | NEG73 | 1.00 | 0.23 | 0.11 | 0.11 | 0.12 | 0.23 | 0.13 | 0.20 | 0.06 | 0.11 | 0.07 | 0.11 |
| ## | NEG76 | 0.23 | 1.00 | 0.04 | 0.01 | -0.01 | 0.01 | 0.07 | 0.06 | 0.00 | 0.02 | 0.04 | -0.01 |
| ## | POS02 | 0.11 | 0.04 | 1.00 | 0.25 | 0.27 | 0.24 | 0.08 | 0.25 | 0.26 | 0.16 | 0.19 | 0.18 |
| ## | POS05 | 0.11 | 0.01 | 0.25 | 1.00 | 0.03 | 0.16 | 0.12 | 0.21 | 0.24 | 0.11 | 0.14 | 0.19 |
| ## | POS08 | 0.12 | -0.01 | 0.27 | 0.03 | 1.00 | 0.14 | 0.09 | 0.08 | 0.12 | 0.08 | 0.09 | 0.16 |
| ## | POS11 | 0.23 | 0.01 | 0.24 | 0.16 | 0.14 | 1.00 | 0.14 | 0.17 | 0.23 | 0.17 | 0.22 | 0.08 |
| ## | POS14 | 0.13 | 0.07 | 0.08 | 0.12 | 0.09 | 0.14 | 1.00 | 0.12 | 0.11 | 0.12 | 0.15 | 0.10 |
| ## | POS17 | 0.20 | 0.06 | 0.25 | 0.21 | 0.08 | 0.17 | 0.12 | 1.00 | 0.12 | 0.16 | 0.22 | 0.20 |
| ## | POS20 | 0.06 | 0.00 | 0.26 | 0.24 | 0.12 | 0.23 | 0.11 | 0.12 | 1.00 | 0.12 | 0.26 | 0.34 |
| ## | POS23 | 0.11 | 0.02 | 0.16 | 0.11 | 0.08 | 0.17 | 0.12 | 0.16 | 0.12 | 1.00 | 0.18 | 0.20 |
| ## | POS26 | 0.07 | 0.04 | 0.19 | 0.14 | 0.09 | 0.22 | 0.15 | 0.22 | 0.26 | 0.18 | 1.00 | 0.25 |
| ## | POS29 | 0.11 | -0.01 | 0.18 | 0.19 | 0.16 | 0.08 | 0.10 | 0.20 | 0.34 | 0.20 | 0.25 | 1.00 |
| ## | POS32 | 0.18 | 0.08 | 0.24 | 0.17 | -0.01 | 0.26 | 0.14 | 0.26 | 0.07 | 0.26 | 0.18 | 0.14 |
| ## | POS35 | 0.15 | 0.06 | 0.11 | 0.04 | 0.17 | 0.22 | 0.11 | 0.04 | 0.18 | 0.10 | 0.14 | 0.19 |
| ## | POS38 | 0.13 | 0.03 | 0.23 | -0.03 | 0.19 | 0.20 | 0.06 | 0.12 | 0.11 | 0.18 | 0.12 | 0.10 |
| ## | POS41 | 0.27 | 0.11 | 0.17 | 0.10 | 0.09 | 0.24 | 0.09 | 0.21 | 0.21 | 0.18 | 0.17 | 0.19 |
| ## | POS44 | 0.15 | 0.04 | 0.14 | 0.21 | 0.12 | 0.19 | 0.06 | 0.14 | 0.28 | 0.21 | 0.23 | 0.35 |
| ## | POS47 | 0.16 | 0.03 | 0.13 | 0.06 | 0.18 | 0.25 | 0.17 | 0.10 | 0.22 | 0.06 | 0.27 | 0.14 |
| ## | POS50 | 0.28 | 0.06 | 0.23 | 0.10 | 0.12 | 0.41 | 0.12 | 0.14 | 0.15 | 0.19 | 0.14 | 0.12 |
| ## | POS53 | 0.18 | 0.00 | 0.11 | 0.20 | -0.01 | 0.20 | 0.18 | 0.25 | 0.10 | 0.39 | 0.19 | 0.21 |
| ## | POS56 | 0.17 | 0.07 | 0.22 | 0.25 | 0.13 | 0.19 | 0.23 | 0.25 | 0.19 | 0.20 | 0.16 | 0.22 |
| ## | POS59 | 0.24 | 0.04 | 0.08 | 0.17 | 0.04 | 0.12 | 0.10 | 0.27 | 0.13 | 0.27 | 0.17 | 0.19 |
| ## | POS62 | 0.18 | 0.07 | 0.21 | 0.21 | 0.18 | 0.25 | 0.16 | 0.25 | 0.37 | 0.17 | 0.33 | 0.37 |
| ## | POS65 | 0.11 | 0.05 | 0.32 | -0.02 | 0.53 | 0.20 | 0.09 | 0.16 | 0.10 | 0.25 | 0.15 | 0.11 |
| ## | POS68 | 0.13 | 0.03 | 0.40 | 0.17 | 0.20 | 0.37 | 0.21 | 0.19 | 0.23 | 0.05 | 0.16 | 0.14 |
| ## | POS71 | 0.13 | 0.05 | 0.22 | 0.15 | 0.07 | 0.19 | 0.22 | 0.19 | 0.20 | 0.24 | 0.23 | 0.22 |
| ## | POS74 | 0.16 | 0.05 | 0.25 | 0.11 | 0.22 | 0.30 | 0.20 | 0.23 | 0.18 | 0.28 | 0.22 | 0.26 |
| ## | POS77 | 0.16 | 0.02 | 0.12 | 0.14 | 0.08 | 0.17 | 0.12 | 0.22 | 0.12 | 0.16 | 0.18 | 0.17 |
| ## | DIS03 | 0.19 | 0.06 | 0.12 | 0.10 | 0.25 | 0.16 | 0.01 | 0.07 | 0.16 | 0.07 | 0.12 | 0.12 |
| ## | DIS06 | 0.28 | 0.05 | 0.20 | 0.08 | 0.22 | 0.21 | 0.10 | 0.20 | 0.10 | 0.20 | 0.15 | 0.13 |
| ## | DIS09 | 0.29 | 0.03 | 0.19 | 0.06 | 0.19 | 0.23 | 0.03 | 0.16 | 0.14 | 0.10 | 0.15 | 0.14 |
| ## | DIS12 | 0.15 | 0.06 | 0.13 | 0.01 | 0.07 | 0.18 | 0.00 | 0.09 | 0.10 | 0.11 | 0.12 | 0.13 |
| ## | DIS15 | 0.28 | 0.02 | 0.11 | 0.08 | 0.10 | 0.19 | 0.02 | 0.13 | 0.11 | 0.13 | 0.07 | 0.06 |
| ## | DIS18 | 0.23 | 0.07 | 0.20 | 0.03 | 0.17 | 0.16 | 0.04 | 0.09 | 0.15 | 0.09 | 0.13 | 0.11 |
| ## | DIS21 | 0.39 | 0.09 | 0.10 | 0.11 | 0.05 | 0.22 | 0.12 | 0.19 | 0.12 | 0.19 | 0.12 | 0.17 |
| ## | DIS24 | 0.17 | 0.05 | 0.12 | 0.04 | 0.08 | 0.20 | 0.05 | 0.09 | 0.15 | 0.11 | 0.12 | 0.13 |
| ## | DIS27 | 0.32 | 0.05 | 0.15 | 0.05 | 0.17 | 0.13 | 0.02 | 0.17 | 0.18 | 0.09 | 0.13 | 0.16 |
| ## | DIS30 | 0.24 | 0.01 | 0.15 | 0.14 | 0.14 | 0.30 | 0.13 | 0.18 | 0.17 | 0.11 | 0.24 | 0.14 |
| ## | DIS33 | 0.32 | 0.04 | 0.15 | 0.07 | 0.13 | 0.20 | -0.02 | 0.12 | 0.11 | 0.07 | 0.10 | 0.12 |
| ## | DIS36 | 0.22 | 0.04 | 0.09 | 0.07 | 0.16 | 0.15 | 0.07 | 0.08 | 0.15 | 0.08 | 0.10 | 0.12 |
| ## | DIS39 | 0.44 | 0.05 | 0.17 | 0.08 | 0.23 | 0.23 | -0.02 | 0.14 | 0.13 | 0.14 | 0.10 | 0.11 |
| ## | DIS42 | 0.36 | 0.14 | 0.15 | 0.07 | 0.25 | 0.30 | 0.05 | 0.15 | 0.08 | 0.15 | 0.16 | 0.13 |
| ## | DIS45 | 0.28 | 0.05 | 0.20 | 0.08 | 0.22 | 0.28 | 0.02 | 0.16 | 0.18 | 0.13 | 0.13 | 0.15 |
| ## | DIS48 | 0.26 | 0.04 | 0.12 | 0.04 | 0.12 | 0.13 | -0.03 | 0.11 | 0.11 | 0.11 | 0.11 | 0.18 |
| ## | DIS51 | 0.38 | 0.03 | 0.13 | 0.10 | 0.25 | 0.28 | 0.07 | 0.19 | 0.16 | 0.19 | 0.11 | 0.20 |

|  |  |  |  |  |  |  |  |  |  |  |  |  |  |
| --- | --- | --- | --- | --- | --- | --- | --- | --- | --- | --- | --- | --- | --- |
| ## | DIS54 | 0.14 | 0.04 | 0.01 | 0.03 | 0.03 | 0.13 | 0.00 | 0.03 | 0.11 | 0.05 | 0.03 | 0.10 |
| ## | DIS57 | 0.43 | 0.05 | 0.21 | 0.08 | 0.22 | 0.30 | 0.04 | 0.18 | 0.14 | 0.08 | 0.09 | 0.11 |
| ## | DIS60 | 0.37 | 0.04 | 0.16 | 0.05 | 0.21 | 0.25 | 0.04 | 0.08 | 0.11 | 0.17 | 0.10 | 0.10 |
| ## | DIS63 | 0.21 | 0.10 | 0.04 | 0.04 | 0.07 | 0.12 | 0.00 | 0.05 | 0.10 | 0.08 | 0.06 | 0.10 |
| ## | DIS66 | 0.25 | 0.01 | 0.06 | 0.03 | 0.15 | 0.15 | -0.02 | 0.14 | 0.07 | 0.09 | 0.04 | 0.12 |
| ## | DIS69 | 0.25 | 0.04 | 0.22 | 0.10 | 0.13 | 0.24 | 0.14 | 0.26 | 0.18 | 0.21 | 0.15 | 0.14 |
| ## | DIS72 | 0.46 | 0.08 | 0.17 | 0.08 | 0.22 | 0.30 | 0.15 | 0.18 | 0.10 | 0.18 | 0.13 | 0.13 |
| ## | DIS75 | 0.09 | 0.17 | 0.04 | 0.09 | 0.01 | 0.08 | 0.09 | 0.10 | 0.08 | 0.05 | 0.09 | 0.01 |
| ## |  | POS32 | POS35 | POS38 | POS41 | POS44 | POS47 | POS50 | POS53 | POS56 | POS59 | POS62 | POS65 |
| ## | NEG01 | 0.03 | 0.02 | 0.19 | 0.09 | 0.05 | 0.09 | 0.07 | 0.04 | 0.06 | 0.09 | 0.13 | 0.14 |
| ## | NEG04 | 0.00 | 0.08 | 0.03 | 0.03 | 0.02 | 0.00 | 0.03 | 0.03 | 0.01 | 0.00 | 0.02 | 0.04 |
| ## | NEG07 | 0.01 | 0.00 | 0.04 | 0.00 | 0.00 | 0.00 | 0.02 | -0.01 | 0.03 | 0.00 | 0.01 | 0.04 |
| ## | NEG10 | 0.01 | 0.07 | 0.09 | 0.11 | 0.02 | 0.06 | 0.11 | 0.10 | 0.09 | 0.06 | 0.07 | 0.13 |
| ## | NEG13 | 0.08 | 0.10 | 0.16 | 0.10 | 0.12 | 0.15 | 0.14 | 0.16 | 0.13 | 0.17 | 0.16 | 0.17 |
| ## | NEG16 | 0.11 | 0.05 | 0.10 | 0.08 | 0.08 | 0.19 | 0.14 | 0.24 | 0.12 | 0.14 | 0.19 | 0.02 |
| ## | NEG19 | 0.02 | 0.06 | 0.03 | 0.07 | 0.05 | 0.07 | 0.03 | 0.06 | 0.09 | 0.08 | 0.08 | 0.11 |
| ## | NEG22 | 0.01 | 0.04 | 0.05 | 0.03 | 0.06 | 0.03 | 0.01 | -0.06 | 0.06 | 0.01 | 0.06 | 0.10 |
| ## | NEG25 | 0.07 | 0.16 | 0.10 | 0.20 | 0.12 | 0.16 | 0.21 | 0.14 | 0.14 | 0.08 | 0.17 | 0.09 |
| ## | NEG28 | 0.09 | 0.09 | 0.06 | 0.09 | 0.04 | 0.04 | 0.22 | 0.16 | 0.12 | 0.13 | 0.03 | 0.05 |
| ## | NEG31 | 0.03 | 0.06 | 0.08 | 0.04 | 0.02 | 0.00 | 0.06 | 0.00 | 0.04 | -0.01 | 0.00 | 0.11 |
| ## | NEG34 | 0.04 | 0.03 | 0.07 | 0.05 | 0.01 | 0.09 | 0.06 | 0.01 | 0.05 | 0.03 | 0.03 | 0.11 |
| ## | NEG37 | 0.03 | -0.02 | 0.08 | -0.01 | 0.03 | 0.07 | 0.00 | 0.08 | 0.08 | 0.02 | 0.15 | 0.05 |
| ## | NEG40 | 0.10 | 0.08 | 0.10 | 0.18 | 0.08 | 0.12 | 0.17 | 0.10 | 0.18 | 0.12 | 0.11 | 0.14 |
| ## | NEG43 | 0.10 | 0.07 | 0.17 | 0.15 | 0.10 | 0.11 | 0.13 | 0.10 | 0.20 | 0.09 | 0.11 | 0.13 |
| ## | NEG46 | 0.09 | 0.06 | 0.11 | 0.10 | 0.04 | 0.11 | 0.09 | 0.03 | 0.07 | 0.09 | 0.11 | 0.13 |
| ## | NEG49 | -0.06 | -0.06 | 0.02 | -0.06 | 0.00 | 0.02 | -0.07 | -0.04 | 0.00 | -0.04 | -0.05 | 0.02 |
| ## | NEG52 | 0.05 | 0.10 | 0.08 | 0.10 | 0.11 | 0.05 | 0.09 | 0.09 | 0.13 | 0.14 | 0.15 | 0.17 |
| ## | NEG55 | 0.06 | 0.12 | 0.06 | 0.22 | 0.11 | 0.15 | 0.34 | 0.17 | 0.11 | 0.17 | 0.15 | 0.04 |
| ## | NEG58 | 0.06 | 0.01 | 0.06 | 0.10 | 0.04 | 0.04 | 0.15 | 0.08 | 0.14 | 0.05 | 0.03 | 0.09 |
| ## | NEG61 | 0.01 | 0.11 | 0.13 | 0.11 | 0.10 | 0.03 | 0.15 | 0.12 | 0.12 | 0.08 | 0.16 | 0.13 |
| ## | NEG64 | 0.11 | 0.13 | 0.11 | 0.20 | 0.10 | 0.10 | 0.24 | 0.12 | 0.09 | 0.08 | 0.11 | 0.10 |
| ## | NEG67 | 0.07 | 0.02 | 0.16 | 0.14 | 0.07 | 0.14 | 0.14 | 0.18 | 0.18 | 0.17 | 0.18 | 0.15 |
| ## | NEG70 | -0.02 | 0.04 | 0.17 | 0.03 | 0.02 | 0.05 | 0.09 | -0.02 | 0.06 | 0.04 | 0.10 | 0.15 |
| ## | NEG73 | 0.18 | 0.15 | 0.13 | 0.27 | 0.15 | 0.16 | 0.28 | 0.18 | 0.17 | 0.24 | 0.18 | 0.11 |
| ## | NEG76 | 0.08 | 0.06 | 0.03 | 0.11 | 0.04 | 0.03 | 0.06 | 0.00 | 0.07 | 0.04 | 0.07 | 0.05 |
| ## | POS02 | 0.24 | 0.11 | 0.23 | 0.17 | 0.14 | 0.13 | 0.23 | 0.11 | 0.22 | 0.08 | 0.21 | 0.32 |
| ## | POS05 | 0.17 | 0.04 | -0.03 | 0.10 | 0.21 | 0.06 | 0.10 | 0.20 | 0.25 | 0.17 | 0.21 | -0.02 |
| ## | POS08 | -0.01 | 0.17 | 0.19 | 0.09 | 0.12 | 0.18 | 0.12 | -0.01 | 0.13 | 0.04 | 0.18 | 0.53 |
| ## | POS11 | 0.26 | 0.22 | 0.20 | 0.24 | 0.19 | 0.25 | 0.41 | 0.20 | 0.19 | 0.12 | 0.25 | 0.20 |
| ## | POS14 | 0.14 | 0.11 | 0.06 | 0.09 | 0.06 | 0.17 | 0.12 | 0.18 | 0.23 | 0.10 | 0.16 | 0.09 |
| ## | POS17 | 0.26 | 0.04 | 0.12 | 0.21 | 0.14 | 0.10 | 0.14 | 0.25 | 0.25 | 0.27 | 0.25 | 0.16 |
| ## | POS20 | 0.07 | 0.18 | 0.11 | 0.21 | 0.28 | 0.22 | 0.15 | 0.10 | 0.19 | 0.13 | 0.37 | 0.10 |
| ## | POS23 | 0.26 | 0.10 | 0.18 | 0.18 | 0.21 | 0.06 | 0.19 | 0.39 | 0.20 | 0.27 | 0.17 | 0.25 |
| ## | POS26 | 0.18 | 0.14 | 0.12 | 0.17 | 0.23 | 0.27 | 0.14 | 0.19 | 0.16 | 0.17 | 0.33 | 0.15 |
| ## | POS29 | 0.14 | 0.19 | 0.10 | 0.19 | 0.35 | 0.14 | 0.12 | 0.21 | 0.22 | 0.19 | 0.37 | 0.11 |
| ## | POS32 | 1.00 | -0.01 | 0.06 | 0.25 | 0.12 | 0.17 | 0.27 | 0.30 | 0.24 | 0.28 | 0.17 | 0.09 |
| ## | POS35 | -0.01 | 1.00 | 0.05 | 0.10 | 0.18 | 0.27 | 0.18 | 0.16 | 0.15 | 0.19 | 0.14 | 0.07 |
| ## | POS38 | 0.06 | 0.05 | 1.00 | 0.20 | 0.11 | 0.21 | 0.16 | 0.23 | 0.13 | 0.10 | 0.22 | 0.47 |
| ## | POS41 | 0.25 | 0.10 | 0.20 | 1.00 | 0.24 | 0.23 | 0.35 | 0.20 | 0.20 | 0.26 | 0.29 | 0.13 |
| ## | POS44 | 0.12 | 0.18 | 0.11 | 0.24 | 1.00 | 0.14 | 0.13 | 0.19 | 0.22 | 0.18 | 0.28 | 0.11 |
| ## | POS47 | 0.17 | 0.27 | 0.21 | 0.23 | 0.14 | 1.00 | 0.27 | 0.23 | 0.26 | 0.20 | 0.24 | 0.17 |
| ## | POS50 | 0.27 | 0.18 | 0.16 | 0.35 | 0.13 | 0.27 | 1.00 | 0.23 | 0.13 | 0.17 | 0.16 | 0.17 |
| ## | POS53 | 0.30 | 0.16 | 0.23 | 0.20 | 0.19 | 0.23 | 0.23 | 1.00 | 0.32 | 0.37 | 0.21 | 0.12 |
| ## | POS56 | 0.24 | 0.15 | 0.13 | 0.20 | 0.22 | 0.26 | 0.13 | 0.32 | 1.00 | 0.28 | 0.22 | 0.18 |

|  |  |  |  |  |  |  |  |  |  |  |  |  |
| --- | --- | --- | --- | --- | --- | --- | --- | --- | --- | --- | --- | --- |
| ## POS59 | 0.28 | 0.19 | 0.10 | 0.26 | 0.18 | 0.20 | 0.17 | 0.37 | 0.28 | 1.00 | 0.18 | 0.09 |
| ## POS62 | 0.17 | 0.14 | 0.22 | 0.29 | 0.28 | 0.24 | 0.16 | 0.21 | 0.22 | 0.18 | 1.00 | 0.16 |
| ## POS65 | 0.09 | 0.07 | 0.47 | 0.13 | 0.11 | 0.17 | 0.17 | 0.12 | 0.18 | 0.09 | 0.16 | 1.00 |
| ## POS68 | 0.30 | 0.12 | 0.14 | 0.25 | 0.15 | 0.22 | 0.32 | 0.14 | 0.19 | 0.11 | 0.17 | 0.19 |
| ## POS71 | 0.22 | 0.08 | 0.16 | 0.15 | 0.15 | 0.16 | 0.23 | 0.30 | 0.25 | 0.20 | 0.30 | 0.06 |
| ## POS74 | 0.33 | 0.18 | 0.15 | 0.31 | 0.16 | 0.15 | 0.29 | 0.17 | 0.17 | 0.22 | 0.26 | 0.21 |
| ## POS77 | 0.33 | 0.16 | 0.12 | 0.28 | 0.12 | 0.23 | 0.27 | 0.35 | 0.25 | 0.31 | 0.17 | 0.08 |
| ## DIS03 | 0.09 | 0.24 | 0.09 | 0.22 | 0.13 | 0.18 | 0.25 | 0.09 | 0.12 | 0.15 | 0.15 | 0.23 |
| ## DIS06 | 0.24 | 0.05 | 0.14 | 0.19 | 0.15 | 0.21 | 0.31 | 0.26 | 0.22 | 0.15 | 0.13 | 0.20 |
| ## DIS09 | 0.13 | 0.13 | 0.21 | 0.31 | 0.16 | 0.23 | 0.26 | 0.19 | 0.16 | 0.17 | 0.18 | 0.18 |
| ## DIS12 | 0.16 | 0.10 | 0.10 | 0.21 | 0.13 | 0.23 | 0.20 | 0.16 | 0.16 | 0.19 | 0.12 | 0.10 |
| ## DIS15 | 0.20 | 0.05 | 0.15 | 0.20 | 0.11 | 0.21 | 0.21 | 0.26 | 0.17 | 0.18 | 0.12 | 0.21 |
| ## DIS18 | 0.11 | 0.18 | 0.14 | 0.25 | 0.19 | 0.24 | 0.24 | 0.15 | 0.17 | 0.14 | 0.18 | 0.20 |
| ## DIS21 | 0.28 | 0.14 | 0.17 | 0.33 | 0.16 | 0.24 | 0.33 | 0.27 | 0.17 | 0.23 | 0.17 | 0.11 |
| ## DIS24 | 0.12 | 0.16 | 0.11 | 0.23 | 0.15 | 0.21 | 0.22 | 0.21 | 0.12 | 0.19 | 0.13 | 0.10 |
| ## DIS27 | 0.11 | 0.17 | 0.15 | 0.30 | 0.15 | 0.22 | 0.24 | 0.09 | 0.12 | 0.16 | 0.16 | 0.16 |
| ## DIS30 | 0.17 | 0.17 | 0.24 | 0.25 | 0.19 | 0.23 | 0.32 | 0.23 | 0.22 | 0.19 | 0.30 | 0.13 |
| ## DIS33 | 0.20 | 0.21 | 0.14 | 0.26 | 0.12 | 0.20 | 0.23 | 0.15 | 0.18 | 0.20 | 0.15 | 0.12 |
| ## DIS36 | 0.17 | 0.14 | 0.16 | 0.21 | 0.12 | 0.25 | 0.22 | 0.18 | 0.13 | 0.17 | 0.17 | 0.15 |
| ## DIS39 | 0.16 | 0.23 | 0.15 | 0.26 | 0.11 | 0.23 | 0.30 | 0.17 | 0.17 | 0.20 | 0.11 | 0.21 |
| ## DIS42 | 0.24 | 0.15 | 0.23 | 0.26 | 0.13 | 0.18 | 0.37 | 0.24 | 0.19 | 0.22 | 0.13 | 0.32 |
| ## DIS45 | 0.28 | 0.20 | 0.19 | 0.28 | 0.12 | 0.26 | 0.23 | 0.23 | 0.22 | 0.20 | 0.23 | 0.15 |
| ## DIS48 | 0.17 | 0.17 | 0.13 | 0.28 | 0.11 | 0.18 | 0.24 | 0.18 | 0.11 | 0.21 | 0.12 | 0.11 |
| ## DIS51 | 0.22 | 0.14 | 0.16 | 0.32 | 0.13 | 0.30 | 0.32 | 0.23 | 0.25 | 0.28 | 0.23 | 0.17 |
| ## DIS54 | 0.08 | 0.11 | 0.12 | 0.17 | 0.14 | 0.15 | 0.17 | 0.16 | 0.12 | 0.16 | 0.14 | 0.06 |
| ## DIS57 | 0.21 | 0.18 | 0.15 | 0.34 | 0.14 | 0.22 | 0.33 | 0.21 | 0.28 | 0.22 | 0.15 | 0.21 |
| ## DIS60 | 0.20 | 0.22 | 0.14 | 0.27 | 0.11 | 0.21 | 0.31 | 0.20 | 0.19 | 0.18 | 0.12 | 0.20 |
| ## DIS63 | 0.17 | 0.16 | 0.10 | 0.22 | 0.12 | 0.20 | 0.21 | 0.18 | 0.20 | 0.18 | 0.13 | 0.06 |
| ## DIS66 | 0.04 | 0.24 | 0.16 | 0.15 | 0.11 | 0.20 | 0.20 | 0.18 | 0.09 | 0.17 | 0.07 | 0.14 |
| ## DIS69 | 0.25 | 0.11 | 0.24 | 0.26 | 0.12 | 0.27 | 0.27 | 0.26 | 0.26 | 0.26 | 0.17 | 0.26 |
| ## DIS72 | 0.27 | 0.23 | 0.20 | 0.37 | 0.16 | 0.30 | 0.36 | 0.25 | 0.24 | 0.22 | 0.19 | 0.21 |
| ## DIS75 | 0.07 | 0.05 | 0.07 | 0.06 | 0.03 | 0.18 | 0.14 | 0.09 | 0.11 | 0.10 | 0.08 | 0.08 |
| ## POS68 | POS71 | POS74 | POS77 | DIS03 | DIS06 | DIS09 | DIS12 | DIS15 | DIS18 | DIS21 | DIS24 |  |
| ## NEG01 | 0.03 | 0.11 | 0.08 | 0.05 | 0.10 | 0.16 | 0.07 | 0.11 | 0.09 | 0.12 | 0.10 | 0.08 |
| ## NEG04 | 0.04 | 0.04 | 0.04 | 0.03 | 0.01 | 0.06 | 0.00 | 0.01 | 0.03 | 0.02 | -0.03 | -0.02 |
| ## NEG07 | 0.04 | 0.03 | -0.02 | -0.02 | 0.04 | 0.09 | 0.05 | 0.04 | 0.02 | 0.00 | 0.02 | -0.01 |
| ## NEG10 | 0.10 | 0.06 | 0.08 | 0.08 | 0.03 | 0.13 | 0.03 | 0.10 | 0.07 | 0.03 | 0.09 | 0.05 |
| ## NEG13 | 0.12 | 0.08 | 0.11 | 0.10 | 0.15 | 0.20 | 0.18 | 0.12 | 0.21 | 0.15 | 0.21 | 0.10 |
| ## NEG16 | 0.07 | 0.20 | 0.06 | 0.15 | 0.10 | 0.24 | 0.14 | 0.13 | 0.16 | 0.12 | 0.13 | 0.09 |
| ## NEG19 | 0.00 | 0.04 | 0.07 | 0.05 | 0.07 | 0.09 | 0.05 | 0.08 | 0.04 | 0.08 | 0.01 | 0.07 |
| ## NEG22 | 0.03 | 0.02 | 0.01 | 0.03 | 0.00 | 0.04 | -0.01 | -0.02 | -0.02 | 0.01 | 0.01 | -0.01 |
| ## NEG25 | 0.11 | 0.17 | 0.07 | 0.09 | 0.19 | 0.23 | 0.29 | 0.17 | 0.26 | 0.23 | 0.21 | 0.16 |
| ## NEG28 | 0.03 | 0.03 | 0.09 | 0.13 | 0.07 | 0.17 | 0.08 | 0.15 | 0.05 | 0.05 | 0.13 | 0.07 |
| ## NEG31 | -0.02 | -0.03 | 0.01 | 0.02 | 0.11 | 0.08 | 0.03 | 0.01 | 0.02 | 0.07 | 0.10 | 0.03 |
| ## NEG34 | 0.01 | 0.05 | 0.07 | 0.00 | 0.01 | 0.03 | -0.02 | 0.05 | 0.05 | 0.05 | 0.04 | 0.05 |
| ## NEG37 | 0.03 | 0.02 | -0.03 | -0.02 | 0.04 | 0.08 | 0.02 | 0.07 | 0.11 | 0.03 | 0.07 | 0.01 |
| ## NEG40 | 0.14 | 0.03 | 0.09 | 0.05 | 0.08 | 0.16 | 0.11 | 0.18 | 0.14 | 0.12 | 0.15 | 0.16 |
| ## NEG43 | 0.14 | 0.03 | 0.08 | 0.08 | 0.09 | 0.15 | 0.12 | 0.12 | 0.14 | 0.11 | 0.14 | 0.12 |
| ## NEG46 | 0.06 | 0.05 | 0.07 | 0.04 | 0.11 | 0.07 | 0.09 | 0.08 | 0.11 | 0.11 | 0.12 | 0.08 |
| ## NEG49 | -0.03 | -0.05 | -0.04 | -0.08 | -0.05 | -0.02 | -0.03 | 0.00 | -0.06 | -0.07 | -0.05 | -0.03 |
| ## NEG52 | 0.20 | 0.08 | 0.05 | 0.04 | 0.10 | 0.27 | 0.09 | 0.04 | 0.14 | 0.09 | 0.10 | 0.07 |
| ## NEG55 | 0.15 | 0.14 | 0.07 | 0.09 | 0.19 | 0.27 | 0.21 | 0.15 | 0.25 | 0.20 | 0.32 | 0.15 |
| ## NEG58 | 0.11 | 0.04 | 0.06 | 0.12 | 0.08 | 0.15 | 0.10 | 0.08 | 0.05 | 0.06 | 0.07 | 0.04 |
| ## NEG61 | 0.09 | 0.09 | 0.08 | 0.08 | 0.13 | 0.15 | 0.10 | 0.04 | 0.09 | 0.12 | 0.06 | 0.09 |

|  |  |  |  |  |  |  |  |  |  |  |  |  |  |
| --- | --- | --- | --- | --- | --- | --- | --- | --- | --- | --- | --- | --- | --- |
| ## | NEG64 | 0.07 | 0.08 | 0.04 | 0.09 | 0.18 | 0.21 | 0.22 | 0.16 | 0.21 | 0.17 | 0.28 | 0.18 |
| ## | NEG67 | 0.11 | 0.12 | 0.11 | 0.11 | 0.11 | 0.17 | 0.16 | 0.13 | 0.17 | 0.15 | 0.14 | 0.10 |
| ## | NEG70 | 0.11 | 0.03 | 0.07 | -0.02 | 0.03 | 0.04 | 0.01 | 0.03 | 0.01 | 0.02 | 0.05 | 0.02 |
| ## | NEG73 | 0.13 | 0.13 | 0.16 | 0.16 | 0.19 | 0.28 | 0.29 | 0.15 | 0.28 | 0.23 | 0.39 | 0.17 |
| ## | NEG76 | 0.03 | 0.05 | 0.05 | 0.02 | 0.06 | 0.05 | 0.03 | 0.06 | 0.02 | 0.07 | 0.09 | 0.05 |
| ## | POS02 | 0.40 | 0.22 | 0.25 | 0.12 | 0.12 | 0.20 | 0.19 | 0.13 | 0.11 | 0.20 | 0.10 | 0.12 |
| ## | POS05 | 0.17 | 0.15 | 0.11 | 0.14 | 0.10 | 0.08 | 0.06 | 0.01 | 0.08 | 0.03 | 0.11 | 0.04 |
| ## | POS08 | 0.20 | 0.07 | 0.22 | 0.08 | 0.25 | 0.22 | 0.19 | 0.07 | 0.10 | 0.17 | 0.05 | 0.08 |
| ## | POS11 | 0.37 | 0.19 | 0.30 | 0.17 | 0.16 | 0.21 | 0.23 | 0.18 | 0.19 | 0.16 | 0.22 | 0.20 |
| ## | POS14 | 0.21 | 0.22 | 0.20 | 0.12 | 0.01 | 0.10 | 0.03 | 0.00 | 0.02 | 0.04 | 0.12 | 0.05 |
| ## | POS17 | 0.19 | 0.19 | 0.23 | 0.22 | 0.07 | 0.20 | 0.16 | 0.09 | 0.13 | 0.09 | 0.19 | 0.09 |
| ## | POS20 | 0.23 | 0.20 | 0.18 | 0.12 | 0.16 | 0.10 | 0.14 | 0.10 | 0.11 | 0.15 | 0.12 | 0.15 |
| ## | POS23 | 0.05 | 0.24 | 0.28 | 0.16 | 0.07 | 0.20 | 0.10 | 0.11 | 0.13 | 0.09 | 0.19 | 0.11 |
| ## | POS26 | 0.16 | 0.23 | 0.22 | 0.18 | 0.12 | 0.15 | 0.15 | 0.12 | 0.07 | 0.13 | 0.12 | 0.12 |
| ## | POS29 | 0.14 | 0.22 | 0.26 | 0.17 | 0.12 | 0.13 | 0.14 | 0.13 | 0.06 | 0.11 | 0.17 | 0.13 |
| ## | POS32 | 0.30 | 0.22 | 0.33 | 0.33 | 0.09 | 0.24 | 0.13 | 0.16 | 0.20 | 0.11 | 0.28 | 0.12 |
| ## | POS35 | 0.12 | 0.08 | 0.18 | 0.16 | 0.24 | 0.05 | 0.13 | 0.10 | 0.05 | 0.18 | 0.14 | 0.16 |
| ## | POS38 | 0.14 | 0.16 | 0.15 | 0.12 | 0.09 | 0.14 | 0.21 | 0.10 | 0.15 | 0.14 | 0.17 | 0.11 |
| ## | POS41 | 0.25 | 0.15 | 0.31 | 0.28 | 0.22 | 0.19 | 0.31 | 0.21 | 0.20 | 0.25 | 0.33 | 0.23 |
| ## | POS44 | 0.15 | 0.15 | 0.16 | 0.12 | 0.13 | 0.15 | 0.16 | 0.13 | 0.11 | 0.19 | 0.16 | 0.15 |
| ## | POS47 | 0.22 | 0.16 | 0.15 | 0.23 | 0.18 | 0.21 | 0.23 | 0.23 | 0.21 | 0.24 | 0.24 | 0.21 |
| ## | POS50 | 0.32 | 0.23 | 0.29 | 0.27 | 0.25 | 0.31 | 0.26 | 0.20 | 0.21 | 0.24 | 0.33 | 0.22 |
| ## | POS53 | 0.14 | 0.30 | 0.17 | 0.35 | 0.09 | 0.26 | 0.19 | 0.16 | 0.26 | 0.15 | 0.27 | 0.21 |
| ## | POS56 | 0.19 | 0.25 | 0.17 | 0.25 | 0.12 | 0.22 | 0.16 | 0.16 | 0.17 | 0.17 | 0.17 | 0.12 |
| ## | POS59 | 0.11 | 0.20 | 0.22 | 0.31 | 0.15 | 0.15 | 0.17 | 0.19 | 0.18 | 0.14 | 0.23 | 0.19 |
| ## | POS62 | 0.17 | 0.30 | 0.26 | 0.17 | 0.15 | 0.13 | 0.18 | 0.12 | 0.12 | 0.18 | 0.17 | 0.13 |
| ## | POS65 | 0.19 | 0.06 | 0.21 | 0.08 | 0.23 | 0.20 | 0.18 | 0.10 | 0.21 | 0.20 | 0.11 | 0.10 |
| ## | POS68 | 1.00 | 0.16 | 0.33 | 0.12 | 0.09 | 0.15 | 0.16 | 0.11 | 0.15 | 0.15 | 0.18 | 0.16 |
| ## | POS71 | 0.16 | 1.00 | 0.16 | 0.14 | 0.09 | 0.20 | 0.16 | 0.08 | 0.14 | 0.13 | 0.16 | 0.06 |
| ## | POS74 | 0.33 | 0.16 | 1.00 | 0.28 | 0.20 | 0.13 | 0.26 | 0.17 | 0.16 | 0.25 | 0.30 | 0.17 |
| ## | POS77 | 0.12 | 0.14 | 0.28 | 1.00 | 0.07 | 0.20 | 0.13 | 0.09 | 0.13 | 0.12 | 0.15 | 0.13 |
| ## | DIS03 | 0.09 | 0.09 | 0.20 | 0.07 | 1.00 | 0.26 | 0.51 | 0.37 | 0.37 | 0.37 | 0.38 | 0.28 |
| ## | DIS06 | 0.15 | 0.20 | 0.13 | 0.20 | 0.26 | 1.00 | 0.33 | 0.22 | 0.37 | 0.26 | 0.27 | 0.18 |
| ## | DIS09 | 0.16 | 0.16 | 0.26 | 0.13 | 0.51 | 0.33 | 1.00 | 0.54 | 0.34 | 0.44 | 0.42 | 0.29 |
| ## | DIS12 | 0.11 | 0.08 | 0.17 | 0.09 | 0.37 | 0.22 | 0.54 | 1.00 | 0.24 | 0.32 | 0.29 | 0.31 |
| ## | DIS15 | 0.15 | 0.14 | 0.16 | 0.13 | 0.37 | 0.37 | 0.34 | 0.24 | 1.00 | 0.39 | 0.44 | 0.24 |
| ## | DIS18 | 0.15 | 0.13 | 0.25 | 0.12 | 0.37 | 0.26 | 0.44 | 0.32 | 0.39 | 1.00 | 0.34 | 0.33 |
| ## | DIS21 | 0.18 | 0.16 | 0.30 | 0.15 | 0.38 | 0.27 | 0.42 | 0.29 | 0.44 | 0.34 | 1.00 | 0.30 |
| ## | DIS24 | 0.16 | 0.06 | 0.17 | 0.13 | 0.28 | 0.18 | 0.29 | 0.31 | 0.24 | 0.33 | 0.30 | 1.00 |
| ## | DIS27 | 0.11 | 0.15 | 0.27 | 0.09 | 0.48 | 0.29 | 0.52 | 0.36 | 0.37 | 0.36 | 0.48 | 0.24 |
| ## | DIS30 | 0.21 | 0.27 | 0.29 | 0.14 | 0.34 | 0.33 | 0.47 | 0.35 | 0.29 | 0.41 | 0.31 | 0.22 |
| ## | DIS33 | 0.20 | 0.14 | 0.21 | 0.12 | 0.36 | 0.17 | 0.41 | 0.31 | 0.33 | 0.35 | 0.41 | 0.19 |
| ## | DIS36 | 0.14 | 0.17 | 0.16 | 0.14 | 0.43 | 0.27 | 0.34 | 0.29 | 0.35 | 0.39 | 0.38 | 0.31 |
| ## | DIS39 | 0.16 | 0.16 | 0.28 | 0.19 | 0.44 | 0.33 | 0.44 | 0.29 | 0.47 | 0.44 | 0.50 | 0.26 |
| ## | DIS42 | 0.18 | 0.18 | 0.22 | 0.15 | 0.32 | 0.30 | 0.33 | 0.20 | 0.34 | 0.35 | 0.51 | 0.22 |
| ## | DIS45 | 0.19 | 0.17 | 0.26 | 0.20 | 0.26 | 0.35 | 0.37 | 0.22 | 0.35 | 0.28 | 0.35 | 0.21 |
| ## | DIS48 | 0.07 | 0.14 | 0.24 | 0.14 | 0.36 | 0.19 | 0.40 | 0.34 | 0.31 | 0.33 | 0.47 | 0.38 |
| ## | DIS51 | 0.17 | 0.23 | 0.26 | 0.19 | 0.27 | 0.26 | 0.36 | 0.24 | 0.34 | 0.32 | 0.42 | 0.26 |
| ## | DIS54 | 0.05 | 0.13 | 0.12 | 0.08 | 0.32 | 0.22 | 0.32 | 0.29 | 0.28 | 0.30 | 0.31 | 0.41 |
| ## | DIS57 | 0.21 | 0.24 | 0.23 | 0.23 | 0.46 | 0.32 | 0.48 | 0.30 | 0.39 | 0.41 | 0.48 | 0.23 |
| ## | DIS60 | 0.15 | 0.11 | 0.30 | 0.13 | 0.38 | 0.31 | 0.39 | 0.29 | 0.34 | 0.36 | 0.50 | 0.20 |
| ## | DIS63 | 0.10 | 0.11 | 0.20 | 0.11 | 0.35 | 0.15 | 0.37 | 0.33 | 0.32 | 0.35 | 0.39 | 0.38 |
| ## | DIS66 | 0.10 | 0.07 | 0.16 | 0.14 | 0.27 | 0.20 | 0.33 | 0.28 | 0.31 | 0.26 | 0.32 | 0.21 |
| ## | DIS69 | 0.30 | 0.26 | 0.33 | 0.12 | 0.28 | 0.26 | 0.33 | 0.25 | 0.48 | 0.37 | 0.44 | 0.26 |

|  |  |  |  |  |  |  |  |  |  |  |  |  |  |
| --- | --- | --- | --- | --- | --- | --- | --- | --- | --- | --- | --- | --- | --- |
| ## | DIS72 | 0.27 | 0.20 | 0.40 | 0.18 | 0.43 | 0.32 | 0.50 | 0.30 | 0.42 | 0.41 | 0.63 | 0.26 |
| ## | DIS75 | 0.10 | 0.12 | 0.09 | 0.05 | 0.16 | 0.10 | 0.17 | 0.13 | 0.20 | 0.18 | 0.18 | 0.10 |
| ## | DIS27 | DIS30 | DIS33 | DIS36 | DIS39 | DIS42 | DIS45 | DIS48 | DIS51 | DIS54 | DIS57 | DIS60 |  |
| ## | NEG01 | 0.12 | 0.15 | 0.09 | 0.11 | 0.13 | 0.13 | 0.05 | 0.10 | 0.13 | 0.10 | 0.10 | 0.12 |
| ## | NEG04 | 0.01 | 0.03 | -0.01 | -0.01 | 0.05 | 0.04 | -0.02 | 0.00 | -0.03 | -0.01 | 0.02 | 0.01 |
| ## | NEG07 | 0.06 | 0.04 | 0.03 | 0.03 | 0.07 | 0.04 | 0.03 | 0.03 | 0.03 | -0.02 | 0.02 | 0.08 |
| ## | NEG10 | 0.03 | 0.11 | 0.07 | 0.08 | 0.09 | 0.14 | 0.09 | 0.04 | 0.11 | 0.03 | 0.08 | 0.11 |
| ## | NEG13 | 0.10 | 0.18 | 0.17 | 0.16 | 0.19 | 0.21 | 0.20 | 0.10 | 0.23 | 0.11 | 0.18 | 0.18 |
| ## | NEG16 | 0.11 | 0.26 | 0.05 | 0.11 | 0.17 | 0.11 | 0.16 | 0.11 | 0.17 | 0.04 | 0.09 | 0.08 |
| ## | NEG19 | 0.05 | 0.04 | 0.03 | 0.09 | 0.04 | 0.05 | -0.01 | 0.08 | 0.04 | 0.00 | 0.04 | 0.02 |
| ## | NEG22 | 0.01 | 0.09 | 0.01 | 0.03 | 0.08 | 0.07 | 0.02 | 0.01 | 0.01 | -0.04 | 0.03 | 0.05 |
| ## | NEG25 | 0.19 | 0.20 | 0.21 | 0.19 | 0.27 | 0.37 | 0.28 | 0.19 | 0.26 | 0.18 | 0.29 | 0.22 |
| ## | NEG28 | 0.03 | 0.15 | 0.04 | 0.03 | 0.05 | 0.12 | 0.10 | 0.15 | 0.04 | 0.11 | 0.07 | 0.09 |
| ## | NEG31 | 0.08 | 0.09 | 0.11 | 0.04 | 0.13 | 0.18 | 0.05 | 0.06 | 0.00 | 0.02 | 0.12 | 0.11 |
| ## | NEG34 | -0.01 | 0.05 | 0.01 | 0.04 | 0.02 | 0.11 | -0.03 | 0.06 | 0.06 | 0.00 | 0.04 | 0.01 |
| ## | NEG37 | 0.00 | 0.09 | 0.04 | 0.00 | 0.01 | 0.15 | 0.05 | 0.00 | 0.03 | 0.01 | 0.01 | 0.05 |
| ## | NEG40 | 0.01 | 0.18 | 0.08 | 0.09 | 0.09 | 0.14 | 0.16 | 0.12 | 0.14 | 0.12 | 0.12 | 0.17 |
| ## | NEG43 | 0.07 | 0.13 | 0.13 | 0.11 | 0.18 | 0.17 | 0.18 | 0.08 | 0.16 | 0.07 | 0.14 | 0.19 |
| ## | NEG46 | 0.05 | 0.21 | 0.07 | 0.10 | 0.12 | 0.16 | 0.17 | 0.10 | 0.15 | 0.09 | 0.13 | 0.12 |
| ## | NEG49 | -0.03 | -0.03 | -0.04 | -0.02 | -0.06 | -0.01 | -0.07 | -0.06 | -0.07 | -0.04 | -0.08 | -0.04 |
| ## | NEG52 | 0.13 | 0.15 | 0.09 | 0.08 | 0.16 | 0.19 | 0.10 | 0.08 | 0.09 | 0.10 | 0.10 | 0.18 |
| ## | NEG55 | 0.26 | 0.28 | 0.23 | 0.17 | 0.33 | 0.33 | 0.18 | 0.19 | 0.28 | 0.13 | 0.32 | 0.28 |
| ## | NEG58 | 0.06 | 0.08 | 0.14 | 0.09 | 0.10 | 0.08 | 0.10 | 0.06 | 0.09 | 0.01 | 0.19 | 0.09 |
| ## | NEG61 | 0.07 | 0.21 | 0.10 | 0.08 | 0.09 | 0.17 | 0.11 | 0.08 | 0.10 | 0.04 | 0.11 | 0.08 |
| ## | NEG64 | 0.22 | 0.18 | 0.21 | 0.20 | 0.31 | 0.35 | 0.21 | 0.24 | 0.24 | 0.13 | 0.34 | 0.29 |
| ## | NEG67 | 0.07 | 0.19 | 0.12 | 0.12 | 0.14 | 0.16 | 0.19 | 0.13 | 0.20 | 0.12 | 0.16 | 0.15 |
| ## | NEG70 | 0.01 | 0.19 | 0.02 | 0.04 | 0.03 | 0.14 | 0.01 | 0.02 | 0.09 | 0.02 | 0.03 | 0.02 |
| ## | NEG73 | 0.32 | 0.24 | 0.32 | 0.22 | 0.44 | 0.36 | 0.28 | 0.26 | 0.38 | 0.14 | 0.43 | 0.37 |
| ## | NEG76 | 0.05 | 0.01 | 0.04 | 0.04 | 0.05 | 0.14 | 0.05 | 0.04 | 0.03 | 0.04 | 0.05 | 0.04 |
| ## | POS02 | 0.15 | 0.15 | 0.15 | 0.09 | 0.17 | 0.15 | 0.20 | 0.12 | 0.13 | 0.01 | 0.21 | 0.16 |
| ## | POS05 | 0.05 | 0.14 | 0.07 | 0.07 | 0.08 | 0.07 | 0.08 | 0.04 | 0.10 | 0.03 | 0.08 | 0.05 |
| ## | POS08 | 0.17 | 0.14 | 0.13 | 0.16 | 0.23 | 0.25 | 0.22 | 0.12 | 0.25 | 0.03 | 0.22 | 0.21 |
| ## | POS11 | 0.13 | 0.30 | 0.20 | 0.15 | 0.23 | 0.30 | 0.28 | 0.13 | 0.28 | 0.13 | 0.30 | 0.25 |
| ## | POS14 | 0.02 | 0.13 | -0.02 | 0.07 | -0.02 | 0.05 | 0.02 | -0.03 | 0.07 | 0.00 | 0.04 | 0.04 |
| ## | POS17 | 0.17 | 0.18 | 0.12 | 0.08 | 0.14 | 0.15 | 0.16 | 0.11 | 0.19 | 0.03 | 0.18 | 0.08 |
| ## | POS20 | 0.18 | 0.17 | 0.11 | 0.15 | 0.13 | 0.08 | 0.18 | 0.11 | 0.16 | 0.11 | 0.14 | 0.11 |
| ## | POS23 | 0.09 | 0.11 | 0.07 | 0.08 | 0.14 | 0.15 | 0.13 | 0.11 | 0.19 | 0.05 | 0.08 | 0.17 |
| ## | POS26 | 0.13 | 0.24 | 0.10 | 0.10 | 0.10 | 0.16 | 0.13 | 0.11 | 0.11 | 0.03 | 0.09 | 0.10 |
| ## | POS29 | 0.16 | 0.14 | 0.12 | 0.12 | 0.11 | 0.13 | 0.15 | 0.18 | 0.20 | 0.10 | 0.11 | 0.10 |
| ## | POS32 | 0.11 | 0.17 | 0.20 | 0.17 | 0.16 | 0.24 | 0.28 | 0.17 | 0.22 | 0.08 | 0.21 | 0.20 |
| ## | POS35 | 0.17 | 0.17 | 0.21 | 0.14 | 0.23 | 0.15 | 0.20 | 0.17 | 0.14 | 0.11 | 0.18 | 0.22 |
| ## | POS38 | 0.15 | 0.24 | 0.14 | 0.16 | 0.15 | 0.23 | 0.19 | 0.13 | 0.16 | 0.12 | 0.15 | 0.14 |
| ## | POS41 | 0.30 | 0.25 | 0.26 | 0.21 | 0.26 | 0.26 | 0.28 | 0.28 | 0.32 | 0.17 | 0.34 | 0.27 |
| ## | POS44 | 0.15 | 0.19 | 0.12 | 0.12 | 0.11 | 0.13 | 0.12 | 0.11 | 0.13 | 0.14 | 0.14 | 0.11 |
| ## | POS47 | 0.22 | 0.23 | 0.20 | 0.25 | 0.23 | 0.18 | 0.26 | 0.18 | 0.30 | 0.15 | 0.22 | 0.21 |
| ## | POS50 | 0.24 | 0.32 | 0.23 | 0.22 | 0.30 | 0.37 | 0.23 | 0.24 | 0.32 | 0.17 | 0.33 | 0.31 |
| ## | POS53 | 0.09 | 0.23 | 0.15 | 0.18 | 0.17 | 0.24 | 0.23 | 0.18 | 0.23 | 0.16 | 0.21 | 0.20 |
| ## | POS56 | 0.12 | 0.22 | 0.18 | 0.13 | 0.17 | 0.19 | 0.22 | 0.11 | 0.25 | 0.12 | 0.28 | 0.19 |
| ## | POS59 | 0.16 | 0.19 | 0.20 | 0.17 | 0.20 | 0.22 | 0.20 | 0.21 | 0.28 | 0.16 | 0.22 | 0.18 |
| ## | POS62 | 0.16 | 0.30 | 0.15 | 0.17 | 0.11 | 0.13 | 0.23 | 0.12 | 0.23 | 0.14 | 0.15 | 0.12 |
| ## | POS65 | 0.16 | 0.13 | 0.12 | 0.15 | 0.21 | 0.32 | 0.15 | 0.11 | 0.17 | 0.06 | 0.21 | 0.20 |
| ## | POS68 | 0.11 | 0.21 | 0.20 | 0.14 | 0.16 | 0.18 | 0.19 | 0.07 | 0.17 | 0.05 | 0.21 | 0.15 |
| ## | POS71 | 0.15 | 0.27 | 0.14 | 0.17 | 0.16 | 0.18 | 0.17 | 0.14 | 0.23 | 0.13 | 0.24 | 0.11 |
| ## | POS74 | 0.27 | 0.29 | 0.21 | 0.16 | 0.28 | 0.22 | 0.26 | 0.24 | 0.26 | 0.12 | 0.23 | 0.30 |

|  |  |  |  |  |  |  |  |  |  |  |  |  |
| --- | --- | --- | --- | --- | --- | --- | --- | --- | --- | --- | --- | --- |
| ## POS77 | 0.09 | 0.14 | 0.12 | 0.14 | 0.19 | 0.15 | 0.20 | 0.14 | 0.19 | 0.08 | 0.23 | 0.13 |
| ## DIS03 | 0.48 | 0.34 | 0.36 | 0.43 | 0.44 | 0.32 | 0.26 | 0.36 | 0.27 | 0.32 | 0.46 | 0.38 |
| ## DIS06 | 0.29 | 0.33 | 0.17 | 0.27 | 0.33 | 0.30 | 0.35 | 0.19 | 0.26 | 0.22 | 0.32 | 0.31 |
| ## DIS09 | 0.52 | 0.47 | 0.41 | 0.34 | 0.44 | 0.33 | 0.37 | 0.40 | 0.36 | 0.32 | 0.48 | 0.39 |
| ## DIS12 | 0.36 | 0.35 | 0.31 | 0.29 | 0.29 | 0.20 | 0.22 | 0.34 | 0.24 | 0.29 | 0.30 | 0.29 |
| ## DIS15 | 0.37 | 0.29 | 0.33 | 0.35 | 0.47 | 0.34 | 0.35 | 0.31 | 0.34 | 0.28 | 0.39 | 0.34 |
| ## DIS18 | 0.36 | 0.41 | 0.35 | 0.39 | 0.44 | 0.35 | 0.28 | 0.33 | 0.32 | 0.30 | 0.41 | 0.36 |
| ## DIS21 | 0.48 | 0.31 | 0.41 | 0.38 | 0.50 | 0.51 | 0.35 | 0.47 | 0.42 | 0.31 | 0.48 | 0.50 |
| ## DIS24 | 0.24 | 0.22 | 0.19 | 0.31 | 0.26 | 0.22 | 0.21 | 0.38 | 0.26 | 0.41 | 0.23 | 0.20 |
| ## DIS27 | 1.00 | 0.37 | 0.37 | 0.34 | 0.66 | 0.35 | 0.32 | 0.39 | 0.30 | 0.22 | 0.48 | 0.52 |
| ## DIS30 | 0.37 | 1.00 | 0.34 | 0.28 | 0.35 | 0.30 | 0.37 | 0.30 | 0.39 | 0.24 | 0.40 | 0.30 |
| ## DIS33 | 0.37 | 0.34 | 1.00 | 0.33 | 0.46 | 0.38 | 0.38 | 0.35 | 0.30 | 0.25 | 0.53 | 0.47 |
| ## DIS36 | 0.34 | 0.28 | 0.33 | 1.00 | 0.39 | 0.24 | 0.30 | 0.37 | 0.28 | 0.40 | 0.38 | 0.34 |
| ## DIS39 | 0.66 | 0.35 | 0.46 | 0.39 | 1.00 | 0.47 | 0.43 | 0.45 | 0.41 | 0.23 | 0.59 | 0.73 |
| ## DIS42 | 0.35 | 0.30 | 0.38 | 0.24 | 0.47 | 1.00 | 0.33 | 0.33 | 0.33 | 0.19 | 0.55 | 0.44 |
| ## DIS45 | 0.32 | 0.37 | 0.38 | 0.30 | 0.43 | 0.33 | 1.00 | 0.31 | 0.42 | 0.24 | 0.39 | 0.43 |
| ## DIS48 | 0.39 | 0.30 | 0.35 | 0.37 | 0.45 | 0.33 | 0.31 | 1.00 | 0.37 | 0.48 | 0.46 | 0.42 |
| ## DIS51 | 0.30 | 0.39 | 0.30 | 0.28 | 0.41 | 0.33 | 0.42 | 0.37 | 1.00 | 0.24 | 0.43 | 0.38 |
| ## DIS54 | 0.22 | 0.24 | 0.25 | 0.40 | 0.23 | 0.19 | 0.24 | 0.48 | 0.24 | 1.00 | 0.23 | 0.27 |
| ## DIS57 | 0.48 | 0.40 | 0.53 | 0.38 | 0.59 | 0.55 | 0.39 | 0.46 | 0.43 | 0.23 | 1.00 | 0.55 |
| ## DIS60 | 0.52 | 0.30 | 0.47 | 0.34 | 0.73 | 0.44 | 0.43 | 0.42 | 0.38 | 0.27 | 0.55 | 1.00 |
| ## DIS63 | 0.30 | 0.25 | 0.36 | 0.40 | 0.41 | 0.24 | 0.29 | 0.54 | 0.32 | 0.53 | 0.35 | 0.38 |
| ## DIS66 | 0.36 | 0.21 | 0.31 | 0.27 | 0.34 | 0.28 | 0.27 | 0.28 | 0.27 | 0.22 | 0.33 | 0.28 |
| ## DIS69 | 0.37 | 0.34 | 0.30 | 0.30 | 0.38 | 0.38 | 0.38 | 0.31 | 0.40 | 0.22 | 0.41 | 0.32 |
| ## DIS72 | 0.51 | 0.37 | 0.49 | 0.41 | 0.63 | 0.55 | 0.45 | 0.49 | 0.47 | 0.28 | 0.70 | 0.59 |
| ## DIS75 | 0.14 | 0.20 | 0.06 | 0.11 | 0.16 | 0.11 | 0.15 | 0.13 | 0.14 | 0.09 | 0.17 | 0.14 |
| ## | DIS63 | DIS66 | DIS69 | DIS72 | DIS75 |  |  |  |  |  |  |  |
| ## NEG01 | 0.09 | 0.16 | 0.09 | 0.13 | 0.01 |  |  |  |  |  |  |  |
| ## NEG04 | 0.03 | -0.02 | -0.02 | 0.03 | 0.01 |  |  |  |  |  |  |  |
| ## NEG07 | -0.02 | 0.09 | 0.01 | 0.02 | 0.00 |  |  |  |  |  |  |  |
| ## NEG10 | 0.06 | 0.06 | 0.07 | 0.08 | 0.02 |  |  |  |  |  |  |  |
| ## NEG13 | 0.09 | 0.23 | 0.17 | 0.18 | 0.09 |  |  |  |  |  |  |  |
| ## NEG16 | 0.07 | 0.20 | 0.12 | 0.14 | 0.08 |  |  |  |  |  |  |  |
| ## NEG19 | 0.06 | 0.02 | 0.03 | 0.05 | 0.02 |  |  |  |  |  |  |  |
| ## NEG22 | 0.01 | 0.01 | 0.00 | 0.01 | -0.03 |  |  |  |  |  |  |  |
| ## NEG25 | 0.19 | 0.28 | 0.24 | 0.29 | 0.10 |  |  |  |  |  |  |  |
| ## NEG28 | 0.10 | 0.08 | 0.06 | 0.07 | 0.12 |  |  |  |  |  |  |  |
| ## NEG31 | 0.04 | 0.05 | 0.04 | 0.12 | 0.05 |  |  |  |  |  |  |  |
| ## NEG34 | 0.05 | 0.02 | 0.01 | 0.04 | 0.01 |  |  |  |  |  |  |  |
| ## NEG37 | 0.04 | 0.14 | 0.04 | 0.05 | 0.02 |  |  |  |  |  |  |  |
| ## NEG40 | 0.14 | 0.07 | 0.16 | 0.15 | 0.06 |  |  |  |  |  |  |  |
| ## NEG43 | 0.11 | 0.12 | 0.16 | 0.17 | 0.08 |  |  |  |  |  |  |  |
| ## NEG46 | 0.11 | 0.05 | 0.12 | 0.13 | 0.03 |  |  |  |  |  |  |  |
| ## NEG49 | -0.05 | -0.04 | -0.04 | -0.07 | -0.04 |  |  |  |  |  |  |  |
| ## NEG52 | 0.01 | 0.05 | 0.12 | 0.13 | 0.06 |  |  |  |  |  |  |  |
| ## NEG55 | 0.15 | 0.20 | 0.23 | 0.32 | 0.11 |  |  |  |  |  |  |  |
| ## NEG58 | 0.05 | 0.14 | 0.08 | 0.16 | 0.07 |  |  |  |  |  |  |  |
| ## NEG61 | 0.06 | 0.09 | 0.10 | 0.11 | 0.02 |  |  |  |  |  |  |  |
| ## NEG64 | 0.18 | 0.14 | 0.15 | 0.30 | 0.04 |  |  |  |  |  |  |  |
| ## NEG67 | 0.14 | 0.17 | 0.16 | 0.18 | 0.07 |  |  |  |  |  |  |  |
| ## NEG70 | -0.01 | 0.03 | 0.06 | 0.07 | 0.00 |  |  |  |  |  |  |  |
| ## NEG73 | 0.21 | 0.25 | 0.25 | 0.46 | 0.09 |  |  |  |  |  |  |  |
| ## NEG76 | 0.10 | 0.01 | 0.04 | 0.08 | 0.17 |  |  |  |  |  |  |  |
| ## POS02 | 0.04 | 0.06 | 0.22 | 0.17 | 0.04 |  |  |  |  |  |  |  |

|  |  |  |  |  |  |
| --- | --- | --- | --- | --- | --- |
| ## POS05 | 0.04 | 0.03 | 0.10 | 0.08 | 0.09 |
| ## POS08 | 0.07 | 0.15 | 0.13 | 0.22 | 0.01 |
| ## POS11 | 0.12 | 0.15 | 0.24 | 0.30 | 0.08 |
| ## POS14 | 0.00 | -0.02 | 0.14 | 0.15 | 0.09 |
| ## POS17 | 0.05 | 0.14 | 0.26 | 0.18 | 0.10 |
| ## POS20 | 0.10 | 0.07 | 0.18 | 0.10 | 0.08 |
| ## POS23 | 0.08 | 0.09 | 0.21 | 0.18 | 0.05 |
| ## POS26 | 0.06 | 0.04 | 0.15 | 0.13 | 0.09 |
| ## POS29 | 0.10 | 0.12 | 0.14 | 0.13 | 0.01 |
| ## POS32 | 0.17 | 0.04 | 0.25 | 0.27 | 0.07 |
| ## POS35 | 0.16 | 0.24 | 0.11 | 0.23 | 0.05 |
| ## POS38 | 0.10 | 0.16 | 0.24 | 0.20 | 0.07 |
| ## POS41 | 0.22 | 0.15 | 0.26 | 0.37 | 0.06 |
| ## POS44 | 0.12 | 0.11 | 0.12 | 0.16 | 0.03 |
| ## POS47 | 0.20 | 0.20 | 0.27 | 0.30 | 0.18 |
| ## POS50 | 0.21 | 0.20 | 0.27 | 0.36 | 0.14 |
| ## POS53 | 0.18 | 0.18 | 0.26 | 0.25 | 0.09 |
| ## POS56 | 0.20 | 0.09 | 0.26 | 0.24 | 0.11 |
| ## POS59 | 0.18 | 0.17 | 0.26 | 0.22 | 0.10 |
| ## POS62 | 0.13 | 0.07 | 0.17 | 0.19 | 0.08 |
| ## POS65 | 0.06 | 0.14 | 0.26 | 0.21 | 0.08 |
| ## POS68 | 0.10 | 0.10 | 0.30 | 0.27 | 0.10 |
| ## POS71 | 0.11 | 0.07 | 0.26 | 0.20 | 0.12 |
| ## POS74 | 0.20 | 0.16 | 0.33 | 0.40 | 0.09 |
| ## POS77 | 0.11 | 0.14 | 0.12 | 0.18 | 0.05 |
| ## DIS03 | 0.35 | 0.27 | 0.28 | 0.43 | 0.16 |
| ## DIS06 | 0.15 | 0.20 | 0.26 | 0.32 | 0.10 |
| ## DIS09 | 0.37 | 0.33 | 0.33 | 0.50 | 0.17 |
| ## DIS12 | 0.33 | 0.28 | 0.25 | 0.30 | 0.13 |
| ## DIS15 | 0.32 | 0.31 | 0.48 | 0.42 | 0.20 |
| ## DIS18 | 0.35 | 0.26 | 0.37 | 0.41 | 0.18 |
| ## DIS21 | 0.39 | 0.32 | 0.44 | 0.63 | 0.18 |
| ## DIS24 | 0.38 | 0.21 | 0.26 | 0.26 | 0.10 |
| ## DIS27 | 0.30 | 0.36 | 0.37 | 0.51 | 0.14 |
| ## DIS30 | 0.25 | 0.21 | 0.34 | 0.37 | 0.20 |
| ## DIS33 | 0.36 | 0.31 | 0.30 | 0.49 | 0.06 |
| ## DIS36 | 0.40 | 0.27 | 0.30 | 0.41 | 0.11 |
| ## DIS39 | 0.41 | 0.34 | 0.38 | 0.63 | 0.16 |
| ## DIS42 | 0.24 | 0.28 | 0.38 | 0.55 | 0.11 |
| ## DIS45 | 0.29 | 0.27 | 0.38 | 0.45 | 0.15 |
| ## DIS48 | 0.54 | 0.28 | 0.31 | 0.49 | 0.13 |
| ## DIS51 | 0.32 | 0.27 | 0.40 | 0.47 | 0.14 |
| ## DIS54 | 0.53 | 0.22 | 0.22 | 0.28 | 0.09 |
| ## DIS57 | 0.35 | 0.33 | 0.41 | 0.70 | 0.17 |
| ## DIS60 | 0.38 | 0.28 | 0.32 | 0.59 | 0.14 |
| ## DIS63 | 1.00 | 0.24 | 0.30 | 0.45 | 0.08 |
| ## DIS66 | 0.24 | 1.00 | 0.26 | 0.42 | 0.10 |
| ## DIS69 | 0.30 | 0.26 | 1.00 | 0.49 | 0.36 |
| ## DIS72 | 0.45 | 0.42 | 0.49 | 1.00 | 0.17 |
| ## DIS75 | 0.08 | 0.10 | 0.36 | 0.17 | 1.00 |

|  |  |  |  |  |  |  |  |  |  |  |  |  |
| --- | --- | --- | --- | --- | --- | --- | --- | --- | --- | --- | --- | --- |
| ## | NEG01 | NEG04 | NEG07 | NEG10 | NEG13 | NEG16 | NEG19 | NEG22 | NEG25 | NEG28 | NEG31 | NEG34 |
| ## NEG01 | 1.00 | 0.09 | 0.14 | 0.23 | 0.15 | 0.37 | 0.14 | 0.14 | 0.17 | 0.18 | 0.21 | 0.11 |
| ## NEG04 | 0.09 | 1.00 | 0.18 | 0.13 | 0.05 | 0.05 | 0.34 | 0.14 | 0.08 | 0.07 | 0.09 | 0.15 |

```
## NEG07  0.14  0.18  1.00  0.26  0.10  0.18  0.24  0.27  0.14  0.08  0.25  0.22
## NEG10  0.23  0.13  0.26  1.00  0.21  0.17  0.26  0.24  0.20  0.18  0.17  0.24
## NEG13  0.15  0.05  0.10  0.21  1.00  0.10  0.10  0.11  0.18  0.10  0.15  0.11
## NEG16  0.37  0.05  0.18  0.17  0.10  1.00  0.11  0.14  0.28  0.26  0.09  0.00
##      NEG37 NEG40 NEG43 NEG46 NEG49 NEG52 NEG55 NEG58 NEG61 NEG64 NEG67 NEG70
## NEG01  0.24  0.15  0.12  0.16  0.10  0.19  0.22  0.12  0.19  0.15  0.41  0.23
## NEG04  0.03  0.07  0.08  0.17  0.23  0.09  0.10  0.09  0.04  0.07  0.10  0.05
## NEG07  0.14  0.07  0.13  0.17  0.23  0.21  0.14  0.06  0.18  0.10  0.24  0.23
## NEG10  0.10  0.25  0.20  0.31  0.13  0.29  0.24  0.15  0.28  0.21  0.25  0.26
## NEG13  0.13  0.22  0.21  0.19  0.01  0.19  0.23  0.09  0.18  0.16  0.18  0.13
## NEG16  0.35  0.20  0.24  0.16  0.05  0.27  0.28  0.10  0.31  0.17  0.58  0.24
##      NEG73 NEG76 POS02 POS05 POS08 POS11 POS14 POS17 POS20 POS23 POS26 POS29
## NEG01  0.23  0.10  0.11  0.03  0.11  0.05  0.03  0.11  0.05  0.11  0.07  0.04
## NEG04  0.06  0.09  0.01 -0.01  0.01  0.04  0.01 -0.06  0.02  0.00 -0.01 -0.02
## NEG07  0.11  0.16  0.05  0.04  0.08  0.02  0.09  0.07  0.03 -0.05 -0.01  0.01
## NEG10  0.25  0.24  0.10  0.07  0.09  0.14  0.09  0.04  0.01  0.01  0.01  0.04
## NEG13  0.25  0.06  0.10  0.08  0.12  0.24 -0.02  0.06  0.10  0.10  0.09  0.07
## NEG16  0.27  0.06  0.07  0.18  0.07  0.08  0.17  0.18  0.07  0.12  0.17  0.09
##      POS32 POS35 POS38 POS41 POS44 POS47 POS50 POS53 POS56 POS59 POS62 POS65
## NEG01  0.03  0.02  0.19  0.09  0.05  0.09  0.07  0.04  0.06  0.09  0.13  0.14
## NEG04  0.00  0.08  0.03  0.03  0.02  0.00  0.03  0.03  0.01  0.00  0.02  0.04
## NEG07  0.01  0.00  0.04  0.00  0.00  0.00  0.02 -0.01  0.03  0.00  0.01  0.04
## NEG10  0.01  0.07  0.09  0.11  0.02  0.06  0.11  0.10  0.09  0.06  0.07  0.13
## NEG13  0.08  0.10  0.16  0.10  0.12  0.15  0.14  0.16  0.13  0.17  0.16  0.17
## NEG16  0.11  0.05  0.10  0.08  0.08  0.19  0.14  0.24  0.12  0.14  0.19  0.02
##      POS68 POS71 POS74 POS77 DIS03 DIS06 DIS09 DIS12 DIS15 DIS18 DIS21 DIS24
## NEG01  0.03  0.11  0.08  0.05  0.10  0.16  0.07  0.11  0.09  0.12  0.10  0.08
## NEG04  0.04  0.04  0.04  0.03  0.01  0.06  0.00  0.01  0.03  0.02 -0.03 -0.02
## NEG07  0.04  0.03 -0.02 -0.02  0.04  0.09  0.05  0.04  0.02  0.00  0.02 -0.01
## NEG10  0.10  0.06  0.08  0.08  0.03  0.13  0.03  0.10  0.07  0.03  0.09  0.05
## NEG13  0.12  0.08  0.11  0.10  0.15  0.20  0.18  0.12  0.21  0.15  0.21  0.10
## NEG16  0.07  0.20  0.06  0.15  0.10  0.24  0.14  0.13  0.16  0.12  0.13  0.09
##      DIS27 DIS30 DIS33 DIS36 DIS39 DIS42 DIS45 DIS48 DIS51 DIS54 DIS57 DIS60
## NEG01  0.12  0.15  0.09  0.11  0.13  0.13  0.05  0.10  0.13  0.10  0.10  0.12
## NEG04  0.01  0.03 -0.01 -0.01  0.05  0.04 -0.02  0.00 -0.03 -0.01  0.02  0.01
## NEG07  0.06  0.04  0.03  0.03  0.07  0.04  0.03  0.03  0.03 -0.02  0.02  0.08
## NEG10  0.03  0.11  0.07  0.08  0.09  0.14  0.09  0.04  0.11  0.03  0.08  0.11
## NEG13  0.10  0.18  0.17  0.16  0.19  0.21  0.20  0.10  0.23  0.11  0.18  0.18
## NEG16  0.11  0.26  0.05  0.11  0.17  0.11  0.16  0.11  0.17  0.04  0.09  0.08
##      DIS63 DIS66 DIS69 DIS72 DIS75
## NEG01  0.09  0.16  0.09  0.13  0.01
## NEG04  0.03 -0.02 -0.02  0.03  0.01
## NEG07 -0.02  0.09  0.01  0.02  0.00
## NEG10  0.06  0.06  0.07  0.08  0.02
## NEG13  0.09  0.23  0.17  0.18  0.09
## NEG16  0.07  0.20  0.12  0.14  0.08
```

*# Check if the 2 matrices are statistically different:*

*# Jennrich test for equality of correlation matrices.*

```
library(cocor)
library(psych)
nMSS <- 77
corrMSS_result <- cortest.jennrich(mat.MSS, phi_matrix_MSS, nMSS, nMSS)
print(corrMSS_result) # The two matrices are not statistically different
## $chi2
## [1] 4.446466
##
## $prob
## [1] 1
# Between the 2 is better to use the one with more decimal digits --> spearman
```

#### 2. FUNCTION TO CREATE THE EGA object with EBIC glasso

```
ega.object <- function (matrix, dataset)
{
  cor.data <- matrix
  glasso.ebic <- EBICglasso(matrix,n=nrow(dataset))
  graph.glasso <- as.igraph(qgraph(abs(glasso.ebic), layout = "spring",
                                vsize = 3, DoNotPlot = TRUE))
  wc <- cluster_walktrap(graph.glasso)
  n.dim <- max(wc$membership)
  a <- list()
  a$n.dim <- n.dim
  a$correlation <- cor.data
  a$glasso <- glasso.ebic
  a$wc <- wc$membership
  dim.variables <- data.frame(items = colnames(dataset), dimension = a$wc)
  dim.variables <- dim.variables[order(dim.variables[, 2]), ]
  a$dim.variables <- dim.variables
  class(a) <- "EGA"

  return(a)
}

# Create object
ega_MSS <- ega.object(mat.MSS, MSS)

# Extrapolate dimensions and labeling them
dim.MSS <- ega_MSS$dim.variables

oneMSS <- which(dim.MSS$dimension==1)
dim.MSS$dimension[oneMSS] <- rep("Negative",length(oneMSS))
twoMSS <- which(dim.MSS$dimension==2)
dim.MSS$dimension[twoMSS] <- rep("Positive",length(twoMSS))
threeMSS <- which(dim.MSS$dimension==3)
dim.MSS$dimension[threeMSS] <- rep("Disorganised",length(threeMSS))
```

#### 3. REPRESENTATION

```
graph1 <- ega_MSS$glasso[match(dim.MSS$items, colnames(MSS)),
                         match(dim.MSS$items,colnames(MSS))]
```

```
A_MSS <- qgraph(graph1, groups = dim.MSS$dimension,
  label.prop = 0.8 ,
  color = c("#fde725", "#31688e", "#440154"), layout = "spring",
  title = "EGA model of the MSS")
```

#### EGA model of the MSS

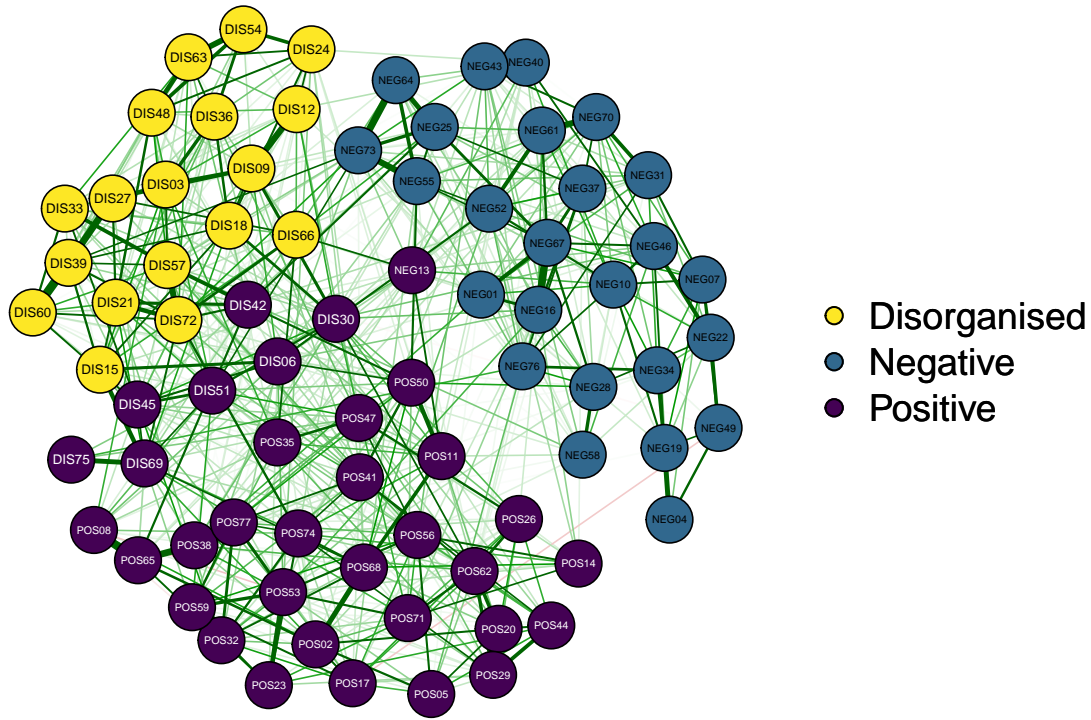

```
# This is the division of the items found with the EGA algorithm.
# It is a 3 dimensional structure.
# 8 Items are wrongly categorized.
# NEG13;DIS06;DIS30;DIS42;DIS45;DIS51;DIS69;DIS75.

# Representing these 8 items as a different dimension
groups4dim <- c(rep("Negative",25), rep("Disconnection",1), rep("Positive",26),
  rep("Disconnection",7), rep("Disorganised",18))

A_MSS_4factors <- qgraph(graph1, groups = groups4dim,
  label.prop = 0.8 ,
  color = c("#35b779", "#fde725", "#31688e", "#440154"),
  layout = "spring",
  vsize = 4, title = "EGA model of the MSS - 4 FACTORS")
```

#### EGA model of the MSS – 4 FACTORS

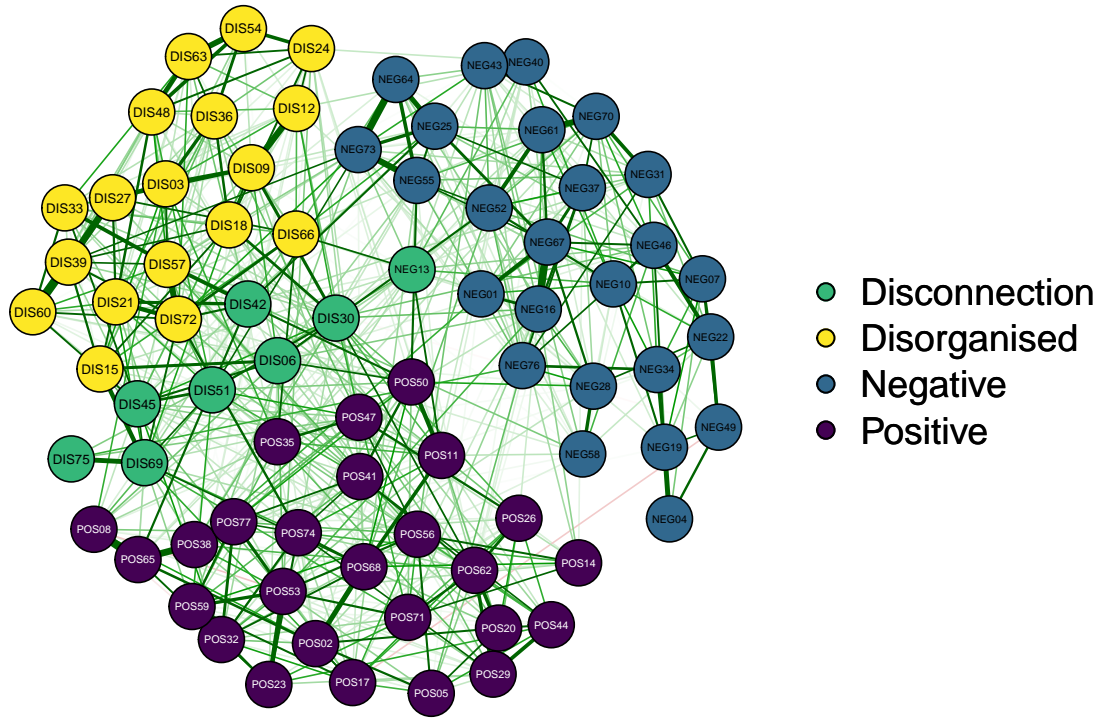

```
# Centrality measures of the network and representation in 4 FACTOR NETWORK
centralityTableMSS <- centralityTable(A_MSS_4factors, relative = TRUE,
                                     standardized = FALSE)

centralityTableMSS
```

| ## | graph | type | node | measure | value |
| --- | --- | --- | --- | --- | --- |
| ## 1 | graph 1 | NA | NEG01 | Betweenness | 0.147058824 |
| ## 2 | graph 1 | NA | NEG04 | Betweenness | 0.014705882 |
| ## 3 | graph 1 | NA | NEG07 | Betweenness | 0.014705882 |
| ## 4 | graph 1 | NA | NEG10 | Betweenness | 0.294117647 |
| ## 5 | graph 1 | NA | NEG16 | Betweenness | 0.464705882 |
| ## 6 | graph 1 | NA | NEG19 | Betweenness | 0.226470588 |
| ## 7 | graph 1 | NA | NEG22 | Betweenness | 0.270588235 |
| ## 8 | graph 1 | NA | NEG25 | Betweenness | 0.638235294 |
| ## 9 | graph 1 | NA | NEG28 | Betweenness | 0.264705882 |
| ## 10 | graph 1 | NA | NEG31 | Betweenness | 0.035294118 |
| ## 11 | graph 1 | NA | NEG34 | Betweenness | 0.064705882 |
| ## 12 | graph 1 | NA | NEG37 | Betweenness | 0.085294118 |
| ## 13 | graph 1 | NA | NEG40 | Betweenness | 0.067647059 |
| ## 14 | graph 1 | NA | NEG43 | Betweenness | 0.182352941 |
| ## 15 | graph 1 | NA | NEG46 | Betweenness | 0.623529412 |
| ## 16 | graph 1 | NA | NEG49 | Betweenness | 0.017647059 |
| ## 17 | graph 1 | NA | NEG52 | Betweenness | 0.558823529 |
| ## 18 | graph 1 | NA | NEG55 | Betweenness | 0.247058824 |
| ## 19 | graph 1 | NA | NEG58 | Betweenness | 0.000000000 |
| ## 20 | graph 1 | NA | NEG61 | Betweenness | 0.350000000 |

|  |  |  |  |  |
| --- | --- | --- | --- | --- |
| ## 21 | graph 1 | NA NEG64 | Betweenness | 0.000000000 |
| ## 22 | graph 1 | NA NEG67 | Betweenness | 1.000000000 |
| ## 23 | graph 1 | NA NEG70 | Betweenness | 0.200000000 |
| ## 24 | graph 1 | NA NEG73 | Betweenness | 0.502941176 |
| ## 25 | graph 1 | NA NEG76 | Betweenness | 0.250000000 |
| ## 26 | graph 1 | NA NEG13 | Betweenness | 0.076470588 |
| ## 27 | graph 1 | NA POS02 | Betweenness | 0.335294118 |
| ## 28 | graph 1 | NA POS05 | Betweenness | 0.067647059 |
| ## 29 | graph 1 | NA POS08 | Betweenness | 0.088235294 |
| ## 30 | graph 1 | NA POS11 | Betweenness | 0.314705882 |
| ## 31 | graph 1 | NA POS14 | Betweenness | 0.000000000 |
| ## 32 | graph 1 | NA POS17 | Betweenness | 0.038235294 |
| ## 33 | graph 1 | NA POS20 | Betweenness | 0.082352941 |
| ## 34 | graph 1 | NA POS23 | Betweenness | 0.123529412 |
| ## 35 | graph 1 | NA POS26 | Betweenness | 0.026470588 |
| ## 36 | graph 1 | NA POS29 | Betweenness | 0.135294118 |
| ## 37 | graph 1 | NA POS32 | Betweenness | 0.038235294 |
| ## 38 | graph 1 | NA POS35 | Betweenness | 0.032352941 |
| ## 39 | graph 1 | NA POS38 | Betweenness | 0.155882353 |
| ## 40 | graph 1 | NA POS41 | Betweenness | 0.141176471 |
| ## 41 | graph 1 | NA POS44 | Betweenness | 0.017647059 |
| ## 42 | graph 1 | NA POS47 | Betweenness | 0.105882353 |
| ## 43 | graph 1 | NA POS50 | Betweenness | 0.667647059 |
| ## 44 | graph 1 | NA POS53 | Betweenness | 0.464705882 |
| ## 45 | graph 1 | NA POS56 | Betweenness | 0.167647059 |
| ## 46 | graph 1 | NA POS59 | Betweenness | 0.052941176 |
| ## 47 | graph 1 | NA POS62 | Betweenness | 0.332352941 |
| ## 48 | graph 1 | NA POS65 | Betweenness | 0.426470588 |
| ## 49 | graph 1 | NA POS68 | Betweenness | 0.514705882 |
| ## 50 | graph 1 | NA POS71 | Betweenness | 0.102941176 |
| ## 51 | graph 1 | NA POS74 | Betweenness | 0.397058824 |
| ## 52 | graph 1 | NA POS77 | Betweenness | 0.082352941 |
| ## 53 | graph 1 | NA DIS06 | Betweenness | 0.461764706 |
| ## 54 | graph 1 | NA DIS30 | Betweenness | 0.700000000 |
| ## 55 | graph 1 | NA DIS42 | Betweenness | 0.641176471 |
| ## 56 | graph 1 | NA DIS45 | Betweenness | 0.100000000 |
| ## 57 | graph 1 | NA DIS51 | Betweenness | 0.191176471 |
| ## 58 | graph 1 | NA DIS69 | Betweenness | 0.529411765 |
| ## 59 | graph 1 | NA DIS75 | Betweenness | 0.223529412 |
| ## 60 | graph 1 | NA DIS03 | Betweenness | 0.232352941 |
| ## 61 | graph 1 | NA DIS09 | Betweenness | 0.329411765 |
| ## 62 | graph 1 | NA DIS12 | Betweenness | 0.017647059 |
| ## 63 | graph 1 | NA DIS15 | Betweenness | 0.364705882 |
| ## 64 | graph 1 | NA DIS18 | Betweenness | 0.247058824 |
| ## 65 | graph 1 | NA DIS21 | Betweenness | 0.441176471 |
| ## 66 | graph 1 | NA DIS24 | Betweenness | 0.020588235 |
| ## 67 | graph 1 | NA DIS27 | Betweenness | 0.220588235 |
| ## 68 | graph 1 | NA DIS33 | Betweenness | 0.020588235 |
| ## 69 | graph 1 | NA DIS36 | Betweenness | 0.111764706 |
| ## 70 | graph 1 | NA DIS39 | Betweenness | 0.423529412 |
| ## 71 | graph 1 | NA DIS48 | Betweenness | 0.282352941 |
| ## 72 | graph 1 | NA DIS54 | Betweenness | 0.064705882 |
| ## 73 | graph 1 | NA DIS57 | Betweenness | 0.444117647 |
| ## 74 | graph 1 | NA DIS60 | Betweenness | 0.035294118 |

|  |  |  |  |  |
| --- | --- | --- | --- | --- |
| ## 75 | graph 1 | NA DIS63 | Betweenness | 0.005882353 |
| ## 76 | graph 1 | NA DIS66 | Betweenness | 0.167647059 |
| ## 77 | graph 1 | NA DIS72 | Betweenness | 0.673529412 |
| ## 78 | graph 1 | NA NEG01 | Closeness | 0.810933750 |
| ## 79 | graph 1 | NA NEG04 | Closeness | 0.563024065 |
| ## 80 | graph 1 | NA NEG07 | Closeness | 0.671037444 |
| ## 81 | graph 1 | NA NEG10 | Closeness | 0.736896017 |
| ## 82 | graph 1 | NA NEG16 | Closeness | 0.892940129 |
| ## 83 | graph 1 | NA NEG19 | Closeness | 0.618036993 |
| ## 84 | graph 1 | NA NEG22 | Closeness | 0.693924876 |
| ## 85 | graph 1 | NA NEG25 | Closeness | 0.928949846 |
| ## 86 | graph 1 | NA NEG28 | Closeness | 0.790051053 |
| ## 87 | graph 1 | NA NEG31 | Closeness | 0.668859110 |
| ## 88 | graph 1 | NA NEG34 | Closeness | 0.614829453 |
| ## 89 | graph 1 | NA NEG37 | Closeness | 0.788064273 |
| ## 90 | graph 1 | NA NEG40 | Closeness | 0.715023041 |
| ## 91 | graph 1 | NA NEG43 | Closeness | 0.731517974 |
| ## 92 | graph 1 | NA NEG46 | Closeness | 0.775716221 |
| ## 93 | graph 1 | NA NEG49 | Closeness | 0.601234975 |
| ## 94 | graph 1 | NA NEG52 | Closeness | 0.865231490 |
| ## 95 | graph 1 | NA NEG55 | Closeness | 0.931983321 |
| ## 96 | graph 1 | NA NEG58 | Closeness | 0.692887455 |
| ## 97 | graph 1 | NA NEG61 | Closeness | 0.806632738 |
| ## 98 | graph 1 | NA NEG64 | Closeness | 0.903856927 |
| ## 99 | graph 1 | NA NEG67 | Closeness | 0.922249711 |
| ## 100 | graph 1 | NA NEG70 | Closeness | 0.750953068 |
| ## 101 | graph 1 | NA NEG73 | Closeness | 0.945944536 |
| ## 102 | graph 1 | NA NEG76 | Closeness | 0.717202342 |
| ## 103 | graph 1 | NA NEG13 | Closeness | 0.795356543 |
| ## 104 | graph 1 | NA POS02 | Closeness | 0.871066208 |
| ## 105 | graph 1 | NA POS05 | Closeness | 0.761162587 |
| ## 106 | graph 1 | NA POS08 | Closeness | 0.842075549 |
| ## 107 | graph 1 | NA POS11 | Closeness | 0.927726762 |
| ## 108 | graph 1 | NA POS14 | Closeness | 0.703727355 |
| ## 109 | graph 1 | NA POS17 | Closeness | 0.749152242 |
| ## 110 | graph 1 | NA POS20 | Closeness | 0.749138382 |
| ## 111 | graph 1 | NA POS23 | Closeness | 0.828220825 |
| ## 112 | graph 1 | NA POS26 | Closeness | 0.692293898 |
| ## 113 | graph 1 | NA POS29 | Closeness | 0.732929324 |
| ## 114 | graph 1 | NA POS32 | Closeness | 0.781679026 |
| ## 115 | graph 1 | NA POS35 | Closeness | 0.749380099 |
| ## 116 | graph 1 | NA POS38 | Closeness | 0.868291215 |
| ## 117 | graph 1 | NA POS41 | Closeness | 0.833788945 |
| ## 118 | graph 1 | NA POS44 | Closeness | 0.698232679 |
| ## 119 | graph 1 | NA POS47 | Closeness | 0.758445031 |
| ## 120 | graph 1 | NA POS50 | Closeness | 0.996781999 |
| ## 121 | graph 1 | NA POS53 | Closeness | 0.873469996 |
| ## 122 | graph 1 | NA POS56 | Closeness | 0.776765429 |
| ## 123 | graph 1 | NA POS59 | Closeness | 0.786069759 |
| ## 124 | graph 1 | NA POS62 | Closeness | 0.807356363 |
| ## 125 | graph 1 | NA POS65 | Closeness | 0.893837547 |
| ## 126 | graph 1 | NA POS68 | Closeness | 0.922696844 |
| ## 127 | graph 1 | NA POS71 | Closeness | 0.772552670 |
| ## 128 | graph 1 | NA POS74 | Closeness | 0.888695863 |

|  |  |  |  |  |
| --- | --- | --- | --- | --- |
| ## 129 | graph 1 | NA POS77 | Closeness | 0.804375270 |
| ## 130 | graph 1 | NA DIS06 | Closeness | 0.957112103 |
| ## 131 | graph 1 | NA DIS30 | Closeness | 0.944229438 |
| ## 132 | graph 1 | NA DIS42 | Closeness | 0.999512769 |
| ## 133 | graph 1 | NA DIS45 | Closeness | 0.867499633 |
| ## 134 | graph 1 | NA DIS51 | Closeness | 0.932295811 |
| ## 135 | graph 1 | NA DIS69 | Closeness | 0.928256151 |
| ## 136 | graph 1 | NA DIS75 | Closeness | 0.843336762 |
| ## 137 | graph 1 | NA DIS03 | Closeness | 0.863668284 |
| ## 138 | graph 1 | NA DIS09 | Closeness | 0.880750043 |
| ## 139 | graph 1 | NA DIS12 | Closeness | 0.799323500 |
| ## 140 | graph 1 | NA DIS15 | Closeness | 0.943992968 |
| ## 141 | graph 1 | NA DIS18 | Closeness | 0.832308979 |
| ## 142 | graph 1 | NA DIS21 | Closeness | 0.963037725 |
| ## 143 | graph 1 | NA DIS24 | Closeness | 0.683815773 |
| ## 144 | graph 1 | NA DIS27 | Closeness | 0.878846969 |
| ## 145 | graph 1 | NA DIS33 | Closeness | 0.850217575 |
| ## 146 | graph 1 | NA DIS36 | Closeness | 0.751126936 |
| ## 147 | graph 1 | NA DIS39 | Closeness | 0.909211213 |
| ## 148 | graph 1 | NA DIS48 | Closeness | 0.771224946 |
| ## 149 | graph 1 | NA DIS54 | Closeness | 0.692261021 |
| ## 150 | graph 1 | NA DIS57 | Closeness | 0.974876463 |
| ## 151 | graph 1 | NA DIS60 | Closeness | 0.851656772 |
| ## 152 | graph 1 | NA DIS63 | Closeness | 0.703676073 |
| ## 153 | graph 1 | NA DIS66 | Closeness | 0.884726219 |
| ## 154 | graph 1 | NA DIS72 | Closeness | 1.000000000 |
| ## 155 | graph 1 | NA NEG01 | Strength | 0.400963920 |
| ## 156 | graph 1 | NA NEG04 | Strength | 0.270209057 |
| ## 157 | graph 1 | NA NEG07 | Strength | 0.479374872 |
| ## 158 | graph 1 | NA NEG10 | Strength | 0.632403574 |
| ## 159 | graph 1 | NA NEG16 | Strength | 0.721598121 |
| ## 160 | graph 1 | NA NEG19 | Strength | 0.500886204 |
| ## 161 | graph 1 | NA NEG22 | Strength | 0.494300201 |
| ## 162 | graph 1 | NA NEG25 | Strength | 0.664519355 |
| ## 163 | graph 1 | NA NEG28 | Strength | 0.433440912 |
| ## 164 | graph 1 | NA NEG31 | Strength | 0.444506540 |
| ## 165 | graph 1 | NA NEG34 | Strength | 0.377961168 |
| ## 166 | graph 1 | NA NEG37 | Strength | 0.435075906 |
| ## 167 | graph 1 | NA NEG40 | Strength | 0.601057816 |
| ## 168 | graph 1 | NA NEG43 | Strength | 0.653582826 |
| ## 169 | graph 1 | NA NEG46 | Strength | 0.569580856 |
| ## 170 | graph 1 | NA NEG49 | Strength | 0.301074412 |
| ## 171 | graph 1 | NA NEG52 | Strength | 0.606577444 |
| ## 172 | graph 1 | NA NEG55 | Strength | 0.722982662 |
| ## 173 | graph 1 | NA NEG58 | Strength | 0.266443817 |
| ## 174 | graph 1 | NA NEG61 | Strength | 0.562768309 |
| ## 175 | graph 1 | NA NEG64 | Strength | 0.592397572 |
| ## 176 | graph 1 | NA NEG67 | Strength | 0.874909377 |
| ## 177 | graph 1 | NA NEG70 | Strength | 0.652145476 |
| ## 178 | graph 1 | NA NEG73 | Strength | 0.862055724 |
| ## 179 | graph 1 | NA NEG76 | Strength | 0.434298709 |
| ## 180 | graph 1 | NA NEG13 | Strength | 0.391954514 |
| ## 181 | graph 1 | NA POS02 | Strength | 0.596001325 |
| ## 182 | graph 1 | NA POS05 | Strength | 0.355248640 |

|  |  |  |  |  |  |  |  |
| --- | --- | --- | --- | --- | --- | --- | --- |
| ## | 183 | graph | 1 | NA | POS08 | Strength | 0.504339731 |
| ## | 184 | graph | 1 | NA | POS11 | Strength | 0.643122594 |
| ## | 185 | graph | 1 | NA | POS14 | Strength | 0.278798996 |
| ## | 186 | graph | 1 | NA | POS17 | Strength | 0.496667929 |
| ## | 187 | graph | 1 | NA | POS20 | Strength | 0.532591605 |
| ## | 188 | graph | 1 | NA | POS23 | Strength | 0.468700167 |
| ## | 189 | graph | 1 | NA | POS26 | Strength | 0.461994631 |
| ## | 190 | graph | 1 | NA | POS29 | Strength | 0.546845851 |
| ## | 191 | graph | 1 | NA | POS32 | Strength | 0.586985380 |
| ## | 192 | graph | 1 | NA | POS35 | Strength | 0.363320915 |
| ## | 193 | graph | 1 | NA | POS38 | Strength | 0.480852889 |
| ## | 194 | graph | 1 | NA | POS41 | Strength | 0.585310697 |
| ## | 195 | graph | 1 | NA | POS44 | Strength | 0.474870911 |
| ## | 196 | graph | 1 | NA | POS47 | Strength | 0.608646752 |
| ## | 197 | graph | 1 | NA | POS50 | Strength | 0.694104734 |
| ## | 198 | graph | 1 | NA | POS53 | Strength | 0.782924001 |
| ## | 199 | graph | 1 | NA | POS56 | Strength | 0.624918276 |
| ## | 200 | graph | 1 | NA | POS59 | Strength | 0.558154848 |
| ## | 201 | graph | 1 | NA | POS62 | Strength | 0.708032650 |
| ## | 202 | graph | 1 | NA | POS65 | Strength | 0.698217440 |
| ## | 203 | graph | 1 | NA | POS68 | Strength | 0.617639660 |
| ## | 204 | graph | 1 | NA | POS71 | Strength | 0.509928932 |
| ## | 205 | graph | 1 | NA | POS74 | Strength | 0.652618228 |
| ## | 206 | graph | 1 | NA | POS77 | Strength | 0.492503718 |
| ## | 207 | graph | 1 | NA | DIS06 | Strength | 0.596091448 |
| ## | 208 | graph | 1 | NA | DIS30 | Strength | 0.766375455 |
| ## | 209 | graph | 1 | NA | DIS42 | Strength | 0.651115553 |
| ## | 210 | graph | 1 | NA | DIS45 | Strength | 0.622470018 |
| ## | 211 | graph | 1 | NA | DIS51 | Strength | 0.677663301 |
| ## | 212 | graph | 1 | NA | DIS69 | Strength | 0.832795366 |
| ## | 213 | graph | 1 | NA | DIS75 | Strength | 0.233562708 |
| ## | 214 | graph | 1 | NA | DIS03 | Strength | 0.627418557 |
| ## | 215 | graph | 1 | NA | DIS09 | Strength | 0.816042220 |
| ## | 216 | graph | 1 | NA | DIS12 | Strength | 0.511551035 |
| ## | 217 | graph | 1 | NA | DIS15 | Strength | 0.616392867 |
| ## | 218 | graph | 1 | NA | DIS18 | Strength | 0.589513631 |
| ## | 219 | graph | 1 | NA | DIS21 | Strength | 0.721595180 |
| ## | 220 | graph | 1 | NA | DIS24 | Strength | 0.486041928 |
| ## | 221 | graph | 1 | NA | DIS27 | Strength | 0.620399426 |
| ## | 222 | graph | 1 | NA | DIS33 | Strength | 0.520369234 |
| ## | 223 | graph | 1 | NA | DIS36 | Strength | 0.564697604 |
| ## | 224 | graph | 1 | NA | DIS39 | Strength | 0.880240914 |
| ## | 225 | graph | 1 | NA | DIS48 | Strength | 0.645905642 |
| ## | 226 | graph | 1 | NA | DIS54 | Strength | 0.531665929 |
| ## | 227 | graph | 1 | NA | DIS57 | Strength | 0.832974204 |
| ## | 228 | graph | 1 | NA | DIS60 | Strength | 0.633756665 |
| ## | 229 | graph | 1 | NA | DIS63 | Strength | 0.635795442 |
| ## | 230 | graph | 1 | NA | DIS66 | Strength | 0.457535008 |
| ## | 231 | graph | 1 | NA | DIS72 | Strength | 1.000000000 |
| ## | 232 | graph | 1 | NA | NEG01 | ExpectedInfluence | 0.400963920 |
| ## | 233 | graph | 1 | NA | NEG04 | ExpectedInfluence | 0.270209057 |
| ## | 234 | graph | 1 | NA | NEG07 | ExpectedInfluence | 0.479374872 |
| ## | 235 | graph | 1 | NA | NEG10 | ExpectedInfluence | 0.632403574 |
| ## | 236 | graph | 1 | NA | NEG16 | ExpectedInfluence | 0.721598121 |

|  |  |  |  |  |  |  |  |
| --- | --- | --- | --- | --- | --- | --- | --- |
| ## | 237 | graph | 1 | NA | NEG19 | ExpectedInfluence | 0.500886204 |
| ## | 238 | graph | 1 | NA | NEG22 | ExpectedInfluence | 0.494300201 |
| ## | 239 | graph | 1 | NA | NEG25 | ExpectedInfluence | 0.664519355 |
| ## | 240 | graph | 1 | NA | NEG28 | ExpectedInfluence | 0.433440912 |
| ## | 241 | graph | 1 | NA | NEG31 | ExpectedInfluence | 0.444506540 |
| ## | 242 | graph | 1 | NA | NEG34 | ExpectedInfluence | 0.377961168 |
| ## | 243 | graph | 1 | NA | NEG37 | ExpectedInfluence | 0.435075906 |
| ## | 244 | graph | 1 | NA | NEG40 | ExpectedInfluence | 0.601057816 |
| ## | 245 | graph | 1 | NA | NEG43 | ExpectedInfluence | 0.653582826 |
| ## | 246 | graph | 1 | NA | NEG46 | ExpectedInfluence | 0.569580856 |
| ## | 247 | graph | 1 | NA | NEG49 | ExpectedInfluence | 0.279154454 |
| ## | 248 | graph | 1 | NA | NEG52 | ExpectedInfluence | 0.606577444 |
| ## | 249 | graph | 1 | NA | NEG55 | ExpectedInfluence | 0.722982662 |
| ## | 250 | graph | 1 | NA | NEG58 | ExpectedInfluence | 0.266443817 |
| ## | 251 | graph | 1 | NA | NEG61 | ExpectedInfluence | 0.562768309 |
| ## | 252 | graph | 1 | NA | NEG64 | ExpectedInfluence | 0.592397572 |
| ## | 253 | graph | 1 | NA | NEG67 | ExpectedInfluence | 0.874909377 |
| ## | 254 | graph | 1 | NA | NEG70 | ExpectedInfluence | 0.652145476 |
| ## | 255 | graph | 1 | NA | NEG73 | ExpectedInfluence | 0.862055724 |
| ## | 256 | graph | 1 | NA | NEG76 | ExpectedInfluence | 0.434298709 |
| ## | 257 | graph | 1 | NA | NEG13 | ExpectedInfluence | 0.391954514 |
| ## | 258 | graph | 1 | NA | POS02 | ExpectedInfluence | 0.596001325 |
| ## | 259 | graph | 1 | NA | POS05 | ExpectedInfluence | 0.355248640 |
| ## | 260 | graph | 1 | NA | POS08 | ExpectedInfluence | 0.474273276 |
| ## | 261 | graph | 1 | NA | POS11 | ExpectedInfluence | 0.643122594 |
| ## | 262 | graph | 1 | NA | POS14 | ExpectedInfluence | 0.278798996 |
| ## | 263 | graph | 1 | NA | POS17 | ExpectedInfluence | 0.476874935 |
| ## | 264 | graph | 1 | NA | POS20 | ExpectedInfluence | 0.532591605 |
| ## | 265 | graph | 1 | NA | POS23 | ExpectedInfluence | 0.468700167 |
| ## | 266 | graph | 1 | NA | POS26 | ExpectedInfluence | 0.461994631 |
| ## | 267 | graph | 1 | NA | POS29 | ExpectedInfluence | 0.546845851 |
| ## | 268 | graph | 1 | NA | POS32 | ExpectedInfluence | 0.578645858 |
| ## | 269 | graph | 1 | NA | POS35 | ExpectedInfluence | 0.363320915 |
| ## | 270 | graph | 1 | NA | POS38 | ExpectedInfluence | 0.480852889 |
| ## | 271 | graph | 1 | NA | POS41 | ExpectedInfluence | 0.585310697 |
| ## | 272 | graph | 1 | NA | POS44 | ExpectedInfluence | 0.474870911 |
| ## | 273 | graph | 1 | NA | POS47 | ExpectedInfluence | 0.608646752 |
| ## | 274 | graph | 1 | NA | POS50 | ExpectedInfluence | 0.694104734 |
| ## | 275 | graph | 1 | NA | POS53 | ExpectedInfluence | 0.761197068 |
| ## | 276 | graph | 1 | NA | POS56 | ExpectedInfluence | 0.624918276 |
| ## | 277 | graph | 1 | NA | POS59 | ExpectedInfluence | 0.558154848 |
| ## | 278 | graph | 1 | NA | POS62 | ExpectedInfluence | 0.708032650 |
| ## | 279 | graph | 1 | NA | POS65 | ExpectedInfluence | 0.698217440 |
| ## | 280 | graph | 1 | NA | POS68 | ExpectedInfluence | 0.617639660 |
| ## | 281 | graph | 1 | NA | POS71 | ExpectedInfluence | 0.509928932 |
| ## | 282 | graph | 1 | NA | POS74 | ExpectedInfluence | 0.652618228 |
| ## | 283 | graph | 1 | NA | POS77 | ExpectedInfluence | 0.492503718 |
| ## | 284 | graph | 1 | NA | DIS06 | ExpectedInfluence | 0.596091448 |
| ## | 285 | graph | 1 | NA | DIS30 | ExpectedInfluence | 0.766375455 |
| ## | 286 | graph | 1 | NA | DIS42 | ExpectedInfluence | 0.651115553 |
| ## | 287 | graph | 1 | NA | DIS45 | ExpectedInfluence | 0.622470018 |
| ## | 288 | graph | 1 | NA | DIS51 | ExpectedInfluence | 0.677663301 |
| ## | 289 | graph | 1 | NA | DIS69 | ExpectedInfluence | 0.832795366 |
| ## | 290 | graph | 1 | NA | DIS75 | ExpectedInfluence | 0.233562708 |

```
## 291 graph 1 NA DIS03 ExpectedInfluence 0.627418557
## 292 graph 1 NA DIS09 ExpectedInfluence 0.816042220
## 293 graph 1 NA DIS12 ExpectedInfluence 0.511551035
## 294 graph 1 NA DIS15 ExpectedInfluence 0.616392867
## 295 graph 1 NA DIS18 ExpectedInfluence 0.589513631
## 296 graph 1 NA DIS21 ExpectedInfluence 0.721595180
## 297 graph 1 NA DIS24 ExpectedInfluence 0.486041928
## 298 graph 1 NA DIS27 ExpectedInfluence 0.620399426
## 299 graph 1 NA DIS33 ExpectedInfluence 0.520369234
## 300 graph 1 NA DIS36 ExpectedInfluence 0.564697604
## 301 graph 1 NA DIS39 ExpectedInfluence 0.880240914
## 302 graph 1 NA DIS48 ExpectedInfluence 0.645905642
## 303 graph 1 NA DIS54 ExpectedInfluence 0.531665929
## 304 graph 1 NA DIS57 ExpectedInfluence 0.830847240
## 305 graph 1 NA DIS60 ExpectedInfluence 0.633756665
## 306 graph 1 NA DIS63 ExpectedInfluence 0.635795442
## 307 graph 1 NA DIS66 ExpectedInfluence 0.457535008
## 308 graph 1 NA DIS72 ExpectedInfluence 1.000000000
```

```
centralityPlot(A_MSS_4factors, include = c("Strength", "Closeness", "Betweenness",
                                           "ExpectedInfluence"),
              scale = "relative", orderBy = "ExpectedInfluence" )
```

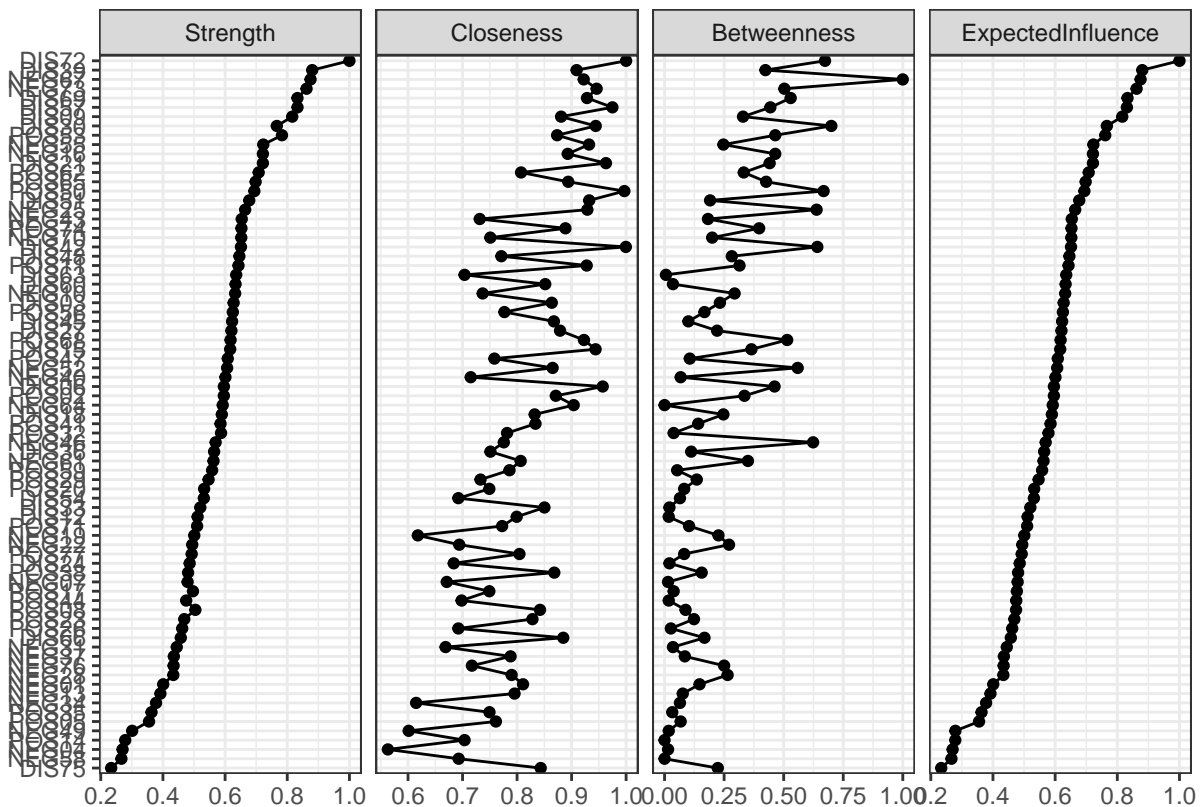

```
A_MSS_4factors <- qgraph(graph1, groups = groups4dim,
                        label.prop = 0.8 ,
```

```
color = c("#35b779", "#fde725", "#31688e", "#440154"),
layout = "spring",
vsize = centralityTableMSS[1:77, 5] * 5 + 3, # BETWEENNESS CENTRALITY
title = "EGA model of the MSS - 4 FACTORS + Betweenness centrality")
```

EGA model of the MSS – 4 FACTORS + Betweenness centrality

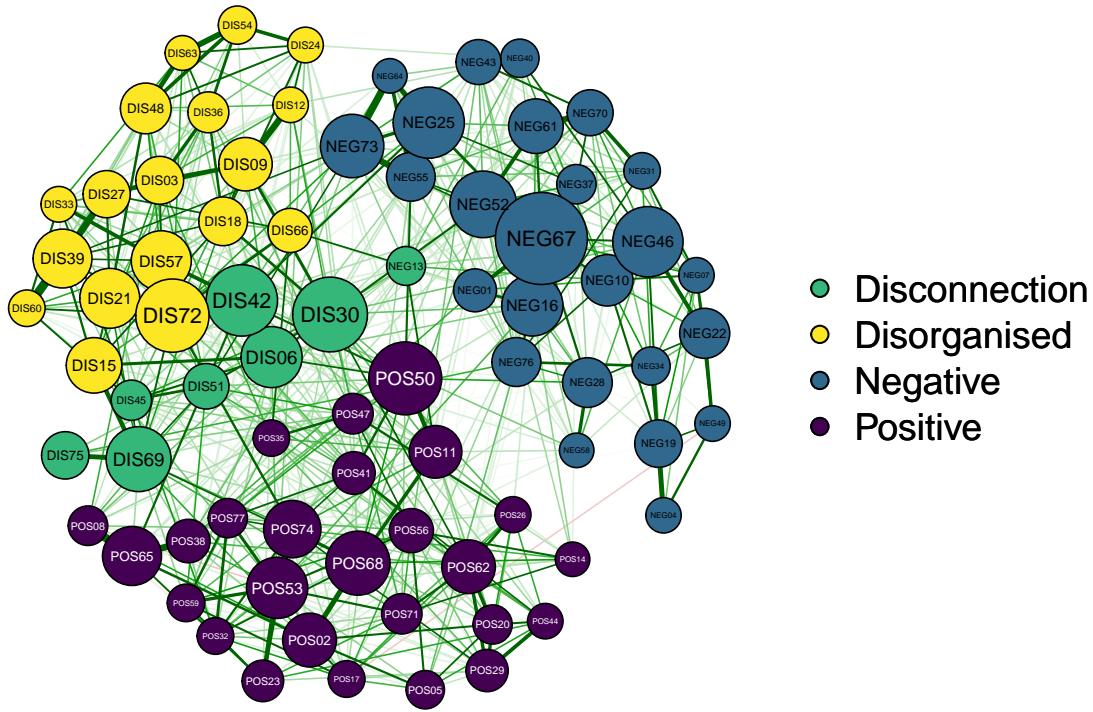

```
# Printing the final graph
# pdf("EGA_MSS_4dim.pdf", width = 10)
# dev.off()
```

###### 4. CONFIRMATORY FACTOR ANALYSIS - CFA for MSS

```
dim.MSS$items
```

```
## [1] "NEG01" "NEG04" "NEG07" "NEG10" "NEG16" "NEG19" "NEG22" "NEG25" "NEG28"
## [10] "NEG31" "NEG34" "NEG37" "NEG40" "NEG43" "NEG46" "NEG49" "NEG52" "NEG55"
## [19] "NEG58" "NEG61" "NEG64" "NEG67" "NEG70" "NEG73" "NEG76" "NEG13" "POS02"
## [28] "POS05" "POS08" "POS11" "POS14" "POS17" "POS20" "POS23" "POS26" "POS29"
## [37] "POS32" "POS35" "POS38" "POS41" "POS44" "POS47" "POS50" "POS53" "POS56"
## [46] "POS59" "POS62" "POS65" "POS68" "POS71" "POS74" "POS77" "DIS06" "DIS30"
## [55] "DIS42" "DIS45" "DIS51" "DIS69" "DIS75" "DIS03" "DIS09" "DIS12" "DIS15"
## [64] "DIS18" "DIS21" "DIS24" "DIS27" "DIS33" "DIS36" "DIS39" "DIS48" "DIS54"
## [73] "DIS57" "DIS60" "DIS63" "DIS66" "DIS72"
```

```
dim.MSS$dimension
```

```
## [1] "Negative"      "Negative"      "Negative"      "Negative"      "Negative"
## [6] "Negative"      "Negative"      "Negative"      "Negative"      "Negative"
## [11] "Negative"      "Negative"      "Negative"      "Negative"      "Negative"
## [16] "Negative"      "Negative"      "Negative"      "Negative"      "Negative"
## [21] "Negative"      "Negative"      "Negative"      "Negative"      "Negative"
## [26] "Positive"      "Positive"      "Positive"      "Positive"      "Positive"
## [31] "Positive"      "Positive"      "Positive"      "Positive"      "Positive"
## [36] "Positive"      "Positive"      "Positive"      "Positive"      "Positive"
## [41] "Positive"      "Positive"      "Positive"      "Positive"      "Positive"
## [46] "Positive"      "Positive"      "Positive"      "Positive"      "Positive"
## [51] "Positive"      "Positive"      "Positive"      "Positive"      "Positive"
## [56] "Positive"      "Positive"      "Positive"      "Positive"      "Disorganised"
## [61] "Disorganised" "Disorganised" "Disorganised" "Disorganised" "Disorganised"
## [66] "Disorganised" "Disorganised" "Disorganised" "Disorganised" "Disorganised"
## [71] "Disorganised" "Disorganised" "Disorganised" "Disorganised" "Disorganised"
## [76] "Disorganised" "Disorganised"
```

```
# This is the original model of MSS (Kwapil, Gross, Silvia, Raulin, et al., 2018)
```

```
model_MSS_original <- '
Negative =~ `NEG01` + `NEG04` + `NEG07` + `NEG10` + `NEG13` + `NEG16` +
`NEG19` + `NEG22` + `NEG25` + `NEG28` + `NEG31` + `NEG34` +
`NEG37` + `NEG40` + `NEG43` + `NEG46` + `NEG49` + `NEG52` +
`NEG55` + `NEG58` + `NEG61` + `NEG64` + `NEG67` + `NEG70` +
`NEG73` + `NEG76`
Positive =~ `POS02` + `POS05` + `POS08` + `POS11` + `POS14` + `POS17` +
`POS20` + `POS23` + `POS26` + `POS29` + `POS32` + `POS35` +
`POS38` + `POS41` + `POS44` + `POS47` + `POS50` + `POS53` +
`POS56` + `POS59` + `POS62` + `POS65` + `POS68` + `POS71` +
`POS74` + `POS77`
Disorganised =~ `DIS03` + `DIS06` + `DIS09` + `DIS12` + `DIS15` + `DIS18` +
`DIS21` + `DIS24` + `DIS27` + `DIS30` + `DIS33` + `DIS36` +
`DIS39` + `DIS42` + `DIS45` + `DIS48` + `DIS51` + `DIS54` +
`DIS57` + `DIS60` + `DIS63` + `DIS66` + `DIS69` + `DIS72` +
`DIS75`
'
```

```
# Model found with Exploratory Graph Analysis - EGA (3 DIMENSIONS)
```

```
model_MSS_ega <- '
Negative =~ `NEG01` + `NEG04` + `NEG07` + `NEG10` + `NEG16` + `NEG19` +
`NEG22` + `NEG25` + `NEG28` + `NEG31` + `NEG34` + `NEG37` +
`NEG40` + `NEG43` + `NEG46` + `NEG49` + `NEG52` + `NEG55` +
`NEG58` + `NEG61` + `NEG64` + `NEG67` + `NEG70` + `NEG73` + `NEG76`
Positive =~ `POS02` + `POS05` + `DIS06` + `POS08` + `POS11` + `NEG13` +
`POS14` + `POS17` + `POS20` + `POS23` + `POS26` + `POS29` +
`DIS30` + `POS32` + `POS35` + `POS38` + `POS41` + `DIS42` +
`POS44` + `DIS45` + `POS47` + `POS50` + `DIS51` + `POS53` +
`POS56` + `POS59` + `POS62` + `POS65` + `POS68` + `DIS69` +
`POS71` + `POS74` + `DIS75` + `POS77`
Disorganised =~ `DIS03` + `DIS09` + `DIS12` + `DIS15` + `DIS18` + `DIS21` +
`DIS24` + `DIS27` + `DIS33` + `DIS36` + `DIS39` + `DIS48` +
`DIS54` + `DIS57` + `DIS60` + `DIS63` + `DIS66` + `DIS72`
```

```
# Model with one more dimension for the misclassified items based on EGA
model_MSS_4dim <- '
Negative =~ `NEG01` + `NEG04` + `NEG07` + `NEG10` + `NEG16` + `NEG19` +
`NEG22` + `NEG25` + `NEG28` + `NEG31` + `NEG34` + `NEG37` +
`NEG40` + `NEG43` + `NEG46` + `NEG49` + `NEG52` + `NEG55` +
`NEG58` + `NEG61` + `NEG64` + `NEG67` + `NEG70` + `NEG73` +
`NEG76`
Positive =~ `POS02` + `POS05` + `POS08` + `POS11` + `POS14` + `POS17` +
`POS20` + `POS23` + `POS26` + `POS29` + `POS32` + `POS35` +
`POS38` + `POS41` + `POS44` + `POS47` + `POS50` + `POS53` +
`POS56` + `POS59` + `POS62` + `POS65` + `POS68` +
`POS71` + `POS74` + `POS77`
Disconnected = ~ `NEG13` + `DIS06` + `DIS30` + `DIS42` + `DIS45` + `DIS51` +
`DIS69` + `DIS75`
Disorganised =~ `DIS03` + `DIS09` + `DIS12` + `DIS15` + `DIS18` + `DIS21` +
`DIS24` + `DIS27` + `DIS33` + `DIS36` + `DIS39` + `DIS48` +
`DIS54` + `DIS57` + `DIS60` + `DIS63` + `DIS66` + `DIS72`
'
```

#### 5. FITTING THE MODELS AND COMPARISON OF THE RESULTING INDICES

```
# These are not computed in the Markdown because they would take too much time.

# Original MSS model
# fit.MSS.original <- cfa(model_MSS_original, data = MSS, estimator = "wlsmv",
#                          meanstructure = TRUE, ordered = TRUE)

# ordered = TRUE is setted because otherwise the variables would be treated as
# continuous

# # EGA model 3 dimensions
# fit.MSS.ega <- CFA(ega_MSS, estimator= "WLSMW", data = MSS,
#                   meanstructure = TRUE,
#                   ordered = TRUE, plot.CFA = FALSE)

# # Model with 4 dimensions
# fit.MSS.4dim <- cfa(model_MSS_4dim, data = MSS, estimator = "wlsmv",
#                   meanstructure = TRUE,
#                   ordered = TRUE)

fitMeasures(fit.MSS.original, c("chisq","df","pvalue","pvalue.scaled","srmr",
                                "cfi","cfi.scaled", "rmsea",
                                "rmsea.scaled", "tli", "tli.scaled"))
```

|  |  |  |  |  |  |
| --- | --- | --- | --- | --- | --- |
| ## | chisq | df | pvalue | pvalue.scaled | srmr |
| ## | 4899.174 | 2846.000 | 0.000 | 0.000 | 0.123 |
| ## | cfi | cfi.scaled | rmsea | rmsea.scaled | tli |
| ## | 0.979 | 0.941 | 0.026 | 0.015 | 0.978 |
| ## | tli.scaled |  |  |  |  |

```
##          0.940
```

```
fitMeasures(fit.MSS.ega$fit, c("chisq","df","pvalue","pvalue.scaled","srmr",
                               "cfi","cfi.scaled", "rmsea",
                               "rmsea.scaled", "tli", "tli.scaled"))
```

```
##          chisq          df      pvalue pvalue.scaled      srmr
##    5080.538    2846.000      0.000      0.000      0.124
##          cfi    cfi.scaled      rmsea  rmsea.scaled      tli
##          0.977          0.937      0.027      0.016      0.976
##    tli.scaled
##          0.935
```

```
fitMeasures(fit.MSS.4dim, c("chisq","df","pvalue","pvalue.scaled","srmr",
                             "cfi","cfi.scaled", "rmsea",
                             "rmsea.scaled", "tli", "tli.scaled"))
```

```
##          chisq          df      pvalue pvalue.scaled      srmr
##    4697.093    2843.000      0.000      0.000      0.120
##          cfi    cfi.scaled      rmsea  rmsea.scaled      tli
##          0.981          0.945      0.025      0.015      0.980
##    tli.scaled
##          0.943
```

#### 6.CHECK FOR SIGNIFICANT DIFFERENCES between the 3 MODELS

```
lavTestLRT(fit.MSS.4dim, fit.MSS.original, method="satorra.bentler.2010")
```

```
##
## Scaled Chi-Squared Difference Test (method = "satorra.bentler.2010")
##
## lavaan NOTE:
##   The "Chisq" column contains standard test statistics, not the
##   robust test that should be reported per model. A robust difference
##   test is a function of two standard (not robust) statistics.
##
##           Df AIC BIC  Chisq Chisq diff Df diff Pr(>Chisq)
## fit.MSS.4dim    2843      4697.1
## fit.MSS.original 2846      4899.2   -0.32282      3      1
```

```
lavTestLRT(fit.MSS.4dim, fit.MSS.ega$fit, method="satorra.bentler.2010")
```

```
##
## Scaled Chi-Squared Difference Test (method = "satorra.bentler.2010")
##
## lavaan NOTE:
##   The "Chisq" column contains standard test statistics, not the
##   robust test that should be reported per model. A robust difference
##   test is a function of two standard (not robust) statistics.
##
##           Df AIC BIC  Chisq Chisq diff Df diff Pr(>Chisq)
## fit.MSS.4dim    2843      4697.1
## fit.MSS.ega$fit 2846    5080.5   -0.13363      3      1
```

```
# McDonald's Omega and Chronback alpha of the 4 dimensional model
MSS_negative <- MSS[ , c("NEG01", "NEG04", "NEG07", "NEG10", "NEG16", "NEG19",
                        "NEG22", "NEG25", "NEG28", "NEG31", "NEG34", "NEG37",
                        "NEG40", "NEG43", "NEG46", "NEG49", "NEG52", "NEG55",
                        "NEG58", "NEG61", "NEG64", "NEG67", "NEG70", "NEG73",
                        "NEG76")]
MSS_positive <- MSS[ , c("POS02", "POS05", "POS08", "POS11", "POS14", "POS17",
                        "POS20", "POS23", "POS26", "POS29", "POS32", "POS35",
                        "POS38", "POS41", "POS44", "POS47", "POS50", "POS53",
                        "POS56", "POS59", "POS62", "POS65", "POS68",
                        "POS71", "POS74", "POS77")]
MSS_disorganised <- MSS[ , c("DIS03", "DIS09", "DIS12", "DIS15", "DIS18",
                        "DIS21", "DIS24", "DIS27", "DIS33", "DIS36",
                        "DIS39", "DIS48", "DIS54", "DIS57", "DIS60",
                        "DIS63", "DIS66", "DIS72")]
MSS_disconnected <- MSS[ , c("NEG13", "DIS06", "DIS30", "DIS42", "DIS45",
                        "DIS51", "DIS69", "DIS75")]

omega_negative <- omega(MSS_negative, plot = FALSE)
print(omega_negative)
```

```
## Omega
## Call: omegah(m = m, nfactors = nfactors, fm = fm, key = key, flip = flip,
##      digits = digits, title = title, sl = sl, labels = labels,
##      plot = plot, n.obs = n.obs, rotate = rotate, Phi = Phi, option = option,
##      covar = covar)
## Alpha:          0.85
## G.6:            0.88
## Omega Hierarchical: 0.57
## Omega H asymptotic: 0.65
## Omega Total      0.87
##
## Schmid Leiman Factor loadings greater than 0.2
##      g    F1*   F2*   F3*   h2   u2   p2
## NEG01 0.38 0.26                0.22 0.78 0.65
## NEG04                0.38 0.16 0.84 0.10
## NEG07 0.28                0.38 0.23 0.77 0.34
## NEG10 0.37                0.37 0.29 0.71 0.48
## NEG16 0.50 0.45                0.46 0.54 0.55
## NEG19                0.51 0.31 0.69 0.12
## NEG22 0.26                0.40 0.23 0.77 0.29
## NEG25 0.48          0.47                0.46 0.54 0.49
## NEG28 0.31                0.15 0.85 0.67
## NEG31 0.30                0.25 0.17 0.83 0.55
## NEG34                0.43 0.23 0.77 0.16
## NEG37 0.40 0.33                0.27 0.73 0.59
## NEG40 0.35                0.18 0.82 0.68
## NEG43 0.40                0.23 0.77 0.69
## NEG46 0.35                0.33 0.25 0.75 0.49
## NEG49                0.38 0.17 0.83 0.09
## NEG52 0.41 0.25                0.25 0.75 0.68
## NEG55 0.50          0.51                0.52 0.48 0.48
## NEG58 0.20                0.10 0.90 0.41
```

```
## NEG61  0.46  0.30                0.31 0.69 0.68
## NEG64  0.43                0.65        0.62 0.38 0.30
## NEG67  0.59  0.43                0.54 0.46 0.65
## NEG70  0.44  0.26                0.29 0.71 0.66
## NEG73  0.52                0.64        0.67 0.33 0.40
## NEG76  0.27                0.21 0.15 0.85 0.48
##
## With Sums of squares of:
##   g  F1*  F2*  F3*
## 3.51 0.91 1.46 1.56
##
## general/max  2.25   max/min =   1.71
## mean percent general =  0.47   with sd =  0.2 and cv of  0.42
## Explained Common Variance of the general factor =  0.47
##
## The degrees of freedom are 228 and the fit is  1.47
## The number of observations was 1059 with Chi Square = 1535.22 with prob < 2.4e-192
## The root mean square of the residuals is  0.05
## The df corrected root mean square of the residuals is  0.06
## RMSEA index =  0.074 and the 10 % confidence intervals are  0.07 0.077
## BIC = -52.81
##
## Compare this with the adequacy of just a general factor and no group factors
## The degrees of freedom for just the general factor are 275 and the fit is  2.99
## The number of observations was 1059 with Chi Square = 3134.27 with prob < 0
## The root mean square of the residuals is  0.1
## The df corrected root mean square of the residuals is  0.11
##
## RMSEA index =  0.099 and the 10 % confidence intervals are  0.096 0.102
## BIC = 1218.87
##
## Measures of factor score adequacy
##
##           g  F1*  F2*  F3*
## Correlation of scores with factors      0.79  0.61 0.79 0.78
## Multiple R square of scores with factors      0.63  0.37 0.62 0.61
## Minimum correlation of factor score estimates 0.26 -0.27 0.25 0.21
##
## Total, General and Subset omega for each subset
##
##           g  F1*  F2*  F3*
## Omega total for total scores and subscales      0.87 0.77 0.80 0.72
## Omega general for total scores and subscales      0.57 0.53 0.34 0.28
## Omega group for total scores and subscales      0.20 0.25 0.46 0.44

omega_positive <- omega(MSS_positive, plot = FALSE)
print(omega_positive)

## Omega
## Call: omegah(m = m, nfactors = nfactors, fm = fm, key = key, flip = flip,
##   digits = digits, title = title, sl = sl, labels = labels,
##   plot = plot, n.obs = n.obs, rotate = rotate, Phi = Phi, option = option,
##   covar = covar)
## Alpha:                0.85
## G.6:                  0.87
## Omega Hierarchical:    0.56
```

```
## Omega H asymptotic:    0.65
## Omega Total           0.87
##
## Schmid Leiman Factor loadings greater than 0.2
##      g   F1*   F2*   F3*   h2   u2   p2
## POS02 0.38                0.31 0.27 0.73 0.53
## POS05 0.30          0.22      0.17 0.83 0.53
## POS08 0.26                0.54 0.38 0.62 0.18
## POS11 0.38 0.25          0.21 0.25 0.75 0.57
## POS14 0.24                0.09 0.91 0.62
## POS17 0.34 0.27                0.20 0.80 0.58
## POS20 0.44          0.43      0.38 0.62 0.51
## POS23 0.31 0.31                0.20 0.80 0.49
## POS26 0.39          0.24      0.22 0.78 0.67
## POS29 0.43          0.38      0.33 0.67 0.57
## POS32 0.33 0.49                0.35 0.65 0.31
## POS35 0.26                0.10 0.90 0.68
## POS38 0.26                0.40 0.24 0.76 0.29
## POS41 0.38 0.28                0.24 0.76 0.60
## POS44 0.39          0.31      0.25 0.75 0.60
## POS47 0.35                0.19 0.81 0.64
## POS50 0.34 0.34                0.26 0.74 0.45
## POS53 0.37 0.48                0.37 0.63 0.37
## POS56 0.39 0.27                0.24 0.76 0.62
## POS59 0.33 0.40                0.28 0.72 0.40
## POS62 0.49          0.36      0.37 0.63 0.63
## POS65 0.29                0.74 0.63 0.37 0.13
## POS68 0.37                0.24 0.24 0.76 0.57
## POS71 0.36 0.23                0.21 0.79 0.62
## POS74 0.40 0.28                0.28 0.72 0.57
## POS77 0.32 0.42                0.28 0.72 0.37
##
## With Sums of squares of:
##      g   F1*   F2*   F3*
## 3.26 1.62 0.82 1.34
##
## general/max 2.02   max/min = 1.98
## mean percent general = 0.5   with sd = 0.15 and cv of 0.3
## Explained Common Variance of the general factor = 0.46
##
## The degrees of freedom are 250 and the fit is 1.31
## The number of observations was 1059 with Chi Square = 1375.42 with prob < 1.2e-154
## The root mean square of the residuals is 0.05
## The df corrected root mean square of the residuals is 0.05
## RMSEA index = 0.065 and the 10 % confidence intervals are 0.062 0.069
## BIC = -365.85
##
## Compare this with the adequacy of just a general factor and no group factors
## The degrees of freedom for just the general factor are 299 and the fit is 2.39
## The number of observations was 1059 with Chi Square = 2504.15 with prob < 0
## The root mean square of the residuals is 0.1
## The df corrected root mean square of the residuals is 0.1
##
## RMSEA index = 0.083 and the 10 % confidence intervals are 0.08 0.087
```

```
## BIC = 421.59
##
## Measures of factor score adequacy
##
##           g  F1*  F2*  F3*
## Correlation of scores with factors    0.77 0.72 0.59 0.82
## Multiple R square of scores with factors    0.59 0.53 0.35 0.67
## Minimum correlation of factor score estimates 0.18 0.05 -0.29 0.34
##
## Total, General and Subset omega for each subset
##
##           g  F1*  F2*  F3*
## Omega total for total scores and subscales    0.87 0.78 0.69 0.68
## Omega general for total scores and subscales    0.56 0.43 0.43 0.22
## Omega group for total scores and subscales    0.19 0.35 0.26 0.46

omega_disorganised <- omega(MSS_disorganised, plot = FALSE)
print(omega_disorganised)

## Omega
## Call: omegah(m = m, nfactors = nfactors, fm = fm, key = key, flip = flip,
##   digits = digits, title = title, sl = sl, labels = labels,
##   plot = plot, n.obs = n.obs, rotate = rotate, Phi = Phi, option = option,
##   covar = covar)
## Alpha:          0.92
## G.6:            0.93
## Omega Hierarchical: 0.73
## Omega H asymptotic: 0.78
## Omega Total      0.93
##
## Schmid Leiman Factor loadings greater than 0.2
##           g  F1*  F2*  F3*  h2  u2  p2
## DIS03 0.58          0.25 0.42 0.58 0.79
## DIS09 0.67          0.46 0.67 0.33 0.68
## DIS12 0.52          0.38 0.42 0.58 0.63
## DIS15 0.49 0.24          0.31 0.69 0.76
## DIS18 0.52          0.34 0.66 0.80
## DIS21 0.58 0.35          0.48 0.52 0.70
## DIS24 0.41          0.38          0.32 0.68 0.51
## DIS27 0.61 0.32          0.20 0.53 0.47 0.71
## DIS33 0.52 0.30          0.37 0.63 0.74
## DIS36 0.51          0.27          0.36 0.64 0.72
## DIS39 0.65 0.54          0.71 0.29 0.59
## DIS48 0.56          0.39          0.50 0.50 0.63
## DIS54 0.45          0.60          0.57 0.43 0.36
## DIS57 0.62 0.44          0.58 0.42 0.66
## DIS60 0.58 0.50          0.58 0.42 0.57
## DIS63 0.53          0.50          0.54 0.46 0.52
## DIS66 0.42          0.23 0.77 0.79
## DIS72 0.66 0.49          0.68 0.32 0.64
##
## With Sums of squares of:
##   g  F1*  F2*  F3*
## 5.51 1.49 1.06 0.54
##
## general/max 3.7  max/min = 2.76
```

```
## mean percent general = 0.66    with sd = 0.12 and cv of 0.18
## Explained Common Variance of the general factor = 0.64
##
## The degrees of freedom are 102 and the fit is 0.63
## The number of observations was 1059 with Chi Square = 656.89 with prob < 5.9e-82
## The root mean square of the residuals is 0.03
## The df corrected root mean square of the residuals is 0.04
## RMSEA index = 0.072 and the 10 % confidence intervals are 0.067 0.077
## BIC = -53.55
##
## Compare this with the adequacy of just a general factor and no group factors
## The degrees of freedom for just the general factor are 135 and the fit is 1.85
## The number of observations was 1059 with Chi Square = 1948.4 with prob < 1.099998e-318
## The root mean square of the residuals is 0.11
## The df corrected root mean square of the residuals is 0.11
##
## RMSEA index = 0.113 and the 10 % confidence intervals are 0.108 0.117
## BIC = 1008.11
##
## Measures of factor score adequacy
##
##           g  F1*  F2*  F3*
## Correlation of scores with factors      0.86 0.71 0.76 0.61
## Multiple R square of scores with factors 0.74 0.50 0.58 0.38
## Minimum correlation of factor score estimates 0.49 0.01 0.15 -0.25
##
## Total, General and Subset omega for each subset
##
##           g  F1*  F2*  F3*
## Omega total for total scores and subscales 0.93 0.88 0.79 0.75
## Omega general for total scores and subscales 0.73 0.62 0.45 0.58
## Omega group for total scores and subscales 0.13 0.26 0.34 0.17

omega_disconnected <- omega(MSS_disconnected, plot = FALSE)
print(omega_disconnected)

## Omega
## Call: omegah(m = m, nfactors = nfactors, fm = fm, key = key, flip = flip,
## digits = digits, title = title, sl = sl, labels = labels,
## plot = plot, n.obs = n.obs, rotate = rotate, Phi = Phi, option = option,
## covar = covar)
## Alpha:          0.74
## G.6:            0.73
## Omega Hierarchical: 0.72
## Omega H asymptotic: 0.89
## Omega Total      0.81
##
## Schmid Leiman Factor loadings greater than 0.2
##           g  F1*  F2*  F3*  h2  u2  p2
## NEG13 0.37                0.14 0.86 0.97
## DIS06 0.51                0.27 0.73 0.98
## DIS30 0.58                0.81 1.00 0.00 0.34
## DIS42 0.55                0.31 0.69 0.99
## DIS45 0.64                0.41 0.59 1.00
## DIS51 0.60                0.37 0.63 0.98
## DIS69 0.60          0.80      1.00 0.00 0.36
```

```
## DIS75 0.22          0.29          0.14 0.86 0.35
##
## With Sums of squares of:
##   g  F1*  F2*  F3*
## 2.21 0.00 0.73 0.67
##
## general/max 3.01   max/min =   170.25
## mean percent general = 0.75   with sd = 0.33 and cv of 0.44
## Explained Common Variance of the general factor = 0.61
##
## The degrees of freedom are 7 and the fit is 0.02
## The number of observations was 1059 with Chi Square = 18.32 with prob < 0.011
## The root mean square of the residuals is 0.02
## The df corrected root mean square of the residuals is 0.03
## RMSEA index = 0.039 and the 10 % confidence intervals are 0.018 0.061
## BIC = -30.43
##
## Compare this with the adequacy of just a general factor and no group factors
## The degrees of freedom for just the general factor are 20 and the fit is 0.12
## The number of observations was 1059 with Chi Square = 128.82 with prob < 6.5e-18
## The root mean square of the residuals is 0.05
## The df corrected root mean square of the residuals is 0.06
##
## RMSEA index = 0.072 and the 10 % confidence intervals are 0.06 0.084
## BIC = -10.48
##
## Measures of factor score adequacy
##
##               g  F1*  F2*  F3*
## Correlation of scores with factors      0.87 0.06 0.93 0.93
## Multiple R square of scores with factors 0.76 0.00 0.86 0.87
## Minimum correlation of factor score estimates 0.52 -0.99 0.73 0.75
##
## Total, General and Subset omega for each subset
##
##               g  F1*  F2*  F3*
## Omega total for total scores and subscales 0.81 0.41 0.65 0.77
## Omega general for total scores and subscales 0.72 0.41 0.54 0.50
## Omega group for total scores and subscales 0.08 0.00 0.11 0.27
```

*# And the original model*

```
MSS_negative_orig <- MSS[, c("NEG01", "NEG04", "NEG07", "NEG10", "NEG13",
                             "NEG16", "NEG19", "NEG22", "NEG25", "NEG28",
                             "NEG31", "NEG34", "NEG37", "NEG40", "NEG43",
                             "NEG46", "NEG49", "NEG52", "NEG55", "NEG58",
                             "NEG61", "NEG64", "NEG67", "NEG70", "NEG73",
                             "NEG76")]
MSS_positive_orig <- MSS[, c("POS02", "POS05", "POS08", "POS11", "POS14",
                             "POS17", "POS20", "POS23", "POS26", "POS29",
                             "POS32", "POS35", "POS38", "POS41", "POS44",
                             "POS47", "POS50", "POS53", "POS56", "POS59",
                             "POS62", "POS65", "POS68", "POS71", "POS74",
                             "POS77")]
MSS_disorganised_orig <- MSS[, c("DIS03", "DIS06", "DIS09", "DIS12", "DIS15",
                                 "DIS18", "DIS21", "DIS24", "DIS27", "DIS30",
                                 "DIS33", "DIS36", "DIS39", "DIS42", "DIS45",
```

```
                                "DIS48", "DIS51", "DIS54", "DIS57", "DIS60",
                                "DIS63", "DIS66", "DIS69", "DIS72", "DIS75"]
alpha(MSS_negative_orig)
```

```
##
## Reliability analysis
## Call: alpha(x = MSS_negative_orig)
##
##      raw_alpha std.alpha G6(smc) average_r S/N   ase mean   sd median_r
##      0.82      0.85      0.88      0.18 5.9 0.008  0.1 0.12      0.17
##
##      95% confidence boundaries
##              lower alpha upper
## Feldt      0.8  0.82  0.83
## Duhachek    0.8  0.82  0.83
##
## Reliability if an item is dropped:
##      raw_alpha std.alpha G6(smc) average_r S/N alpha se  var.r med.r
## NEG01      0.81      0.85      0.87      0.18 5.6  0.0083 0.0110  0.17
## NEG04      0.82      0.85      0.88      0.19 5.9  0.0078 0.0105  0.18
## NEG07      0.81      0.85      0.87      0.18 5.7  0.0084 0.0111  0.17
## NEG10      0.81      0.85      0.87      0.18 5.5  0.0084 0.0111  0.17
## NEG13      0.81      0.85      0.88      0.19 5.8  0.0081 0.0110  0.17
## NEG16      0.81      0.85      0.87      0.18 5.6  0.0083 0.0101  0.17
## NEG19      0.81      0.85      0.88      0.19 5.7  0.0082 0.0108  0.18
## NEG22      0.81      0.85      0.87      0.19 5.7  0.0083 0.0111  0.17
## NEG25      0.81      0.85      0.87      0.18 5.5  0.0083 0.0097  0.17
## NEG28      0.81      0.85      0.87      0.18 5.7  0.0083 0.0111  0.17
## NEG31      0.81      0.85      0.87      0.18 5.7  0.0082 0.0111  0.17
## NEG34      0.81      0.85      0.88      0.19 5.8  0.0082 0.0109  0.18
## NEG37      0.81      0.85      0.87      0.18 5.7  0.0082 0.0106  0.17
## NEG40      0.81      0.85      0.87      0.18 5.6  0.0083 0.0103  0.17
## NEG43      0.81      0.85      0.87      0.18 5.5  0.0083 0.0104  0.17
## NEG46      0.81      0.85      0.87      0.18 5.6  0.0085 0.0111  0.17
## NEG49      0.82      0.85      0.88      0.19 5.9  0.0078 0.0104  0.18
## NEG52      0.81      0.85      0.87      0.18 5.6  0.0083 0.0108  0.17
## NEG55      0.81      0.84      0.87      0.18 5.4  0.0083 0.0096  0.17
## NEG58      0.81      0.85      0.88      0.19 5.8  0.0081 0.0108  0.18
## NEG61      0.81      0.85      0.87      0.18 5.5  0.0083 0.0105  0.17
## NEG64      0.81      0.85      0.87      0.18 5.5  0.0083 0.0096  0.17
## NEG67      0.80      0.84      0.87      0.18 5.4  0.0086 0.0098  0.17
## NEG70      0.81      0.85      0.87      0.18 5.5  0.0082 0.0106  0.17
## NEG73      0.81      0.84      0.87      0.18 5.4  0.0083 0.0092  0.17
## NEG76      0.81      0.85      0.88      0.19 5.7  0.0082 0.0110  0.17
##
## Item statistics
##      n raw.r std.r r.cor r.drop mean  sd
## NEG01 1059  0.45  0.46  0.42  0.37 0.073 0.26
## NEG04 1059  0.42  0.30  0.25  0.27 0.530 0.50
## NEG07 1059  0.48  0.43  0.39  0.39 0.115 0.32
## NEG10 1059  0.53  0.53  0.51  0.47 0.048 0.21
## NEG13 1059  0.33  0.38  0.33  0.28 0.033 0.18
## NEG16 1059  0.47  0.50  0.48  0.41 0.065 0.25
```

```
## NEG19 1059 0.49 0.39 0.35 0.37 0.297 0.46
## NEG22 1059 0.51 0.43 0.39 0.39 0.254 0.44
## NEG25 1059 0.48 0.55 0.54 0.43 0.040 0.20
## NEG28 1059 0.43 0.43 0.39 0.35 0.081 0.27
## NEG31 1059 0.40 0.44 0.41 0.36 0.030 0.17
## NEG34 1059 0.39 0.37 0.33 0.32 0.061 0.24
## NEG37 1059 0.38 0.44 0.41 0.34 0.027 0.16
## NEG40 1059 0.43 0.47 0.45 0.37 0.044 0.21
## NEG43 1059 0.47 0.52 0.51 0.42 0.048 0.21
## NEG46 1059 0.52 0.50 0.47 0.45 0.087 0.28
## NEG49 1059 0.41 0.29 0.23 0.26 0.505 0.50
## NEG52 1059 0.46 0.50 0.47 0.40 0.055 0.23
## NEG55 1059 0.51 0.58 0.57 0.46 0.041 0.20
## NEG58 1059 0.32 0.34 0.28 0.26 0.042 0.20
## NEG61 1059 0.48 0.53 0.50 0.42 0.049 0.22
## NEG64 1059 0.45 0.52 0.51 0.40 0.038 0.19
## NEG67 1059 0.59 0.62 0.62 0.53 0.069 0.25
## NEG70 1059 0.46 0.52 0.50 0.43 0.019 0.14
## NEG73 1059 0.51 0.59 0.60 0.47 0.029 0.17
## NEG76 1059 0.37 0.41 0.37 0.32 0.031 0.17
##
## Non missing response frequency for each item
##      0      1 miss
## NEG01 0.93 0.07 0
## NEG04 0.47 0.53 0
## NEG07 0.88 0.12 0
## NEG10 0.95 0.05 0
## NEG13 0.97 0.03 0
## NEG16 0.93 0.07 0
## NEG19 0.70 0.30 0
## NEG22 0.75 0.25 0
## NEG25 0.96 0.04 0
## NEG28 0.92 0.08 0
## NEG31 0.97 0.03 0
## NEG34 0.94 0.06 0
## NEG37 0.97 0.03 0
## NEG40 0.96 0.04 0
## NEG43 0.95 0.05 0
## NEG46 0.91 0.09 0
## NEG49 0.49 0.51 0
## NEG52 0.95 0.05 0
## NEG55 0.96 0.04 0
## NEG58 0.96 0.04 0
## NEG61 0.95 0.05 0
## NEG64 0.96 0.04 0
## NEG67 0.93 0.07 0
## NEG70 0.98 0.02 0
## NEG73 0.97 0.03 0
## NEG76 0.97 0.03 0
```

```
alpha(MSS_positive_orig)
```

```
##
## Reliability analysis
```

```
## Call: alpha(x = MSS_positive_orig)
##
##      raw_alpha std.alpha G6(smc) average_r S/N      ase mean      sd median_r
##      0.84      0.85      0.87      0.18 5.9 0.0068 0.038 0.084      0.18
##
##      95% confidence boundaries
##              lower alpha upper
## Feldt      0.82 0.84 0.85
## Duhachek 0.83 0.84 0.85
##
## Reliability if an item is dropped:
##      raw_alpha std.alpha G6(smc) average_r S/N alpha se var.r med.r
## POS02      0.83      0.85      0.87      0.18 5.6 0.0070 0.0065 0.18
## POS05      0.84      0.85      0.87      0.19 5.8 0.0069 0.0063 0.18
## POS08      0.84      0.85      0.87      0.19 5.8 0.0069 0.0058 0.18
## POS11      0.83      0.85      0.87      0.18 5.6 0.0071 0.0065 0.18
## POS14      0.84      0.85      0.87      0.19 5.8 0.0069 0.0066 0.18
## POS17      0.83      0.85      0.87      0.18 5.7 0.0070 0.0067 0.18
## POS20      0.83      0.85      0.87      0.18 5.7 0.0071 0.0065 0.18
## POS23      0.83      0.85      0.87      0.19 5.7 0.0070 0.0065 0.18
## POS26      0.83      0.85      0.87      0.18 5.7 0.0070 0.0068 0.18
## POS29      0.83      0.85      0.87      0.18 5.6 0.0072 0.0065 0.18
## POS32      0.83      0.85      0.87      0.18 5.6 0.0070 0.0062 0.18
## POS35      0.84      0.85      0.87      0.19 5.8 0.0069 0.0064 0.18
## POS38      0.84      0.85      0.87      0.19 5.8 0.0069 0.0062 0.18
## POS41      0.83      0.85      0.87      0.18 5.6 0.0071 0.0066 0.18
## POS44      0.83      0.85      0.87      0.19 5.7 0.0070 0.0066 0.18
## POS47      0.83      0.85      0.87      0.18 5.7 0.0070 0.0067 0.18
## POS50      0.83      0.85      0.87      0.18 5.6 0.0070 0.0064 0.18
## POS53      0.83      0.85      0.87      0.18 5.6 0.0071 0.0063 0.18
## POS56      0.83      0.85      0.87      0.18 5.6 0.0071 0.0068 0.18
## POS59      0.83      0.85      0.87      0.18 5.7 0.0070 0.0065 0.18
## POS62      0.83      0.85      0.87      0.18 5.5 0.0073 0.0065 0.18
## POS65      0.84      0.85      0.87      0.19 5.7 0.0069 0.0058 0.18
## POS68      0.83      0.85      0.87      0.18 5.6 0.0070 0.0064 0.18
## POS71      0.83      0.85      0.87      0.18 5.7 0.0070 0.0067 0.18
## POS74      0.83      0.85      0.87      0.18 5.5 0.0071 0.0065 0.18
## POS77      0.83      0.85      0.87      0.18 5.7 0.0070 0.0065 0.18
##
## Item statistics
##      n raw.r std.r r.cor r.drop mean sd
## POS02 1059 0.47 0.50 0.48 0.41 0.0274 0.163
## POS05 1059 0.41 0.37 0.33 0.32 0.0510 0.220
## POS08 1059 0.29 0.36 0.33 0.26 0.0066 0.081
## POS11 1059 0.50 0.52 0.50 0.44 0.0246 0.155
## POS14 1059 0.32 0.35 0.30 0.27 0.0132 0.114
## POS17 1059 0.44 0.46 0.43 0.39 0.0161 0.126
## POS20 1059 0.55 0.47 0.44 0.43 0.1161 0.321
## POS23 1059 0.42 0.46 0.43 0.38 0.0161 0.126
## POS26 1059 0.53 0.47 0.44 0.42 0.1058 0.308
## POS29 1059 0.54 0.48 0.45 0.45 0.0689 0.253
## POS32 1059 0.44 0.48 0.46 0.40 0.0123 0.110
## POS35 1059 0.35 0.36 0.31 0.29 0.0170 0.129
## POS38 1059 0.35 0.40 0.36 0.30 0.0132 0.114
```

```
## POS41 1059 0.52 0.50 0.48 0.44 0.0500 0.218
## POS44 1059 0.53 0.45 0.41 0.41 0.1152 0.319
## POS47 1059 0.47 0.47 0.44 0.40 0.0321 0.176
## POS50 1059 0.47 0.50 0.47 0.40 0.0331 0.179
## POS53 1059 0.50 0.52 0.51 0.44 0.0274 0.163
## POS56 1059 0.50 0.52 0.49 0.44 0.0283 0.166
## POS59 1059 0.47 0.47 0.44 0.39 0.0415 0.200
## POS62 1059 0.61 0.55 0.53 0.51 0.0916 0.289
## POS65 1059 0.35 0.43 0.41 0.31 0.0076 0.087
## POS68 1059 0.45 0.50 0.47 0.41 0.0123 0.110
## POS71 1059 0.46 0.47 0.43 0.41 0.0208 0.143
## POS74 1059 0.51 0.55 0.53 0.46 0.0227 0.149
## POS77 1059 0.44 0.47 0.44 0.39 0.0161 0.126
```

```
##
## Non missing response frequency for each item
```

```
##      0      1 miss
## POS02 0.97 0.03 0
## POS05 0.95 0.05 0
## POS08 0.99 0.01 0
## POS11 0.98 0.02 0
## POS14 0.99 0.01 0
## POS17 0.98 0.02 0
## POS20 0.88 0.12 0
## POS23 0.98 0.02 0
## POS26 0.89 0.11 0
## POS29 0.93 0.07 0
## POS32 0.99 0.01 0
## POS35 0.98 0.02 0
## POS38 0.99 0.01 0
## POS41 0.95 0.05 0
## POS44 0.88 0.12 0
## POS47 0.97 0.03 0
## POS50 0.97 0.03 0
## POS53 0.97 0.03 0
## POS56 0.97 0.03 0
## POS59 0.96 0.04 0
## POS62 0.91 0.09 0
## POS65 0.99 0.01 0
## POS68 0.99 0.01 0
## POS71 0.98 0.02 0
## POS74 0.98 0.02 0
## POS77 0.98 0.02 0
```

```
alpha(MSS_disorganised_orig)
```

```
##
## Reliability analysis
## Call: alpha(x = MSS_disorganised_orig)
##
##      raw_alpha std.alpha G6(smc) average_r S/N      ase mean   sd median_r
##      0.91      0.93      0.94      0.34  13 0.0039 0.057 0.13      0.33
##
##      95% confidence boundaries
##      lower alpha upper
```

```
## Feldt      0.9  0.91  0.92
## Duhachek   0.9  0.91  0.92
##
## Reliability if an item is dropped:
##      raw_alpha std.alpha G6(smc) average_r S/N alpha se  var.r med.r
## DIS03      0.91      0.92      0.94      0.34 12   0.0041 0.0124  0.33
## DIS06      0.91      0.93      0.94      0.34 13   0.0040 0.0120  0.34
## DIS09      0.90      0.92      0.93      0.33 12   0.0042 0.0121  0.33
## DIS12      0.91      0.93      0.94      0.34 12   0.0040 0.0123  0.34
## DIS15      0.91      0.92      0.94      0.34 12   0.0041 0.0126  0.33
## DIS18      0.91      0.92      0.94      0.34 12   0.0041 0.0127  0.33
## DIS21      0.90      0.92      0.93      0.33 12   0.0041 0.0119  0.33
## DIS24      0.91      0.93      0.94      0.34 13   0.0038 0.0120  0.34
## DIS27      0.91      0.92      0.93      0.33 12   0.0041 0.0117  0.33
## DIS30      0.91      0.92      0.94      0.34 12   0.0041 0.0126  0.33
## DIS33      0.91      0.92      0.94      0.34 12   0.0040 0.0121  0.33
## DIS36      0.91      0.92      0.94      0.34 12   0.0041 0.0126  0.33
## DIS39      0.91      0.92      0.93      0.33 12   0.0041 0.0108  0.33
## DIS42      0.91      0.92      0.94      0.34 12   0.0040 0.0120  0.34
## DIS45      0.91      0.92      0.94      0.34 12   0.0040 0.0125  0.33
## DIS48      0.90      0.92      0.93      0.34 12   0.0042 0.0122  0.33
## DIS51      0.91      0.92      0.94      0.34 12   0.0040 0.0125  0.34
## DIS54      0.91      0.93      0.94      0.34 13   0.0040 0.0119  0.34
## DIS57      0.91      0.92      0.93      0.33 12   0.0041 0.0111  0.33
## DIS60      0.91      0.92      0.93      0.33 12   0.0041 0.0114  0.33
## DIS63      0.91      0.92      0.93      0.34 12   0.0041 0.0121  0.33
## DIS66      0.91      0.93      0.94      0.34 13   0.0040 0.0123  0.34
## DIS69      0.91      0.92      0.93      0.34 12   0.0040 0.0126  0.33
## DIS72      0.90      0.92      0.93      0.33 12   0.0041 0.0105  0.33
## DIS75      0.91      0.93      0.94      0.35 13   0.0037 0.0093  0.34
##
## Item statistics
##      n raw.r std.r r.cor r.drop mean  sd
## DIS03 1059  0.63  0.63  0.61  0.59 0.051 0.22
## DIS06 1059  0.47  0.48  0.45  0.42 0.041 0.20
## DIS09 1059  0.70  0.69  0.68  0.66 0.077 0.27
## DIS12 1059  0.58  0.54  0.51  0.50 0.128 0.33
## DIS15 1059  0.60  0.61  0.59  0.56 0.040 0.20
## DIS18 1059  0.63  0.62  0.60  0.58 0.062 0.24
## DIS21 1059  0.67  0.70  0.69  0.64 0.031 0.17
## DIS24 1059  0.55  0.48  0.45  0.45 0.193 0.39
## DIS27 1059  0.65  0.67  0.66  0.61 0.037 0.19
## DIS30 1059  0.58  0.58  0.55  0.53 0.050 0.22
## DIS33 1059  0.58  0.61  0.59  0.55 0.026 0.16
## DIS36 1059  0.61  0.59  0.57  0.55 0.069 0.25
## DIS39 1059  0.70  0.75  0.75  0.68 0.022 0.15
## DIS42 1059  0.55  0.60  0.58  0.52 0.018 0.13
## DIS45 1059  0.57  0.59  0.56  0.52 0.041 0.20
## DIS48 1059  0.66  0.65  0.63  0.62 0.068 0.25
## DIS51 1059  0.57  0.59  0.57  0.53 0.033 0.18
## DIS54 1059  0.57  0.51  0.49  0.49 0.144 0.35
## DIS57 1059  0.69  0.73  0.73  0.66 0.023 0.15
## DIS60 1059  0.64  0.68  0.68  0.61 0.025 0.15
## DIS63 1059  0.63  0.60  0.59  0.58 0.072 0.26
```

```
## DIS66 1059 0.48 0.51 0.47 0.45 0.021 0.14
## DIS69 1059 0.59 0.61 0.59 0.56 0.026 0.16
## DIS72 1059 0.74 0.78 0.78 0.71 0.023 0.15
## DIS75 1059 0.32 0.30 0.25 0.23 0.107 0.31
##
## Non missing response frequency for each item
##      0      1 miss
## DIS03 0.95 0.05 0
## DIS06 0.96 0.04 0
## DIS09 0.92 0.08 0
## DIS12 0.87 0.13 0
## DIS15 0.96 0.04 0
## DIS18 0.94 0.06 0
## DIS21 0.97 0.03 0
## DIS24 0.81 0.19 0
## DIS27 0.96 0.04 0
## DIS30 0.95 0.05 0
## DIS33 0.97 0.03 0
## DIS36 0.93 0.07 0
## DIS39 0.98 0.02 0
## DIS42 0.98 0.02 0
## DIS45 0.96 0.04 0
## DIS48 0.93 0.07 0
## DIS51 0.97 0.03 0
## DIS54 0.86 0.14 0
## DIS57 0.98 0.02 0
## DIS60 0.98 0.02 0
## DIS63 0.93 0.07 0
## DIS66 0.98 0.02 0
## DIS69 0.97 0.03 0
## DIS72 0.98 0.02 0
## DIS75 0.89 0.11 0
```

#### OXFORD-LIVERPOOL INVENTORY FOR FEELINGS AND EXPERIENCES

##### 1. CORRELATIONAL METHOD

```
mat.OLIFE <- cor(OLIFE, method = "spearman")
row.names(mat.OLIFE) <- colnames(mat.OLIFE)

# Get all pairs of columns of OLIFE
pairsOLIFE <- combn(ncol(OLIFE), 2)

# Calculate Phi coefficient for each pair
phi_matrix_OLIFE <- matrix(NA, ncol(OLIFE), ncol(OLIFE))
colnames(phi_matrix_OLIFE) <- colnames(OLIFE)
rownames(phi_matrix_OLIFE) <- colnames(OLIFE)

for (i in 1:ncol(pairsOLIFE)) {
  var1 <- pairsOLIFE[1, i]
  var2 <- pairsOLIFE[2, i]
  phi_valueOLIFE <- calculate_phi(OLIFE[[var1]], OLIFE[[var2]])
  phi_matrix_OLIFE[var1, var2] <- phi_valueOLIFE
  phi_matrix_OLIFE[var2, var1] <- phi_valueOLIFE
}

# Set diagonal to 1 as the Phi coefficient of a variable with itself is 1
diag(phi_matrix_OLIFE) <- 1

# Display the Phi coefficient matrix
head(print(phi_matrix_OLIFE))
```

| ## | COG001 | COG008 | COG016 | COG021 | COG022 | COG023 | COG028 | COG032 | COG036 | COG037 |
| --- | --- | --- | --- | --- | --- | --- | --- | --- | --- | --- |
| ## COG001 | 1.00 | 0.23 | 0.28 | 0.23 | 0.12 | 0.21 | 0.28 | 0.18 | 0.22 | 0.16 |
| ## COG008 | 0.23 | 1.00 | 0.24 | 0.20 | 0.31 | 0.17 | 0.26 | 0.21 | 0.24 | 0.32 |
| ## COG016 | 0.28 | 0.24 | 1.00 | 0.16 | 0.22 | 0.26 | 0.32 | 0.24 | 0.26 | 0.17 |
| ## COG021 | 0.23 | 0.20 | 0.16 | 1.00 | 0.21 | 0.18 | 0.23 | 0.18 | 0.19 | 0.24 |
| ## COG022 | 0.12 | 0.31 | 0.22 | 0.21 | 1.00 | 0.14 | 0.26 | 0.24 | 0.25 | 0.32 |
| ## COG023 | 0.21 | 0.17 | 0.26 | 0.18 | 0.14 | 1.00 | 0.49 | 0.27 | 0.22 | 0.20 |
| ## COG028 | 0.28 | 0.26 | 0.32 | 0.23 | 0.26 | 0.49 | 1.00 | 0.33 | 0.33 | 0.28 |
| ## COG032 | 0.18 | 0.21 | 0.24 | 0.18 | 0.24 | 0.27 | 0.33 | 1.00 | 0.23 | 0.23 |
| ## COG036 | 0.22 | 0.24 | 0.26 | 0.19 | 0.25 | 0.22 | 0.33 | 0.23 | 1.00 | 0.20 |
| ## COG037 | 0.16 | 0.32 | 0.17 | 0.24 | 0.32 | 0.20 | 0.28 | 0.23 | 0.20 | 1.00 |
| ## COG038 | 0.32 | 0.22 | 0.21 | 0.19 | 0.16 | 0.22 | 0.28 | 0.19 | 0.16 | 0.19 |
| ## COG043 | 0.16 | 0.23 | 0.22 | 0.22 | 0.36 | 0.18 | 0.26 | 0.22 | 0.17 | 0.28 |
| ## COG054 | 0.13 | 0.28 | 0.21 | 0.18 | 0.25 | 0.18 | 0.28 | 0.25 | 0.24 | 0.28 |
| ## COG055 | 0.19 | 0.14 | 0.34 | 0.11 | 0.20 | 0.23 | 0.28 | 0.22 | 0.24 | 0.14 |
| ## COG061 | 0.19 | 0.30 | 0.20 | 0.21 | 0.41 | 0.19 | 0.23 | 0.23 | 0.26 | 0.25 |
| ## COG062 | 0.24 | 0.25 | 0.24 | 0.34 | 0.33 | 0.25 | 0.35 | 0.32 | 0.32 | 0.26 |
| ## COG066 | 0.29 | 0.17 | 0.43 | 0.07 | 0.10 | 0.28 | 0.30 | 0.19 | 0.22 | 0.13 |
| ## COG073 | 0.21 | 0.26 | 0.28 | 0.11 | 0.20 | 0.22 | 0.29 | 0.30 | 0.20 | 0.24 |
| ## COG075 | 0.19 | 0.26 | 0.23 | 0.15 | 0.30 | 0.16 | 0.29 | 0.31 | 0.21 | 0.32 |
| ## COG081 | 0.19 | 0.29 | 0.28 | 0.19 | 0.30 | 0.23 | 0.37 | 0.31 | 0.26 | 0.26 |

|  |  |  |  |  |  |  |  |  |  |  |
| --- | --- | --- | --- | --- | --- | --- | --- | --- | --- | --- |
| ## COG085 | 0.20 | 0.15 | 0.21 | 0.15 | 0.20 | 0.24 | 0.23 | 0.19 | 0.34 | 0.23 |
| ## COG091 | 0.23 | 0.27 | 0.32 | 0.17 | 0.16 | 0.28 | 0.30 | 0.23 | 0.21 | 0.24 |
| ## COG095 | 0.19 | 0.16 | 0.17 | 0.14 | 0.30 | 0.13 | 0.25 | 0.21 | 0.18 | 0.21 |
| ## COG101 | 0.18 | 0.28 | 0.21 | 0.28 | 0.32 | 0.23 | 0.27 | 0.26 | 0.19 | 0.34 |
| ## IMP002 | 0.12 | 0.18 | 0.22 | 0.06 | 0.22 | 0.19 | 0.22 | 0.20 | 0.14 | 0.19 |
| ## IMP007 | -0.29 | -0.09 | -0.09 | -0.14 | 0.03 | -0.07 | -0.13 | -0.02 | -0.09 | 0.03 |
| ## IMP010 | 0.02 | 0.18 | 0.09 | 0.07 | 0.16 | 0.03 | 0.07 | 0.03 | 0.06 | 0.19 |
| ## IMP012 | 0.17 | 0.16 | 0.25 | 0.14 | 0.31 | 0.12 | 0.24 | 0.30 | 0.17 | 0.24 |
| ## IMP013 | 0.02 | 0.04 | 0.00 | -0.02 | 0.03 | 0.01 | 0.05 | -0.05 | 0.05 | 0.03 |
| ## IMP015 | 0.09 | 0.17 | 0.15 | 0.11 | 0.22 | 0.18 | 0.17 | 0.18 | 0.21 | 0.17 |
| ## IMP018 | -0.10 | -0.10 | -0.10 | -0.08 | -0.06 | -0.04 | -0.11 | -0.03 | -0.08 | -0.08 |
| ## IMP019 | 0.10 | 0.15 | 0.09 | 0.13 | 0.20 | 0.07 | 0.13 | 0.12 | 0.09 | 0.19 |
| ## IMP024 | 0.01 | 0.06 | 0.06 | -0.02 | 0.13 | 0.10 | 0.12 | 0.14 | 0.05 | 0.14 |
| ## IMP031 | 0.05 | 0.08 | 0.10 | 0.04 | 0.07 | 0.09 | 0.07 | 0.11 | 0.11 | 0.10 |
| ## IMP051 | -0.05 | -0.04 | -0.04 | -0.01 | -0.02 | -0.11 | -0.07 | -0.04 | -0.06 | 0.00 |
| ## IMP052 | -0.03 | 0.05 | -0.03 | 0.06 | 0.15 | -0.02 | 0.04 | 0.06 | 0.05 | 0.19 |
| ## IMP053 | 0.13 | 0.19 | 0.21 | 0.13 | 0.21 | 0.20 | 0.23 | 0.27 | 0.17 | 0.20 |
| ## IMP056 | 0.11 | 0.16 | 0.20 | 0.12 | 0.19 | 0.18 | 0.24 | 0.27 | 0.22 | 0.22 |
| ## IMP057 | -0.27 | -0.06 | -0.11 | -0.10 | -0.03 | -0.14 | -0.15 | -0.09 | -0.13 | -0.09 |
| ## IMP068 | -0.02 | 0.01 | -0.06 | 0.04 | 0.08 | -0.01 | -0.03 | 0.03 | -0.04 | 0.03 |
| ## IMP069 | 0.00 | 0.04 | 0.06 | 0.02 | 0.05 | 0.10 | 0.11 | 0.04 | 0.07 | 0.08 |
| ## IMP070 | 0.04 | 0.11 | 0.08 | 0.07 | 0.11 | 0.08 | 0.12 | 0.07 | 0.12 | 0.16 |
| ## IMP074 | 0.08 | 0.12 | 0.11 | 0.07 | 0.16 | 0.11 | 0.12 | 0.12 | 0.13 | 0.15 |
| ## IMP084 | 0.01 | 0.04 | 0.03 | 0.03 | 0.09 | 0.08 | 0.03 | 0.07 | -0.01 | 0.15 |
| ## IMP088 | 0.00 | 0.10 | 0.10 | 0.10 | 0.12 | 0.05 | 0.09 | 0.08 | 0.08 | 0.13 |
| ## IMP096 | 0.14 | 0.24 | 0.20 | 0.12 | 0.27 | 0.16 | 0.23 | 0.19 | 0.14 | 0.24 |
| ## IMP100 | 0.11 | 0.18 | 0.18 | 0.11 | 0.28 | 0.15 | 0.19 | 0.16 | 0.11 | 0.20 |
| ## UNEX003 | 0.04 | 0.01 | 0.07 | 0.06 | 0.09 | 0.06 | 0.03 | 0.06 | 0.02 | 0.05 |
| ## UNEX005 | 0.09 | 0.09 | 0.18 | 0.11 | 0.23 | 0.10 | 0.19 | 0.18 | 0.15 | 0.10 |
| ## UNEX006 | 0.07 | 0.04 | 0.14 | 0.04 | 0.22 | 0.07 | 0.05 | 0.11 | 0.09 | 0.08 |
| ## UNEX017 | 0.15 | 0.18 | 0.17 | 0.15 | 0.34 | 0.18 | 0.21 | 0.26 | 0.18 | 0.28 |
| ## UNEX020 | 0.04 | 0.04 | 0.03 | 0.04 | 0.16 | 0.06 | 0.11 | 0.08 | 0.04 | 0.09 |
| ## UNEX025 | 0.06 | 0.13 | 0.08 | 0.08 | 0.20 | 0.09 | 0.09 | 0.17 | 0.11 | 0.18 |
| ## UNEX027 | 0.10 | 0.17 | 0.22 | 0.16 | 0.29 | 0.13 | 0.20 | 0.21 | 0.22 | 0.20 |
| ## UNEX029 | 0.08 | -0.02 | 0.10 | 0.02 | 0.16 | 0.08 | 0.09 | 0.07 | 0.03 | 0.09 |
| ## UNEX030 | 0.10 | 0.15 | 0.22 | 0.08 | 0.32 | 0.13 | 0.17 | 0.17 | 0.19 | 0.18 |
| ## UNEX033 | 0.10 | 0.15 | 0.11 | 0.12 | 0.11 | 0.10 | 0.14 | 0.16 | 0.15 | 0.19 |
| ## UNEX040 | 0.04 | 0.05 | 0.10 | 0.07 | 0.20 | 0.06 | 0.10 | 0.09 | 0.08 | 0.13 |
| ## UNEX044 | 0.05 | 0.12 | 0.12 | 0.09 | 0.19 | 0.07 | 0.06 | 0.09 | 0.11 | 0.11 |
| ## UNEX047 | 0.17 | 0.16 | 0.16 | 0.27 | 0.23 | 0.17 | 0.20 | 0.18 | 0.20 | 0.16 |
| ## UNEX058 | 0.12 | 0.07 | 0.13 | 0.11 | 0.15 | 0.12 | 0.20 | 0.12 | 0.10 | 0.14 |
| ## UNEX060 | 0.02 | 0.10 | 0.11 | 0.08 | 0.19 | 0.09 | 0.11 | 0.13 | 0.10 | 0.16 |
| ## UNEX063 | 0.11 | 0.11 | 0.15 | 0.10 | 0.21 | 0.11 | 0.18 | 0.15 | 0.17 | 0.15 |
| ## UNEX065 | 0.04 | 0.09 | 0.12 | 0.05 | 0.15 | 0.05 | 0.12 | 0.13 | 0.12 | 0.15 |
| ## UNEX071 | 0.07 | 0.09 | 0.08 | 0.11 | 0.13 | 0.07 | 0.08 | 0.08 | 0.15 | 0.12 |
| ## UNEX072 | 0.05 | 0.10 | 0.10 | 0.09 | 0.21 | 0.11 | 0.18 | 0.14 | 0.14 | 0.15 |
| ## UNEX076 | 0.02 | 0.07 | 0.03 | 0.04 | 0.09 | 0.06 | 0.06 | 0.01 | 0.06 | 0.05 |
| ## UNEX078 | 0.12 | 0.16 | 0.17 | 0.20 | 0.24 | 0.19 | 0.22 | 0.16 | 0.20 | 0.21 |
| ## UNEX079 | 0.10 | 0.07 | 0.13 | 0.02 | 0.11 | 0.09 | 0.13 | 0.12 | 0.11 | 0.09 |
| ## UNEX082 | 0.06 | 0.12 | 0.09 | 0.09 | 0.20 | 0.09 | 0.11 | 0.06 | 0.13 | 0.15 |
| ## UNEX083 | 0.14 | 0.18 | 0.17 | 0.17 | 0.28 | 0.22 | 0.24 | 0.22 | 0.15 | 0.21 |
| ## UNEX087 | 0.13 | 0.19 | 0.12 | 0.16 | 0.29 | 0.13 | 0.14 | 0.14 | 0.20 | 0.14 |
| ## UNEX089 | 0.10 | 0.15 | 0.14 | 0.13 | 0.19 | 0.18 | 0.21 | 0.16 | 0.18 | 0.19 |
| ## UNEX090 | 0.07 | 0.18 | 0.11 | 0.06 | 0.20 | 0.07 | 0.13 | 0.19 | 0.14 | 0.18 |

|  |  |  |  |  |  |  |  |  |  |  |
| --- | --- | --- | --- | --- | --- | --- | --- | --- | --- | --- |
| ## UNEX093 | 0.05 | 0.05 | 0.05 | 0.18 | 0.16 | 0.10 | 0.12 | 0.10 | 0.08 | 0.13 |
| ## UNEX099 | 0.07 | 0.13 | 0.20 | 0.01 | 0.24 | 0.12 | 0.16 | 0.17 | 0.18 | 0.14 |
| ## UNEX103 | 0.02 | 0.02 | 0.02 | -0.02 | 0.06 | 0.03 | 0.04 | 0.11 | 0.03 | 0.08 |
| ## INT004 | 0.12 | 0.14 | 0.10 | 0.09 | 0.08 | 0.10 | 0.14 | 0.13 | 0.13 | 0.19 |
| ## INT009 | 0.11 | 0.04 | 0.06 | 0.07 | 0.03 | 0.04 | 0.04 | 0.00 | 0.06 | 0.08 |
| ## INT011 | 0.08 | 0.07 | 0.04 | 0.02 | 0.04 | 0.04 | 0.07 | 0.06 | 0.08 | 0.05 |
| ## INT014 | 0.15 | 0.20 | 0.15 | 0.09 | 0.15 | 0.12 | 0.20 | 0.13 | 0.14 | 0.28 |
| ## INT026 | 0.08 | 0.10 | 0.10 | 0.15 | 0.17 | 0.11 | 0.15 | 0.10 | 0.08 | 0.23 |
| ## INT034 | 0.05 | 0.05 | 0.06 | 0.05 | -0.01 | -0.04 | 0.02 | 0.06 | 0.03 | 0.09 |
| ## INT035 | 0.19 | 0.14 | 0.13 | 0.15 | 0.03 | 0.10 | 0.17 | 0.01 | 0.12 | 0.04 |
| ## INT039 | 0.01 | 0.05 | 0.00 | -0.03 | 0.05 | 0.01 | -0.04 | 0.02 | -0.01 | 0.09 |
| ## INT041 | 0.07 | 0.06 | 0.06 | 0.03 | 0.09 | 0.02 | 0.08 | 0.05 | 0.07 | 0.10 |
| ## INT042 | -0.05 | -0.03 | -0.07 | -0.01 | -0.05 | -0.08 | -0.10 | -0.10 | -0.10 | -0.02 |
| ## INT045 | 0.05 | 0.07 | 0.03 | 0.06 | 0.11 | 0.03 | 0.08 | 0.04 | 0.01 | 0.12 |
| ## INT046 | 0.03 | 0.08 | 0.01 | 0.05 | 0.06 | 0.06 | 0.07 | 0.07 | 0.04 | 0.12 |
| ## INT048 | 0.20 | 0.17 | 0.21 | 0.22 | 0.23 | 0.18 | 0.21 | 0.18 | 0.17 | 0.27 |
| ## INT049 | 0.15 | 0.20 | 0.12 | 0.05 | 0.12 | 0.04 | 0.16 | 0.09 | 0.09 | 0.17 |
| ## INT050 | 0.18 | 0.09 | 0.07 | 0.10 | 0.04 | 0.06 | 0.15 | 0.06 | 0.08 | 0.12 |
| ## INT059 | 0.11 | 0.21 | 0.13 | 0.09 | 0.07 | 0.10 | 0.17 | 0.14 | 0.15 | 0.19 |
| ## INT064 | 0.12 | 0.14 | 0.09 | 0.09 | 0.12 | 0.08 | 0.16 | 0.05 | 0.09 | 0.21 |
| ## INT067 | 0.09 | 0.08 | 0.08 | 0.06 | 0.06 | 0.04 | 0.02 | 0.04 | 0.06 | 0.07 |
| ## INT077 | 0.13 | 0.04 | 0.07 | 0.07 | 0.04 | 0.06 | 0.08 | -0.01 | 0.04 | 0.06 |
| ## INT080 | 0.12 | 0.12 | 0.10 | 0.13 | 0.21 | 0.09 | 0.18 | 0.14 | 0.08 | 0.24 |
| ## INT086 | 0.22 | 0.07 | 0.12 | 0.17 | 0.04 | 0.13 | 0.14 | 0.04 | 0.11 | 0.12 |
| ## INT092 | 0.00 | 0.00 | -0.07 | 0.05 | 0.01 | -0.04 | 0.04 | 0.00 | -0.02 | 0.03 |
| ## INT094 | 0.07 | 0.11 | 0.08 | 0.04 | 0.05 | 0.08 | 0.06 | 0.03 | 0.11 | 0.11 |
| ## INT097 | 0.21 | 0.09 | 0.01 | 0.18 | 0.05 | 0.03 | 0.08 | 0.04 | 0.07 | 0.10 |
| ## INT098 | 0.11 | 0.10 | 0.10 | 0.08 | 0.08 | 0.06 | 0.12 | 0.09 | 0.07 | 0.18 |
| ## INT102 | 0.14 | 0.13 | 0.08 | 0.12 | 0.14 | 0.06 | 0.13 | 0.14 | 0.09 | 0.18 |
| ## INT104 | 0.17 | 0.09 | 0.08 | 0.13 | 0.07 | 0.14 | 0.14 | 0.10 | 0.06 | 0.09 |
| ## | COG038 | COG043 | COG054 | COG055 | COG061 | COG062 | COG066 | COG073 | COG075 | COG081 |
| ## COG001 | 0.32 | 0.16 | 0.13 | 0.19 | 0.19 | 0.24 | 0.29 | 0.21 | 0.19 | 0.19 |
| ## COG008 | 0.22 | 0.23 | 0.28 | 0.14 | 0.30 | 0.25 | 0.17 | 0.26 | 0.26 | 0.29 |
| ## COG016 | 0.21 | 0.22 | 0.21 | 0.34 | 0.20 | 0.24 | 0.43 | 0.28 | 0.23 | 0.28 |
| ## COG021 | 0.19 | 0.22 | 0.18 | 0.11 | 0.21 | 0.34 | 0.07 | 0.11 | 0.15 | 0.19 |
| ## COG022 | 0.16 | 0.36 | 0.25 | 0.20 | 0.41 | 0.33 | 0.10 | 0.20 | 0.30 | 0.30 |
| ## COG023 | 0.22 | 0.18 | 0.18 | 0.23 | 0.19 | 0.25 | 0.28 | 0.22 | 0.16 | 0.23 |
| ## COG028 | 0.28 | 0.26 | 0.28 | 0.28 | 0.23 | 0.35 | 0.30 | 0.29 | 0.29 | 0.37 |
| ## COG032 | 0.19 | 0.22 | 0.25 | 0.22 | 0.23 | 0.32 | 0.19 | 0.30 | 0.31 | 0.31 |
| ## COG036 | 0.16 | 0.17 | 0.24 | 0.24 | 0.26 | 0.32 | 0.22 | 0.20 | 0.21 | 0.26 |
| ## COG037 | 0.19 | 0.28 | 0.28 | 0.14 | 0.25 | 0.26 | 0.13 | 0.24 | 0.32 | 0.26 |
| ## COG038 | 1.00 | 0.15 | 0.18 | 0.16 | 0.17 | 0.23 | 0.23 | 0.20 | 0.22 | 0.19 |
| ## COG043 | 0.15 | 1.00 | 0.31 | 0.18 | 0.30 | 0.29 | 0.13 | 0.31 | 0.36 | 0.25 |
| ## COG054 | 0.18 | 0.31 | 1.00 | 0.23 | 0.29 | 0.25 | 0.17 | 0.29 | 0.41 | 0.33 |
| ## COG055 | 0.16 | 0.18 | 0.23 | 1.00 | 0.22 | 0.19 | 0.25 | 0.23 | 0.22 | 0.28 |
| ## COG061 | 0.17 | 0.30 | 0.29 | 0.22 | 1.00 | 0.30 | 0.16 | 0.21 | 0.27 | 0.25 |
| ## COG062 | 0.23 | 0.29 | 0.25 | 0.19 | 0.30 | 1.00 | 0.16 | 0.22 | 0.23 | 0.27 |
| ## COG066 | 0.23 | 0.13 | 0.17 | 0.25 | 0.16 | 0.16 | 1.00 | 0.18 | 0.17 | 0.21 |
| ## COG073 | 0.20 | 0.31 | 0.29 | 0.23 | 0.21 | 0.22 | 0.18 | 1.00 | 0.48 | 0.34 |
| ## COG075 | 0.22 | 0.36 | 0.41 | 0.22 | 0.27 | 0.23 | 0.17 | 0.48 | 1.00 | 0.33 |
| ## COG081 | 0.19 | 0.25 | 0.33 | 0.28 | 0.25 | 0.27 | 0.21 | 0.34 | 0.33 | 1.00 |
| ## COG085 | 0.16 | 0.21 | 0.16 | 0.17 | 0.19 | 0.27 | 0.19 | 0.18 | 0.17 | 0.26 |
| ## COG091 | 0.24 | 0.18 | 0.18 | 0.21 | 0.21 | 0.22 | 0.32 | 0.23 | 0.19 | 0.19 |
| ## COG095 | 0.19 | 0.27 | 0.19 | 0.15 | 0.18 | 0.26 | 0.10 | 0.23 | 0.28 | 0.17 |

|  |  |  |  |  |  |  |  |  |  |  |
| --- | --- | --- | --- | --- | --- | --- | --- | --- | --- | --- |
| ## COG101 | 0.15 | 0.23 | 0.17 | 0.19 | 0.34 | 0.33 | 0.14 | 0.23 | 0.20 | 0.25 |
| ## IMP002 | 0.11 | 0.23 | 0.23 | 0.15 | 0.14 | 0.12 | 0.17 | 0.19 | 0.26 | 0.27 |
| ## IMP007 | -0.19 | 0.00 | -0.01 | -0.07 | -0.04 | -0.09 | -0.14 | -0.02 | 0.00 | -0.04 |
| ## IMP010 | 0.06 | 0.12 | 0.06 | 0.06 | 0.15 | 0.07 | 0.07 | 0.06 | 0.07 | 0.05 |
| ## IMP012 | 0.14 | 0.25 | 0.27 | 0.14 | 0.24 | 0.22 | 0.07 | 0.29 | 0.37 | 0.31 |
| ## IMP013 | 0.00 | 0.01 | 0.04 | 0.04 | 0.02 | -0.04 | 0.02 | -0.02 | 0.01 | 0.03 |
| ## IMP015 | 0.10 | 0.16 | 0.26 | 0.14 | 0.24 | 0.15 | 0.11 | 0.17 | 0.26 | 0.23 |
| ## IMP018 | -0.15 | -0.07 | -0.06 | -0.04 | -0.05 | -0.10 | -0.07 | -0.11 | -0.09 | -0.14 |
| ## IMP019 | 0.08 | 0.17 | 0.21 | 0.07 | 0.19 | 0.10 | 0.08 | 0.12 | 0.22 | 0.15 |
| ## IMP024 | 0.07 | 0.14 | 0.11 | 0.04 | 0.07 | 0.05 | 0.05 | 0.14 | 0.12 | 0.10 |
| ## IMP031 | 0.01 | 0.08 | 0.04 | 0.12 | 0.07 | 0.05 | 0.13 | 0.07 | 0.09 | 0.11 |
| ## IMP051 | -0.08 | -0.02 | -0.01 | -0.13 | -0.03 | -0.04 | -0.11 | -0.06 | 0.01 | -0.03 |
| ## IMP052 | 0.00 | 0.10 | 0.10 | -0.02 | 0.11 | 0.12 | -0.12 | 0.04 | 0.06 | 0.09 |
| ## IMP053 | 0.15 | 0.14 | 0.23 | 0.18 | 0.15 | 0.18 | 0.12 | 0.17 | 0.25 | 0.25 |
| ## IMP056 | 0.17 | 0.17 | 0.24 | 0.12 | 0.15 | 0.19 | 0.16 | 0.23 | 0.31 | 0.20 |
| ## IMP057 | -0.18 | -0.06 | -0.04 | -0.07 | -0.02 | -0.10 | -0.17 | -0.09 | -0.09 | -0.02 |
| ## IMP068 | -0.02 | 0.07 | 0.06 | -0.03 | 0.07 | 0.00 | -0.05 | 0.04 | 0.07 | 0.02 |
| ## IMP069 | 0.00 | -0.01 | 0.12 | 0.06 | 0.06 | 0.01 | 0.06 | 0.10 | 0.07 | 0.09 |
| ## IMP070 | 0.05 | 0.13 | 0.21 | 0.08 | 0.13 | 0.07 | 0.07 | 0.14 | 0.21 | 0.20 |
| ## IMP074 | 0.12 | 0.21 | 0.21 | 0.10 | 0.13 | 0.14 | 0.06 | 0.16 | 0.23 | 0.19 |
| ## IMP084 | 0.06 | 0.06 | 0.07 | -0.02 | 0.08 | 0.00 | 0.02 | 0.07 | 0.09 | 0.11 |
| ## IMP088 | 0.00 | 0.10 | 0.08 | 0.09 | 0.09 | 0.06 | 0.03 | 0.01 | 0.02 | 0.07 |
| ## IMP096 | 0.13 | 0.20 | 0.27 | 0.15 | 0.21 | 0.19 | 0.10 | 0.19 | 0.28 | 0.31 |
| ## IMP100 | 0.13 | 0.24 | 0.12 | 0.13 | 0.16 | 0.18 | 0.10 | 0.21 | 0.26 | 0.27 |
| ## UNEX003 | 0.04 | 0.09 | 0.04 | 0.10 | 0.08 | 0.04 | 0.02 | 0.04 | 0.08 | 0.12 |
| ## UNEX005 | 0.13 | 0.16 | 0.17 | 0.17 | 0.17 | 0.16 | 0.09 | 0.14 | 0.17 | 0.22 |
| ## UNEX006 | 0.04 | 0.18 | 0.07 | 0.12 | 0.21 | 0.09 | 0.00 | 0.05 | 0.13 | 0.17 |
| ## UNEX017 | 0.14 | 0.27 | 0.21 | 0.17 | 0.26 | 0.20 | 0.13 | 0.20 | 0.29 | 0.26 |
| ## UNEX020 | 0.01 | 0.12 | 0.10 | 0.08 | 0.11 | 0.08 | 0.00 | 0.11 | 0.14 | 0.13 |
| ## UNEX025 | 0.04 | 0.20 | 0.08 | 0.07 | 0.14 | 0.20 | 0.01 | 0.15 | 0.15 | 0.13 |
| ## UNEX027 | 0.06 | 0.20 | 0.14 | 0.17 | 0.25 | 0.20 | 0.07 | 0.13 | 0.20 | 0.22 |
| ## UNEX029 | 0.05 | 0.11 | 0.03 | 0.09 | 0.11 | 0.12 | 0.02 | 0.09 | 0.14 | 0.09 |
| ## UNEX030 | 0.14 | 0.29 | 0.21 | 0.23 | 0.24 | 0.24 | 0.15 | 0.18 | 0.27 | 0.25 |
| ## UNEX033 | 0.07 | 0.24 | 0.20 | 0.14 | 0.16 | 0.14 | 0.08 | 0.20 | 0.27 | 0.15 |
| ## UNEX040 | 0.02 | 0.12 | 0.15 | 0.10 | 0.09 | 0.08 | 0.00 | 0.12 | 0.17 | 0.13 |
| ## UNEX044 | 0.08 | 0.17 | 0.21 | 0.14 | 0.14 | 0.16 | 0.04 | 0.14 | 0.18 | 0.18 |
| ## UNEX047 | 0.17 | 0.16 | 0.17 | 0.16 | 0.22 | 0.23 | 0.14 | 0.13 | 0.21 | 0.24 |
| ## UNEX058 | 0.09 | 0.17 | 0.09 | 0.14 | 0.11 | 0.16 | 0.12 | 0.09 | 0.10 | 0.15 |
| ## UNEX060 | 0.02 | 0.16 | 0.15 | 0.10 | 0.13 | 0.14 | 0.05 | 0.19 | 0.24 | 0.25 |
| ## UNEX063 | 0.14 | 0.21 | 0.13 | 0.22 | 0.18 | 0.15 | 0.11 | 0.17 | 0.26 | 0.21 |
| ## UNEX065 | 0.09 | 0.14 | 0.12 | 0.12 | 0.11 | 0.10 | 0.04 | 0.13 | 0.18 | 0.16 |
| ## UNEX071 | 0.03 | 0.14 | 0.12 | 0.07 | 0.15 | 0.15 | 0.06 | 0.09 | 0.10 | 0.14 |
| ## UNEX072 | 0.07 | 0.21 | 0.13 | 0.11 | 0.12 | 0.17 | 0.03 | 0.14 | 0.23 | 0.16 |
| ## UNEX076 | 0.04 | 0.05 | 0.09 | 0.02 | 0.05 | 0.07 | 0.04 | 0.04 | 0.02 | 0.07 |
| ## UNEX078 | 0.13 | 0.22 | 0.21 | 0.14 | 0.19 | 0.26 | 0.10 | 0.14 | 0.21 | 0.25 |
| ## UNEX079 | 0.07 | 0.17 | 0.06 | 0.19 | 0.15 | 0.09 | 0.12 | 0.11 | 0.17 | 0.21 |
| ## UNEX082 | 0.11 | 0.15 | 0.12 | 0.10 | 0.11 | 0.15 | 0.07 | 0.09 | 0.23 | 0.10 |
| ## UNEX083 | 0.14 | 0.20 | 0.17 | 0.19 | 0.18 | 0.25 | 0.09 | 0.16 | 0.24 | 0.20 |
| ## UNEX087 | 0.12 | 0.23 | 0.19 | 0.08 | 0.21 | 0.23 | 0.09 | 0.13 | 0.26 | 0.22 |
| ## UNEX089 | 0.14 | 0.21 | 0.16 | 0.17 | 0.18 | 0.15 | 0.14 | 0.16 | 0.21 | 0.20 |
| ## UNEX090 | 0.14 | 0.18 | 0.15 | 0.18 | 0.14 | 0.10 | 0.10 | 0.16 | 0.24 | 0.21 |
| ## UNEX093 | 0.06 | 0.07 | 0.14 | 0.12 | 0.15 | 0.13 | 0.03 | 0.08 | 0.08 | 0.13 |
| ## UNEX099 | 0.11 | 0.19 | 0.17 | 0.25 | 0.21 | 0.18 | 0.14 | 0.18 | 0.22 | 0.28 |
| ## UNEX103 | 0.06 | 0.08 | 0.01 | 0.01 | 0.05 | 0.05 | 0.00 | 0.08 | 0.12 | 0.07 |

|  |  |  |  |  |  |  |  |  |  |  |  |
| --- | --- | --- | --- | --- | --- | --- | --- | --- | --- | --- | --- |
| ## | INT004 | 0.10 | 0.12 | 0.19 | 0.08 | 0.13 | 0.14 | 0.10 | 0.19 | 0.22 | 0.22 |
| ## | INT009 | 0.11 | 0.06 | 0.03 | 0.01 | -0.02 | 0.05 | 0.07 | 0.07 | 0.06 | 0.05 |
| ## | INT011 | 0.09 | -0.02 | 0.02 | 0.02 | 0.06 | 0.07 | 0.03 | 0.10 | 0.09 | 0.04 |
| ## | INT014 | 0.17 | 0.20 | 0.30 | 0.14 | 0.12 | 0.12 | 0.13 | 0.28 | 0.40 | 0.23 |
| ## | INT026 | 0.17 | 0.16 | 0.19 | 0.08 | 0.16 | 0.12 | 0.09 | 0.14 | 0.24 | 0.22 |
| ## | INT034 | 0.03 | 0.02 | 0.15 | -0.03 | 0.03 | 0.02 | 0.04 | 0.03 | 0.08 | 0.04 |
| ## | INT035 | 0.17 | 0.07 | 0.13 | 0.00 | 0.07 | 0.08 | 0.12 | 0.15 | 0.18 | 0.17 |
| ## | INT039 | 0.04 | -0.02 | 0.05 | -0.09 | -0.03 | -0.01 | 0.02 | 0.05 | 0.03 | 0.04 |
| ## | INT041 | 0.12 | 0.05 | 0.13 | 0.07 | 0.06 | 0.07 | 0.07 | 0.10 | 0.20 | 0.14 |
| ## | INT042 | -0.02 | -0.04 | 0.00 | -0.04 | -0.08 | -0.09 | -0.08 | -0.01 | -0.02 | -0.08 |
| ## | INT045 | 0.14 | 0.09 | 0.11 | 0.07 | 0.03 | 0.09 | 0.00 | 0.07 | 0.10 | 0.09 |
| ## | INT046 | 0.11 | 0.12 | 0.07 | 0.00 | 0.05 | 0.11 | 0.06 | 0.04 | 0.07 | 0.09 |
| ## | INT048 | 0.17 | 0.18 | 0.23 | 0.15 | 0.16 | 0.20 | 0.14 | 0.20 | 0.29 | 0.24 |
| ## | INT049 | 0.22 | 0.14 | 0.21 | 0.04 | 0.12 | 0.12 | 0.15 | 0.15 | 0.24 | 0.15 |
| ## | INT050 | 0.16 | 0.06 | 0.10 | 0.07 | 0.03 | 0.11 | 0.10 | 0.19 | 0.15 | 0.17 |
| ## | INT059 | 0.17 | 0.13 | 0.25 | 0.08 | 0.12 | 0.14 | 0.12 | 0.18 | 0.27 | 0.19 |
| ## | INT064 | 0.14 | 0.18 | 0.20 | 0.07 | 0.13 | 0.09 | 0.08 | 0.16 | 0.19 | 0.16 |
| ## | INT067 | 0.09 | 0.05 | 0.07 | 0.01 | 0.06 | 0.00 | 0.08 | 0.02 | 0.06 | 0.03 |
| ## | INT077 | 0.14 | 0.00 | 0.06 | 0.09 | 0.01 | 0.03 | 0.09 | 0.07 | 0.10 | 0.08 |
| ## | INT080 | 0.15 | 0.16 | 0.19 | 0.10 | 0.15 | 0.13 | 0.05 | 0.19 | 0.30 | 0.20 |
| ## | INT086 | 0.24 | 0.09 | 0.13 | 0.03 | 0.08 | 0.14 | 0.16 | 0.14 | 0.18 | 0.10 |
| ## | INT092 | -0.04 | 0.00 | 0.01 | -0.01 | -0.01 | 0.00 | 0.01 | 0.04 | 0.05 | 0.01 |
| ## | INT094 | 0.08 | 0.03 | 0.05 | 0.02 | 0.05 | 0.08 | 0.05 | 0.10 | 0.08 | 0.08 |
| ## | INT097 | 0.21 | 0.05 | 0.10 | 0.01 | 0.02 | 0.14 | 0.08 | 0.09 | 0.09 | 0.08 |
| ## | INT098 | 0.07 | 0.12 | 0.13 | 0.01 | 0.08 | 0.09 | 0.08 | 0.19 | 0.20 | 0.13 |
| ## | INT102 | 0.14 | 0.19 | 0.20 | 0.10 | 0.14 | 0.13 | 0.06 | 0.15 | 0.26 | 0.15 |
| ## | INT104 | 0.20 | 0.09 | 0.15 | 0.06 | 0.03 | 0.14 | 0.12 | 0.08 | 0.13 | 0.18 |
| ## |  | COG085 | COG091 | COG095 | COG101 | IMP002 | IMP007 | IMP010 | IMP012 | IMP013 | IMP015 |
| ## | COG001 | 0.20 | 0.23 | 0.19 | 0.18 | 0.12 | -0.29 | 0.02 | 0.17 | 0.02 | 0.09 |
| ## | COG008 | 0.15 | 0.27 | 0.16 | 0.28 | 0.18 | -0.09 | 0.18 | 0.16 | 0.04 | 0.17 |
| ## | COG016 | 0.21 | 0.32 | 0.17 | 0.21 | 0.22 | -0.09 | 0.09 | 0.25 | 0.00 | 0.15 |
| ## | COG021 | 0.15 | 0.17 | 0.14 | 0.28 | 0.06 | -0.14 | 0.07 | 0.14 | -0.02 | 0.11 |
| ## | COG022 | 0.20 | 0.16 | 0.30 | 0.32 | 0.22 | 0.03 | 0.16 | 0.31 | 0.03 | 0.22 |
| ## | COG023 | 0.24 | 0.28 | 0.13 | 0.23 | 0.19 | -0.07 | 0.03 | 0.12 | 0.01 | 0.18 |
| ## | COG028 | 0.23 | 0.30 | 0.25 | 0.27 | 0.22 | -0.13 | 0.07 | 0.24 | 0.05 | 0.17 |
| ## | COG032 | 0.19 | 0.23 | 0.21 | 0.26 | 0.20 | -0.02 | 0.03 | 0.30 | -0.05 | 0.18 |
| ## | COG036 | 0.34 | 0.21 | 0.18 | 0.19 | 0.14 | -0.09 | 0.06 | 0.17 | 0.05 | 0.21 |
| ## | COG037 | 0.23 | 0.24 | 0.21 | 0.34 | 0.19 | 0.03 | 0.19 | 0.24 | 0.03 | 0.17 |
| ## | COG038 | 0.16 | 0.24 | 0.19 | 0.15 | 0.11 | -0.19 | 0.06 | 0.14 | 0.00 | 0.10 |
| ## | COG043 | 0.21 | 0.18 | 0.27 | 0.23 | 0.23 | 0.00 | 0.12 | 0.25 | 0.01 | 0.16 |
| ## | COG054 | 0.16 | 0.18 | 0.19 | 0.17 | 0.23 | -0.01 | 0.06 | 0.27 | 0.04 | 0.26 |
| ## | COG055 | 0.17 | 0.21 | 0.15 | 0.19 | 0.15 | -0.07 | 0.06 | 0.14 | 0.04 | 0.14 |
| ## | COG061 | 0.19 | 0.21 | 0.18 | 0.34 | 0.14 | -0.04 | 0.15 | 0.24 | 0.02 | 0.24 |
| ## | COG062 | 0.27 | 0.22 | 0.26 | 0.33 | 0.12 | -0.09 | 0.07 | 0.22 | -0.04 | 0.15 |
| ## | COG066 | 0.19 | 0.32 | 0.10 | 0.14 | 0.17 | -0.14 | 0.07 | 0.07 | 0.02 | 0.11 |
| ## | COG073 | 0.18 | 0.23 | 0.23 | 0.23 | 0.19 | -0.02 | 0.06 | 0.29 | -0.02 | 0.17 |
| ## | COG075 | 0.17 | 0.19 | 0.28 | 0.20 | 0.26 | 0.00 | 0.07 | 0.37 | 0.01 | 0.26 |
| ## | COG081 | 0.26 | 0.19 | 0.17 | 0.25 | 0.27 | -0.04 | 0.05 | 0.31 | 0.03 | 0.23 |
| ## | COG085 | 1.00 | 0.19 | 0.18 | 0.23 | 0.16 | -0.04 | 0.02 | 0.17 | -0.01 | 0.15 |
| ## | COG091 | 0.19 | 1.00 | 0.18 | 0.20 | 0.12 | -0.15 | 0.16 | 0.13 | -0.07 | 0.09 |
| ## | COG095 | 0.18 | 0.18 | 1.00 | 0.22 | 0.18 | -0.04 | 0.09 | 0.26 | -0.01 | 0.13 |
| ## | COG101 | 0.23 | 0.20 | 0.22 | 1.00 | 0.17 | -0.01 | 0.10 | 0.17 | -0.02 | 0.15 |
| ## | IMP002 | 0.16 | 0.12 | 0.18 | 0.17 | 1.00 | 0.06 | 0.07 | 0.25 | 0.01 | 0.22 |
| ## | IMP007 | -0.04 | -0.15 | -0.04 | -0.01 | 0.06 | 1.00 | 0.03 | 0.01 | 0.00 | 0.00 |

|  |  |  |  |  |  |  |  |  |  |  |
| --- | --- | --- | --- | --- | --- | --- | --- | --- | --- | --- |
| ## IMP010 | 0.02 | 0.16 | 0.09 | 0.10 | 0.07 | 0.03 | 1.00 | 0.11 | 0.04 | 0.05 |
| ## IMP012 | 0.17 | 0.13 | 0.26 | 0.17 | 0.25 | 0.01 | 0.11 | 1.00 | 0.00 | 0.28 |
| ## IMP013 | -0.01 | -0.07 | -0.01 | -0.02 | 0.01 | 0.00 | 0.04 | 0.00 | 1.00 | 0.11 |
| ## IMP015 | 0.15 | 0.09 | 0.13 | 0.15 | 0.22 | 0.00 | 0.05 | 0.28 | 0.11 | 1.00 |
| ## IMP018 | -0.05 | -0.07 | -0.03 | -0.03 | -0.03 | 0.05 | 0.00 | -0.08 | -0.02 | -0.06 |
| ## IMP019 | 0.11 | 0.09 | 0.09 | 0.13 | 0.23 | -0.02 | 0.04 | 0.15 | 0.12 | 0.29 |
| ## IMP024 | 0.11 | 0.04 | 0.11 | 0.14 | 0.12 | 0.12 | 0.08 | 0.09 | 0.04 | 0.10 |
| ## IMP031 | 0.08 | 0.03 | 0.01 | 0.06 | 0.11 | 0.05 | 0.00 | 0.05 | 0.26 | 0.16 |
| ## IMP051 | -0.06 | -0.06 | -0.01 | -0.01 | -0.01 | 0.09 | -0.02 | 0.02 | -0.01 | 0.01 |
| ## IMP052 | 0.03 | 0.00 | 0.05 | 0.07 | 0.08 | 0.10 | 0.12 | 0.11 | 0.05 | 0.10 |
| ## IMP053 | 0.12 | 0.07 | 0.12 | 0.18 | 0.21 | -0.03 | 0.08 | 0.17 | 0.11 | 0.32 |
| ## IMP056 | 0.12 | 0.19 | 0.16 | 0.12 | 0.10 | -0.09 | -0.03 | 0.18 | 0.00 | 0.32 |
| ## IMP057 | -0.09 | -0.20 | -0.14 | -0.04 | 0.01 | 0.24 | 0.00 | 0.00 | 0.07 | -0.01 |
| ## IMP068 | -0.05 | -0.05 | -0.01 | 0.01 | 0.05 | 0.19 | 0.02 | 0.07 | 0.06 | 0.12 |
| ## IMP069 | 0.06 | 0.03 | 0.04 | 0.08 | 0.06 | 0.08 | 0.00 | 0.06 | 0.21 | 0.18 |
| ## IMP070 | 0.07 | -0.01 | 0.09 | 0.03 | 0.09 | 0.06 | 0.00 | 0.15 | 0.06 | 0.16 |
| ## IMP074 | 0.19 | 0.06 | 0.17 | 0.14 | 0.12 | 0.02 | 0.06 | 0.18 | 0.05 | 0.16 |
| ## IMP084 | 0.04 | 0.01 | 0.02 | 0.10 | 0.07 | 0.13 | 0.11 | 0.14 | 0.06 | 0.13 |
| ## IMP088 | 0.07 | 0.04 | 0.10 | 0.10 | 0.08 | 0.00 | 0.07 | 0.08 | 0.16 | 0.14 |
| ## IMP096 | 0.15 | 0.14 | 0.21 | 0.20 | 0.21 | 0.00 | 0.07 | 0.30 | 0.03 | 0.35 |
| ## IMP100 | 0.18 | 0.10 | 0.19 | 0.20 | 0.22 | 0.03 | 0.05 | 0.29 | -0.04 | 0.15 |
| ## UNEX003 | 0.07 | 0.00 | 0.06 | 0.03 | 0.04 | -0.02 | 0.02 | 0.12 | 0.01 | 0.03 |
| ## UNEX005 | 0.07 | 0.09 | 0.18 | 0.10 | 0.09 | -0.04 | 0.06 | 0.24 | 0.00 | 0.21 |
| ## UNEX006 | 0.12 | 0.00 | 0.08 | 0.11 | 0.08 | 0.04 | 0.05 | 0.23 | 0.01 | 0.11 |
| ## UNEX017 | 0.17 | 0.13 | 0.18 | 0.19 | 0.21 | -0.07 | 0.08 | 0.29 | 0.03 | 0.25 |
| ## UNEX020 | 0.07 | -0.03 | 0.07 | 0.08 | 0.12 | 0.05 | -0.06 | 0.11 | 0.03 | 0.08 |
| ## UNEX025 | 0.07 | 0.06 | 0.14 | 0.12 | 0.16 | 0.08 | 0.05 | 0.26 | 0.01 | 0.11 |
| ## UNEX027 | 0.18 | 0.10 | 0.21 | 0.17 | 0.11 | 0.01 | 0.07 | 0.27 | 0.03 | 0.21 |
| ## UNEX029 | 0.00 | 0.01 | 0.13 | 0.09 | -0.01 | 0.02 | 0.04 | 0.11 | -0.03 | 0.04 |
| ## UNEX030 | 0.13 | 0.11 | 0.23 | 0.13 | 0.16 | 0.01 | 0.08 | 0.28 | 0.03 | 0.18 |
| ## UNEX033 | 0.08 | 0.09 | 0.10 | 0.13 | 0.10 | -0.01 | 0.06 | 0.21 | 0.08 | 0.23 |
| ## UNEX040 | 0.16 | 0.03 | 0.16 | 0.09 | 0.09 | 0.03 | -0.01 | 0.22 | 0.01 | 0.11 |
| ## UNEX044 | 0.09 | 0.04 | 0.11 | 0.13 | 0.10 | 0.03 | 0.07 | 0.27 | 0.04 | 0.11 |
| ## UNEX047 | 0.15 | 0.14 | 0.15 | 0.20 | 0.11 | -0.07 | 0.06 | 0.16 | 0.02 | 0.17 |
| ## UNEX058 | 0.10 | 0.11 | 0.14 | 0.10 | 0.09 | 0.04 | 0.04 | 0.20 | -0.01 | 0.10 |
| ## UNEX060 | 0.08 | 0.03 | 0.17 | 0.11 | 0.11 | 0.06 | 0.05 | 0.29 | 0.07 | 0.19 |
| ## UNEX063 | 0.17 | 0.12 | 0.21 | 0.15 | 0.12 | -0.05 | 0.08 | 0.19 | 0.05 | 0.13 |
| ## UNEX065 | 0.05 | 0.05 | 0.12 | 0.08 | 0.11 | 0.02 | 0.03 | 0.14 | 0.04 | 0.12 |
| ## UNEX071 | 0.09 | 0.03 | 0.10 | 0.10 | 0.13 | -0.01 | 0.02 | 0.12 | 0.02 | 0.11 |
| ## UNEX072 | 0.11 | 0.07 | 0.18 | 0.13 | 0.04 | -0.02 | 0.00 | 0.18 | 0.02 | 0.22 |
| ## UNEX076 | 0.00 | 0.04 | 0.02 | 0.08 | 0.03 | -0.05 | 0.02 | -0.02 | 0.07 | 0.06 |
| ## UNEX078 | 0.15 | 0.10 | 0.26 | 0.20 | 0.19 | -0.04 | 0.05 | 0.23 | 0.04 | 0.19 |
| ## UNEX079 | 0.07 | 0.04 | 0.12 | 0.12 | 0.14 | 0.02 | 0.06 | 0.12 | 0.09 | 0.10 |
| ## UNEX082 | 0.11 | 0.10 | 0.16 | 0.11 | 0.09 | -0.02 | 0.04 | 0.10 | 0.04 | 0.20 |
| ## UNEX083 | 0.17 | 0.16 | 0.24 | 0.17 | 0.19 | -0.04 | 0.04 | 0.25 | 0.05 | 0.23 |
| ## UNEX087 | 0.16 | 0.16 | 0.25 | 0.19 | 0.09 | -0.07 | 0.10 | 0.23 | 0.03 | 0.16 |
| ## UNEX089 | 0.13 | 0.08 | 0.21 | 0.23 | 0.20 | -0.04 | 0.06 | 0.13 | 0.07 | 0.14 |
| ## UNEX090 | 0.07 | 0.08 | 0.21 | 0.16 | 0.22 | 0.03 | 0.09 | 0.17 | 0.00 | 0.14 |
| ## UNEX093 | 0.05 | 0.10 | 0.12 | 0.15 | 0.05 | -0.03 | 0.06 | 0.03 | -0.02 | 0.10 |
| ## UNEX099 | 0.13 | 0.08 | 0.18 | 0.12 | 0.12 | 0.03 | 0.08 | 0.20 | 0.03 | 0.14 |
| ## UNEX103 | 0.03 | -0.03 | 0.05 | 0.05 | 0.02 | 0.09 | 0.03 | 0.10 | -0.05 | -0.01 |
| ## INT004 | 0.11 | 0.14 | 0.14 | 0.16 | 0.13 | 0.04 | -0.02 | 0.16 | -0.01 | 0.16 |
| ## INT009 | 0.02 | 0.01 | 0.02 | 0.01 | 0.05 | -0.09 | 0.01 | 0.05 | 0.05 | 0.00 |
| ## INT011 | 0.07 | 0.07 | 0.09 | 0.11 | -0.02 | 0.00 | -0.03 | 0.03 | 0.05 | 0.07 |

|  |  |  |  |  |  |  |  |  |  |  |  |
| --- | --- | --- | --- | --- | --- | --- | --- | --- | --- | --- | --- |
| ## | INT014 | 0.06 | 0.17 | 0.18 | 0.17 | 0.16 | 0.00 | 0.04 | 0.25 | 0.02 | 0.19 |
| ## | INT026 | 0.07 | 0.11 | 0.10 | 0.13 | 0.08 | 0.05 | 0.03 | 0.25 | -0.01 | 0.20 |
| ## | INT034 | -0.03 | 0.01 | 0.03 | 0.00 | 0.05 | 0.04 | 0.02 | 0.15 | -0.02 | 0.09 |
| ## | INT035 | 0.02 | 0.10 | 0.09 | 0.01 | 0.05 | -0.12 | -0.02 | 0.08 | -0.01 | 0.12 |
| ## | INT039 | 0.00 | -0.01 | -0.02 | 0.02 | 0.02 | 0.01 | 0.01 | 0.08 | 0.00 | 0.03 |
| ## | INT041 | 0.02 | 0.11 | 0.06 | 0.08 | -0.01 | 0.01 | -0.04 | 0.18 | -0.01 | 0.08 |
| ## | INT042 | -0.03 | -0.03 | -0.08 | -0.03 | -0.05 | 0.03 | 0.02 | -0.03 | 0.00 | -0.07 |
| ## | INT045 | 0.03 | 0.05 | 0.10 | 0.10 | 0.05 | 0.00 | 0.05 | 0.13 | 0.01 | 0.09 |
| ## | INT046 | 0.05 | 0.03 | 0.01 | 0.11 | 0.04 | 0.04 | 0.06 | 0.05 | -0.02 | 0.07 |
| ## | INT048 | 0.16 | 0.14 | 0.20 | 0.19 | 0.14 | -0.10 | 0.02 | 0.25 | -0.01 | 0.22 |
| ## | INT049 | 0.01 | 0.12 | 0.08 | 0.09 | 0.12 | -0.07 | 0.06 | 0.19 | 0.03 | 0.18 |
| ## | INT050 | 0.07 | 0.07 | 0.15 | 0.06 | 0.01 | -0.08 | -0.03 | 0.12 | -0.06 | 0.10 |
| ## | INT059 | 0.08 | 0.14 | 0.13 | 0.10 | 0.12 | -0.02 | 0.04 | 0.18 | 0.01 | 0.14 |
| ## | INT064 | 0.11 | 0.09 | 0.17 | 0.14 | 0.12 | 0.00 | 0.02 | 0.19 | -0.03 | 0.22 |
| ## | INT067 | 0.04 | 0.02 | 0.02 | 0.05 | 0.06 | -0.01 | 0.02 | 0.03 | 0.01 | 0.04 |
| ## | INT077 | 0.06 | 0.09 | 0.05 | 0.07 | 0.09 | -0.09 | 0.01 | 0.12 | 0.01 | 0.08 |
| ## | INT080 | 0.12 | 0.12 | 0.24 | 0.17 | 0.14 | 0.04 | 0.02 | 0.24 | -0.02 | 0.19 |
| ## | INT086 | 0.08 | 0.13 | 0.11 | 0.05 | 0.03 | -0.08 | -0.03 | 0.11 | -0.06 | 0.10 |
| ## | INT092 | -0.03 | 0.03 | -0.03 | -0.01 | -0.01 | 0.01 | -0.09 | -0.01 | 0.01 | 0.01 |
| ## | INT094 | 0.06 | 0.05 | 0.06 | 0.11 | 0.03 | -0.05 | -0.02 | 0.07 | 0.06 | 0.04 |
| ## | INT097 | 0.07 | 0.05 | 0.08 | 0.07 | -0.03 | -0.12 | -0.05 | 0.10 | -0.05 | 0.06 |
| ## | INT098 | 0.08 | 0.05 | 0.11 | 0.09 | 0.11 | 0.03 | 0.01 | 0.19 | -0.05 | 0.21 |
| ## | INT102 | 0.09 | 0.12 | 0.19 | 0.13 | 0.08 | -0.01 | -0.03 | 0.26 | -0.04 | 0.18 |
| ## | INT104 | 0.12 | 0.05 | 0.06 | 0.08 | 0.06 | -0.08 | -0.05 | 0.10 | 0.01 | 0.10 |
| ## |  | IMP018 | IMP019 | IMP024 | IMP031 | IMP051 | IMP052 | IMP053 | IMP056 | IMP057 | IMP068 |
| ## | COG001 | -0.10 | 0.10 | 0.01 | 0.05 | -0.05 | -0.03 | 0.13 | 0.11 | -0.27 | -0.02 |
| ## | COG008 | -0.10 | 0.15 | 0.06 | 0.08 | -0.04 | 0.05 | 0.19 | 0.16 | -0.06 | 0.01 |
| ## | COG016 | -0.10 | 0.09 | 0.06 | 0.10 | -0.04 | -0.03 | 0.21 | 0.20 | -0.11 | -0.06 |
| ## | COG021 | -0.08 | 0.13 | -0.02 | 0.04 | -0.01 | 0.06 | 0.13 | 0.12 | -0.10 | 0.04 |
| ## | COG022 | -0.06 | 0.20 | 0.13 | 0.07 | -0.02 | 0.15 | 0.21 | 0.19 | -0.03 | 0.08 |
| ## | COG023 | -0.04 | 0.07 | 0.10 | 0.09 | -0.11 | -0.02 | 0.20 | 0.18 | -0.14 | -0.01 |
| ## | COG028 | -0.11 | 0.13 | 0.12 | 0.07 | -0.07 | 0.04 | 0.23 | 0.24 | -0.15 | -0.03 |
| ## | COG032 | -0.03 | 0.12 | 0.14 | 0.11 | -0.04 | 0.06 | 0.27 | 0.27 | -0.09 | 0.03 |
| ## | COG036 | -0.08 | 0.09 | 0.05 | 0.11 | -0.06 | 0.05 | 0.17 | 0.22 | -0.13 | -0.04 |
| ## | COG037 | -0.08 | 0.19 | 0.14 | 0.10 | 0.00 | 0.19 | 0.20 | 0.22 | -0.09 | 0.03 |
| ## | COG038 | -0.15 | 0.08 | 0.07 | 0.01 | -0.08 | 0.00 | 0.15 | 0.17 | -0.18 | -0.02 |
| ## | COG043 | -0.07 | 0.17 | 0.14 | 0.08 | -0.02 | 0.10 | 0.14 | 0.17 | -0.06 | 0.07 |
| ## | COG054 | -0.06 | 0.21 | 0.11 | 0.04 | -0.01 | 0.10 | 0.23 | 0.24 | -0.04 | 0.06 |
| ## | COG055 | -0.04 | 0.07 | 0.04 | 0.12 | -0.13 | -0.02 | 0.18 | 0.12 | -0.07 | -0.03 |
| ## | COG061 | -0.05 | 0.19 | 0.07 | 0.07 | -0.03 | 0.11 | 0.15 | 0.15 | -0.02 | 0.07 |
| ## | COG062 | -0.10 | 0.10 | 0.05 | 0.05 | -0.04 | 0.12 | 0.18 | 0.19 | -0.10 | 0.00 |
| ## | COG066 | -0.07 | 0.08 | 0.05 | 0.13 | -0.11 | -0.12 | 0.12 | 0.16 | -0.17 | -0.05 |
| ## | COG073 | -0.11 | 0.12 | 0.14 | 0.07 | -0.06 | 0.04 | 0.17 | 0.23 | -0.09 | 0.04 |
| ## | COG075 | -0.09 | 0.22 | 0.12 | 0.09 | 0.01 | 0.06 | 0.25 | 0.31 | -0.09 | 0.07 |
| ## | COG081 | -0.14 | 0.15 | 0.10 | 0.11 | -0.03 | 0.09 | 0.25 | 0.20 | -0.02 | 0.02 |
| ## | COG085 | -0.05 | 0.11 | 0.11 | 0.08 | -0.06 | 0.03 | 0.12 | 0.12 | -0.09 | -0.05 |
| ## | COG091 | -0.07 | 0.09 | 0.04 | 0.03 | -0.06 | 0.00 | 0.07 | 0.19 | -0.20 | -0.05 |
| ## | COG095 | -0.03 | 0.09 | 0.11 | 0.01 | -0.01 | 0.05 | 0.12 | 0.16 | -0.14 | -0.01 |
| ## | COG101 | -0.03 | 0.13 | 0.14 | 0.06 | -0.01 | 0.07 | 0.18 | 0.12 | -0.04 | 0.01 |
| ## | IMP002 | -0.03 | 0.23 | 0.12 | 0.11 | -0.01 | 0.08 | 0.21 | 0.10 | 0.01 | 0.05 |
| ## | IMP007 | 0.05 | -0.02 | 0.12 | 0.05 | 0.09 | 0.10 | -0.03 | -0.09 | 0.24 | 0.19 |
| ## | IMP010 | 0.00 | 0.04 | 0.08 | 0.00 | -0.02 | 0.12 | 0.08 | -0.03 | 0.00 | 0.02 |
| ## | IMP012 | -0.08 | 0.15 | 0.09 | 0.05 | 0.02 | 0.11 | 0.17 | 0.18 | 0.00 | 0.07 |
| ## | IMP013 | -0.02 | 0.12 | 0.04 | 0.26 | -0.01 | 0.05 | 0.11 | 0.00 | 0.07 | 0.06 |

|  |  |  |  |  |  |  |  |  |  |  |
| --- | --- | --- | --- | --- | --- | --- | --- | --- | --- | --- |
| ## IMP015 | -0.06 | 0.29 | 0.10 | 0.16 | 0.01 | 0.10 | 0.32 | 0.32 | -0.01 | 0.12 |
| ## IMP018 | 1.00 | -0.02 | 0.06 | -0.01 | 0.02 | -0.03 | 0.02 | -0.04 | 0.11 | 0.01 |
| ## IMP019 | -0.02 | 1.00 | 0.07 | 0.10 | 0.05 | 0.20 | 0.19 | 0.17 | 0.05 | 0.13 |
| ## IMP024 | 0.06 | 0.07 | 1.00 | 0.08 | 0.02 | 0.07 | 0.12 | 0.08 | 0.00 | 0.01 |
| ## IMP031 | -0.01 | 0.10 | 0.08 | 1.00 | -0.03 | 0.07 | 0.19 | 0.07 | -0.01 | 0.02 |
| ## IMP051 | 0.02 | 0.05 | 0.02 | -0.03 | 1.00 | 0.03 | -0.06 | -0.07 | 0.07 | 0.05 |
| ## IMP052 | -0.03 | 0.20 | 0.07 | 0.07 | 0.03 | 1.00 | 0.08 | 0.01 | 0.07 | 0.20 |
| ## IMP053 | 0.02 | 0.19 | 0.12 | 0.19 | -0.06 | 0.08 | 1.00 | 0.26 | -0.05 | 0.05 |
| ## IMP056 | -0.04 | 0.17 | 0.08 | 0.07 | -0.07 | 0.01 | 0.26 | 1.00 | -0.06 | 0.03 |
| ## IMP057 | 0.11 | 0.05 | 0.00 | -0.01 | 0.07 | 0.07 | -0.05 | -0.06 | 1.00 | 0.09 |
| ## IMP068 | 0.01 | 0.13 | 0.01 | 0.02 | 0.05 | 0.20 | 0.05 | 0.03 | 0.09 | 1.00 |
| ## IMP069 | -0.02 | 0.13 | 0.03 | 0.32 | 0.00 | 0.05 | 0.14 | 0.12 | -0.02 | 0.08 |
| ## IMP070 | -0.04 | 0.10 | 0.07 | 0.14 | 0.07 | 0.03 | 0.11 | 0.14 | 0.00 | 0.09 |
| ## IMP074 | -0.04 | 0.11 | 0.14 | 0.07 | 0.03 | 0.04 | 0.21 | 0.11 | -0.06 | 0.08 |
| ## IMP084 | -0.01 | 0.11 | 0.16 | 0.12 | -0.01 | 0.09 | 0.13 | 0.05 | 0.06 | 0.07 |
| ## IMP088 | -0.05 | 0.16 | 0.09 | 0.24 | -0.05 | 0.10 | 0.18 | 0.10 | 0.05 | -0.03 |
| ## IMP096 | -0.07 | 0.15 | 0.17 | 0.12 | -0.01 | 0.09 | 0.26 | 0.18 | -0.03 | 0.07 |
| ## IMP100 | -0.05 | 0.16 | 0.14 | 0.08 | 0.00 | 0.16 | 0.24 | 0.16 | -0.02 | 0.10 |
| ## UNEX003 | -0.03 | 0.02 | 0.04 | 0.01 | 0.03 | 0.11 | 0.05 | 0.05 | 0.03 | 0.03 |
| ## UNEX005 | -0.02 | 0.09 | 0.09 | 0.06 | 0.01 | 0.09 | 0.22 | 0.15 | -0.04 | 0.09 |
| ## UNEX006 | -0.01 | 0.14 | 0.01 | -0.01 | 0.03 | 0.08 | 0.07 | 0.05 | 0.04 | 0.10 |
| ## UNEX017 | -0.05 | 0.12 | 0.07 | 0.07 | -0.02 | 0.05 | 0.15 | 0.15 | 0.00 | 0.08 |
| ## UNEX020 | -0.07 | 0.07 | 0.04 | 0.02 | -0.01 | 0.10 | 0.07 | 0.10 | 0.02 | 0.12 |
| ## UNEX025 | -0.09 | 0.18 | 0.04 | 0.03 | 0.03 | 0.17 | 0.08 | 0.15 | 0.02 | 0.10 |
| ## UNEX027 | -0.08 | 0.18 | 0.09 | 0.08 | -0.02 | 0.09 | 0.18 | 0.17 | 0.00 | 0.09 |
| ## UNEX029 | -0.06 | 0.09 | 0.00 | 0.00 | 0.02 | 0.07 | 0.05 | 0.03 | 0.03 | 0.06 |
| ## UNEX030 | -0.12 | 0.14 | 0.09 | 0.06 | -0.02 | 0.11 | 0.17 | 0.17 | -0.01 | 0.03 |
| ## UNEX033 | -0.06 | 0.15 | 0.10 | 0.11 | -0.05 | 0.08 | 0.15 | 0.19 | 0.00 | 0.05 |
| ## UNEX040 | -0.07 | 0.11 | 0.07 | 0.00 | 0.03 | 0.10 | 0.09 | 0.15 | -0.01 | 0.05 |
| ## UNEX044 | -0.08 | 0.12 | 0.07 | 0.04 | 0.04 | 0.06 | 0.13 | 0.09 | 0.04 | 0.01 |
| ## UNEX047 | -0.09 | 0.09 | 0.03 | 0.09 | -0.01 | 0.11 | 0.15 | 0.19 | -0.08 | 0.05 |
| ## UNEX058 | -0.03 | 0.09 | 0.03 | 0.05 | -0.02 | 0.09 | 0.09 | 0.09 | -0.05 | 0.09 |
| ## UNEX060 | -0.10 | 0.13 | 0.09 | 0.10 | 0.04 | 0.12 | 0.18 | 0.08 | 0.01 | 0.09 |
| ## UNEX063 | -0.10 | 0.09 | -0.01 | 0.05 | -0.05 | 0.03 | 0.12 | 0.17 | -0.07 | 0.01 |
| ## UNEX065 | -0.01 | 0.10 | 0.04 | 0.09 | 0.02 | 0.09 | 0.15 | 0.16 | 0.00 | 0.08 |
| ## UNEX071 | -0.04 | 0.05 | 0.03 | 0.05 | -0.03 | 0.08 | 0.13 | 0.11 | 0.02 | 0.04 |
| ## UNEX072 | -0.01 | 0.11 | 0.02 | 0.07 | 0.04 | 0.08 | 0.15 | 0.21 | -0.05 | 0.01 |
| ## UNEX076 | -0.02 | 0.04 | -0.01 | 0.01 | -0.03 | 0.04 | 0.11 | 0.10 | -0.03 | 0.03 |
| ## UNEX078 | -0.08 | 0.14 | 0.10 | 0.07 | -0.02 | 0.12 | 0.14 | 0.13 | -0.01 | 0.01 |
| ## UNEX079 | -0.04 | 0.07 | 0.07 | 0.10 | -0.07 | 0.10 | 0.11 | 0.09 | 0.00 | 0.07 |
| ## UNEX082 | -0.06 | 0.13 | 0.05 | 0.03 | -0.01 | 0.05 | 0.11 | 0.21 | -0.09 | 0.00 |
| ## UNEX083 | -0.04 | 0.14 | 0.09 | 0.06 | -0.07 | 0.11 | 0.26 | 0.30 | -0.09 | 0.05 |
| ## UNEX087 | -0.09 | 0.15 | 0.12 | 0.05 | 0.01 | 0.10 | 0.15 | 0.16 | -0.06 | 0.05 |
| ## UNEX089 | -0.04 | 0.10 | 0.11 | 0.06 | -0.04 | 0.04 | 0.18 | 0.21 | -0.03 | 0.04 |
| ## UNEX090 | -0.06 | 0.09 | 0.10 | 0.08 | -0.02 | 0.09 | 0.19 | 0.21 | -0.03 | 0.09 |
| ## UNEX093 | -0.02 | 0.07 | -0.02 | 0.02 | 0.01 | 0.10 | 0.10 | 0.04 | -0.03 | 0.07 |
| ## UNEX099 | -0.09 | 0.07 | 0.09 | 0.07 | -0.02 | 0.09 | 0.16 | 0.16 | -0.03 | 0.02 |
| ## UNEX103 | 0.01 | 0.01 | 0.06 | 0.05 | 0.02 | 0.11 | 0.04 | 0.03 | 0.01 | 0.07 |
| ## INT004 | -0.04 | 0.06 | 0.17 | 0.06 | 0.06 | 0.03 | 0.12 | 0.12 | -0.07 | 0.02 |
| ## INT009 | -0.11 | 0.05 | 0.02 | 0.03 | 0.01 | -0.02 | 0.08 | -0.02 | -0.02 | 0.02 |
| ## INT011 | -0.05 | 0.01 | 0.12 | 0.04 | -0.02 | -0.05 | 0.11 | 0.08 | -0.07 | -0.03 |
| ## INT014 | -0.15 | 0.14 | 0.14 | 0.06 | 0.02 | 0.05 | 0.15 | 0.16 | -0.08 | 0.04 |
| ## INT026 | -0.12 | 0.11 | 0.09 | 0.04 | 0.01 | 0.10 | 0.13 | 0.11 | -0.03 | 0.14 |
| ## INT034 | -0.11 | 0.03 | 0.05 | -0.06 | 0.09 | -0.01 | -0.01 | 0.02 | 0.00 | 0.07 |

|  |  |  |  |  |  |  |  |  |  |  |  |
| --- | --- | --- | --- | --- | --- | --- | --- | --- | --- | --- | --- |
| ## | INT035 | -0.20 | 0.06 | 0.00 | 0.03 | 0.04 | -0.04 | 0.07 | 0.07 | -0.13 | 0.04 |
| ## | INT039 | -0.02 | 0.02 | 0.07 | 0.03 | 0.04 | -0.02 | -0.01 | -0.01 | 0.02 | -0.02 |
| ## | INT041 | -0.06 | 0.03 | 0.11 | 0.03 | 0.04 | -0.01 | 0.05 | 0.12 | -0.05 | 0.06 |
| ## | INT042 | 0.02 | -0.05 | 0.01 | -0.07 | 0.07 | -0.03 | -0.11 | -0.11 | 0.00 | 0.05 |
| ## | INT045 | -0.08 | 0.06 | 0.07 | 0.00 | 0.01 | 0.07 | 0.10 | 0.02 | -0.03 | 0.09 |
| ## | INT046 | -0.03 | 0.02 | 0.06 | 0.03 | 0.04 | 0.06 | 0.04 | 0.01 | -0.03 | 0.07 |
| ## | INT048 | -0.08 | 0.14 | 0.05 | 0.01 | -0.03 | 0.06 | 0.21 | 0.19 | -0.12 | 0.01 |
| ## | INT049 | -0.16 | 0.10 | 0.12 | 0.06 | 0.04 | 0.04 | 0.13 | 0.13 | -0.09 | 0.02 |
| ## | INT050 | -0.17 | -0.03 | 0.05 | 0.04 | 0.01 | -0.02 | 0.12 | 0.12 | -0.11 | 0.00 |
| ## | INT059 | -0.08 | 0.16 | 0.09 | 0.05 | -0.02 | 0.03 | 0.14 | 0.15 | -0.05 | -0.04 |
| ## | INT064 | -0.15 | 0.10 | 0.07 | 0.03 | 0.08 | 0.08 | 0.08 | 0.05 | -0.09 | 0.05 |
| ## | INT067 | -0.04 | 0.03 | 0.02 | -0.01 | 0.00 | -0.01 | 0.05 | 0.02 | -0.06 | -0.05 |
| ## | INT077 | -0.05 | 0.06 | 0.00 | 0.02 | 0.01 | 0.03 | 0.12 | -0.01 | -0.09 | 0.02 |
| ## | INT080 | -0.11 | 0.11 | 0.11 | 0.02 | 0.08 | 0.07 | 0.11 | 0.15 | -0.06 | 0.08 |
| ## | INT086 | -0.13 | -0.03 | 0.06 | 0.00 | -0.01 | -0.03 | 0.08 | 0.13 | -0.17 | 0.01 |
| ## | INT092 | -0.08 | 0.00 | 0.04 | 0.02 | 0.03 | 0.01 | -0.02 | -0.01 | -0.04 | 0.03 |
| ## | INT094 | -0.06 | 0.05 | 0.07 | 0.04 | -0.04 | -0.02 | 0.11 | 0.10 | -0.05 | -0.05 |
| ## | INT097 | -0.13 | -0.04 | 0.01 | -0.01 | -0.03 | -0.01 | 0.05 | 0.08 | -0.13 | -0.04 |
| ## | INT098 | -0.17 | 0.08 | 0.12 | 0.02 | 0.13 | 0.05 | 0.10 | 0.12 | -0.08 | 0.08 |
| ## | INT102 | -0.09 | 0.11 | 0.12 | 0.02 | 0.07 | 0.04 | 0.13 | 0.12 | -0.09 | 0.07 |
| ## | INT104 | -0.13 | 0.03 | 0.09 | 0.05 | -0.02 | 0.00 | 0.11 | 0.09 | -0.12 | 0.01 |
| ## |  | IMP069 | IMP070 | IMP074 | IMP084 | IMP088 | IMP096 | IMP100 | UNEX003 | UNEX005 |  |
| ## | COG001 | 0.00 | 0.04 | 0.08 | 0.01 | 0.00 | 0.14 | 0.11 | 0.04 | 0.09 |  |
| ## | COG008 | 0.04 | 0.11 | 0.12 | 0.04 | 0.10 | 0.24 | 0.18 | 0.01 | 0.09 |  |
| ## | COG016 | 0.06 | 0.08 | 0.11 | 0.03 | 0.10 | 0.20 | 0.18 | 0.07 | 0.18 |  |
| ## | COG021 | 0.02 | 0.07 | 0.07 | 0.03 | 0.10 | 0.12 | 0.11 | 0.06 | 0.11 |  |
| ## | COG022 | 0.05 | 0.11 | 0.16 | 0.09 | 0.12 | 0.27 | 0.28 | 0.09 | 0.23 |  |
| ## | COG023 | 0.10 | 0.08 | 0.11 | 0.08 | 0.05 | 0.16 | 0.15 | 0.06 | 0.10 |  |
| ## | COG028 | 0.11 | 0.12 | 0.12 | 0.03 | 0.09 | 0.23 | 0.19 | 0.03 | 0.19 |  |
| ## | COG032 | 0.04 | 0.07 | 0.12 | 0.07 | 0.08 | 0.19 | 0.16 | 0.06 | 0.18 |  |
| ## | COG036 | 0.07 | 0.12 | 0.13 | -0.01 | 0.08 | 0.14 | 0.11 | 0.02 | 0.15 |  |
| ## | COG037 | 0.08 | 0.16 | 0.15 | 0.15 | 0.13 | 0.24 | 0.20 | 0.05 | 0.10 |  |
| ## | COG038 | 0.00 | 0.05 | 0.12 | 0.06 | 0.00 | 0.13 | 0.13 | 0.04 | 0.13 |  |
| ## | COG043 | -0.01 | 0.13 | 0.21 | 0.06 | 0.10 | 0.20 | 0.24 | 0.09 | 0.16 |  |
| ## | COG054 | 0.12 | 0.21 | 0.21 | 0.07 | 0.08 | 0.27 | 0.12 | 0.04 | 0.17 |  |
| ## | COG055 | 0.06 | 0.08 | 0.10 | -0.02 | 0.09 | 0.15 | 0.13 | 0.10 | 0.17 |  |
| ## | COG061 | 0.06 | 0.13 | 0.13 | 0.08 | 0.09 | 0.21 | 0.16 | 0.08 | 0.17 |  |
| ## | COG062 | 0.01 | 0.07 | 0.14 | 0.00 | 0.06 | 0.19 | 0.18 | 0.04 | 0.16 |  |
| ## | COG066 | 0.06 | 0.07 | 0.06 | 0.02 | 0.03 | 0.10 | 0.10 | 0.02 | 0.09 |  |
| ## | COG073 | 0.10 | 0.14 | 0.16 | 0.07 | 0.01 | 0.19 | 0.21 | 0.04 | 0.14 |  |
| ## | COG075 | 0.07 | 0.21 | 0.23 | 0.09 | 0.02 | 0.28 | 0.26 | 0.08 | 0.17 |  |
| ## | COG081 | 0.09 | 0.20 | 0.19 | 0.11 | 0.07 | 0.31 | 0.27 | 0.12 | 0.22 |  |
| ## | COG085 | 0.06 | 0.07 | 0.19 | 0.04 | 0.07 | 0.15 | 0.18 | 0.07 | 0.07 |  |
| ## | COG091 | 0.03 | -0.01 | 0.06 | 0.01 | 0.04 | 0.14 | 0.10 | 0.00 | 0.09 |  |
| ## | COG095 | 0.04 | 0.09 | 0.17 | 0.02 | 0.10 | 0.21 | 0.19 | 0.06 | 0.18 |  |
| ## | COG101 | 0.08 | 0.03 | 0.14 | 0.10 | 0.10 | 0.20 | 0.20 | 0.03 | 0.10 |  |
| ## | IMP002 | 0.06 | 0.09 | 0.12 | 0.07 | 0.08 | 0.21 | 0.22 | 0.04 | 0.09 |  |
| ## | IMP007 | 0.08 | 0.06 | 0.02 | 0.13 | 0.00 | 0.00 | 0.03 | -0.02 | -0.04 |  |
| ## | IMP010 | 0.00 | 0.00 | 0.06 | 0.11 | 0.07 | 0.07 | 0.05 | 0.02 | 0.06 |  |
| ## | IMP012 | 0.06 | 0.15 | 0.18 | 0.14 | 0.08 | 0.30 | 0.29 | 0.12 | 0.24 |  |
| ## | IMP013 | 0.21 | 0.06 | 0.05 | 0.06 | 0.16 | 0.03 | -0.04 | 0.01 | 0.00 |  |
| ## | IMP015 | 0.18 | 0.16 | 0.16 | 0.13 | 0.14 | 0.35 | 0.15 | 0.03 | 0.21 |  |
| ## | IMP018 | -0.02 | -0.04 | -0.04 | -0.01 | -0.05 | -0.07 | -0.05 | -0.03 | -0.02 |  |
| ## | IMP019 | 0.13 | 0.10 | 0.11 | 0.11 | 0.16 | 0.15 | 0.16 | 0.02 | 0.09 |  |

|  |  |  |  |  |  |  |  |  |  |
| --- | --- | --- | --- | --- | --- | --- | --- | --- | --- |
| ## IMP024 | 0.03 | 0.07 | 0.14 | 0.16 | 0.09 | 0.17 | 0.14 | 0.04 | 0.09 |
| ## IMP031 | 0.32 | 0.14 | 0.07 | 0.12 | 0.24 | 0.12 | 0.08 | 0.01 | 0.06 |
| ## IMP051 | 0.00 | 0.07 | 0.03 | -0.01 | -0.05 | -0.01 | 0.00 | 0.03 | 0.01 |
| ## IMP052 | 0.05 | 0.03 | 0.04 | 0.09 | 0.10 | 0.09 | 0.16 | 0.11 | 0.09 |
| ## IMP053 | 0.14 | 0.11 | 0.21 | 0.13 | 0.18 | 0.26 | 0.24 | 0.05 | 0.22 |
| ## IMP056 | 0.12 | 0.14 | 0.11 | 0.05 | 0.10 | 0.18 | 0.16 | 0.05 | 0.15 |
| ## IMP057 | -0.02 | 0.00 | -0.06 | 0.06 | 0.05 | -0.03 | -0.02 | 0.03 | -0.04 |
| ## IMP068 | 0.08 | 0.09 | 0.08 | 0.07 | -0.03 | 0.07 | 0.10 | 0.03 | 0.09 |
| ## IMP069 | 1.00 | 0.16 | 0.04 | 0.12 | 0.18 | 0.12 | 0.09 | 0.03 | 0.03 |
| ## IMP070 | 0.16 | 1.00 | 0.29 | 0.07 | 0.05 | 0.16 | 0.09 | 0.06 | 0.09 |
| ## IMP074 | 0.04 | 0.29 | 1.00 | 0.11 | 0.07 | 0.16 | 0.18 | 0.02 | 0.11 |
| ## IMP084 | 0.12 | 0.07 | 0.11 | 1.00 | 0.10 | 0.18 | 0.13 | 0.07 | 0.07 |
| ## IMP088 | 0.18 | 0.05 | 0.07 | 0.10 | 1.00 | 0.19 | 0.07 | 0.07 | 0.08 |
| ## IMP096 | 0.12 | 0.16 | 0.16 | 0.18 | 0.19 | 1.00 | 0.23 | 0.11 | 0.20 |
| ## IMP100 | 0.09 | 0.09 | 0.18 | 0.13 | 0.07 | 0.23 | 1.00 | 0.08 | 0.15 |
| ## UNEX003 | 0.03 | 0.06 | 0.02 | 0.07 | 0.07 | 0.11 | 0.08 | 1.00 | 0.27 |
| ## UNEX005 | 0.03 | 0.09 | 0.11 | 0.07 | 0.08 | 0.20 | 0.15 | 0.27 | 1.00 |
| ## UNEX006 | 0.01 | 0.05 | 0.07 | 0.03 | 0.08 | 0.15 | 0.10 | 0.21 | 0.29 |
| ## UNEX017 | 0.04 | 0.11 | 0.16 | 0.10 | 0.11 | 0.26 | 0.18 | 0.11 | 0.34 |
| ## UNEX020 | 0.09 | 0.10 | 0.07 | 0.01 | 0.06 | 0.16 | 0.12 | 0.15 | 0.20 |
| ## UNEX025 | 0.07 | 0.09 | 0.10 | 0.04 | 0.05 | 0.10 | 0.11 | 0.14 | 0.21 |
| ## UNEX027 | 0.09 | 0.11 | 0.14 | 0.07 | 0.15 | 0.28 | 0.21 | 0.13 | 0.28 |
| ## UNEX029 | 0.01 | 0.08 | 0.08 | 0.06 | 0.02 | 0.08 | 0.11 | 0.07 | 0.18 |
| ## UNEX030 | 0.06 | 0.16 | 0.17 | 0.10 | 0.12 | 0.21 | 0.19 | 0.14 | 0.33 |
| ## UNEX033 | 0.10 | 0.15 | 0.13 | 0.04 | 0.15 | 0.17 | 0.09 | 0.03 | 0.15 |
| ## UNEX040 | 0.09 | 0.10 | 0.10 | 0.03 | 0.07 | 0.13 | 0.16 | 0.09 | 0.18 |
| ## UNEX044 | 0.06 | 0.16 | 0.25 | 0.09 | 0.06 | 0.21 | 0.16 | 0.21 | 0.20 |
| ## UNEX047 | 0.02 | 0.12 | 0.13 | 0.08 | 0.06 | 0.20 | 0.14 | 0.09 | 0.18 |
| ## UNEX058 | 0.09 | 0.11 | 0.04 | 0.08 | 0.06 | 0.24 | 0.13 | 0.13 | 0.23 |
| ## UNEX060 | 0.14 | 0.23 | 0.15 | 0.10 | 0.08 | 0.25 | 0.19 | 0.08 | 0.17 |
| ## UNEX063 | 0.12 | 0.04 | 0.12 | 0.06 | 0.11 | 0.14 | 0.17 | 0.12 | 0.26 |
| ## UNEX065 | 0.04 | 0.10 | 0.17 | 0.05 | 0.14 | 0.16 | 0.15 | 0.10 | 0.21 |
| ## UNEX071 | 0.09 | 0.06 | 0.10 | 0.10 | 0.11 | 0.12 | 0.09 | 0.09 | 0.12 |
| ## UNEX072 | 0.04 | 0.14 | 0.18 | 0.01 | 0.07 | 0.21 | 0.09 | 0.07 | 0.18 |
| ## UNEX076 | 0.07 | 0.07 | 0.05 | 0.04 | 0.07 | 0.13 | 0.07 | 0.04 | 0.11 |
| ## UNEX078 | 0.06 | 0.11 | 0.18 | 0.13 | 0.15 | 0.25 | 0.17 | 0.10 | 0.22 |
| ## UNEX079 | 0.12 | 0.04 | 0.15 | 0.10 | 0.10 | 0.17 | 0.16 | 0.11 | 0.18 |
| ## UNEX082 | 0.02 | 0.07 | 0.08 | 0.01 | 0.10 | 0.19 | 0.07 | 0.03 | 0.12 |
| ## UNEX083 | 0.10 | 0.09 | 0.18 | 0.08 | 0.17 | 0.27 | 0.19 | 0.07 | 0.29 |
| ## UNEX087 | 0.08 | 0.14 | 0.16 | 0.09 | 0.12 | 0.26 | 0.15 | 0.10 | 0.21 |
| ## UNEX089 | 0.08 | 0.09 | 0.12 | 0.09 | 0.11 | 0.24 | 0.14 | 0.12 | 0.24 |
| ## UNEX090 | 0.10 | 0.12 | 0.13 | 0.11 | 0.12 | 0.16 | 0.25 | 0.11 | 0.22 |
| ## UNEX093 | 0.06 | 0.08 | 0.06 | 0.05 | 0.07 | 0.12 | 0.12 | 0.10 | 0.15 |
| ## UNEX099 | 0.06 | 0.11 | 0.13 | 0.08 | 0.14 | 0.23 | 0.16 | 0.23 | 0.27 |
| ## UNEX103 | 0.06 | 0.05 | 0.05 | 0.08 | 0.03 | 0.12 | 0.12 | 0.15 | 0.14 |
| ## INT004 | 0.06 | 0.11 | 0.10 | 0.03 | 0.02 | 0.15 | 0.15 | 0.05 | 0.07 |
| ## INT009 | 0.05 | 0.09 | 0.02 | 0.03 | 0.03 | 0.02 | 0.09 | 0.01 | 0.01 |
| ## INT011 | 0.02 | 0.03 | 0.10 | 0.04 | 0.06 | 0.10 | 0.05 | -0.01 | 0.03 |
| ## INT014 | 0.05 | 0.23 | 0.17 | 0.05 | 0.07 | 0.21 | 0.19 | 0.06 | 0.12 |
| ## INT026 | 0.05 | 0.14 | 0.14 | 0.13 | 0.02 | 0.25 | 0.12 | 0.04 | 0.14 |
| ## INT034 | 0.08 | 0.10 | 0.01 | 0.04 | -0.02 | 0.10 | -0.01 | -0.01 | 0.05 |
| ## INT035 | 0.02 | 0.08 | 0.09 | -0.03 | -0.04 | 0.11 | 0.08 | -0.03 | 0.00 |
| ## INT039 | 0.03 | 0.05 | 0.03 | 0.02 | 0.00 | 0.09 | 0.00 | 0.03 | -0.03 |
| ## INT041 | 0.05 | 0.16 | 0.14 | 0.01 | -0.02 | 0.16 | 0.08 | 0.07 | 0.04 |

|  |  |  |  |  |  |  |  |  |  |  |
| --- | --- | --- | --- | --- | --- | --- | --- | --- | --- | --- |
| ## | INT042 | -0.07 | -0.01 | -0.02 | -0.03 | -0.11 | -0.03 | -0.05 | 0.00 | -0.10 |
| ## | INT045 | 0.03 | 0.10 | 0.21 | 0.10 | 0.03 | 0.13 | 0.12 | 0.06 | 0.14 |
| ## | INT046 | 0.04 | 0.03 | 0.04 | 0.05 | -0.02 | 0.06 | 0.02 | 0.00 | -0.04 |
| ## | INT048 | 0.03 | 0.05 | 0.12 | 0.07 | 0.09 | 0.24 | 0.15 | 0.08 | 0.15 |
| ## | INT049 | 0.02 | 0.08 | 0.09 | 0.02 | 0.09 | 0.13 | 0.12 | 0.02 | 0.10 |
| ## | INT050 | 0.05 | 0.06 | 0.04 | 0.05 | 0.00 | 0.12 | 0.11 | 0.07 | 0.07 |
| ## | INT059 | 0.05 | 0.13 | 0.14 | 0.04 | 0.06 | 0.15 | 0.11 | 0.03 | 0.10 |
| ## | INT064 | 0.07 | 0.12 | 0.09 | 0.06 | 0.03 | 0.23 | 0.10 | 0.01 | 0.07 |
| ## | INT067 | 0.01 | 0.04 | 0.06 | 0.04 | 0.07 | 0.06 | 0.01 | 0.03 | 0.02 |
| ## | INT077 | 0.02 | 0.03 | 0.03 | 0.09 | 0.00 | 0.10 | 0.09 | 0.06 | 0.02 |
| ## | INT080 | 0.08 | 0.11 | 0.15 | 0.12 | 0.03 | 0.25 | 0.18 | 0.04 | 0.11 |
| ## | INT086 | -0.01 | 0.09 | 0.11 | -0.01 | -0.04 | 0.10 | 0.07 | 0.00 | 0.08 |
| ## | INT092 | -0.01 | 0.06 | 0.04 | -0.07 | 0.01 | -0.02 | -0.03 | -0.08 | -0.05 |
| ## | INT094 | 0.03 | 0.02 | 0.06 | 0.06 | 0.08 | 0.14 | 0.05 | 0.02 | 0.03 |
| ## | INT097 | -0.04 | 0.04 | 0.04 | -0.01 | -0.01 | 0.07 | 0.04 | 0.00 | -0.01 |
| ## | INT098 | 0.06 | 0.13 | 0.11 | 0.05 | -0.02 | 0.16 | 0.11 | 0.02 | 0.02 |
| ## | INT102 | 0.01 | 0.18 | 0.25 | 0.09 | 0.02 | 0.21 | 0.16 | 0.09 | 0.12 |
| ## | INT104 | 0.01 | 0.09 | 0.05 | 0.07 | 0.00 | 0.16 | 0.14 | -0.02 | 0.07 |
| ## |  | UNEX006 | UNEX017 | UNEX020 | UNEX025 | UNEX027 | UNEX029 | UNEX030 | UNEX033 | UNEX040 |
| ## | COG001 | 0.07 | 0.15 | 0.04 | 0.06 | 0.10 | 0.08 | 0.10 | 0.10 | 0.04 |
| ## | COG008 | 0.04 | 0.18 | 0.04 | 0.13 | 0.17 | -0.02 | 0.15 | 0.15 | 0.05 |
| ## | COG016 | 0.14 | 0.17 | 0.03 | 0.08 | 0.22 | 0.10 | 0.22 | 0.11 | 0.10 |
| ## | COG021 | 0.04 | 0.15 | 0.04 | 0.08 | 0.16 | 0.02 | 0.08 | 0.12 | 0.07 |
| ## | COG022 | 0.22 | 0.34 | 0.16 | 0.20 | 0.29 | 0.16 | 0.32 | 0.11 | 0.20 |
| ## | COG023 | 0.07 | 0.18 | 0.06 | 0.09 | 0.13 | 0.08 | 0.13 | 0.10 | 0.06 |
| ## | COG028 | 0.05 | 0.21 | 0.11 | 0.09 | 0.20 | 0.09 | 0.17 | 0.14 | 0.10 |
| ## | COG032 | 0.11 | 0.26 | 0.08 | 0.17 | 0.21 | 0.07 | 0.17 | 0.16 | 0.09 |
| ## | COG036 | 0.09 | 0.18 | 0.04 | 0.11 | 0.22 | 0.03 | 0.19 | 0.15 | 0.08 |
| ## | COG037 | 0.08 | 0.28 | 0.09 | 0.18 | 0.20 | 0.09 | 0.18 | 0.19 | 0.13 |
| ## | COG038 | 0.04 | 0.14 | 0.01 | 0.04 | 0.06 | 0.05 | 0.14 | 0.07 | 0.02 |
| ## | COG043 | 0.18 | 0.27 | 0.12 | 0.20 | 0.20 | 0.11 | 0.29 | 0.24 | 0.12 |
| ## | COG054 | 0.07 | 0.21 | 0.10 | 0.08 | 0.14 | 0.03 | 0.21 | 0.20 | 0.15 |
| ## | COG055 | 0.12 | 0.17 | 0.08 | 0.07 | 0.17 | 0.09 | 0.23 | 0.14 | 0.10 |
| ## | COG061 | 0.21 | 0.26 | 0.11 | 0.14 | 0.25 | 0.11 | 0.24 | 0.16 | 0.09 |
| ## | COG062 | 0.09 | 0.20 | 0.08 | 0.20 | 0.20 | 0.12 | 0.24 | 0.14 | 0.08 |
| ## | COG066 | 0.00 | 0.13 | 0.00 | 0.01 | 0.07 | 0.02 | 0.15 | 0.08 | 0.00 |
| ## | COG073 | 0.05 | 0.20 | 0.11 | 0.15 | 0.13 | 0.09 | 0.18 | 0.20 | 0.12 |
| ## | COG075 | 0.13 | 0.29 | 0.14 | 0.15 | 0.20 | 0.14 | 0.27 | 0.27 | 0.17 |
| ## | COG081 | 0.17 | 0.26 | 0.13 | 0.13 | 0.22 | 0.09 | 0.25 | 0.15 | 0.13 |
| ## | COG085 | 0.12 | 0.17 | 0.07 | 0.07 | 0.18 | 0.00 | 0.13 | 0.08 | 0.16 |
| ## | COG091 | 0.00 | 0.13 | -0.03 | 0.06 | 0.10 | 0.01 | 0.11 | 0.09 | 0.03 |
| ## | COG095 | 0.08 | 0.18 | 0.07 | 0.14 | 0.21 | 0.13 | 0.23 | 0.10 | 0.16 |
| ## | COG101 | 0.11 | 0.19 | 0.08 | 0.12 | 0.17 | 0.09 | 0.13 | 0.13 | 0.09 |
| ## | IMP002 | 0.08 | 0.21 | 0.12 | 0.16 | 0.11 | -0.01 | 0.16 | 0.10 | 0.09 |
| ## | IMP007 | 0.04 | -0.07 | 0.05 | 0.08 | 0.01 | 0.02 | 0.01 | -0.01 | 0.03 |
| ## | IMP010 | 0.05 | 0.08 | -0.06 | 0.05 | 0.07 | 0.04 | 0.08 | 0.06 | -0.01 |
| ## | IMP012 | 0.23 | 0.29 | 0.11 | 0.26 | 0.27 | 0.11 | 0.28 | 0.21 | 0.22 |
| ## | IMP013 | 0.01 | 0.03 | 0.03 | 0.01 | 0.03 | -0.03 | 0.03 | 0.08 | 0.01 |
| ## | IMP015 | 0.11 | 0.25 | 0.08 | 0.11 | 0.21 | 0.04 | 0.18 | 0.23 | 0.11 |
| ## | IMP018 | -0.01 | -0.05 | -0.07 | -0.09 | -0.08 | -0.06 | -0.12 | -0.06 | -0.07 |
| ## | IMP019 | 0.14 | 0.12 | 0.07 | 0.18 | 0.18 | 0.09 | 0.14 | 0.15 | 0.11 |
| ## | IMP024 | 0.01 | 0.07 | 0.04 | 0.04 | 0.09 | 0.00 | 0.09 | 0.10 | 0.07 |
| ## | IMP031 | -0.01 | 0.07 | 0.02 | 0.03 | 0.08 | 0.00 | 0.06 | 0.11 | 0.00 |
| ## | IMP051 | 0.03 | -0.02 | -0.01 | 0.03 | -0.02 | 0.02 | -0.02 | -0.05 | 0.03 |

|  |  |  |  |  |  |  |  |  |  |
| --- | --- | --- | --- | --- | --- | --- | --- | --- | --- |
| ## IMP052 | 0.08 | 0.05 | 0.10 | 0.17 | 0.09 | 0.07 | 0.11 | 0.08 | 0.10 |
| ## IMP053 | 0.07 | 0.15 | 0.07 | 0.08 | 0.18 | 0.05 | 0.17 | 0.15 | 0.09 |
| ## IMP056 | 0.05 | 0.15 | 0.10 | 0.15 | 0.17 | 0.03 | 0.17 | 0.19 | 0.15 |
| ## IMP057 | 0.04 | 0.00 | 0.02 | 0.02 | 0.00 | 0.03 | -0.01 | 0.00 | -0.01 |
| ## IMP068 | 0.10 | 0.08 | 0.12 | 0.10 | 0.09 | 0.06 | 0.03 | 0.05 | 0.05 |
| ## IMP069 | 0.01 | 0.04 | 0.09 | 0.07 | 0.09 | 0.01 | 0.06 | 0.10 | 0.09 |
| ## IMP070 | 0.05 | 0.11 | 0.10 | 0.09 | 0.11 | 0.08 | 0.16 | 0.15 | 0.10 |
| ## IMP074 | 0.07 | 0.16 | 0.07 | 0.10 | 0.14 | 0.08 | 0.17 | 0.13 | 0.10 |
| ## IMP084 | 0.03 | 0.10 | 0.01 | 0.04 | 0.07 | 0.06 | 0.10 | 0.04 | 0.03 |
| ## IMP088 | 0.08 | 0.11 | 0.06 | 0.05 | 0.15 | 0.02 | 0.12 | 0.15 | 0.07 |
| ## IMP096 | 0.15 | 0.26 | 0.16 | 0.10 | 0.28 | 0.08 | 0.21 | 0.17 | 0.13 |
| ## IMP100 | 0.10 | 0.18 | 0.12 | 0.11 | 0.21 | 0.11 | 0.19 | 0.09 | 0.16 |
| ## UNEX003 | 0.21 | 0.11 | 0.15 | 0.14 | 0.13 | 0.07 | 0.14 | 0.03 | 0.09 |
| ## UNEX005 | 0.29 | 0.34 | 0.20 | 0.21 | 0.28 | 0.18 | 0.33 | 0.15 | 0.18 |
| ## UNEX006 | 1.00 | 0.21 | 0.17 | 0.21 | 0.37 | 0.21 | 0.22 | 0.08 | 0.12 |
| ## UNEX017 | 0.21 | 1.00 | 0.12 | 0.25 | 0.25 | 0.15 | 0.31 | 0.18 | 0.18 |
| ## UNEX020 | 0.17 | 0.12 | 1.00 | 0.31 | 0.18 | 0.20 | 0.21 | 0.14 | 0.40 |
| ## UNEX025 | 0.21 | 0.25 | 0.31 | 1.00 | 0.21 | 0.20 | 0.26 | 0.15 | 0.28 |
| ## UNEX027 | 0.37 | 0.25 | 0.18 | 0.21 | 1.00 | 0.19 | 0.27 | 0.14 | 0.18 |
| ## UNEX029 | 0.21 | 0.15 | 0.20 | 0.20 | 0.19 | 1.00 | 0.22 | 0.06 | 0.21 |
| ## UNEX030 | 0.22 | 0.31 | 0.21 | 0.26 | 0.27 | 0.22 | 1.00 | 0.19 | 0.27 |
| ## UNEX033 | 0.08 | 0.18 | 0.14 | 0.15 | 0.14 | 0.06 | 0.19 | 1.00 | 0.10 |
| ## UNEX040 | 0.12 | 0.18 | 0.40 | 0.28 | 0.18 | 0.21 | 0.27 | 0.10 | 1.00 |
| ## UNEX044 | 0.12 | 0.19 | 0.17 | 0.25 | 0.15 | 0.19 | 0.31 | 0.15 | 0.23 |
| ## UNEX047 | 0.14 | 0.29 | 0.08 | 0.15 | 0.18 | 0.08 | 0.20 | 0.17 | 0.06 |
| ## UNEX058 | 0.18 | 0.20 | 0.23 | 0.37 | 0.25 | 0.16 | 0.22 | 0.14 | 0.17 |
| ## UNEX060 | 0.15 | 0.19 | 0.27 | 0.24 | 0.21 | 0.12 | 0.28 | 0.23 | 0.32 |
| ## UNEX063 | 0.17 | 0.24 | 0.19 | 0.21 | 0.19 | 0.16 | 0.25 | 0.14 | 0.15 |
| ## UNEX065 | 0.17 | 0.20 | 0.24 | 0.26 | 0.23 | 0.18 | 0.21 | 0.12 | 0.21 |
| ## UNEX071 | 0.08 | 0.09 | 0.10 | 0.15 | 0.09 | 0.04 | 0.19 | 0.09 | 0.08 |
| ## UNEX072 | 0.12 | 0.19 | 0.26 | 0.28 | 0.19 | 0.18 | 0.18 | 0.24 | 0.24 |
| ## UNEX076 | 0.12 | 0.08 | 0.16 | 0.08 | 0.09 | 0.06 | 0.11 | 0.06 | 0.12 |
| ## UNEX078 | 0.19 | 0.30 | 0.12 | 0.19 | 0.21 | 0.09 | 0.22 | 0.18 | 0.13 |
| ## UNEX079 | 0.17 | 0.19 | 0.24 | 0.22 | 0.21 | 0.14 | 0.24 | 0.11 | 0.13 |
| ## UNEX082 | 0.04 | 0.15 | 0.08 | 0.11 | 0.12 | 0.08 | 0.16 | 0.17 | 0.19 |
| ## UNEX083 | 0.15 | 0.26 | 0.16 | 0.23 | 0.26 | 0.11 | 0.27 | 0.21 | 0.22 |
| ## UNEX087 | 0.18 | 0.28 | 0.13 | 0.25 | 0.23 | 0.13 | 0.32 | 0.21 | 0.17 |
| ## UNEX089 | 0.11 | 0.27 | 0.13 | 0.19 | 0.19 | 0.09 | 0.15 | 0.12 | 0.13 |
| ## UNEX090 | 0.13 | 0.29 | 0.23 | 0.28 | 0.17 | 0.14 | 0.28 | 0.13 | 0.22 |
| ## UNEX093 | 0.16 | 0.19 | 0.12 | 0.08 | 0.14 | 0.10 | 0.07 | 0.06 | 0.01 |
| ## UNEX099 | 0.24 | 0.17 | 0.21 | 0.24 | 0.23 | 0.17 | 0.33 | 0.15 | 0.16 |
| ## UNEX103 | 0.13 | 0.09 | 0.28 | 0.24 | 0.11 | 0.13 | 0.10 | 0.07 | 0.14 |
| ## INT004 | -0.05 | 0.12 | 0.00 | 0.07 | 0.00 | -0.02 | 0.06 | 0.15 | 0.00 |
| ## INT009 | -0.01 | 0.01 | -0.04 | 0.02 | 0.07 | 0.01 | 0.02 | 0.07 | -0.05 |
| ## INT011 | -0.01 | 0.09 | 0.02 | -0.01 | 0.05 | 0.04 | 0.02 | 0.07 | 0.07 |
| ## INT014 | 0.06 | 0.23 | 0.11 | 0.17 | 0.09 | 0.12 | 0.16 | 0.21 | 0.12 |
| ## INT026 | 0.11 | 0.21 | 0.12 | 0.11 | 0.18 | 0.11 | 0.11 | 0.17 | 0.07 |
| ## INT034 | 0.02 | 0.05 | 0.01 | -0.01 | 0.01 | 0.06 | 0.03 | 0.10 | 0.05 |
| ## INT035 | -0.09 | 0.07 | 0.03 | -0.01 | -0.02 | -0.03 | 0.01 | 0.07 | -0.02 |
| ## INT039 | -0.04 | -0.02 | 0.02 | -0.05 | 0.02 | 0.02 | 0.03 | 0.06 | 0.01 |
| ## INT041 | 0.00 | 0.08 | 0.00 | 0.12 | 0.04 | -0.01 | 0.05 | 0.07 | 0.10 |
| ## INT042 | -0.08 | -0.08 | -0.04 | -0.09 | -0.11 | -0.03 | -0.06 | -0.01 | 0.00 |
| ## INT045 | 0.03 | 0.08 | 0.08 | 0.08 | 0.05 | 0.05 | 0.12 | 0.05 | 0.13 |
| ## INT046 | 0.03 | 0.07 | 0.09 | 0.00 | 0.00 | 0.00 | 0.04 | 0.02 | -0.02 |

|  |  |  |  |  |  |  |  |  |  |  |
| --- | --- | --- | --- | --- | --- | --- | --- | --- | --- | --- |
| ## | INT048 | 0.12 | 0.21 | 0.05 | 0.14 | 0.15 | 0.08 | 0.12 | 0.19 | 0.08 |
| ## | INT049 | 0.00 | 0.16 | 0.08 | 0.14 | 0.08 | 0.01 | 0.13 | 0.08 | 0.08 |
| ## | INT050 | -0.02 | 0.07 | 0.07 | 0.03 | 0.05 | 0.02 | 0.05 | 0.09 | 0.07 |
| ## | INT059 | 0.00 | 0.15 | 0.06 | 0.13 | 0.10 | 0.05 | 0.14 | 0.19 | 0.16 |
| ## | INT064 | 0.01 | 0.14 | 0.11 | 0.13 | 0.10 | 0.13 | 0.06 | 0.13 | 0.15 |
| ## | INT067 | 0.00 | 0.03 | -0.01 | -0.02 | -0.02 | 0.01 | 0.04 | 0.09 | -0.02 |
| ## | INT077 | 0.01 | 0.02 | 0.03 | 0.02 | 0.03 | -0.05 | -0.02 | 0.00 | 0.00 |
| ## | INT080 | 0.02 | 0.12 | 0.11 | 0.13 | 0.12 | 0.08 | 0.18 | 0.18 | 0.19 |
| ## | INT086 | 0.03 | 0.05 | 0.02 | 0.02 | 0.07 | -0.02 | 0.08 | 0.16 | 0.04 |
| ## | INT092 | -0.07 | -0.04 | 0.00 | 0.00 | -0.01 | 0.01 | -0.05 | 0.04 | -0.03 |
| ## | INT094 | 0.03 | 0.07 | 0.05 | 0.06 | 0.05 | 0.01 | 0.02 | 0.05 | 0.08 |
| ## | INT097 | -0.07 | 0.03 | 0.03 | 0.01 | 0.05 | -0.05 | 0.01 | 0.07 | 0.02 |
| ## | INT098 | -0.01 | 0.08 | 0.07 | 0.08 | 0.04 | 0.01 | 0.05 | 0.11 | 0.12 |
| ## | INT102 | 0.06 | 0.18 | 0.11 | 0.13 | 0.12 | 0.06 | 0.19 | 0.10 | 0.19 |
| ## | INT104 | -0.02 | 0.11 | 0.01 | 0.03 | 0.04 | 0.00 | 0.07 | 0.08 | 0.06 |
| ## |  | UNEX044 | UNEX047 | UNEX058 | UNEX060 | UNEX063 | UNEX065 | UNEX071 | UNEX072 | UNEX076 |
| ## | COG001 | 0.05 | 0.17 | 0.12 | 0.02 | 0.11 | 0.04 | 0.07 | 0.05 | 0.02 |
| ## | COG008 | 0.12 | 0.16 | 0.07 | 0.10 | 0.11 | 0.09 | 0.09 | 0.10 | 0.07 |
| ## | COG016 | 0.12 | 0.16 | 0.13 | 0.11 | 0.15 | 0.12 | 0.08 | 0.10 | 0.03 |
| ## | COG021 | 0.09 | 0.27 | 0.11 | 0.08 | 0.10 | 0.05 | 0.11 | 0.09 | 0.04 |
| ## | COG022 | 0.19 | 0.23 | 0.15 | 0.19 | 0.21 | 0.15 | 0.13 | 0.21 | 0.09 |
| ## | COG023 | 0.07 | 0.17 | 0.12 | 0.09 | 0.11 | 0.05 | 0.07 | 0.11 | 0.06 |
| ## | COG028 | 0.06 | 0.20 | 0.20 | 0.11 | 0.18 | 0.12 | 0.08 | 0.18 | 0.06 |
| ## | COG032 | 0.09 | 0.18 | 0.12 | 0.13 | 0.15 | 0.13 | 0.08 | 0.14 | 0.01 |
| ## | COG036 | 0.11 | 0.20 | 0.10 | 0.10 | 0.17 | 0.12 | 0.15 | 0.14 | 0.06 |
| ## | COG037 | 0.11 | 0.16 | 0.14 | 0.16 | 0.15 | 0.15 | 0.12 | 0.15 | 0.05 |
| ## | COG038 | 0.08 | 0.17 | 0.09 | 0.02 | 0.14 | 0.09 | 0.03 | 0.07 | 0.04 |
| ## | COG043 | 0.17 | 0.16 | 0.17 | 0.16 | 0.21 | 0.14 | 0.14 | 0.21 | 0.05 |
| ## | COG054 | 0.21 | 0.17 | 0.09 | 0.15 | 0.13 | 0.12 | 0.12 | 0.13 | 0.09 |
| ## | COG055 | 0.14 | 0.16 | 0.14 | 0.10 | 0.22 | 0.12 | 0.07 | 0.11 | 0.02 |
| ## | COG061 | 0.14 | 0.22 | 0.11 | 0.13 | 0.18 | 0.11 | 0.15 | 0.12 | 0.05 |
| ## | COG062 | 0.16 | 0.23 | 0.16 | 0.14 | 0.15 | 0.10 | 0.15 | 0.17 | 0.07 |
| ## | COG066 | 0.04 | 0.14 | 0.12 | 0.05 | 0.11 | 0.04 | 0.06 | 0.03 | 0.04 |
| ## | COG073 | 0.14 | 0.13 | 0.09 | 0.19 | 0.17 | 0.13 | 0.09 | 0.14 | 0.04 |
| ## | COG075 | 0.18 | 0.21 | 0.10 | 0.24 | 0.26 | 0.18 | 0.10 | 0.23 | 0.02 |
| ## | COG081 | 0.18 | 0.24 | 0.15 | 0.25 | 0.21 | 0.16 | 0.14 | 0.16 | 0.07 |
| ## | COG085 | 0.09 | 0.15 | 0.10 | 0.08 | 0.17 | 0.05 | 0.09 | 0.11 | 0.00 |
| ## | COG091 | 0.04 | 0.14 | 0.11 | 0.03 | 0.12 | 0.05 | 0.03 | 0.07 | 0.04 |
| ## | COG095 | 0.11 | 0.15 | 0.14 | 0.17 | 0.21 | 0.12 | 0.10 | 0.18 | 0.02 |
| ## | COG101 | 0.13 | 0.20 | 0.10 | 0.11 | 0.15 | 0.08 | 0.10 | 0.13 | 0.08 |
| ## | IMP002 | 0.10 | 0.11 | 0.09 | 0.11 | 0.12 | 0.11 | 0.13 | 0.04 | 0.03 |
| ## | IMP007 | 0.03 | -0.07 | 0.04 | 0.06 | -0.05 | 0.02 | -0.01 | -0.02 | -0.05 |
| ## | IMP010 | 0.07 | 0.06 | 0.04 | 0.05 | 0.08 | 0.03 | 0.02 | 0.00 | 0.02 |
| ## | IMP012 | 0.27 | 0.16 | 0.20 | 0.29 | 0.19 | 0.14 | 0.12 | 0.18 | -0.02 |
| ## | IMP013 | 0.04 | 0.02 | -0.01 | 0.07 | 0.05 | 0.04 | 0.02 | 0.02 | 0.07 |
| ## | IMP015 | 0.11 | 0.17 | 0.10 | 0.19 | 0.13 | 0.12 | 0.11 | 0.22 | 0.06 |
| ## | IMP018 | -0.08 | -0.09 | -0.03 | -0.10 | -0.10 | -0.01 | -0.04 | -0.01 | -0.02 |
| ## | IMP019 | 0.12 | 0.09 | 0.09 | 0.13 | 0.09 | 0.10 | 0.05 | 0.11 | 0.04 |
| ## | IMP024 | 0.07 | 0.03 | 0.03 | 0.09 | -0.01 | 0.04 | 0.03 | 0.02 | -0.01 |
| ## | IMP031 | 0.04 | 0.09 | 0.05 | 0.10 | 0.05 | 0.09 | 0.05 | 0.07 | 0.01 |
| ## | IMP051 | 0.04 | -0.01 | -0.02 | 0.04 | -0.05 | 0.02 | -0.03 | 0.04 | -0.03 |
| ## | IMP052 | 0.06 | 0.11 | 0.09 | 0.12 | 0.03 | 0.09 | 0.08 | 0.08 | 0.04 |
| ## | IMP053 | 0.13 | 0.15 | 0.09 | 0.18 | 0.12 | 0.15 | 0.13 | 0.15 | 0.11 |
| ## | IMP056 | 0.09 | 0.19 | 0.09 | 0.08 | 0.17 | 0.16 | 0.11 | 0.21 | 0.10 |

|  |  |  |  |  |  |  |  |  |  |
| --- | --- | --- | --- | --- | --- | --- | --- | --- | --- |
| ## IMP057 | 0.04 | -0.08 | -0.05 | 0.01 | -0.07 | 0.00 | 0.02 | -0.05 | -0.03 |
| ## IMP068 | 0.01 | 0.05 | 0.09 | 0.09 | 0.01 | 0.08 | 0.04 | 0.01 | 0.03 |
| ## IMP069 | 0.06 | 0.02 | 0.09 | 0.14 | 0.12 | 0.04 | 0.09 | 0.04 | 0.07 |
| ## IMP070 | 0.16 | 0.12 | 0.11 | 0.23 | 0.04 | 0.10 | 0.06 | 0.14 | 0.07 |
| ## IMP074 | 0.25 | 0.13 | 0.04 | 0.15 | 0.12 | 0.17 | 0.10 | 0.18 | 0.05 |
| ## IMP084 | 0.09 | 0.08 | 0.08 | 0.10 | 0.06 | 0.05 | 0.10 | 0.01 | 0.04 |
| ## IMP088 | 0.06 | 0.06 | 0.06 | 0.08 | 0.11 | 0.14 | 0.11 | 0.07 | 0.07 |
| ## IMP096 | 0.21 | 0.20 | 0.24 | 0.25 | 0.14 | 0.16 | 0.12 | 0.21 | 0.13 |
| ## IMP100 | 0.16 | 0.14 | 0.13 | 0.19 | 0.17 | 0.15 | 0.09 | 0.09 | 0.07 |
| ## UNEX003 | 0.21 | 0.09 | 0.13 | 0.08 | 0.12 | 0.10 | 0.09 | 0.07 | 0.04 |
| ## UNEX005 | 0.20 | 0.18 | 0.23 | 0.17 | 0.26 | 0.21 | 0.12 | 0.18 | 0.11 |
| ## UNEX006 | 0.12 | 0.14 | 0.18 | 0.15 | 0.17 | 0.17 | 0.08 | 0.12 | 0.12 |
| ## UNEX017 | 0.19 | 0.29 | 0.20 | 0.19 | 0.24 | 0.20 | 0.09 | 0.19 | 0.08 |
| ## UNEX020 | 0.17 | 0.08 | 0.23 | 0.27 | 0.19 | 0.24 | 0.10 | 0.26 | 0.16 |
| ## UNEX025 | 0.25 | 0.15 | 0.37 | 0.24 | 0.21 | 0.26 | 0.15 | 0.28 | 0.08 |
| ## UNEX027 | 0.15 | 0.18 | 0.25 | 0.21 | 0.19 | 0.23 | 0.09 | 0.19 | 0.09 |
| ## UNEX029 | 0.19 | 0.08 | 0.16 | 0.12 | 0.16 | 0.18 | 0.04 | 0.18 | 0.06 |
| ## UNEX030 | 0.31 | 0.20 | 0.22 | 0.28 | 0.25 | 0.21 | 0.19 | 0.18 | 0.11 |
| ## UNEX033 | 0.15 | 0.17 | 0.14 | 0.23 | 0.14 | 0.12 | 0.09 | 0.24 | 0.06 |
| ## UNEX040 | 0.23 | 0.06 | 0.17 | 0.32 | 0.15 | 0.21 | 0.08 | 0.24 | 0.12 |
| ## UNEX044 | 1.00 | 0.10 | 0.16 | 0.24 | 0.13 | 0.13 | 0.14 | 0.18 | 0.07 |
| ## UNEX047 | 0.10 | 1.00 | 0.15 | 0.17 | 0.10 | 0.18 | 0.13 | 0.19 | 0.09 |
| ## UNEX058 | 0.16 | 0.15 | 1.00 | 0.23 | 0.21 | 0.22 | 0.13 | 0.19 | 0.13 |
| ## UNEX060 | 0.24 | 0.17 | 0.23 | 1.00 | 0.14 | 0.16 | 0.08 | 0.19 | 0.14 |
| ## UNEX063 | 0.13 | 0.10 | 0.21 | 0.14 | 1.00 | 0.19 | 0.20 | 0.23 | 0.10 |
| ## UNEX065 | 0.13 | 0.18 | 0.22 | 0.16 | 0.19 | 1.00 | 0.18 | 0.18 | 0.16 |
| ## UNEX071 | 0.14 | 0.13 | 0.13 | 0.08 | 0.20 | 0.18 | 1.00 | 0.15 | 0.22 |
| ## UNEX072 | 0.18 | 0.19 | 0.19 | 0.19 | 0.23 | 0.18 | 0.15 | 1.00 | 0.26 |
| ## UNEX076 | 0.07 | 0.09 | 0.13 | 0.14 | 0.10 | 0.16 | 0.22 | 0.26 | 1.00 |
| ## UNEX078 | 0.15 | 0.35 | 0.17 | 0.21 | 0.19 | 0.22 | 0.18 | 0.24 | 0.18 |
| ## UNEX079 | 0.13 | 0.12 | 0.25 | 0.15 | 0.23 | 0.21 | 0.08 | 0.12 | 0.11 |
| ## UNEX082 | 0.14 | 0.17 | 0.08 | 0.09 | 0.12 | 0.11 | 0.08 | 0.32 | 0.20 |
| ## UNEX083 | 0.16 | 0.21 | 0.20 | 0.14 | 0.26 | 0.22 | 0.14 | 0.32 | 0.14 |
| ## UNEX087 | 0.22 | 0.33 | 0.17 | 0.25 | 0.17 | 0.16 | 0.12 | 0.22 | 0.06 |
| ## UNEX089 | 0.16 | 0.21 | 0.17 | 0.10 | 0.24 | 0.18 | 0.16 | 0.18 | 0.17 |
| ## UNEX090 | 0.21 | 0.14 | 0.21 | 0.20 | 0.32 | 0.27 | 0.21 | 0.19 | 0.12 |
| ## UNEX093 | 0.03 | 0.25 | 0.16 | 0.06 | 0.13 | 0.13 | 0.02 | 0.08 | 0.12 |
| ## UNEX099 | 0.25 | 0.19 | 0.32 | 0.17 | 0.33 | 0.20 | 0.11 | 0.21 | 0.07 |
| ## UNEX103 | 0.16 | 0.12 | 0.26 | 0.14 | 0.07 | 0.18 | 0.12 | 0.14 | 0.10 |
| ## INT004 | 0.09 | 0.06 | 0.06 | 0.01 | 0.06 | 0.05 | 0.04 | 0.05 | -0.03 |
| ## INT009 | 0.01 | 0.04 | 0.02 | 0.11 | -0.05 | -0.01 | -0.02 | -0.06 | -0.04 |
| ## INT011 | 0.01 | 0.07 | 0.00 | 0.01 | 0.05 | 0.01 | 0.03 | 0.00 | 0.01 |
| ## INT014 | 0.16 | 0.16 | 0.10 | 0.24 | 0.13 | 0.16 | 0.04 | 0.09 | 0.07 |
| ## INT026 | 0.11 | 0.20 | 0.10 | 0.21 | 0.07 | 0.06 | 0.01 | 0.09 | 0.03 |
| ## INT034 | 0.13 | 0.04 | 0.01 | 0.15 | 0.02 | 0.01 | 0.00 | 0.03 | 0.01 |
| ## INT035 | 0.04 | 0.14 | 0.00 | 0.02 | 0.02 | 0.05 | 0.01 | 0.04 | 0.03 |
| ## INT039 | 0.00 | -0.06 | -0.03 | 0.05 | -0.01 | -0.03 | 0.02 | 0.01 | -0.01 |
| ## INT041 | 0.08 | 0.07 | 0.09 | 0.14 | 0.05 | 0.05 | 0.02 | 0.08 | 0.02 |
| ## INT042 | -0.05 | -0.06 | -0.07 | -0.03 | -0.10 | -0.06 | -0.06 | -0.02 | 0.01 |
| ## INT045 | 0.12 | 0.05 | 0.12 | 0.19 | 0.06 | 0.10 | 0.01 | 0.07 | 0.07 |
| ## INT046 | 0.00 | 0.10 | 0.01 | 0.07 | 0.02 | 0.01 | 0.06 | -0.05 | 0.04 |
| ## INT048 | 0.08 | 0.23 | 0.05 | 0.15 | 0.14 | 0.11 | 0.13 | 0.19 | 0.09 |
| ## INT049 | 0.15 | 0.10 | 0.04 | 0.15 | 0.09 | 0.03 | 0.06 | 0.09 | 0.01 |
| ## INT050 | 0.03 | 0.08 | 0.03 | 0.07 | 0.09 | 0.04 | 0.04 | 0.07 | 0.01 |

|  |  |  |  |  |  |  |  |  |  |
| --- | --- | --- | --- | --- | --- | --- | --- | --- | --- |
| ## INT059 | 0.18 | 0.04 | 0.05 | 0.13 | 0.08 | 0.12 | 0.10 | 0.09 | 0.00 |
| ## INT064 | 0.14 | 0.13 | 0.09 | 0.23 | 0.08 | 0.03 | 0.11 | 0.15 | 0.11 |
| ## INT067 | 0.01 | 0.02 | -0.01 | 0.02 | 0.04 | 0.01 | 0.06 | 0.00 | 0.01 |
| ## INT077 | 0.01 | 0.08 | 0.07 | 0.07 | 0.04 | -0.01 | -0.01 | 0.03 | 0.03 |
| ## INT080 | 0.15 | 0.09 | 0.10 | 0.19 | 0.16 | 0.11 | 0.08 | 0.19 | 0.06 |
| ## INT086 | 0.04 | 0.11 | 0.04 | 0.06 | 0.06 | 0.00 | 0.06 | 0.10 | 0.02 |
| ## INT092 | 0.01 | 0.00 | -0.04 | 0.01 | -0.03 | -0.01 | -0.01 | 0.00 | -0.02 |
| ## INT094 | 0.05 | 0.04 | 0.05 | 0.05 | 0.06 | 0.06 | 0.07 | 0.02 | 0.00 |
| ## INT097 | 0.01 | 0.12 | 0.05 | 0.02 | 0.07 | 0.06 | 0.07 | 0.03 | -0.02 |
| ## INT098 | 0.05 | 0.08 | 0.04 | 0.16 | 0.08 | 0.04 | 0.05 | 0.10 | 0.03 |
| ## INT102 | 0.20 | 0.12 | 0.12 | 0.17 | 0.07 | 0.11 | 0.07 | 0.09 | 0.01 |
| ## INT104 | 0.07 | 0.17 | 0.03 | 0.05 | -0.01 | 0.05 | 0.07 | 0.05 | 0.02 |
| ## | UNEX078 | UNEX079 | UNEX082 | UNEX083 | UNEX087 | UNEX089 | UNEX090 | UNEX093 | UNEX099 |
| ## COG001 | 0.12 | 0.10 | 0.06 | 0.14 | 0.13 | 0.10 | 0.07 | 0.05 | 0.07 |
| ## COG008 | 0.16 | 0.07 | 0.12 | 0.18 | 0.19 | 0.15 | 0.18 | 0.05 | 0.13 |
| ## COG016 | 0.17 | 0.13 | 0.09 | 0.17 | 0.12 | 0.14 | 0.11 | 0.05 | 0.20 |
| ## COG021 | 0.20 | 0.02 | 0.09 | 0.17 | 0.16 | 0.13 | 0.06 | 0.18 | 0.01 |
| ## COG022 | 0.24 | 0.11 | 0.20 | 0.28 | 0.29 | 0.19 | 0.20 | 0.16 | 0.24 |
| ## COG023 | 0.19 | 0.09 | 0.09 | 0.22 | 0.13 | 0.18 | 0.07 | 0.10 | 0.12 |
| ## COG028 | 0.22 | 0.13 | 0.11 | 0.24 | 0.14 | 0.21 | 0.13 | 0.12 | 0.16 |
| ## COG032 | 0.16 | 0.12 | 0.06 | 0.22 | 0.14 | 0.16 | 0.19 | 0.10 | 0.17 |
| ## COG036 | 0.20 | 0.11 | 0.13 | 0.15 | 0.20 | 0.18 | 0.14 | 0.08 | 0.18 |
| ## COG037 | 0.21 | 0.09 | 0.15 | 0.21 | 0.14 | 0.19 | 0.18 | 0.13 | 0.14 |
| ## COG038 | 0.13 | 0.07 | 0.11 | 0.14 | 0.12 | 0.14 | 0.14 | 0.06 | 0.11 |
| ## COG043 | 0.22 | 0.17 | 0.15 | 0.20 | 0.23 | 0.21 | 0.18 | 0.07 | 0.19 |
| ## COG054 | 0.21 | 0.06 | 0.12 | 0.17 | 0.19 | 0.16 | 0.15 | 0.14 | 0.17 |
| ## COG055 | 0.14 | 0.19 | 0.10 | 0.19 | 0.08 | 0.17 | 0.18 | 0.12 | 0.25 |
| ## COG061 | 0.19 | 0.15 | 0.11 | 0.18 | 0.21 | 0.18 | 0.14 | 0.15 | 0.21 |
| ## COG062 | 0.26 | 0.09 | 0.15 | 0.25 | 0.23 | 0.15 | 0.10 | 0.13 | 0.18 |
| ## COG066 | 0.10 | 0.12 | 0.07 | 0.09 | 0.09 | 0.14 | 0.10 | 0.03 | 0.14 |
| ## COG073 | 0.14 | 0.11 | 0.09 | 0.16 | 0.13 | 0.16 | 0.16 | 0.08 | 0.18 |
| ## COG075 | 0.21 | 0.17 | 0.23 | 0.24 | 0.26 | 0.21 | 0.24 | 0.08 | 0.22 |
| ## COG081 | 0.25 | 0.21 | 0.10 | 0.20 | 0.22 | 0.20 | 0.21 | 0.13 | 0.28 |
| ## COG085 | 0.15 | 0.07 | 0.11 | 0.17 | 0.16 | 0.13 | 0.07 | 0.05 | 0.13 |
| ## COG091 | 0.10 | 0.04 | 0.10 | 0.16 | 0.16 | 0.08 | 0.08 | 0.10 | 0.08 |
| ## COG095 | 0.26 | 0.12 | 0.16 | 0.24 | 0.25 | 0.21 | 0.21 | 0.12 | 0.18 |
| ## COG101 | 0.20 | 0.12 | 0.11 | 0.17 | 0.19 | 0.23 | 0.16 | 0.15 | 0.12 |
| ## IMP002 | 0.19 | 0.14 | 0.09 | 0.19 | 0.09 | 0.20 | 0.22 | 0.05 | 0.12 |
| ## IMP007 | -0.04 | 0.02 | -0.02 | -0.04 | -0.07 | -0.04 | 0.03 | -0.03 | 0.03 |
| ## IMP010 | 0.05 | 0.06 | 0.04 | 0.04 | 0.10 | 0.06 | 0.09 | 0.06 | 0.08 |
| ## IMP012 | 0.23 | 0.12 | 0.10 | 0.25 | 0.23 | 0.13 | 0.17 | 0.03 | 0.20 |
| ## IMP013 | 0.04 | 0.09 | 0.04 | 0.05 | 0.03 | 0.07 | 0.00 | -0.02 | 0.03 |
| ## IMP015 | 0.19 | 0.10 | 0.20 | 0.23 | 0.16 | 0.14 | 0.14 | 0.10 | 0.14 |
| ## IMP018 | -0.08 | -0.04 | -0.06 | -0.04 | -0.09 | -0.04 | -0.06 | -0.02 | -0.09 |
| ## IMP019 | 0.14 | 0.07 | 0.13 | 0.14 | 0.15 | 0.10 | 0.09 | 0.07 | 0.07 |
| ## IMP024 | 0.10 | 0.07 | 0.05 | 0.09 | 0.12 | 0.11 | 0.10 | -0.02 | 0.09 |
| ## IMP031 | 0.07 | 0.10 | 0.03 | 0.06 | 0.05 | 0.06 | 0.08 | 0.02 | 0.07 |
| ## IMP051 | -0.02 | -0.07 | -0.01 | -0.07 | 0.01 | -0.04 | -0.02 | 0.01 | -0.02 |
| ## IMP052 | 0.12 | 0.10 | 0.05 | 0.11 | 0.10 | 0.04 | 0.09 | 0.10 | 0.09 |
| ## IMP053 | 0.14 | 0.11 | 0.11 | 0.26 | 0.15 | 0.18 | 0.19 | 0.10 | 0.16 |
| ## IMP056 | 0.13 | 0.09 | 0.21 | 0.30 | 0.16 | 0.21 | 0.21 | 0.04 | 0.16 |
| ## IMP057 | -0.01 | 0.00 | -0.09 | -0.09 | -0.06 | -0.03 | -0.03 | -0.03 | -0.03 |
| ## IMP068 | 0.01 | 0.07 | 0.00 | 0.05 | 0.05 | 0.04 | 0.09 | 0.07 | 0.02 |
| ## IMP069 | 0.06 | 0.12 | 0.02 | 0.10 | 0.08 | 0.08 | 0.10 | 0.06 | 0.06 |

|  |  |  |  |  |  |  |  |  |  |
| --- | --- | --- | --- | --- | --- | --- | --- | --- | --- |
| ## IMP070 | 0.11 | 0.04 | 0.07 | 0.09 | 0.14 | 0.09 | 0.12 | 0.08 | 0.11 |
| ## IMP074 | 0.18 | 0.15 | 0.08 | 0.18 | 0.16 | 0.12 | 0.13 | 0.06 | 0.13 |
| ## IMP084 | 0.13 | 0.10 | 0.01 | 0.08 | 0.09 | 0.09 | 0.11 | 0.05 | 0.08 |
| ## IMP088 | 0.15 | 0.10 | 0.10 | 0.17 | 0.12 | 0.11 | 0.12 | 0.07 | 0.14 |
| ## IMP096 | 0.25 | 0.17 | 0.19 | 0.27 | 0.26 | 0.24 | 0.16 | 0.12 | 0.23 |
| ## IMP100 | 0.17 | 0.16 | 0.07 | 0.19 | 0.15 | 0.14 | 0.25 | 0.12 | 0.16 |
| ## UNEX003 | 0.10 | 0.11 | 0.03 | 0.07 | 0.10 | 0.12 | 0.11 | 0.10 | 0.23 |
| ## UNEX005 | 0.22 | 0.18 | 0.12 | 0.29 | 0.21 | 0.24 | 0.22 | 0.15 | 0.27 |
| ## UNEX006 | 0.19 | 0.17 | 0.04 | 0.15 | 0.18 | 0.11 | 0.13 | 0.16 | 0.24 |
| ## UNEX017 | 0.30 | 0.19 | 0.15 | 0.26 | 0.28 | 0.27 | 0.29 | 0.19 | 0.17 |
| ## UNEX020 | 0.12 | 0.24 | 0.08 | 0.16 | 0.13 | 0.13 | 0.23 | 0.12 | 0.21 |
| ## UNEX025 | 0.19 | 0.22 | 0.11 | 0.23 | 0.25 | 0.19 | 0.28 | 0.08 | 0.24 |
| ## UNEX027 | 0.21 | 0.21 | 0.12 | 0.26 | 0.23 | 0.19 | 0.17 | 0.14 | 0.23 |
| ## UNEX029 | 0.09 | 0.14 | 0.08 | 0.11 | 0.13 | 0.09 | 0.14 | 0.10 | 0.17 |
| ## UNEX030 | 0.22 | 0.24 | 0.16 | 0.27 | 0.32 | 0.15 | 0.28 | 0.07 | 0.33 |
| ## UNEX033 | 0.18 | 0.11 | 0.17 | 0.21 | 0.21 | 0.12 | 0.13 | 0.06 | 0.15 |
| ## UNEX040 | 0.13 | 0.13 | 0.19 | 0.22 | 0.17 | 0.13 | 0.22 | 0.01 | 0.16 |
| ## UNEX044 | 0.15 | 0.13 | 0.14 | 0.16 | 0.22 | 0.16 | 0.21 | 0.03 | 0.25 |
| ## UNEX047 | 0.35 | 0.12 | 0.17 | 0.21 | 0.33 | 0.21 | 0.14 | 0.25 | 0.19 |
| ## UNEX058 | 0.17 | 0.25 | 0.08 | 0.20 | 0.17 | 0.17 | 0.21 | 0.16 | 0.32 |
| ## UNEX060 | 0.21 | 0.15 | 0.09 | 0.14 | 0.25 | 0.10 | 0.20 | 0.06 | 0.17 |
| ## UNEX063 | 0.19 | 0.23 | 0.12 | 0.26 | 0.17 | 0.24 | 0.32 | 0.13 | 0.33 |
| ## UNEX065 | 0.22 | 0.21 | 0.11 | 0.22 | 0.16 | 0.18 | 0.27 | 0.13 | 0.20 |
| ## UNEX071 | 0.18 | 0.08 | 0.08 | 0.14 | 0.12 | 0.16 | 0.21 | 0.02 | 0.11 |
| ## UNEX072 | 0.24 | 0.12 | 0.32 | 0.32 | 0.22 | 0.18 | 0.19 | 0.08 | 0.21 |
| ## UNEX076 | 0.18 | 0.11 | 0.20 | 0.14 | 0.06 | 0.17 | 0.12 | 0.12 | 0.07 |
| ## UNEX078 | 1.00 | 0.20 | 0.19 | 0.27 | 0.20 | 0.20 | 0.27 | 0.18 | 0.20 |
| ## UNEX079 | 0.20 | 1.00 | 0.11 | 0.17 | 0.17 | 0.18 | 0.30 | 0.12 | 0.27 |
| ## UNEX082 | 0.19 | 0.11 | 1.00 | 0.24 | 0.15 | 0.14 | 0.19 | 0.03 | 0.14 |
| ## UNEX083 | 0.27 | 0.17 | 0.24 | 1.00 | 0.22 | 0.25 | 0.23 | 0.19 | 0.28 |
| ## UNEX087 | 0.20 | 0.17 | 0.15 | 0.22 | 1.00 | 0.17 | 0.18 | 0.13 | 0.22 |
| ## UNEX089 | 0.20 | 0.18 | 0.14 | 0.25 | 0.17 | 1.00 | 0.37 | 0.13 | 0.24 |
| ## UNEX090 | 0.27 | 0.30 | 0.19 | 0.23 | 0.18 | 0.37 | 1.00 | 0.09 | 0.30 |
| ## UNEX093 | 0.18 | 0.12 | 0.03 | 0.19 | 0.13 | 0.13 | 0.09 | 1.00 | 0.12 |
| ## UNEX099 | 0.20 | 0.27 | 0.14 | 0.28 | 0.22 | 0.24 | 0.30 | 0.12 | 1.00 |
| ## UNEX103 | 0.14 | 0.28 | 0.01 | 0.10 | 0.18 | 0.10 | 0.17 | 0.11 | 0.20 |
| ## INT004 | 0.13 | 0.06 | 0.13 | 0.13 | 0.07 | 0.11 | 0.10 | 0.08 | 0.06 |
| ## INT009 | 0.05 | 0.03 | -0.04 | 0.00 | 0.00 | -0.02 | -0.01 | 0.02 | -0.04 |
| ## INT011 | 0.06 | 0.00 | 0.01 | 0.03 | 0.02 | 0.05 | 0.07 | -0.02 | 0.00 |
| ## INT014 | 0.17 | 0.06 | 0.14 | 0.14 | 0.17 | 0.11 | 0.21 | 0.05 | 0.09 |
| ## INT026 | 0.15 | 0.09 | 0.19 | 0.09 | 0.13 | 0.12 | 0.16 | 0.06 | 0.10 |
| ## INT034 | 0.02 | -0.03 | 0.03 | 0.02 | 0.07 | 0.05 | 0.07 | 0.05 | 0.01 |
| ## INT035 | 0.07 | 0.00 | 0.04 | 0.04 | 0.06 | 0.01 | 0.02 | 0.03 | 0.03 |
| ## INT039 | 0.00 | -0.09 | 0.05 | -0.01 | 0.04 | -0.02 | 0.00 | 0.00 | -0.01 |
| ## INT041 | 0.02 | 0.03 | 0.04 | 0.05 | 0.12 | 0.07 | 0.10 | 0.05 | 0.05 |
| ## INT042 | -0.07 | -0.10 | -0.05 | -0.08 | -0.06 | -0.07 | -0.05 | -0.06 | -0.11 |
| ## INT045 | 0.08 | 0.10 | 0.09 | 0.08 | 0.10 | 0.06 | 0.10 | 0.04 | 0.09 |
| ## INT046 | 0.06 | 0.07 | -0.01 | -0.03 | 0.01 | 0.03 | 0.08 | 0.02 | -0.02 |
| ## INT048 | 0.26 | 0.05 | 0.21 | 0.19 | 0.20 | 0.20 | 0.12 | 0.13 | 0.15 |
| ## INT049 | 0.16 | 0.02 | 0.12 | 0.09 | 0.14 | 0.06 | 0.12 | 0.03 | 0.08 |
| ## INT050 | 0.12 | 0.02 | 0.08 | 0.06 | 0.12 | 0.02 | 0.07 | 0.07 | 0.07 |
| ## INT059 | 0.13 | 0.04 | 0.11 | 0.16 | 0.17 | 0.14 | 0.17 | 0.01 | 0.09 |
| ## INT064 | 0.16 | 0.02 | 0.18 | 0.08 | 0.16 | 0.13 | 0.06 | 0.04 | 0.05 |
| ## INT067 | 0.04 | -0.06 | 0.02 | -0.02 | 0.07 | 0.04 | 0.02 | 0.00 | -0.02 |

|  |  |  |  |  |  |  |  |  |  |  |
| --- | --- | --- | --- | --- | --- | --- | --- | --- | --- | --- |
| ## | INT077 | 0.07 | 0.01 | 0.06 | 0.03 | 0.05 | 0.02 | 0.01 | 0.07 | 0.02 |
| ## | INT080 | 0.18 | 0.11 | 0.12 | 0.15 | 0.18 | 0.12 | 0.19 | 0.06 | 0.16 |
| ## | INT086 | 0.10 | -0.02 | 0.08 | 0.06 | 0.11 | 0.04 | 0.02 | 0.00 | 0.03 |
| ## | INT092 | 0.01 | -0.09 | 0.00 | -0.01 | -0.01 | -0.01 | 0.03 | -0.03 | -0.05 |
| ## | INT094 | 0.10 | 0.00 | 0.02 | 0.05 | 0.09 | 0.10 | 0.07 | 0.00 | 0.01 |
| ## | INT097 | 0.06 | -0.01 | 0.02 | 0.05 | 0.05 | 0.07 | 0.06 | 0.08 | 0.00 |
| ## | INT098 | 0.08 | 0.04 | 0.12 | 0.07 | 0.09 | 0.04 | 0.08 | 0.03 | 0.03 |
| ## | INT102 | 0.15 | 0.06 | 0.12 | 0.15 | 0.16 | 0.14 | 0.17 | 0.10 | 0.08 |
| ## | INT104 | 0.14 | 0.03 | 0.10 | 0.10 | 0.06 | 0.08 | 0.14 | 0.07 | 0.01 |
| ## | UNEX103 | INT004 | INT009 | INT011 | INT014 | INT026 | INT034 | INT035 | INT039 | INT041 |
| ## | COG001 | 0.02 | 0.12 | 0.11 | 0.08 | 0.15 | 0.08 | 0.05 | 0.19 | 0.01 |
| ## | COG008 | 0.02 | 0.14 | 0.04 | 0.07 | 0.20 | 0.10 | 0.05 | 0.14 | 0.05 |
| ## | COG016 | 0.02 | 0.10 | 0.06 | 0.04 | 0.15 | 0.10 | 0.06 | 0.13 | 0.00 |
| ## | COG021 | -0.02 | 0.09 | 0.07 | 0.02 | 0.09 | 0.15 | 0.05 | 0.15 | -0.03 |
| ## | COG022 | 0.06 | 0.08 | 0.03 | 0.04 | 0.15 | 0.17 | -0.01 | 0.03 | 0.05 |
| ## | COG023 | 0.03 | 0.10 | 0.04 | 0.04 | 0.12 | 0.11 | -0.04 | 0.10 | 0.01 |
| ## | COG028 | 0.04 | 0.14 | 0.04 | 0.07 | 0.20 | 0.15 | 0.02 | 0.17 | -0.04 |
| ## | COG032 | 0.11 | 0.13 | 0.00 | 0.06 | 0.13 | 0.10 | 0.06 | 0.01 | 0.02 |
| ## | COG036 | 0.03 | 0.13 | 0.06 | 0.08 | 0.14 | 0.08 | 0.03 | 0.12 | -0.01 |
| ## | COG037 | 0.08 | 0.19 | 0.08 | 0.05 | 0.28 | 0.23 | 0.09 | 0.04 | 0.09 |
| ## | COG038 | 0.06 | 0.10 | 0.11 | 0.09 | 0.17 | 0.17 | 0.03 | 0.17 | 0.04 |
| ## | COG043 | 0.08 | 0.12 | 0.06 | -0.02 | 0.20 | 0.16 | 0.02 | 0.07 | -0.02 |
| ## | COG054 | 0.01 | 0.19 | 0.03 | 0.02 | 0.30 | 0.19 | 0.15 | 0.13 | 0.05 |
| ## | COG055 | 0.01 | 0.08 | 0.01 | 0.02 | 0.14 | 0.08 | -0.03 | 0.00 | -0.09 |
| ## | COG061 | 0.05 | 0.13 | -0.02 | 0.06 | 0.12 | 0.16 | 0.03 | 0.07 | -0.03 |
| ## | COG062 | 0.05 | 0.14 | 0.05 | 0.07 | 0.12 | 0.12 | 0.02 | 0.08 | -0.01 |
| ## | COG066 | 0.00 | 0.10 | 0.07 | 0.03 | 0.13 | 0.09 | 0.04 | 0.12 | 0.02 |
| ## | COG073 | 0.08 | 0.19 | 0.07 | 0.10 | 0.28 | 0.14 | 0.03 | 0.15 | 0.05 |
| ## | COG075 | 0.12 | 0.22 | 0.06 | 0.09 | 0.40 | 0.24 | 0.08 | 0.18 | 0.03 |
| ## | COG081 | 0.07 | 0.22 | 0.05 | 0.04 | 0.23 | 0.22 | 0.04 | 0.17 | 0.04 |
| ## | COG085 | 0.03 | 0.11 | 0.02 | 0.07 | 0.06 | 0.07 | -0.03 | 0.02 | 0.00 |
| ## | COG091 | -0.03 | 0.14 | 0.01 | 0.07 | 0.17 | 0.11 | 0.01 | 0.10 | -0.01 |
| ## | COG095 | 0.05 | 0.14 | 0.02 | 0.09 | 0.18 | 0.10 | 0.03 | 0.09 | -0.02 |
| ## | COG101 | 0.05 | 0.16 | 0.01 | 0.11 | 0.17 | 0.13 | 0.00 | 0.01 | 0.02 |
| ## | IMP002 | 0.02 | 0.13 | 0.05 | -0.02 | 0.16 | 0.08 | 0.05 | 0.05 | 0.02 |
| ## | IMP007 | 0.09 | 0.04 | -0.09 | 0.00 | 0.00 | 0.05 | 0.04 | -0.12 | 0.01 |
| ## | IMP010 | 0.03 | -0.02 | 0.01 | -0.03 | 0.04 | 0.03 | 0.02 | -0.02 | 0.01 |
| ## | IMP012 | 0.10 | 0.16 | 0.05 | 0.03 | 0.25 | 0.25 | 0.15 | 0.08 | 0.08 |
| ## | IMP013 | -0.05 | -0.01 | 0.05 | 0.05 | 0.02 | -0.01 | -0.02 | -0.01 | 0.00 |
| ## | IMP015 | -0.01 | 0.16 | 0.00 | 0.07 | 0.19 | 0.20 | 0.09 | 0.12 | 0.03 |
| ## | IMP018 | 0.01 | -0.04 | -0.11 | -0.05 | -0.15 | -0.12 | -0.11 | -0.20 | -0.02 |
| ## | IMP019 | 0.01 | 0.06 | 0.05 | 0.01 | 0.14 | 0.11 | 0.03 | 0.06 | 0.02 |
| ## | IMP024 | 0.06 | 0.17 | 0.02 | 0.12 | 0.14 | 0.09 | 0.05 | 0.00 | 0.07 |
| ## | IMP031 | 0.05 | 0.06 | 0.03 | 0.04 | 0.06 | 0.04 | -0.06 | 0.03 | 0.03 |
| ## | IMP051 | 0.02 | 0.06 | 0.01 | -0.02 | 0.02 | 0.01 | 0.09 | 0.04 | 0.04 |
| ## | IMP052 | 0.11 | 0.03 | -0.02 | -0.05 | 0.05 | 0.10 | -0.01 | -0.04 | -0.02 |
| ## | IMP053 | 0.04 | 0.12 | 0.08 | 0.11 | 0.15 | 0.13 | -0.01 | 0.07 | -0.01 |
| ## | IMP056 | 0.03 | 0.12 | -0.02 | 0.08 | 0.16 | 0.11 | 0.02 | 0.07 | -0.01 |
| ## | IMP057 | 0.01 | -0.07 | -0.02 | -0.07 | -0.08 | -0.03 | 0.00 | -0.13 | 0.02 |
| ## | IMP068 | 0.07 | 0.02 | 0.02 | -0.03 | 0.04 | 0.14 | 0.07 | 0.04 | -0.02 |
| ## | IMP069 | 0.06 | 0.06 | 0.05 | 0.02 | 0.05 | 0.05 | 0.08 | 0.02 | 0.03 |
| ## | IMP070 | 0.05 | 0.11 | 0.09 | 0.03 | 0.23 | 0.14 | 0.10 | 0.08 | 0.05 |
| ## | IMP074 | 0.05 | 0.10 | 0.02 | 0.10 | 0.17 | 0.14 | 0.01 | 0.09 | 0.03 |
| ## | IMP084 | 0.08 | 0.03 | 0.03 | 0.04 | 0.05 | 0.13 | 0.04 | -0.03 | 0.02 |

|  |  |  |  |  |  |  |  |  |  |  |
| --- | --- | --- | --- | --- | --- | --- | --- | --- | --- | --- |
| ## IMP088 | 0.03 | 0.02 | 0.03 | 0.06 | 0.07 | 0.02 | -0.02 | -0.04 | 0.00 | -0.02 |
| ## IMP096 | 0.12 | 0.15 | 0.02 | 0.10 | 0.21 | 0.25 | 0.10 | 0.11 | 0.09 | 0.16 |
| ## IMP100 | 0.12 | 0.15 | 0.09 | 0.05 | 0.19 | 0.12 | -0.01 | 0.08 | 0.00 | 0.08 |
| ## UNEX003 | 0.15 | 0.05 | 0.01 | -0.01 | 0.06 | 0.04 | -0.01 | -0.03 | 0.03 | 0.07 |
| ## UNEX005 | 0.14 | 0.07 | 0.01 | 0.03 | 0.12 | 0.14 | 0.05 | 0.00 | -0.03 | 0.04 |
| ## UNEX006 | 0.13 | -0.05 | -0.01 | -0.01 | 0.06 | 0.11 | 0.02 | -0.09 | -0.04 | 0.00 |
| ## UNEX017 | 0.09 | 0.12 | 0.01 | 0.09 | 0.23 | 0.21 | 0.05 | 0.07 | -0.02 | 0.08 |
| ## UNEX020 | 0.28 | 0.00 | -0.04 | 0.02 | 0.11 | 0.12 | 0.01 | 0.03 | 0.02 | 0.00 |
| ## UNEX025 | 0.24 | 0.07 | 0.02 | -0.01 | 0.17 | 0.11 | -0.01 | -0.01 | -0.05 | 0.12 |
| ## UNEX027 | 0.11 | 0.00 | 0.07 | 0.05 | 0.09 | 0.18 | 0.01 | -0.02 | 0.02 | 0.04 |
| ## UNEX029 | 0.13 | -0.02 | 0.01 | 0.04 | 0.12 | 0.11 | 0.06 | -0.03 | 0.02 | -0.01 |
| ## UNEX030 | 0.10 | 0.06 | 0.02 | 0.02 | 0.16 | 0.11 | 0.03 | 0.01 | 0.03 | 0.05 |
| ## UNEX033 | 0.07 | 0.15 | 0.07 | 0.07 | 0.21 | 0.17 | 0.10 | 0.07 | 0.06 | 0.07 |
| ## UNEX040 | 0.14 | 0.00 | -0.05 | 0.07 | 0.12 | 0.07 | 0.05 | -0.02 | 0.01 | 0.10 |
| ## UNEX044 | 0.16 | 0.09 | 0.01 | 0.01 | 0.16 | 0.11 | 0.13 | 0.04 | 0.00 | 0.08 |
| ## UNEX047 | 0.12 | 0.06 | 0.04 | 0.07 | 0.16 | 0.20 | 0.04 | 0.14 | -0.06 | 0.07 |
| ## UNEX058 | 0.26 | 0.06 | 0.02 | 0.00 | 0.10 | 0.10 | 0.01 | 0.00 | -0.03 | 0.09 |
| ## UNEX060 | 0.14 | 0.01 | 0.11 | 0.01 | 0.24 | 0.21 | 0.15 | 0.02 | 0.05 | 0.14 |
| ## UNEX063 | 0.07 | 0.06 | -0.05 | 0.05 | 0.13 | 0.07 | 0.02 | 0.02 | -0.01 | 0.05 |
| ## UNEX065 | 0.18 | 0.05 | -0.01 | 0.01 | 0.16 | 0.06 | 0.01 | 0.05 | -0.03 | 0.05 |
| ## UNEX071 | 0.12 | 0.04 | -0.02 | 0.03 | 0.04 | 0.01 | 0.00 | 0.01 | 0.02 | 0.02 |
| ## UNEX072 | 0.14 | 0.05 | -0.06 | 0.00 | 0.09 | 0.09 | 0.03 | 0.04 | 0.01 | 0.08 |
| ## UNEX076 | 0.10 | -0.03 | -0.04 | 0.01 | 0.07 | 0.03 | 0.01 | 0.03 | -0.01 | 0.02 |
| ## UNEX078 | 0.14 | 0.13 | 0.05 | 0.06 | 0.17 | 0.15 | 0.02 | 0.07 | 0.00 | 0.02 |
| ## UNEX079 | 0.28 | 0.06 | 0.03 | 0.00 | 0.06 | 0.09 | -0.03 | 0.00 | -0.09 | 0.03 |
| ## UNEX082 | 0.01 | 0.13 | -0.04 | 0.01 | 0.14 | 0.19 | 0.03 | 0.04 | 0.05 | 0.04 |
| ## UNEX083 | 0.10 | 0.13 | 0.00 | 0.03 | 0.14 | 0.09 | 0.02 | 0.04 | -0.01 | 0.05 |
| ## UNEX087 | 0.18 | 0.07 | 0.00 | 0.02 | 0.17 | 0.13 | 0.07 | 0.06 | 0.04 | 0.12 |
| ## UNEX089 | 0.10 | 0.11 | -0.02 | 0.05 | 0.11 | 0.12 | 0.05 | 0.01 | -0.02 | 0.07 |
| ## UNEX090 | 0.17 | 0.10 | -0.01 | 0.07 | 0.21 | 0.16 | 0.07 | 0.02 | 0.00 | 0.10 |
| ## UNEX093 | 0.11 | 0.08 | 0.02 | -0.02 | 0.05 | 0.06 | 0.05 | 0.03 | 0.00 | 0.05 |
| ## UNEX099 | 0.20 | 0.06 | -0.04 | 0.00 | 0.09 | 0.10 | 0.01 | 0.03 | -0.01 | 0.05 |
| ## UNEX103 | 1.00 | 0.00 | -0.04 | -0.03 | 0.04 | 0.05 | 0.03 | -0.04 | -0.03 | 0.06 |
| ## INT004 | 0.00 | 1.00 | 0.13 | 0.14 | 0.44 | 0.21 | 0.16 | 0.22 | 0.13 | 0.30 |
| ## INT009 | -0.04 | 0.13 | 1.00 | 0.08 | 0.15 | 0.12 | 0.14 | 0.15 | 0.08 | 0.09 |
| ## INT011 | -0.03 | 0.14 | 0.08 | 1.00 | 0.13 | 0.07 | 0.04 | 0.09 | 0.09 | 0.11 |
| ## INT014 | 0.04 | 0.44 | 0.15 | 0.13 | 1.00 | 0.27 | 0.21 | 0.27 | 0.12 | 0.29 |
| ## INT026 | 0.05 | 0.21 | 0.12 | 0.07 | 0.27 | 1.00 | 0.18 | 0.11 | 0.07 | 0.16 |
| ## INT034 | 0.03 | 0.16 | 0.14 | 0.04 | 0.21 | 0.18 | 1.00 | 0.11 | 0.11 | 0.16 |
| ## INT035 | -0.04 | 0.22 | 0.15 | 0.09 | 0.27 | 0.11 | 0.11 | 1.00 | 0.10 | 0.20 |
| ## INT039 | -0.03 | 0.13 | 0.08 | 0.09 | 0.12 | 0.07 | 0.11 | 0.10 | 1.00 | 0.12 |
| ## INT041 | 0.06 | 0.30 | 0.09 | 0.11 | 0.29 | 0.16 | 0.16 | 0.20 | 0.12 | 1.00 |
| ## INT042 | -0.08 | 0.02 | 0.05 | 0.07 | 0.06 | -0.05 | 0.03 | 0.05 | 0.15 | 0.09 |
| ## INT045 | 0.07 | 0.13 | 0.12 | 0.08 | 0.18 | 0.25 | 0.18 | 0.09 | 0.10 | 0.15 |
| ## INT046 | 0.02 | 0.09 | 0.03 | 0.07 | 0.12 | 0.11 | 0.10 | 0.09 | 0.06 | 0.08 |
| ## INT048 | 0.08 | 0.10 | 0.11 | 0.08 | 0.17 | 0.22 | 0.07 | 0.10 | 0.07 | 0.17 |
| ## INT049 | 0.07 | 0.23 | 0.12 | 0.09 | 0.30 | 0.27 | 0.16 | 0.21 | 0.16 | 0.22 |
| ## INT050 | 0.04 | 0.20 | 0.12 | 0.10 | 0.20 | 0.15 | 0.11 | 0.22 | 0.08 | 0.18 |
| ## INT059 | 0.08 | 0.26 | 0.12 | 0.09 | 0.27 | 0.12 | 0.14 | 0.17 | 0.08 | 0.20 |
| ## INT064 | 0.02 | 0.19 | 0.08 | 0.06 | 0.30 | 0.39 | 0.23 | 0.22 | 0.07 | 0.12 |
| ## INT067 | -0.02 | 0.13 | 0.08 | 0.11 | 0.09 | 0.05 | 0.10 | 0.08 | 0.40 | 0.12 |
| ## INT077 | -0.01 | 0.17 | 0.27 | 0.04 | 0.18 | 0.11 | 0.09 | 0.17 | 0.06 | 0.11 |
| ## INT080 | 0.07 | 0.28 | 0.03 | 0.08 | 0.30 | 0.26 | 0.22 | 0.19 | 0.11 | 0.23 |
| ## INT086 | -0.04 | 0.22 | 0.23 | 0.15 | 0.23 | 0.17 | 0.11 | 0.24 | 0.11 | 0.22 |

|  |  |  |  |  |  |  |  |  |  |  |  |
| --- | --- | --- | --- | --- | --- | --- | --- | --- | --- | --- | --- |
| ## | INT092 | -0.04 | 0.11 | 0.06 | 0.12 | 0.13 | 0.08 | 0.07 | 0.08 | 0.08 | 0.13 |
| ## | INT094 | -0.02 | 0.13 | 0.10 | 0.59 | 0.10 | 0.07 | 0.05 | 0.08 | 0.06 | 0.15 |
| ## | INT097 | -0.02 | 0.19 | 0.19 | 0.14 | 0.19 | 0.14 | 0.06 | 0.23 | 0.12 | 0.14 |
| ## | INT098 | 0.00 | 0.24 | 0.08 | 0.07 | 0.24 | 0.29 | 0.28 | 0.28 | 0.11 | 0.10 |
| ## | INT102 | 0.08 | 0.28 | 0.09 | 0.09 | 0.27 | 0.29 | 0.23 | 0.16 | 0.08 | 0.31 |
| ## | INT104 | -0.02 | 0.25 | 0.16 | 0.10 | 0.25 | 0.16 | 0.13 | 0.22 | 0.06 | 0.21 |
| ## |  | INT042 | INT045 | INT046 | INT048 | INT049 | INT050 | INT059 | INT064 | INT067 | INT077 |
| ## | COG001 | -0.05 | 0.05 | 0.03 | 0.20 | 0.15 | 0.18 | 0.11 | 0.12 | 0.09 | 0.13 |
| ## | COG008 | -0.03 | 0.07 | 0.08 | 0.17 | 0.20 | 0.09 | 0.21 | 0.14 | 0.08 | 0.04 |
| ## | COG016 | -0.07 | 0.03 | 0.01 | 0.21 | 0.12 | 0.07 | 0.13 | 0.09 | 0.08 | 0.07 |
| ## | COG021 | -0.01 | 0.06 | 0.05 | 0.22 | 0.05 | 0.10 | 0.09 | 0.09 | 0.06 | 0.07 |
| ## | COG022 | -0.05 | 0.11 | 0.06 | 0.23 | 0.12 | 0.04 | 0.07 | 0.12 | 0.06 | 0.04 |
| ## | COG023 | -0.08 | 0.03 | 0.06 | 0.18 | 0.04 | 0.06 | 0.10 | 0.08 | 0.04 | 0.06 |
| ## | COG028 | -0.10 | 0.08 | 0.07 | 0.21 | 0.16 | 0.15 | 0.17 | 0.16 | 0.02 | 0.08 |
| ## | COG032 | -0.10 | 0.04 | 0.07 | 0.18 | 0.09 | 0.06 | 0.14 | 0.05 | 0.04 | -0.01 |
| ## | COG036 | -0.10 | 0.01 | 0.04 | 0.17 | 0.09 | 0.08 | 0.15 | 0.09 | 0.06 | 0.04 |
| ## | COG037 | -0.02 | 0.12 | 0.12 | 0.27 | 0.17 | 0.12 | 0.19 | 0.21 | 0.07 | 0.06 |
| ## | COG038 | -0.02 | 0.14 | 0.11 | 0.17 | 0.22 | 0.16 | 0.17 | 0.14 | 0.09 | 0.14 |
| ## | COG043 | -0.04 | 0.09 | 0.12 | 0.18 | 0.14 | 0.06 | 0.13 | 0.18 | 0.05 | 0.00 |
| ## | COG054 | 0.00 | 0.11 | 0.07 | 0.23 | 0.21 | 0.10 | 0.25 | 0.20 | 0.07 | 0.06 |
| ## | COG055 | -0.04 | 0.07 | 0.00 | 0.15 | 0.04 | 0.07 | 0.08 | 0.07 | 0.01 | 0.09 |
| ## | COG061 | -0.08 | 0.03 | 0.05 | 0.16 | 0.12 | 0.03 | 0.12 | 0.13 | 0.06 | 0.01 |
| ## | COG062 | -0.09 | 0.09 | 0.11 | 0.20 | 0.12 | 0.11 | 0.14 | 0.09 | 0.00 | 0.03 |
| ## | COG066 | -0.08 | 0.00 | 0.06 | 0.14 | 0.15 | 0.10 | 0.12 | 0.08 | 0.08 | 0.09 |
| ## | COG073 | -0.01 | 0.07 | 0.04 | 0.20 | 0.15 | 0.19 | 0.18 | 0.16 | 0.02 | 0.07 |
| ## | COG075 | -0.02 | 0.10 | 0.07 | 0.29 | 0.24 | 0.15 | 0.27 | 0.19 | 0.06 | 0.10 |
| ## | COG081 | -0.08 | 0.09 | 0.09 | 0.24 | 0.15 | 0.17 | 0.19 | 0.16 | 0.03 | 0.08 |
| ## | COG085 | -0.03 | 0.03 | 0.05 | 0.16 | 0.01 | 0.07 | 0.08 | 0.11 | 0.04 | 0.06 |
| ## | COG091 | -0.03 | 0.05 | 0.03 | 0.14 | 0.12 | 0.07 | 0.14 | 0.09 | 0.02 | 0.09 |
| ## | COG095 | -0.08 | 0.10 | 0.01 | 0.20 | 0.08 | 0.15 | 0.13 | 0.17 | 0.02 | 0.05 |
| ## | COG101 | -0.03 | 0.10 | 0.11 | 0.19 | 0.09 | 0.06 | 0.10 | 0.14 | 0.05 | 0.07 |
| ## | IMP002 | -0.05 | 0.05 | 0.04 | 0.14 | 0.12 | 0.01 | 0.12 | 0.12 | 0.06 | 0.09 |
| ## | IMP007 | 0.03 | 0.00 | 0.04 | -0.10 | -0.07 | -0.08 | -0.02 | 0.00 | -0.01 | -0.09 |
| ## | IMP010 | 0.02 | 0.05 | 0.06 | 0.02 | 0.06 | -0.03 | 0.04 | 0.02 | 0.02 | 0.01 |
| ## | IMP012 | -0.03 | 0.13 | 0.05 | 0.25 | 0.19 | 0.12 | 0.18 | 0.19 | 0.03 | 0.12 |
| ## | IMP013 | 0.00 | 0.01 | -0.02 | -0.01 | 0.03 | -0.06 | 0.01 | -0.03 | 0.01 | 0.01 |
| ## | IMP015 | -0.07 | 0.09 | 0.07 | 0.22 | 0.18 | 0.10 | 0.14 | 0.22 | 0.04 | 0.08 |
| ## | IMP018 | 0.02 | -0.08 | -0.03 | -0.08 | -0.16 | -0.17 | -0.08 | -0.15 | -0.04 | -0.05 |
| ## | IMP019 | -0.05 | 0.06 | 0.02 | 0.14 | 0.10 | -0.03 | 0.16 | 0.10 | 0.03 | 0.06 |
| ## | IMP024 | 0.01 | 0.07 | 0.06 | 0.05 | 0.12 | 0.05 | 0.09 | 0.07 | 0.02 | 0.00 |
| ## | IMP031 | -0.07 | 0.00 | 0.03 | 0.01 | 0.06 | 0.04 | 0.05 | 0.03 | -0.01 | 0.02 |
| ## | IMP051 | 0.07 | 0.01 | 0.04 | -0.03 | 0.04 | 0.01 | -0.02 | 0.08 | 0.00 | 0.01 |
| ## | IMP052 | -0.03 | 0.07 | 0.06 | 0.06 | 0.04 | -0.02 | 0.03 | 0.08 | -0.01 | 0.03 |
| ## | IMP053 | -0.11 | 0.10 | 0.04 | 0.21 | 0.13 | 0.12 | 0.14 | 0.08 | 0.05 | 0.12 |
| ## | IMP056 | -0.11 | 0.02 | 0.01 | 0.19 | 0.13 | 0.12 | 0.15 | 0.05 | 0.02 | -0.01 |
| ## | IMP057 | 0.00 | -0.03 | -0.03 | -0.12 | -0.09 | -0.11 | -0.05 | -0.09 | -0.06 | -0.09 |
| ## | IMP068 | 0.05 | 0.09 | 0.07 | 0.01 | 0.02 | 0.00 | -0.04 | 0.05 | -0.05 | 0.02 |
| ## | IMP069 | -0.07 | 0.03 | 0.04 | 0.03 | 0.02 | 0.05 | 0.05 | 0.07 | 0.01 | 0.02 |
| ## | IMP070 | -0.01 | 0.10 | 0.03 | 0.05 | 0.08 | 0.06 | 0.13 | 0.12 | 0.04 | 0.03 |
| ## | IMP074 | -0.02 | 0.21 | 0.04 | 0.12 | 0.09 | 0.04 | 0.14 | 0.09 | 0.06 | 0.03 |
| ## | IMP084 | -0.03 | 0.10 | 0.05 | 0.07 | 0.02 | 0.05 | 0.04 | 0.06 | 0.04 | 0.09 |
| ## | IMP088 | -0.11 | 0.03 | -0.02 | 0.09 | 0.09 | 0.00 | 0.06 | 0.03 | 0.07 | 0.00 |
| ## | IMP096 | -0.03 | 0.13 | 0.06 | 0.24 | 0.13 | 0.12 | 0.15 | 0.23 | 0.06 | 0.10 |
| ## | IMP100 | -0.05 | 0.12 | 0.02 | 0.15 | 0.12 | 0.11 | 0.11 | 0.10 | 0.01 | 0.09 |

|  |  |  |  |  |  |  |  |  |  |  |
| --- | --- | --- | --- | --- | --- | --- | --- | --- | --- | --- |
| ## UNEX003 | 0.00 | 0.06 | 0.00 | 0.08 | 0.02 | 0.07 | 0.03 | 0.01 | 0.03 | 0.06 |
| ## UNEX005 | -0.10 | 0.14 | -0.04 | 0.15 | 0.10 | 0.07 | 0.10 | 0.07 | 0.02 | 0.02 |
| ## UNEX006 | -0.08 | 0.03 | 0.03 | 0.12 | 0.00 | -0.02 | 0.00 | 0.01 | 0.00 | 0.01 |
| ## UNEX017 | -0.08 | 0.08 | 0.07 | 0.21 | 0.16 | 0.07 | 0.15 | 0.14 | 0.03 | 0.02 |
| ## UNEX020 | -0.04 | 0.08 | 0.09 | 0.05 | 0.08 | 0.07 | 0.06 | 0.11 | -0.01 | 0.03 |
| ## UNEX025 | -0.09 | 0.08 | 0.00 | 0.14 | 0.14 | 0.03 | 0.13 | 0.13 | -0.02 | 0.02 |
| ## UNEX027 | -0.11 | 0.05 | 0.00 | 0.15 | 0.08 | 0.05 | 0.10 | 0.10 | -0.02 | 0.03 |
| ## UNEX029 | -0.03 | 0.05 | 0.00 | 0.08 | 0.01 | 0.02 | 0.05 | 0.13 | 0.01 | -0.05 |
| ## UNEX030 | -0.06 | 0.12 | 0.04 | 0.12 | 0.13 | 0.05 | 0.14 | 0.06 | 0.04 | -0.02 |
| ## UNEX033 | -0.01 | 0.05 | 0.02 | 0.19 | 0.08 | 0.09 | 0.19 | 0.13 | 0.09 | 0.00 |
| ## UNEX040 | 0.00 | 0.13 | -0.02 | 0.08 | 0.08 | 0.07 | 0.16 | 0.15 | -0.02 | 0.00 |
| ## UNEX044 | -0.05 | 0.12 | 0.00 | 0.08 | 0.15 | 0.03 | 0.18 | 0.14 | 0.01 | 0.01 |
| ## UNEX047 | -0.06 | 0.05 | 0.10 | 0.23 | 0.10 | 0.08 | 0.04 | 0.13 | 0.02 | 0.08 |
| ## UNEX058 | -0.07 | 0.12 | 0.01 | 0.05 | 0.04 | 0.03 | 0.05 | 0.09 | -0.01 | 0.07 |
| ## UNEX060 | -0.03 | 0.19 | 0.07 | 0.15 | 0.15 | 0.07 | 0.13 | 0.23 | 0.02 | 0.07 |
| ## UNEX063 | -0.10 | 0.06 | 0.02 | 0.14 | 0.09 | 0.09 | 0.08 | 0.08 | 0.04 | 0.04 |
| ## UNEX065 | -0.06 | 0.10 | 0.01 | 0.11 | 0.03 | 0.04 | 0.12 | 0.03 | 0.01 | -0.01 |
| ## UNEX071 | -0.06 | 0.01 | 0.06 | 0.13 | 0.06 | 0.04 | 0.10 | 0.11 | 0.06 | -0.01 |
| ## UNEX072 | -0.02 | 0.07 | -0.05 | 0.19 | 0.09 | 0.07 | 0.09 | 0.15 | 0.00 | 0.03 |
| ## UNEX076 | 0.01 | 0.07 | 0.04 | 0.09 | 0.01 | 0.01 | 0.00 | 0.11 | 0.01 | 0.03 |
| ## UNEX078 | -0.07 | 0.08 | 0.06 | 0.26 | 0.16 | 0.12 | 0.13 | 0.16 | 0.04 | 0.07 |
| ## UNEX079 | -0.10 | 0.10 | 0.07 | 0.05 | 0.02 | 0.02 | 0.04 | 0.02 | -0.06 | 0.01 |
| ## UNEX082 | -0.05 | 0.09 | -0.01 | 0.21 | 0.12 | 0.08 | 0.11 | 0.18 | 0.02 | 0.06 |
| ## UNEX083 | -0.08 | 0.08 | -0.03 | 0.19 | 0.09 | 0.06 | 0.16 | 0.08 | -0.02 | 0.03 |
| ## UNEX087 | -0.06 | 0.10 | 0.01 | 0.20 | 0.14 | 0.12 | 0.17 | 0.16 | 0.07 | 0.05 |
| ## UNEX089 | -0.07 | 0.06 | 0.03 | 0.20 | 0.06 | 0.02 | 0.14 | 0.13 | 0.04 | 0.02 |
| ## UNEX090 | -0.05 | 0.10 | 0.08 | 0.12 | 0.12 | 0.07 | 0.17 | 0.06 | 0.02 | 0.01 |
| ## UNEX093 | -0.06 | 0.04 | 0.02 | 0.13 | 0.03 | 0.07 | 0.01 | 0.04 | 0.00 | 0.07 |
| ## UNEX099 | -0.11 | 0.09 | -0.02 | 0.15 | 0.08 | 0.07 | 0.09 | 0.05 | -0.02 | 0.02 |
| ## UNEX103 | -0.08 | 0.07 | 0.02 | 0.08 | 0.07 | 0.04 | 0.08 | 0.02 | -0.02 | -0.01 |
| ## INT004 | 0.02 | 0.13 | 0.09 | 0.10 | 0.23 | 0.20 | 0.26 | 0.19 | 0.13 | 0.17 |
| ## INT009 | 0.05 | 0.12 | 0.03 | 0.11 | 0.12 | 0.12 | 0.12 | 0.08 | 0.08 | 0.27 |
| ## INT011 | 0.07 | 0.08 | 0.07 | 0.08 | 0.09 | 0.10 | 0.09 | 0.06 | 0.11 | 0.04 |
| ## INT014 | 0.06 | 0.18 | 0.12 | 0.17 | 0.30 | 0.20 | 0.27 | 0.30 | 0.09 | 0.18 |
| ## INT026 | -0.05 | 0.25 | 0.11 | 0.22 | 0.27 | 0.15 | 0.12 | 0.39 | 0.05 | 0.11 |
| ## INT034 | 0.03 | 0.18 | 0.10 | 0.07 | 0.16 | 0.11 | 0.14 | 0.23 | 0.10 | 0.09 |
| ## INT035 | 0.05 | 0.09 | 0.09 | 0.10 | 0.21 | 0.22 | 0.17 | 0.22 | 0.08 | 0.17 |
| ## INT039 | 0.15 | 0.10 | 0.06 | 0.07 | 0.16 | 0.08 | 0.08 | 0.07 | 0.40 | 0.06 |
| ## INT041 | 0.09 | 0.15 | 0.08 | 0.17 | 0.22 | 0.18 | 0.20 | 0.12 | 0.12 | 0.11 |
| ## INT042 | 1.00 | 0.06 | 0.00 | -0.02 | 0.08 | -0.03 | -0.01 | 0.00 | 0.10 | 0.08 |
| ## INT045 | 0.06 | 1.00 | 0.11 | 0.12 | 0.17 | 0.15 | 0.14 | 0.21 | 0.06 | 0.15 |
| ## INT046 | 0.00 | 0.11 | 1.00 | 0.03 | 0.10 | 0.11 | 0.03 | 0.14 | 0.05 | 0.05 |
| ## INT048 | -0.02 | 0.12 | 0.03 | 1.00 | 0.20 | 0.14 | 0.16 | 0.17 | 0.17 | 0.13 |
| ## INT049 | 0.08 | 0.17 | 0.10 | 0.20 | 1.00 | 0.22 | 0.12 | 0.33 | 0.12 | 0.21 |
| ## INT050 | -0.03 | 0.15 | 0.11 | 0.14 | 0.22 | 1.00 | 0.12 | 0.18 | 0.06 | 0.14 |
| ## INT059 | -0.01 | 0.14 | 0.03 | 0.16 | 0.12 | 0.12 | 1.00 | 0.15 | 0.12 | 0.04 |
| ## INT064 | 0.00 | 0.21 | 0.14 | 0.17 | 0.33 | 0.18 | 0.15 | 1.00 | 0.12 | 0.12 |
| ## INT067 | 0.10 | 0.06 | 0.05 | 0.17 | 0.12 | 0.06 | 0.12 | 0.12 | 1.00 | 0.07 |
| ## INT077 | 0.08 | 0.15 | 0.05 | 0.13 | 0.21 | 0.14 | 0.04 | 0.12 | 0.07 | 1.00 |
| ## INT080 | 0.00 | 0.23 | 0.07 | 0.18 | 0.23 | 0.17 | 0.20 | 0.32 | 0.08 | 0.06 |
| ## INT086 | 0.04 | 0.09 | 0.08 | 0.19 | 0.19 | 0.24 | 0.14 | 0.20 | 0.16 | 0.16 |
| ## INT092 | 0.11 | 0.03 | 0.06 | 0.03 | 0.10 | 0.04 | 0.03 | 0.06 | 0.08 | 0.01 |
| ## INT094 | 0.00 | 0.03 | 0.05 | 0.10 | 0.13 | 0.10 | 0.13 | 0.09 | 0.10 | 0.03 |
| ## INT097 | 0.04 | 0.11 | 0.11 | 0.16 | 0.19 | 0.35 | 0.09 | 0.11 | 0.08 | 0.21 |

|  |  |  |  |  |  |  |  |  |  |  |  |
| --- | --- | --- | --- | --- | --- | --- | --- | --- | --- | --- | --- |
| ## | INT098 | 0.03 | 0.21 | 0.19 | 0.21 | 0.26 | 0.26 | 0.09 | 0.36 | 0.07 | 0.13 |
| ## | INT102 | 0.00 | 0.26 | 0.10 | 0.17 | 0.24 | 0.21 | 0.22 | 0.22 | 0.10 | 0.10 |
| ## | INT104 | 0.00 | 0.13 | 0.11 | 0.20 | 0.22 | 0.28 | 0.14 | 0.14 | 0.02 | 0.15 |
| ## | INT080 | INT086 | INT092 | INT094 | INT097 | INT098 | INT102 | INT104 |  |  |  |
| ## | COG001 | 0.12 | 0.22 | 0.00 | 0.07 | 0.21 | 0.11 | 0.14 | 0.17 |  |  |
| ## | COG008 | 0.12 | 0.07 | 0.00 | 0.11 | 0.09 | 0.10 | 0.13 | 0.09 |  |  |
| ## | COG016 | 0.10 | 0.12 | -0.07 | 0.08 | 0.01 | 0.10 | 0.08 | 0.08 |  |  |
| ## | COG021 | 0.13 | 0.17 | 0.05 | 0.04 | 0.18 | 0.08 | 0.12 | 0.13 |  |  |
| ## | COG022 | 0.21 | 0.04 | 0.01 | 0.05 | 0.05 | 0.08 | 0.14 | 0.07 |  |  |
| ## | COG023 | 0.09 | 0.13 | -0.04 | 0.08 | 0.03 | 0.06 | 0.06 | 0.14 |  |  |
| ## | COG028 | 0.18 | 0.14 | 0.04 | 0.06 | 0.08 | 0.12 | 0.13 | 0.14 |  |  |
| ## | COG032 | 0.14 | 0.04 | 0.00 | 0.03 | 0.04 | 0.09 | 0.14 | 0.10 |  |  |
| ## | COG036 | 0.08 | 0.11 | -0.02 | 0.11 | 0.07 | 0.07 | 0.09 | 0.06 |  |  |
| ## | COG037 | 0.24 | 0.12 | 0.03 | 0.11 | 0.10 | 0.18 | 0.18 | 0.09 |  |  |
| ## | COG038 | 0.15 | 0.24 | -0.04 | 0.08 | 0.21 | 0.07 | 0.14 | 0.20 |  |  |
| ## | COG043 | 0.16 | 0.09 | 0.00 | 0.03 | 0.05 | 0.12 | 0.19 | 0.09 |  |  |
| ## | COG054 | 0.19 | 0.13 | 0.01 | 0.05 | 0.10 | 0.13 | 0.20 | 0.15 |  |  |
| ## | COG055 | 0.10 | 0.03 | -0.01 | 0.02 | 0.01 | 0.01 | 0.10 | 0.06 |  |  |
| ## | COG061 | 0.15 | 0.08 | -0.01 | 0.05 | 0.02 | 0.08 | 0.14 | 0.03 |  |  |
| ## | COG062 | 0.13 | 0.14 | 0.00 | 0.08 | 0.14 | 0.09 | 0.13 | 0.14 |  |  |
| ## | COG066 | 0.05 | 0.16 | 0.01 | 0.05 | 0.08 | 0.08 | 0.06 | 0.12 |  |  |
| ## | COG073 | 0.19 | 0.14 | 0.04 | 0.10 | 0.09 | 0.19 | 0.15 | 0.08 |  |  |
| ## | COG075 | 0.30 | 0.18 | 0.05 | 0.08 | 0.09 | 0.20 | 0.26 | 0.13 |  |  |
| ## | COG081 | 0.20 | 0.10 | 0.01 | 0.08 | 0.08 | 0.13 | 0.15 | 0.18 |  |  |
| ## | COG085 | 0.12 | 0.08 | -0.03 | 0.06 | 0.07 | 0.08 | 0.09 | 0.12 |  |  |
| ## | COG091 | 0.12 | 0.13 | 0.03 | 0.05 | 0.05 | 0.05 | 0.12 | 0.05 |  |  |
| ## | COG095 | 0.24 | 0.11 | -0.03 | 0.06 | 0.08 | 0.11 | 0.19 | 0.06 |  |  |
| ## | COG101 | 0.17 | 0.05 | -0.01 | 0.11 | 0.07 | 0.09 | 0.13 | 0.08 |  |  |
| ## | IMP002 | 0.14 | 0.03 | -0.01 | 0.03 | -0.03 | 0.11 | 0.08 | 0.06 |  |  |
| ## | IMP007 | 0.04 | -0.08 | 0.01 | -0.05 | -0.12 | 0.03 | -0.01 | -0.08 |  |  |
| ## | IMP010 | 0.02 | -0.03 | -0.09 | -0.02 | -0.05 | 0.01 | -0.03 | -0.05 |  |  |
| ## | IMP012 | 0.24 | 0.11 | -0.01 | 0.07 | 0.10 | 0.19 | 0.26 | 0.10 |  |  |
| ## | IMP013 | -0.02 | -0.06 | 0.01 | 0.06 | -0.05 | -0.05 | -0.04 | 0.01 |  |  |
| ## | IMP015 | 0.19 | 0.10 | 0.01 | 0.04 | 0.06 | 0.21 | 0.18 | 0.10 |  |  |
| ## | IMP018 | -0.11 | -0.13 | -0.08 | -0.06 | -0.13 | -0.17 | -0.09 | -0.13 |  |  |
| ## | IMP019 | 0.11 | -0.03 | 0.00 | 0.05 | -0.04 | 0.08 | 0.11 | 0.03 |  |  |
| ## | IMP024 | 0.11 | 0.06 | 0.04 | 0.07 | 0.01 | 0.12 | 0.12 | 0.09 |  |  |
| ## | IMP031 | 0.02 | 0.00 | 0.02 | 0.04 | -0.01 | 0.02 | 0.02 | 0.05 |  |  |
| ## | IMP051 | 0.08 | -0.01 | 0.03 | -0.04 | -0.03 | 0.13 | 0.07 | -0.02 |  |  |
| ## | IMP052 | 0.07 | -0.03 | 0.01 | -0.02 | -0.01 | 0.05 | 0.04 | 0.00 |  |  |
| ## | IMP053 | 0.11 | 0.08 | -0.02 | 0.11 | 0.05 | 0.10 | 0.13 | 0.11 |  |  |
| ## | IMP056 | 0.15 | 0.13 | -0.01 | 0.10 | 0.08 | 0.12 | 0.12 | 0.09 |  |  |
| ## | IMP057 | -0.06 | -0.17 | -0.04 | -0.05 | -0.13 | -0.08 | -0.09 | -0.12 |  |  |
| ## | IMP068 | 0.08 | 0.01 | 0.03 | -0.05 | -0.04 | 0.08 | 0.07 | 0.01 |  |  |
| ## | IMP069 | 0.08 | -0.01 | -0.01 | 0.03 | -0.04 | 0.06 | 0.01 | 0.01 |  |  |
| ## | IMP070 | 0.11 | 0.09 | 0.06 | 0.02 | 0.04 | 0.13 | 0.18 | 0.09 |  |  |
| ## | IMP074 | 0.15 | 0.11 | 0.04 | 0.06 | 0.04 | 0.11 | 0.25 | 0.05 |  |  |
| ## | IMP084 | 0.12 | -0.01 | -0.07 | 0.06 | -0.01 | 0.05 | 0.09 | 0.07 |  |  |
| ## | IMP088 | 0.03 | -0.04 | 0.01 | 0.08 | -0.01 | -0.02 | 0.02 | 0.00 |  |  |
| ## | IMP096 | 0.25 | 0.10 | -0.02 | 0.14 | 0.07 | 0.16 | 0.21 | 0.16 |  |  |
| ## | IMP100 | 0.18 | 0.07 | -0.03 | 0.05 | 0.04 | 0.11 | 0.16 | 0.14 |  |  |
| ## | UNEX003 | 0.04 | 0.00 | -0.08 | 0.02 | 0.00 | 0.02 | 0.09 | -0.02 |  |  |
| ## | UNEX005 | 0.11 | 0.08 | -0.05 | 0.03 | -0.01 | 0.02 | 0.12 | 0.07 |  |  |
| ## | UNEX006 | 0.02 | 0.03 | -0.07 | 0.03 | -0.07 | -0.01 | 0.06 | -0.02 |  |  |

|  |  |  |  |  |  |  |  |  |
| --- | --- | --- | --- | --- | --- | --- | --- | --- |
| ## UNEX017 | 0.12 | 0.05 | -0.04 | 0.07 | 0.03 | 0.08 | 0.18 | 0.11 |
| ## UNEX020 | 0.11 | 0.02 | 0.00 | 0.05 | 0.03 | 0.07 | 0.11 | 0.01 |
| ## UNEX025 | 0.13 | 0.02 | 0.00 | 0.06 | 0.01 | 0.08 | 0.13 | 0.03 |
| ## UNEX027 | 0.12 | 0.07 | -0.01 | 0.05 | 0.05 | 0.04 | 0.12 | 0.04 |
| ## UNEX029 | 0.08 | -0.02 | 0.01 | 0.01 | -0.05 | 0.01 | 0.06 | 0.00 |
| ## UNEX030 | 0.18 | 0.08 | -0.05 | 0.02 | 0.01 | 0.05 | 0.19 | 0.07 |
| ## UNEX033 | 0.18 | 0.16 | 0.04 | 0.05 | 0.07 | 0.11 | 0.10 | 0.08 |
| ## UNEX040 | 0.19 | 0.04 | -0.03 | 0.08 | 0.02 | 0.12 | 0.19 | 0.06 |
| ## UNEX044 | 0.15 | 0.04 | 0.01 | 0.05 | 0.01 | 0.05 | 0.20 | 0.07 |
| ## UNEX047 | 0.09 | 0.11 | 0.00 | 0.04 | 0.12 | 0.08 | 0.12 | 0.17 |
| ## UNEX058 | 0.10 | 0.04 | -0.04 | 0.05 | 0.05 | 0.04 | 0.12 | 0.03 |
| ## UNEX060 | 0.19 | 0.06 | 0.01 | 0.05 | 0.02 | 0.16 | 0.17 | 0.05 |
| ## UNEX063 | 0.16 | 0.06 | -0.03 | 0.06 | 0.07 | 0.08 | 0.07 | -0.01 |
| ## UNEX065 | 0.11 | 0.00 | -0.01 | 0.06 | 0.06 | 0.04 | 0.11 | 0.05 |
| ## UNEX071 | 0.08 | 0.06 | -0.01 | 0.07 | 0.07 | 0.05 | 0.07 | 0.07 |
| ## UNEX072 | 0.19 | 0.10 | 0.00 | 0.02 | 0.03 | 0.10 | 0.09 | 0.05 |
| ## UNEX076 | 0.06 | 0.02 | -0.02 | 0.00 | -0.02 | 0.03 | 0.01 | 0.02 |
| ## UNEX078 | 0.18 | 0.10 | 0.01 | 0.10 | 0.06 | 0.08 | 0.15 | 0.14 |
| ## UNEX079 | 0.11 | -0.02 | -0.09 | 0.00 | -0.01 | 0.04 | 0.06 | 0.03 |
| ## UNEX082 | 0.12 | 0.08 | 0.00 | 0.02 | 0.02 | 0.12 | 0.12 | 0.10 |
| ## UNEX083 | 0.15 | 0.06 | -0.01 | 0.05 | 0.05 | 0.07 | 0.15 | 0.10 |
| ## UNEX087 | 0.18 | 0.11 | -0.01 | 0.09 | 0.05 | 0.09 | 0.16 | 0.06 |
| ## UNEX089 | 0.12 | 0.04 | -0.01 | 0.10 | 0.07 | 0.04 | 0.14 | 0.08 |
| ## UNEX090 | 0.19 | 0.02 | 0.03 | 0.07 | 0.06 | 0.08 | 0.17 | 0.14 |
| ## UNEX093 | 0.06 | 0.00 | -0.03 | 0.00 | 0.08 | 0.03 | 0.10 | 0.07 |
| ## UNEX099 | 0.16 | 0.03 | -0.05 | 0.01 | 0.00 | 0.03 | 0.08 | 0.01 |
| ## UNEX103 | 0.07 | -0.04 | -0.04 | -0.02 | -0.02 | 0.00 | 0.08 | -0.02 |
| ## INT004 | 0.28 | 0.22 | 0.11 | 0.13 | 0.19 | 0.24 | 0.28 | 0.25 |
| ## INT009 | 0.03 | 0.23 | 0.06 | 0.10 | 0.19 | 0.08 | 0.09 | 0.16 |
| ## INT011 | 0.08 | 0.15 | 0.12 | 0.59 | 0.14 | 0.07 | 0.09 | 0.10 |
| ## INT014 | 0.30 | 0.23 | 0.13 | 0.10 | 0.19 | 0.24 | 0.27 | 0.25 |
| ## INT026 | 0.26 | 0.17 | 0.08 | 0.07 | 0.14 | 0.29 | 0.29 | 0.16 |
| ## INT034 | 0.22 | 0.11 | 0.07 | 0.05 | 0.06 | 0.28 | 0.23 | 0.13 |
| ## INT035 | 0.19 | 0.24 | 0.08 | 0.08 | 0.23 | 0.28 | 0.16 | 0.22 |
| ## INT039 | 0.11 | 0.11 | 0.08 | 0.06 | 0.12 | 0.11 | 0.08 | 0.06 |
| ## INT041 | 0.23 | 0.22 | 0.13 | 0.15 | 0.14 | 0.10 | 0.31 | 0.21 |
| ## INT042 | 0.00 | 0.04 | 0.11 | 0.00 | 0.04 | 0.03 | 0.00 | 0.00 |
| ## INT045 | 0.23 | 0.09 | 0.03 | 0.03 | 0.11 | 0.21 | 0.26 | 0.13 |
| ## INT046 | 0.07 | 0.08 | 0.06 | 0.05 | 0.11 | 0.19 | 0.10 | 0.11 |
| ## INT048 | 0.18 | 0.19 | 0.03 | 0.10 | 0.16 | 0.21 | 0.17 | 0.20 |
| ## INT049 | 0.23 | 0.19 | 0.10 | 0.13 | 0.19 | 0.26 | 0.24 | 0.22 |
| ## INT050 | 0.17 | 0.24 | 0.04 | 0.10 | 0.35 | 0.26 | 0.21 | 0.28 |
| ## INT059 | 0.20 | 0.14 | 0.03 | 0.13 | 0.09 | 0.09 | 0.22 | 0.14 |
| ## INT064 | 0.32 | 0.20 | 0.06 | 0.09 | 0.11 | 0.36 | 0.22 | 0.14 |
| ## INT067 | 0.08 | 0.16 | 0.08 | 0.10 | 0.08 | 0.07 | 0.10 | 0.02 |
| ## INT077 | 0.06 | 0.16 | 0.01 | 0.03 | 0.21 | 0.13 | 0.10 | 0.15 |
| ## INT080 | 1.00 | 0.19 | 0.06 | 0.10 | 0.08 | 0.32 | 0.35 | 0.16 |
| ## INT086 | 0.19 | 1.00 | 0.10 | 0.14 | 0.36 | 0.21 | 0.18 | 0.26 |
| ## INT092 | 0.06 | 0.10 | 1.00 | 0.04 | 0.12 | 0.03 | 0.09 | 0.06 |
| ## INT094 | 0.10 | 0.14 | 0.04 | 1.00 | 0.11 | 0.06 | 0.09 | 0.09 |
| ## INT097 | 0.08 | 0.36 | 0.12 | 0.11 | 1.00 | 0.22 | 0.20 | 0.36 |
| ## INT098 | 0.32 | 0.21 | 0.03 | 0.06 | 0.22 | 1.00 | 0.28 | 0.18 |
| ## INT102 | 0.35 | 0.18 | 0.09 | 0.09 | 0.20 | 0.28 | 1.00 | 0.22 |
| ## INT104 | 0.16 | 0.26 | 0.06 | 0.09 | 0.36 | 0.18 | 0.22 | 1.00 |

|  |  |  |  |  |  |  |  |  |  |  |  |
| --- | --- | --- | --- | --- | --- | --- | --- | --- | --- | --- | --- |
| ## | COG001 | COG008 | COG016 | COG021 | COG022 | COG023 | COG028 | COG032 | COG036 | COG037 |  |
| ## | COG001 | 1.00 | 0.23 | 0.28 | 0.23 | 0.12 | 0.21 | 0.28 | 0.18 | 0.22 | 0.16 |
| ## | COG008 | 0.23 | 1.00 | 0.24 | 0.20 | 0.31 | 0.17 | 0.26 | 0.21 | 0.24 | 0.32 |
| ## | COG016 | 0.28 | 0.24 | 1.00 | 0.16 | 0.22 | 0.26 | 0.32 | 0.24 | 0.26 | 0.17 |
| ## | COG021 | 0.23 | 0.20 | 0.16 | 1.00 | 0.21 | 0.18 | 0.23 | 0.18 | 0.19 | 0.24 |
| ## | COG022 | 0.12 | 0.31 | 0.22 | 0.21 | 1.00 | 0.14 | 0.26 | 0.24 | 0.25 | 0.32 |
| ## | COG023 | 0.21 | 0.17 | 0.26 | 0.18 | 0.14 | 1.00 | 0.49 | 0.27 | 0.22 | 0.20 |
| ## | COG038 | COG043 | COG054 | COG055 | COG061 | COG062 | COG066 | COG073 | COG075 | COG081 |  |
| ## | COG001 | 0.32 | 0.16 | 0.13 | 0.19 | 0.19 | 0.24 | 0.29 | 0.21 | 0.19 | 0.19 |
| ## | COG008 | 0.22 | 0.23 | 0.28 | 0.14 | 0.30 | 0.25 | 0.17 | 0.26 | 0.26 | 0.29 |
| ## | COG016 | 0.21 | 0.22 | 0.21 | 0.34 | 0.20 | 0.24 | 0.43 | 0.28 | 0.23 | 0.28 |
| ## | COG021 | 0.19 | 0.22 | 0.18 | 0.11 | 0.21 | 0.34 | 0.07 | 0.11 | 0.15 | 0.19 |
| ## | COG022 | 0.16 | 0.36 | 0.25 | 0.20 | 0.41 | 0.33 | 0.10 | 0.20 | 0.30 | 0.30 |
| ## | COG023 | 0.22 | 0.18 | 0.18 | 0.23 | 0.19 | 0.25 | 0.28 | 0.22 | 0.16 | 0.23 |
| ## | COG085 | COG091 | COG095 | COG101 | IMP002 | IMP007 | IMP010 | IMP012 | IMP013 | IMP015 |  |
| ## | COG001 | 0.20 | 0.23 | 0.19 | 0.18 | 0.12 | -0.29 | 0.02 | 0.17 | 0.02 | 0.09 |
| ## | COG008 | 0.15 | 0.27 | 0.16 | 0.28 | 0.18 | -0.09 | 0.18 | 0.16 | 0.04 | 0.17 |
| ## | COG016 | 0.21 | 0.32 | 0.17 | 0.21 | 0.22 | -0.09 | 0.09 | 0.25 | 0.00 | 0.15 |
| ## | COG021 | 0.15 | 0.17 | 0.14 | 0.28 | 0.06 | -0.14 | 0.07 | 0.14 | -0.02 | 0.11 |
| ## | COG022 | 0.20 | 0.16 | 0.30 | 0.32 | 0.22 | 0.03 | 0.16 | 0.31 | 0.03 | 0.22 |
| ## | COG023 | 0.24 | 0.28 | 0.13 | 0.23 | 0.19 | -0.07 | 0.03 | 0.12 | 0.01 | 0.18 |
| ## | IMP018 | IMP019 | IMP024 | IMP031 | IMP051 | IMP052 | IMP053 | IMP056 | IMP057 | IMP068 |  |
| ## | COG001 | -0.10 | 0.10 | 0.01 | 0.05 | -0.05 | -0.03 | 0.13 | 0.11 | -0.27 | -0.02 |
| ## | COG008 | -0.10 | 0.15 | 0.06 | 0.08 | -0.04 | 0.05 | 0.19 | 0.16 | -0.06 | 0.01 |
| ## | COG016 | -0.10 | 0.09 | 0.06 | 0.10 | -0.04 | -0.03 | 0.21 | 0.20 | -0.11 | -0.06 |
| ## | COG021 | -0.08 | 0.13 | -0.02 | 0.04 | -0.01 | 0.06 | 0.13 | 0.12 | -0.10 | 0.04 |
| ## | COG022 | -0.06 | 0.20 | 0.13 | 0.07 | -0.02 | 0.15 | 0.21 | 0.19 | -0.03 | 0.08 |
| ## | COG023 | -0.04 | 0.07 | 0.10 | 0.09 | -0.11 | -0.02 | 0.20 | 0.18 | -0.14 | -0.01 |
| ## | IMP069 | IMP070 | IMP074 | IMP084 | IMP088 | IMP096 | IMP100 | UNEX003 | UNEX005 | UNEX006 |  |
| ## | COG001 | 0.00 | 0.04 | 0.08 | 0.01 | 0.00 | 0.14 | 0.11 | 0.04 | 0.09 | 0.07 |
| ## | COG008 | 0.04 | 0.11 | 0.12 | 0.04 | 0.10 | 0.24 | 0.18 | 0.01 | 0.09 | 0.04 |
| ## | COG016 | 0.06 | 0.08 | 0.11 | 0.03 | 0.10 | 0.20 | 0.18 | 0.07 | 0.18 | 0.14 |
| ## | COG021 | 0.02 | 0.07 | 0.07 | 0.03 | 0.10 | 0.12 | 0.11 | 0.06 | 0.11 | 0.04 |
| ## | COG022 | 0.05 | 0.11 | 0.16 | 0.09 | 0.12 | 0.27 | 0.28 | 0.09 | 0.23 | 0.22 |
| ## | COG023 | 0.10 | 0.08 | 0.11 | 0.08 | 0.05 | 0.16 | 0.15 | 0.06 | 0.10 | 0.07 |
| ## | UNEX017 | UNEX020 | UNEX025 | UNEX027 | UNEX029 | UNEX030 | UNEX033 | UNEX040 | UNEX044 |  |  |
| ## | COG001 | 0.15 | 0.04 | 0.06 | 0.10 | 0.08 | 0.10 | 0.10 | 0.04 | 0.05 |  |
| ## | COG008 | 0.18 | 0.04 | 0.13 | 0.17 | -0.02 | 0.15 | 0.15 | 0.05 | 0.12 |  |
| ## | COG016 | 0.17 | 0.03 | 0.08 | 0.22 | 0.10 | 0.22 | 0.11 | 0.10 | 0.12 |  |
| ## | COG021 | 0.15 | 0.04 | 0.08 | 0.16 | 0.02 | 0.08 | 0.12 | 0.07 | 0.09 |  |
| ## | COG022 | 0.34 | 0.16 | 0.20 | 0.29 | 0.16 | 0.32 | 0.11 | 0.20 | 0.19 |  |
| ## | COG023 | 0.18 | 0.06 | 0.09 | 0.13 | 0.08 | 0.13 | 0.10 | 0.06 | 0.07 |  |
| ## | UNEX047 | UNEX058 | UNEX060 | UNEX063 | UNEX065 | UNEX071 | UNEX072 | UNEX076 | UNEX078 |  |  |
| ## | COG001 | 0.17 | 0.12 | 0.02 | 0.11 | 0.04 | 0.07 | 0.05 | 0.02 | 0.12 |  |
| ## | COG008 | 0.16 | 0.07 | 0.10 | 0.11 | 0.09 | 0.09 | 0.10 | 0.07 | 0.16 |  |
| ## | COG016 | 0.16 | 0.13 | 0.11 | 0.15 | 0.12 | 0.08 | 0.10 | 0.03 | 0.17 |  |
| ## | COG021 | 0.27 | 0.11 | 0.08 | 0.10 | 0.05 | 0.11 | 0.09 | 0.04 | 0.20 |  |
| ## | COG022 | 0.23 | 0.15 | 0.19 | 0.21 | 0.15 | 0.13 | 0.21 | 0.09 | 0.24 |  |
| ## | COG023 | 0.17 | 0.12 | 0.09 | 0.11 | 0.05 | 0.07 | 0.11 | 0.06 | 0.19 |  |
| ## | UNEX079 | UNEX082 | UNEX083 | UNEX087 | UNEX089 | UNEX090 | UNEX093 | UNEX099 | UNEX103 |  |  |
| ## | COG001 | 0.10 | 0.06 | 0.14 | 0.13 | 0.10 | 0.07 | 0.05 | 0.07 | 0.02 |  |
| ## | COG008 | 0.07 | 0.12 | 0.18 | 0.19 | 0.15 | 0.18 | 0.05 | 0.13 | 0.02 |  |
| ## | COG016 | 0.13 | 0.09 | 0.17 | 0.12 | 0.14 | 0.11 | 0.05 | 0.20 | 0.02 |  |
| ## | COG021 | 0.02 | 0.09 | 0.17 | 0.16 | 0.13 | 0.06 | 0.18 | 0.01 | -0.02 |  |

```
## COG022    0.11    0.20    0.28    0.29    0.19    0.20    0.16    0.24    0.06
## COG023    0.09    0.09    0.22    0.13    0.18    0.07    0.10    0.12    0.03
##          INTO04 INTO09 INTO11 INTO14 INTO26 INTO34 INTO35 INTO39 INTO41 INTO42
## COG001    0.12    0.11    0.08    0.15    0.08    0.05    0.19    0.01    0.07   -0.05
## COG008    0.14    0.04    0.07    0.20    0.10    0.05    0.14    0.05    0.06   -0.03
## COG016    0.10    0.06    0.04    0.15    0.10    0.06    0.13    0.00    0.06   -0.07
## COG021    0.09    0.07    0.02    0.09    0.15    0.05    0.15   -0.03    0.03   -0.01
## COG022    0.08    0.03    0.04    0.15    0.17   -0.01    0.03    0.05    0.09   -0.05
## COG023    0.10    0.04    0.04    0.12    0.11   -0.04    0.10    0.01    0.02   -0.08
##          INTO45 INTO46 INTO48 INTO49 INTO50 INTO59 INTO64 INTO67 INTO77 INTO80
## COG001    0.05    0.03    0.20    0.15    0.18    0.11    0.12    0.09    0.13    0.12
## COG008    0.07    0.08    0.17    0.20    0.09    0.21    0.14    0.08    0.04    0.12
## COG016    0.03    0.01    0.21    0.12    0.07    0.13    0.09    0.08    0.07    0.10
## COG021    0.06    0.05    0.22    0.05    0.10    0.09    0.09    0.06    0.07    0.13
## COG022    0.11    0.06    0.23    0.12    0.04    0.07    0.12    0.06    0.04    0.21
## COG023    0.03    0.06    0.18    0.04    0.06    0.10    0.08    0.04    0.06    0.09
##          INTO86 INTO92 INTO94 INTO97 INTO98 INT102 INT104
## COG001    0.22    0.00    0.07    0.21    0.11    0.14    0.17
## COG008    0.07    0.00    0.11    0.09    0.10    0.13    0.09
## COG016    0.12   -0.07    0.08    0.01    0.10    0.08    0.08
## COG021    0.17    0.05    0.04    0.18    0.08    0.12    0.13
## COG022    0.04    0.01    0.05    0.05    0.08    0.14    0.07
## COG023    0.13   -0.04    0.08    0.03    0.06    0.06    0.14
```

```
# Check if the 2 matrices are statistically different: Jennrich test for
# equality of correlation matrices.
```

```
nOLIFE <- 104
```

```
corrOLIFE_result <- cortest.jennrich(mat.OLIFE, phi_matrix_OLIFE, nOLIFE,
                                     nOLIFE)
```

```
print(corrOLIFE_result)
```

```
## $chi2
## [1] 6.462794
##
## $prob
## [1] 1
```

```
# The two matrices are not statistically different
```

```
# Between the 2 is better to use the one with more decimal digits --> spearman
# Spearman will be used for in any other situation where a correlation matrix
# is required
```

#### 2. EGA Object

```
ega_OLIFE <- ega.object(mat.OLIFE, OLIFE)
dim.OLIFE <- ega_OLIFE$dim.variables
# 13 DIMENSIONS found with this method
oneOLIFE <- which(dim.OLIFE$dimension == 1)
dim.OLIFE$dimension[oneOLIFE] <- rep("Factor 01", length(oneOLIFE))
twoOLIFE <- which(dim.OLIFE$dimension == 2)
dim.OLIFE$dimension[twoOLIFE] <- rep("Factor 02", length(twoOLIFE))
threeOLIFE <- which(dim.OLIFE$dimension == 3)
dim.OLIFE$dimension[threeOLIFE] <- rep("Factor 03", length(threeOLIFE))
fourOLIFE <- which(dim.OLIFE$dimension == 4)
dim.OLIFE$dimension[fourOLIFE] <- rep("Factor 04", length(fourOLIFE))
fiveOLIFE <- which(dim.OLIFE$dimension == 5)
dim.OLIFE$dimension[fiveOLIFE] <- rep("Factor 05", length(fiveOLIFE))
sixOLIFE <- which(dim.OLIFE$dimension == 6)
dim.OLIFE$dimension[sixOLIFE] <- rep("Factor 06", length(sixOLIFE))
sevenOLIFE <- which(dim.OLIFE$dimension == 7)
dim.OLIFE$dimension[sevenOLIFE] <- rep("Factor 07", length(sevenOLIFE))
eightOLIFE <- which(dim.OLIFE$dimension == 8)
dim.OLIFE$dimension[eightOLIFE] <- rep("Factor 08", length(eightOLIFE))
nineOLIFE <- which(dim.OLIFE$dimension == 9)
dim.OLIFE$dimension[nineOLIFE] <- rep("Factor 09", length(nineOLIFE))
tenOLIFE <- which(dim.OLIFE$dimension == 10)
dim.OLIFE$dimension[tenOLIFE] <- rep("Factor 10", length(tenOLIFE))
elevenOLIFE <- which(dim.OLIFE$dimension == 11)
dim.OLIFE$dimension[elevenOLIFE] <- rep("Factor 11", length(elevenOLIFE))
twelveOLIFE <- which(dim.OLIFE$dimension == 12)
dim.OLIFE$dimension[twelveOLIFE] <- rep("Factor 12", length(twelveOLIFE))
thirteenOLIFE <- which(dim.OLIFE$dimension == 13)
dim.OLIFE$dimension[thirteenOLIFE] <- rep("Factor 13", length(thirteenOLIFE))
```

##### 3. REPRESENTATION

```
graph_OLIFE <- ega_OLIFE$glasso[match(dim.OLIFE$items, colnames(OLIFE)),
                                match(dim.OLIFE$items, colnames(OLIFE))]

A_OLIFE <- qgraph(graph_OLIFE, groups = dim.OLIFE$dimension,
                  label.prop = 0.8 , color = c("#f1605d", "#fed799", "#feb078",
                                                "#fc8961", "#fcfdbf", "#d8456c",
                                                "#b73779", "#932b80", "#721f81",
                                                "#51127c", "#2c115f", "#100b2d",
                                                "#000004"),
                  layout = "spring", vsize = 4,
                  title = "EGA model of the 0-LIFE with 13 factors")
```

#### EGA model of the O-LIFE with 13 factors

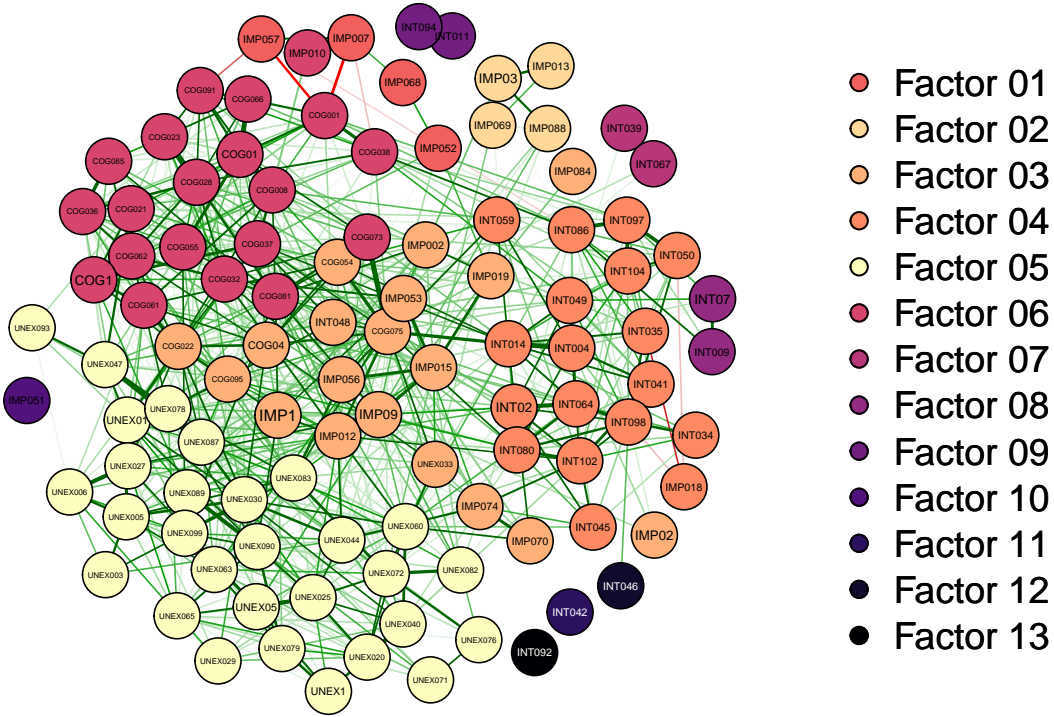

### Just use the 4 factors as are in the literature and see how all the items
### are spread or grouped

```
groups_dimensios_OLIFEega <- c(rep("Impulsive Nonconformity",8),
                                rep("Cognitive disorganisation",5),
                                rep("Impulsive Nonconformity",12),
                                rep("Unusual Experiences", 1),
                                rep("Introvertive Anhedonia",1),
                                rep("Impulsive Nonconformity",1),
                                rep("Introvertive Anhedonia", 17),
                                rep("Unusual Experiences",29),
                                rep("Cognitive disorganisation",19),
                                rep("Impulsive Nonconformity",1),
                                rep("Introvertive Anhedonia", 6),
                                rep("Impulsive Nonconformity",1),
                                rep("Introvertive Anhedonia", 3))

A_OLIFE <- qgraph(graph_OLIFE, groups = groups_dimensios_OLIFEega,
                  label.prop = 0.8 , color = c("#66C2A5", "#fcfdbf", "#8DA0CB",
                                                "#f1605d"),
                  layout = "spring", vsize = 3, labels = dim.OLIFE$items,
                  title = "EGA model of the O-LIFE highlighting 4 factors (original model)")
```

#### EGA model of the O-LIFE highlighting 4 factors (original model)

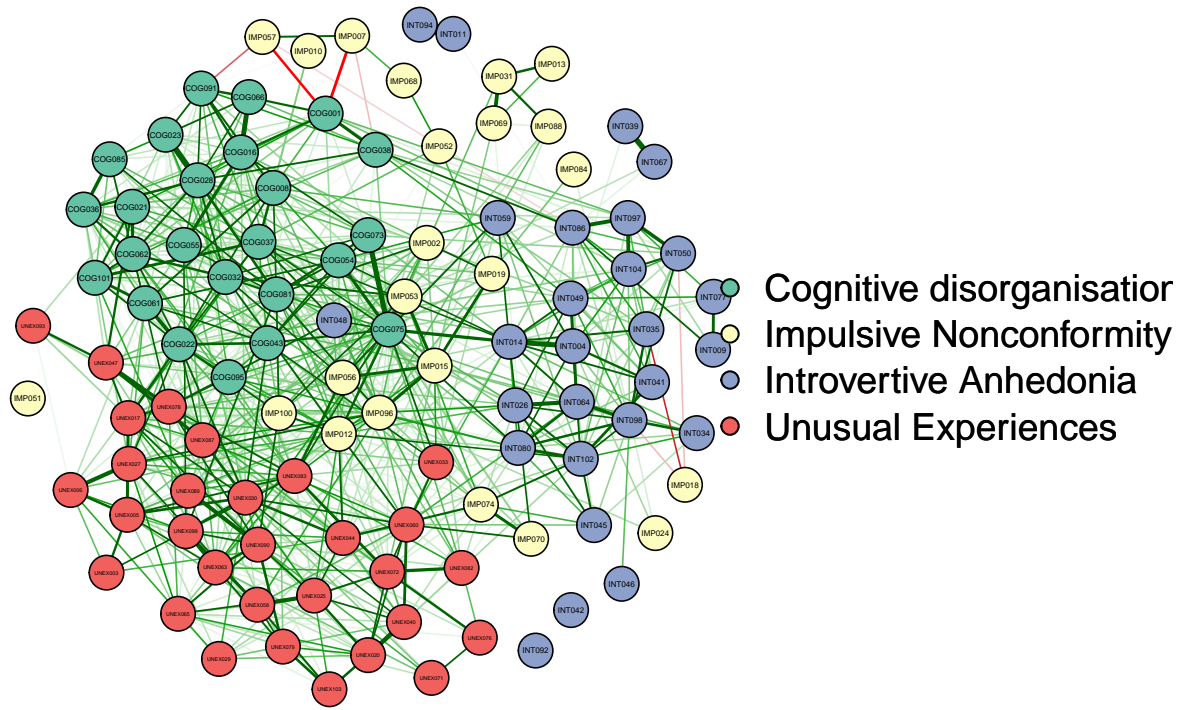

```
# pdf("EGA_OLIFE.pdf", width = 12)
# par(mfrow = c(1,2))
# dev.off()
```

#### 4. EXPLORATIVE FACTOR ANALYSIS TO REDUCE THE NUMBER OF FACTORS

```
# 13 are excessive and many items could create problems
```

```
library(GPArotation)
library(polycor)
```

```
# Running the Kaiser-Meyer-Olkin Test (KMO): determine how suited data is for
# factor analysis
KMO(OLIFE)
```

```
## Kaiser-Meyer-Olkin factor adequacy
```

```
## Call: KMO(r = OLIFE)
```

```
## Overall MSA = 0.9
```

```
## MSA for each item =
```

```
## COG001 COG008 COG016 COG021 COG022 COG023 COG028 COG032 COG036 COG037
## 0.92 0.93 0.92 0.88 0.94 0.89 0.92 0.93 0.93 0.93
## COG038 COG043 COG054 COG055 COG061 COG062 COG066 COG073 COG075 COG081
## 0.92 0.94 0.93 0.92 0.94 0.94 0.89 0.93 0.94 0.95
## COG085 COG091 COG095 COG101 IMP002 IMP007 IMP010 IMP012 IMP013 IMP015
## 0.89 0.90 0.92 0.93 0.89 0.72 0.75 0.93 0.64 0.91
```

```
## IMP018 IMP019 IMP024 IMP031 IMP051 IMP052 IMP053 IMP056 IMP057 IMP068
## 0.83 0.88 0.84 0.75 0.69 0.81 0.92 0.91 0.81 0.75
## IMP069 IMP070 IMP074 IMP084 IMP088 IMP096 IMP100 UNEX003 UNEX005 UNEX006
## 0.76 0.88 0.88 0.82 0.83 0.94 0.92 0.80 0.91 0.87
## UNEX017 UNEX020 UNEX025 UNEX027 UNEX029 UNEX030 UNEX033 UNEX040 UNEX044 UNEX047
## 0.92 0.85 0.89 0.93 0.84 0.94 0.92 0.88 0.90 0.90
## UNEX058 UNEX060 UNEX063 UNEX065 UNEX071 UNEX072 UNEX076 UNEX078 UNEX079 UNEX082
## 0.89 0.91 0.92 0.93 0.85 0.89 0.75 0.93 0.89 0.87
## UNEX083 UNEX087 UNEX089 UNEX090 UNEX093 UNEX099 UNEX103 INT004 INT009 INT011
## 0.94 0.92 0.91 0.90 0.84 0.93 0.82 0.88 0.75 0.64
## INT014 INT026 INT034 INT035 INT039 INT041 INT042 INT045 INT046 INT048
## 0.92 0.90 0.79 0.86 0.65 0.84 0.75 0.89 0.76 0.93
## INT049 INT050 INT059 INT064 INT067 INT077 INT080 INT086 INT092 INT094
## 0.89 0.89 0.91 0.88 0.68 0.81 0.93 0.90 0.67 0.66
## INT097 INT098 INT102 INT104
## 0.83 0.88 0.93 0.88
```

```
# Bartlett's test for Sphericity:
# If the p-value from Bartlett's Test of Sphericity is lower than our chosen
# significance level then our dataset is suitable for a data reduction technique
cortest.bartlett(OLIFE)
```

```
## $chisq
## [1] 26591.78
##
## $p.value
## [1] 0
##
## $df
## [1] 5356
```

```
# Compute the tetrachoric correlation matrix:
# Tetrachoric correlations are used to estimate the correlation between two
# categorical variables,

tetrachoric.corr <- tetrachoric(OLIFE)
```

```
tetracorr.matrix <- tetrachoric.corr$rho

# Determine the number of factors to extract
fa.parallel(tetracorr.matrix, fa = "fa")
```

#### Parallel Analysis Scree Plots

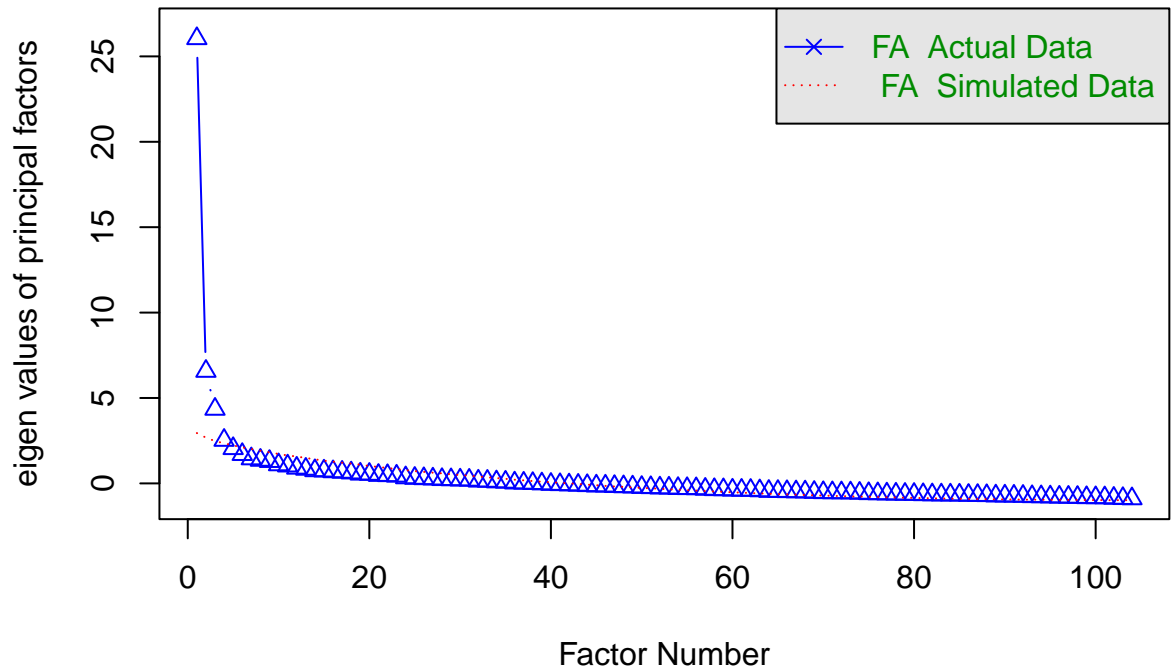

```
## Parallel analysis suggests that the number of factors = 4 and the number of components = NA
```

```
# Perform the factor analysis with the chosen number of factors (e.g., 4)
OLIFEefa_result <- fa(tetracorr.matrix, nfactors = 4, rotate = "promax")
# Oblique rotations allow the factors to be correlated, which often results
# in a more realistic representation of the underlying constructs, in particular
# in social sciences and psychology

# Print the results
print(OLIFEefa_result)
```

```
## Factor Analysis using method = minres
## Call: fa(r = tetracorr.matrix, nfactors = 4, rotate = "promax")
## Standardized loadings (pattern matrix) based upon correlation matrix
##          MR1   MR3   MR2   MR4   h2   u2 com
## COG001 -0.02  0.63  0.22 -0.30  0.46  0.54 1.7
## COG008 -0.15  0.54  0.08  0.25  0.40  0.60 1.6
## COG016 -0.01  0.64 -0.03  0.06  0.42  0.58 1.0
## COG021  0.04  0.48  0.12 -0.09  0.28  0.72 1.2
## COG022  0.28  0.38 -0.05  0.26  0.52  0.48 2.7
## COG023 -0.07  0.67 -0.08  0.05  0.39  0.61 1.1
## COG028 -0.01  0.73  0.03  0.02  0.56  0.44 1.0
## COG032  0.03  0.53 -0.06  0.20  0.40  0.60 1.3
## COG036  0.00  0.62 -0.05  0.07  0.41  0.59 1.0
## COG037  0.01  0.36  0.19  0.33  0.47  0.53 2.5
```

|  |  |  |  |  |  |  |  |  |
| --- | --- | --- | --- | --- | --- | --- | --- | --- |
| ## | COG038 | 0.00 | 0.57 | 0.26 | -0.25 | 0.43 | 0.57 | 1.8 |
| ## | COG043 | 0.19 | 0.46 | 0.01 | 0.22 | 0.52 | 0.48 | 1.8 |
| ## | COG054 | -0.05 | 0.39 | 0.21 | 0.35 | 0.51 | 0.49 | 2.6 |
| ## | COG055 | 0.15 | 0.52 | -0.14 | 0.03 | 0.33 | 0.67 | 1.3 |
| ## | COG061 | 0.11 | 0.46 | -0.07 | 0.24 | 0.41 | 0.59 | 1.7 |
| ## | COG062 | 0.16 | 0.63 | 0.02 | -0.07 | 0.49 | 0.51 | 1.2 |
| ## | COG066 | -0.19 | 0.67 | 0.04 | -0.04 | 0.36 | 0.64 | 1.2 |
| ## | COG073 | 0.00 | 0.47 | 0.18 | 0.18 | 0.44 | 0.56 | 1.6 |
| ## | COG075 | 0.12 | 0.41 | 0.28 | 0.25 | 0.66 | 0.34 | 2.7 |
| ## | COG081 | 0.14 | 0.45 | 0.12 | 0.22 | 0.52 | 0.48 | 1.9 |
| ## | COG085 | -0.02 | 0.57 | -0.06 | 0.09 | 0.33 | 0.67 | 1.1 |
| ## | COG091 | -0.17 | 0.69 | 0.04 | -0.06 | 0.39 | 0.61 | 1.1 |
| ## | COG095 | 0.27 | 0.45 | 0.09 | -0.04 | 0.44 | 0.56 | 1.7 |
| ## | COG101 | 0.04 | 0.46 | 0.01 | 0.19 | 0.35 | 0.65 | 1.3 |
| ## | IMP002 | -0.07 | 0.35 | -0.03 | 0.44 | 0.37 | 0.63 | 2.0 |
| ## | IMP007 | 0.04 | -0.56 | -0.03 | 0.56 | 0.40 | 0.60 | 2.0 |
| ## | IMP010 | -0.06 | 0.19 | -0.14 | 0.26 | 0.09 | 0.91 | 2.6 |
| ## | IMP012 | 0.28 | 0.17 | 0.24 | 0.24 | 0.50 | 0.50 | 3.6 |
| ## | IMP013 | -0.10 | -0.01 | -0.12 | 0.39 | 0.11 | 0.89 | 1.3 |
| ## | IMP015 | 0.02 | 0.22 | 0.16 | 0.45 | 0.44 | 0.56 | 1.7 |
| ## | IMP018 | -0.17 | -0.03 | -0.35 | 0.19 | 0.15 | 0.85 | 2.1 |
| ## | IMP019 | 0.02 | 0.12 | 0.00 | 0.49 | 0.31 | 0.69 | 1.1 |
| ## | IMP024 | -0.11 | 0.03 | 0.16 | 0.42 | 0.21 | 0.79 | 1.4 |
| ## | IMP031 | -0.17 | 0.14 | -0.08 | 0.48 | 0.19 | 0.81 | 1.5 |
| ## | IMP051 | 0.12 | -0.54 | 0.31 | 0.14 | 0.23 | 0.77 | 1.9 |
| ## | IMP052 | 0.28 | -0.18 | -0.04 | 0.29 | 0.19 | 0.81 | 2.7 |
| ## | IMP053 | 0.00 | 0.35 | 0.01 | 0.37 | 0.36 | 0.64 | 2.0 |
| ## | IMP056 | 0.12 | 0.43 | 0.03 | 0.10 | 0.33 | 0.67 | 1.3 |
| ## | IMP057 | 0.03 | -0.43 | -0.20 | 0.41 | 0.28 | 0.72 | 2.4 |
| ## | IMP068 | 0.23 | -0.34 | 0.11 | 0.26 | 0.17 | 0.83 | 3.0 |
| ## | IMP069 | 0.01 | -0.04 | 0.00 | 0.41 | 0.16 | 0.84 | 1.0 |
| ## | IMP070 | 0.12 | -0.04 | 0.29 | 0.43 | 0.42 | 0.58 | 2.0 |
| ## | IMP074 | 0.11 | 0.27 | 0.18 | 0.39 | 0.53 | 0.47 | 2.4 |
| ## | IMP084 | 0.06 | -0.09 | 0.05 | 0.41 | 0.19 | 0.81 | 1.2 |
| ## | IMP088 | 0.08 | 0.13 | -0.15 | 0.33 | 0.17 | 0.83 | 1.8 |
| ## | IMP096 | 0.25 | 0.18 | 0.21 | 0.28 | 0.49 | 0.51 | 3.6 |
| ## | IMP100 | 0.23 | 0.18 | 0.10 | 0.22 | 0.31 | 0.69 | 3.3 |
| ## | UNEX003 | 0.54 | -0.11 | -0.01 | -0.10 | 0.20 | 0.80 | 1.2 |
| ## | UNEX005 | 0.69 | 0.07 | -0.04 | -0.09 | 0.46 | 0.54 | 1.1 |
| ## | UNEX006 | 0.67 | -0.05 | -0.19 | -0.03 | 0.37 | 0.63 | 1.2 |
| ## | UNEX017 | 0.42 | 0.34 | 0.02 | 0.10 | 0.52 | 0.48 | 2.0 |
| ## | UNEX020 | 0.86 | -0.25 | 0.04 | -0.06 | 0.55 | 0.45 | 1.2 |
| ## | UNEX025 | 0.78 | -0.07 | 0.00 | 0.01 | 0.56 | 0.44 | 1.0 |
| ## | UNEX027 | 0.55 | 0.15 | -0.09 | 0.09 | 0.44 | 0.56 | 1.3 |
| ## | UNEX029 | 0.68 | -0.12 | -0.04 | -0.09 | 0.33 | 0.67 | 1.1 |
| ## | UNEX030 | 0.58 | 0.22 | -0.06 | 0.10 | 0.58 | 0.42 | 1.4 |
| ## | UNEX033 | 0.13 | 0.27 | 0.16 | 0.31 | 0.45 | 0.55 | 2.9 |
| ## | UNEX040 | 0.77 | -0.10 | 0.09 | 0.05 | 0.60 | 0.40 | 1.1 |
| ## | UNEX044 | 0.55 | -0.03 | 0.12 | 0.19 | 0.51 | 0.49 | 1.4 |
| ## | UNEX047 | 0.33 | 0.37 | 0.07 | -0.09 | 0.36 | 0.64 | 2.2 |
| ## | UNEX058 | 0.74 | 0.01 | -0.02 | -0.15 | 0.45 | 0.55 | 1.1 |
| ## | UNEX060 | 0.52 | -0.17 | 0.24 | 0.32 | 0.61 | 0.39 | 2.4 |
| ## | UNEX063 | 0.55 | 0.27 | -0.08 | -0.11 | 0.42 | 0.58 | 1.6 |
| ## | UNEX065 | 0.64 | -0.03 | -0.03 | 0.00 | 0.38 | 0.62 | 1.0 |

```
## UNEX071  0.34  0.15 -0.03  0.06  0.22  0.78  1.5
## UNEX072  0.65  0.21  0.00 -0.04  0.57  0.43  1.2
## UNEX076  0.57  0.05 -0.07 -0.07  0.29  0.71  1.1
## UNEX078  0.42  0.33  0.06  0.04  0.49  0.51  2.0
## UNEX079  0.60  0.04 -0.16  0.00  0.35  0.65  1.2
## UNEX082  0.31  0.31  0.13  0.03  0.39  0.61  2.3
## UNEX083  0.45  0.40 -0.09  0.04  0.53  0.47  2.1
## UNEX087  0.46  0.22  0.11  0.04  0.47  0.53  1.6
## UNEX089  0.38  0.34 -0.05  0.06  0.42  0.58  2.1
## UNEX090  0.58  0.08  0.06  0.09  0.49  0.51  1.1
## UNEX093  0.36  0.12  0.00 -0.10  0.15  0.85  1.4
## UNEX099  0.66  0.14 -0.13 -0.01  0.49  0.51  1.2
## UNEX103  0.74 -0.26 -0.05 -0.09  0.37  0.63  1.3
## INT004  -0.22  0.21  0.64  0.16  0.56  0.44  1.6
## INT009  -0.22  0.04  0.47  0.00  0.22  0.78  1.4
## INT011  -0.10  0.10  0.34  0.01  0.13  0.87  1.4
## INT014   0.08  0.12  0.67  0.13  0.68  0.32  1.2
## INT026   0.18 -0.05  0.54  0.13  0.44  0.56  1.4
## INT034   0.07 -0.31  0.68  0.08  0.43  0.57  1.5
## INT035  -0.16  0.18  0.62 -0.17  0.41  0.59  1.5
## INT039  -0.20 -0.17  0.49  0.16  0.24  0.76  1.8
## INT041   0.10 -0.07  0.72 -0.05  0.51  0.49  1.1
## INT042  -0.18 -0.22  0.29  0.01  0.12  0.88  2.6
## INT045   0.31 -0.22  0.56  0.00  0.40  0.60  1.9
## INT046  -0.06 -0.03  0.31  0.08  0.10  0.90  1.2
## INT048   0.16  0.42  0.30 -0.07  0.44  0.56  2.2
## INT049   0.05  0.02  0.62  0.01  0.43  0.57  1.0
## INT050   0.13  0.05  0.62 -0.29  0.39  0.61  1.5
## INT059   0.00  0.24  0.39  0.17  0.38  0.62  2.1
## INT064   0.18 -0.05  0.62  0.05  0.48  0.52  1.2
## INT067  -0.21  0.07  0.37  0.09  0.16  0.84  1.8
## INT077  -0.03  0.05  0.45 -0.12  0.19  0.81  1.2
## INT080   0.24 -0.02  0.57  0.12  0.55  0.45  1.4
## INT086  -0.04  0.24  0.66 -0.31  0.52  0.48  1.7
## INT092  -0.17 -0.10  0.37  0.04  0.12  0.88  1.6
## INT094  -0.04  0.12  0.29  0.01  0.12  0.88  1.4
## INT097   0.02  0.14  0.64 -0.42  0.46  0.54  1.8
## INT098   0.04 -0.11  0.71  0.06  0.49  0.51  1.1
## INT102   0.29 -0.02  0.70 -0.03  0.67  0.33  1.3
## INT104   0.00  0.20  0.61 -0.22  0.44  0.56  1.5
##
##
##          MR1   MR3   MR2  MR4
## SS loadings      12.41 12.14 10.02 5.59
## Proportion Var    0.12 0.12 0.10 0.05
## Cumulative Var    0.12 0.24 0.33 0.39
## Proportion Explained 0.31 0.30 0.25 0.14
## Cumulative Proportion 0.31 0.61 0.86 1.00
##
## With factor correlations of
##      MR1  MR3  MR2  MR4
## MR1 1.00 0.52 0.32 0.55
## MR3 0.52 1.00 0.41 0.37
## MR2 0.32 0.41 1.00 0.34
## MR4 0.55 0.37 0.34 1.00
```

```
##
## Mean item complexity = 1.7
## Test of the hypothesis that 4 factors are sufficient.
##
## df null model = 5356 with the objective function = 550.25
## df of the model are 4946 and the objective function was 507.09
##
## The root mean square of the residuals (RMSR) is 0.06
## The df corrected root mean square of the residuals is 0.06
##
## Fit based upon off diagonal values = 0.95
## Measures of factor score adequacy
##
## Correlation of (regression) scores with factors
## Multiple R square of scores with factors
## Minimum correlation of possible factor scores
```

|  | MR1 | MR3 | MR2 | MR4 |
| --- | --- | --- | --- | --- |
| Correlation of (regression) scores with factors | 0.98 | 0.97 | 0.97 | 0.95 |
| Multiple R square of scores with factors | 0.96 | 0.95 | 0.95 | 0.89 |
| Minimum correlation of possible factor scores | 0.91 | 0.90 | 0.89 | 0.79 |

```
OLIFEefa_result$Phi
```

```
##
## MR1 MR3 MR2 MR4
## MR1 1.0000000 0.5209268 0.3187341 0.5537213
## MR3 0.5209268 1.0000000 0.4138406 0.3743357
## MR2 0.3187341 0.4138406 1.0000000 0.3353848
## MR4 0.5537213 0.3743357 0.3353848 1.0000000
```

```
# Loadings for each item on each factor
print(OLIFEefa_result$loadings)
```

```
##
## Loadings:
## MR1 MR3 MR2 MR4
## COG001 0.629 0.221 -0.302
## COG008 -0.146 0.537 0.252
## COG016 0.641
## COG021 0.481 0.121
## COG022 0.279 0.376 0.258
## COG023 0.672
## COG028 0.734
## COG032 0.531 0.204
## COG036 0.624
## COG037 0.357 0.192 0.330
## COG038 0.569 0.264 -0.245
## COG043 0.190 0.457 0.223
## COG054 0.393 0.214 0.351
## COG055 0.147 0.523 -0.140
## COG061 0.111 0.458 0.243
## COG062 0.159 0.626
## COG066 -0.193 0.672
## COG073 0.469 0.179 0.177
## COG075 0.121 0.412 0.285 0.248
## COG081 0.140 0.445 0.120 0.218
## COG085 0.573
## COG091 -0.168 0.694
```

|  |  |  |  |  |
| --- | --- | --- | --- | --- |
| ## COG095 | 0.269 | 0.451 |  |  |
| ## COG101 |  | 0.464 |  | 0.190 |
| ## IMP002 |  | 0.348 |  | 0.445 |
| ## IMP007 |  | -0.556 |  | 0.565 |
| ## IMP010 |  | 0.185 | -0.143 | 0.256 |
| ## IMP012 | 0.283 | 0.172 | 0.244 | 0.241 |
| ## IMP013 |  |  | -0.123 | 0.389 |
| ## IMP015 |  | 0.215 | 0.156 | 0.451 |
| ## IMP018 | -0.168 |  | -0.346 | 0.188 |
| ## IMP019 |  | 0.120 |  | 0.488 |
| ## IMP024 | -0.107 |  | 0.162 | 0.424 |
| ## IMP031 | -0.167 | 0.142 |  | 0.475 |
| ## IMP051 | 0.123 | -0.540 | 0.311 | 0.140 |
| ## IMP052 | 0.282 | -0.175 |  | 0.288 |
| ## IMP053 |  | 0.351 |  | 0.366 |
| ## IMP056 | 0.120 | 0.433 |  | 0.104 |
| ## IMP057 |  | -0.431 | -0.203 | 0.413 |
| ## IMP068 | 0.228 | -0.336 | 0.114 | 0.264 |
| ## IMP069 |  |  |  | 0.411 |
| ## IMP070 | 0.123 |  | 0.290 | 0.428 |
| ## IMP074 | 0.108 | 0.274 | 0.177 | 0.390 |
| ## IMP084 |  |  |  | 0.411 |
| ## IMP088 |  | 0.126 | -0.152 | 0.335 |
| ## IMP096 | 0.255 | 0.183 | 0.208 | 0.279 |
| ## IMP100 | 0.226 | 0.182 |  | 0.219 |
| ## UNEX003 | 0.538 | -0.107 |  |  |
| ## UNEX005 | 0.692 |  |  |  |
| ## UNEX006 | 0.673 |  | -0.186 |  |
| ## UNEX017 | 0.416 | 0.336 |  |  |
| ## UNEX020 | 0.857 | -0.247 |  |  |
| ## UNEX025 | 0.783 |  |  |  |
| ## UNEX027 | 0.550 | 0.146 |  |  |
| ## UNEX029 | 0.677 | -0.115 |  |  |
| ## UNEX030 | 0.585 | 0.216 |  | 0.105 |
| ## UNEX033 | 0.135 | 0.272 | 0.161 | 0.314 |
| ## UNEX040 | 0.766 | -0.101 |  |  |
| ## UNEX044 | 0.549 |  | 0.123 | 0.191 |
| ## UNEX047 | 0.331 | 0.368 |  |  |
| ## UNEX058 | 0.740 |  |  | -0.149 |
| ## UNEX060 | 0.519 | -0.171 | 0.242 | 0.318 |
| ## UNEX063 | 0.548 | 0.268 |  | -0.106 |
| ## UNEX065 | 0.638 |  |  |  |
| ## UNEX071 | 0.344 | 0.152 |  |  |
| ## UNEX072 | 0.646 | 0.207 |  |  |
| ## UNEX076 | 0.569 |  |  |  |
| ## UNEX078 | 0.420 | 0.326 |  |  |
| ## UNEX079 | 0.605 |  | -0.161 |  |
| ## UNEX082 | 0.314 | 0.312 | 0.126 |  |
| ## UNEX083 | 0.445 | 0.399 |  |  |
| ## UNEX087 | 0.464 | 0.220 | 0.113 |  |
| ## UNEX089 | 0.383 | 0.340 |  |  |
| ## UNEX090 | 0.575 |  |  |  |
| ## UNEX093 | 0.357 | 0.124 |  | -0.104 |
| ## UNEX099 | 0.661 | 0.142 | -0.132 |  |

```
## UNEX103  0.745 -0.257
## INT004  -0.217  0.205  0.643  0.158
## INT009  -0.217          0.475
## INT011  -0.102          0.335
## INT014          0.123  0.671  0.132
## INT026   0.183          0.538  0.128
## INT034          -0.313  0.683
## INT035  -0.162  0.182  0.622 -0.168
## INT039  -0.204 -0.167  0.491  0.159
## INT041          0.723
## INT042  -0.179 -0.217  0.290
## INT045   0.311 -0.216  0.559
## INT046          0.313
## INT048   0.155  0.416  0.298
## INT049          0.625
## INT050   0.126          0.615 -0.292
## INT059          0.242  0.387  0.165
## INT064   0.179          0.615
## INT067  -0.208          0.370
## INT077          0.448 -0.118
## INT080   0.238          0.570  0.122
## INT086          0.237  0.662 -0.307
## INT092  -0.170 -0.104  0.372
## INT094          0.121  0.295
## INT097          0.141  0.640 -0.422
## INT098          -0.106  0.706
## INT102   0.288          0.701
## INT104          0.198  0.609 -0.223
##
##              MR1      MR3      MR2      MR4
## SS loadings  11.406 10.800 9.385 4.949
## Proportion Var 0.110 0.104 0.090 0.048
## Cumulative Var 0.110 0.214 0.304 0.351
```

*# The loadings are divided in the 4 factors but multiple items have high*
*# positive or negative correlation with more than 1 factor at the same time.*
*# This can create problems.*

*# Summarise the model with the Exploratory Factor Analysis*

```
model_OLIFE_efa <- '
Factor1 =~ `IMP012` + `IMP100` + `UNEX003` + `UNEX005` + `UNEX006` + `UNEX017` +
            `UNEX020` + `UNEX025` + `UNEX027` + `UNEX029` + `UNEX030` +
            `UNEX040` + `UNEX044` + `UNEX058` + `UNEX060` + `UNEX063` +
            `UNEX065` + `UNEX071` + `UNEX072` + `UNEX076` + `UNEX078` +
            `UNEX079` + `UNEX082` + `UNEX083` + `UNEX087` + `UNEX089` +
            `UNEX090` + `UNEX093` + `UNEX099` + `UNEX103`
Factor2 =~ `IMP018` + `INT004` + `INT009` + `INT011` + `INT014` + `INT026` +
            `INT034` + `INT035` + `INT039` + `INT041` + `INT042` + `INT045` +
            `INT046` + `INT049` + `INT050` + `INT059` + `INT064` + `INT067` +
            `INT077` + `INT080` + `INT086` + `INT092` + `INT094` + `INT097` +
            `INT098` + `INT102` + `INT104`
Factor3 =~ `COG001` + `COG008` + `COG016` + `COG021` + `COG022` + `COG023` +
            `COG028` + `COG032` + `COG036` + `COG037` + `COG038` + `COG043` +
            `COG054` + `COG055` + `COG061` + `COG062` + `COG066` + `COG073` +
```

```
`COG075` + `COG081` + `COG085` + `COG091` + `COG095` + `COG101` +
`IMP051` + `IMP056` + `IMP057` + `IMP068` + `UNEX047` + `INT048`
Factor4 =~ `IMP002` + `IMP007` + `IMP010` + `IMP013` + `IMP015` + `IMP019` +
`IMP024` + `IMP031` + `IMP052` + `IMP053` + `IMP069` +
`IMP070` + `IMP074` + `IMP084` + `IMP088` + `IMP096` + `UNEX033`
,

# The Kaiser-Meyer-Olkin Test showed multiple items with mediocre or middling
# MSA (measure of sampling adequacy).

# We did an Exploratory Factor Analysis NOT considering the variables with
# MSA < 0.7 (mediocre)

OLIFE_7 <- OLIFE[, -c(29,35,80,85,95,99,100)]

mat.OLIFE_7 <- cor(OLIFE_7, method = "spearman")

ega_OLIFE_7 <- ega.object(mat.OLIFE_7, OLIFE_7)

dim.OLIFE_7 <- ega_OLIFE_7$dim.variables
# From 13 to 9 dimensions ad 4 were the most predominant factors but still some
# problems in the classification

oneOLIFE_7 <- which(dim.OLIFE_7$dimension == 1)
dim.OLIFE_7$dimension[oneOLIFE_7] <- rep("Factor 01", length(oneOLIFE_7))
twoOLIFE_7 <- which(dim.OLIFE_7$dimension == 2)
dim.OLIFE_7$dimension[twoOLIFE_7] <- rep("Factor 02", length(twoOLIFE_7))
threeOLIFE_7 <- which(dim.OLIFE_7$dimension == 3)
dim.OLIFE_7$dimension[threeOLIFE_7] <- rep("Factor 03", length(threeOLIFE_7))
fourOLIFE_7 <- which(dim.OLIFE_7$dimension == 4)
dim.OLIFE_7$dimension[fourOLIFE_7] <- rep("Factor 04", length(fourOLIFE_7))
fiveOLIFE_7 <- which(dim.OLIFE_7$dimension == 5)
dim.OLIFE_7$dimension[fiveOLIFE_7] <- rep("Factor 05", length(fiveOLIFE_7))
sixOLIFE_7 <- which(dim.OLIFE_7$dimension == 6)
dim.OLIFE_7$dimension[sixOLIFE_7] <- rep("Factor 06", length(sixOLIFE_7))
sevenOLIFE_7 <- which(dim.OLIFE_7$dimension == 7)
dim.OLIFE_7$dimension[sevenOLIFE_7] <- rep("Factor 07", length(sevenOLIFE_7))
eightOLIFE_7 <- which(dim.OLIFE_7$dimension == 8)
dim.OLIFE_7$dimension[eightOLIFE_7] <- rep("Factor 08", length(eightOLIFE_7))
nineOLIFE_7 <- which(dim.OLIFE_7$dimension == 9)
dim.OLIFE_7$dimension[nineOLIFE_7] <- rep("Factor 09", length(nineOLIFE_7))

graph_OLIFE_7 <- ega_OLIFE_7$glasso[match(dim.OLIFE_7$items, colnames(OLIFE_7)),
                                     match(dim.OLIFE_7$items, colnames(OLIFE_7))]
A_OLIFE_7 <- qgraph(graph_OLIFE_7, groups = dim.OLIFE_7$dimension,
                    label.prop = 0.8 , color = c("#f1605d", "#fed799", "#feb078",
                                                  "#fc8961", "#fcfdbf", "#d8456c",
                                                  "#b73779", "#932b80", "#721f81"),
                    layout = "spring", vsize = 4, labels = dim.OLIFE_7$items)
```

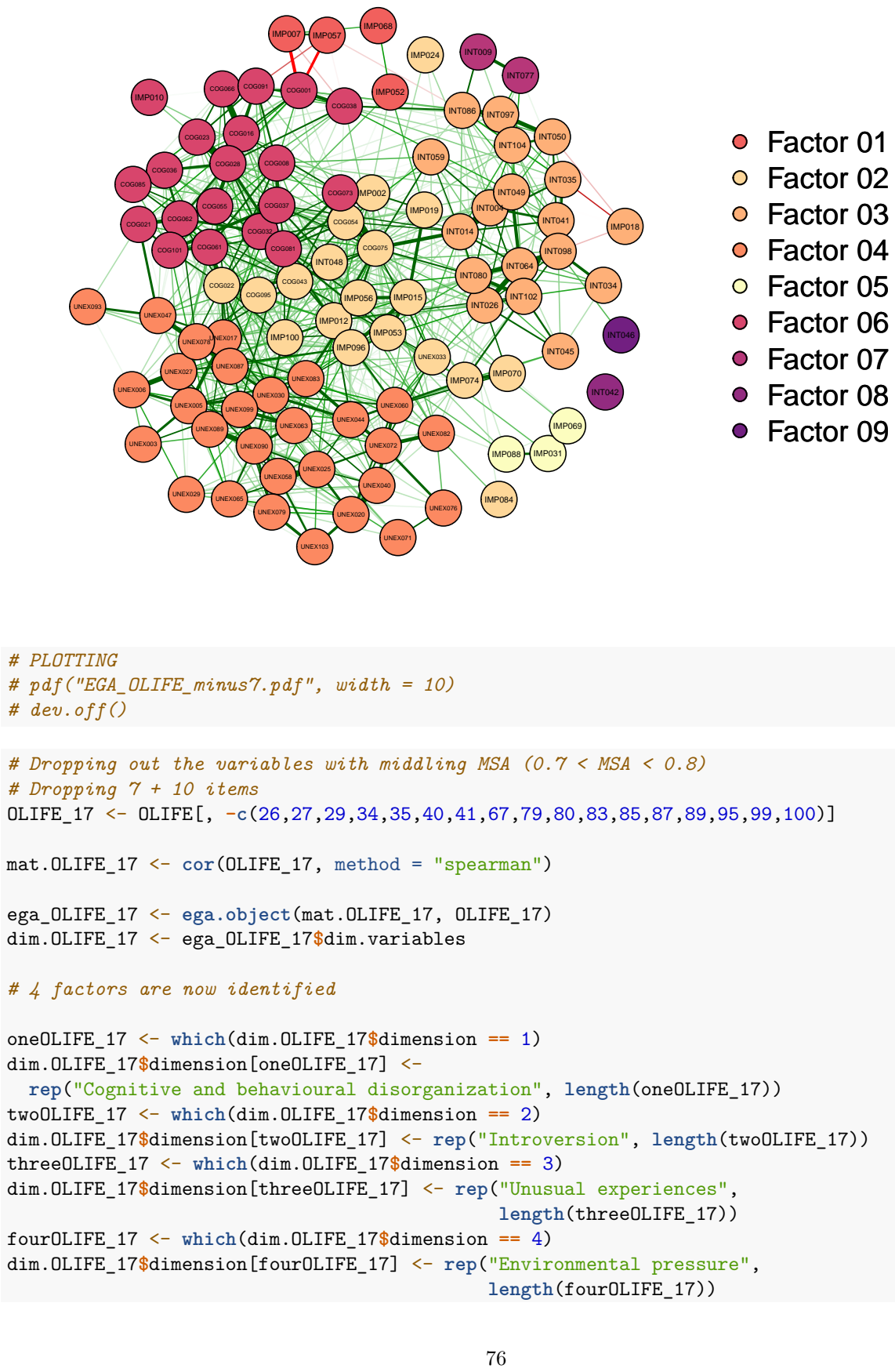

```
# PLOTTING
# pdf("EGA_OLIFE_minus7.pdf",  width = 10)
# dev.off()
# Dropping out  the variables with middling MSA (0.7 < MSA < 0.8)
# Dropping 7 + 10  items
OLIFE_17 <- OLIFE[, -c(26,27,29,34,35,40,41,67,79,80,83,85,87,89,95,99,100)]
mat.OLIFE_17 <- cor(OLIFE_17, method  = "spearman")
ega_OLIFE_17 <- ega.object(mat.OLIFE_17,  OLIFE_17)
dim.OLIFE_17 <- ega_OLIFE_17$dim.variables
# 4 factors are now  identified
oneOLIFE_17 <- which(dim.OLIFE_17$dimension  == 1)
dim.OLIFE_17$dimension[oneOLIFE_17]  <-
  rep("Cognitive and behavioural  disorganization", length(oneOLIFE_17))
twoOLIFE_17 <- which(dim.OLIFE_17$dimension  == 2)
dim.OLIFE_17$dimension[twoOLIFE_17]  <- rep("Introversion", length(twoOLIFE_17))
threeOLIFE_17 <- which(dim.OLIFE_17$dimension  == 3)
dim.OLIFE_17$dimension[threeOLIFE_17]  <- rep("Unusual experiences",
                                              length(threeOLIFE_17))
fourOLIFE_17 <- which(dim.OLIFE_17$dimension  == 4)
dim.OLIFE_17$dimension[fourOLIFE_17]  <- rep("Environmental pressure",
                                             length(fourOLIFE_17))
```

```
graph_OLIFE_17 <- ega_OLIFE_17$glasso[match(dim.OLIFE_17$items,
                                           colnames(OLIFE_17)),
                                           match(dim.OLIFE_17$items,colnames(OLIFE_17))]
```

```
A_OLIFE_17 <- qgraph(graph_OLIFE_17, groups = dim.OLIFE_17$dimension,
                      label.prop = 0.8 , color = c("#fcfdbf", "#fc8961", "#b73779",
                                                    "#1d1147"),
                      layout = "spring", vsize = 4,
                      labels = dim.OLIFE_17$items)
```

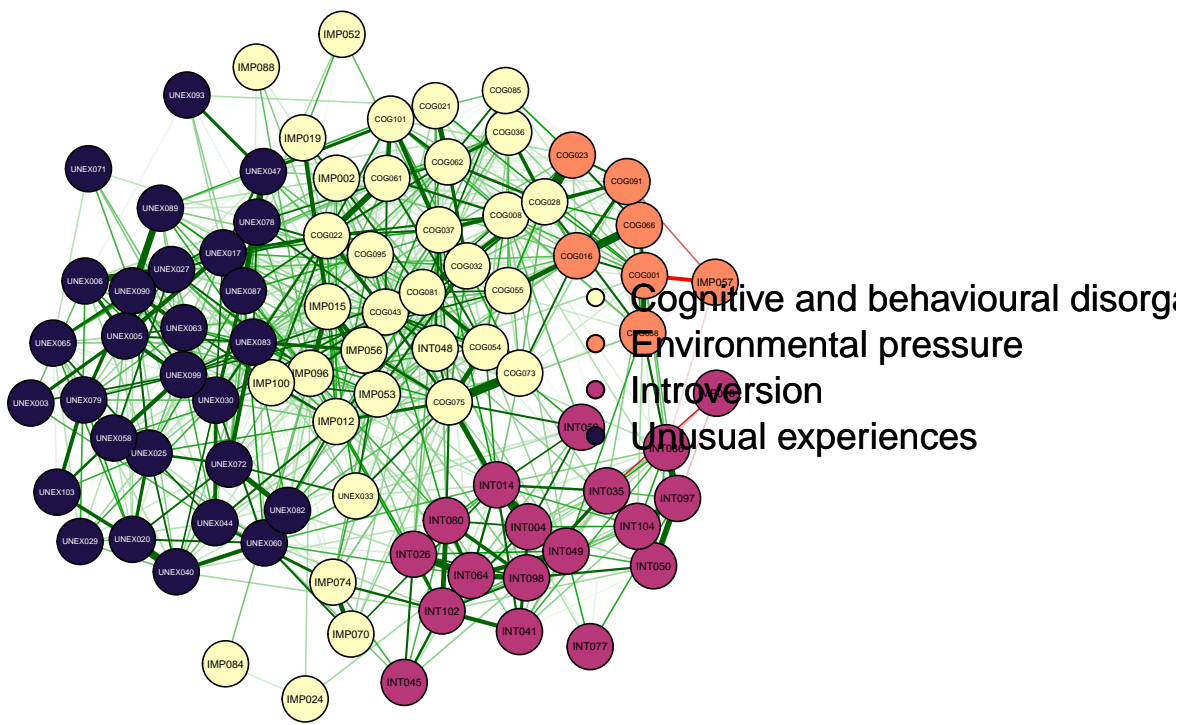

```
# Centrality measures of the network and representation in 4 FACTOR NETWORK
centralityTableOLIFE <- centralityTable(A_OLIFE_17, relative = TRUE,
                                         standardized = FALSE)

centralityTableOLIFE
```

| ## | graph | type | node | measure | value |
| --- | --- | --- | --- | --- | --- |
| ## 1 | graph | 1 | NA | COG008 | Betweenness 0.071030641 |
| ## 2 | graph | 1 | NA | COG021 | Betweenness 0.125348189 |
| ## 3 | graph | 1 | NA | COG022 | Betweenness 0.217270195 |
| ## 4 | graph | 1 | NA | COG028 | Betweenness 0.383008357 |
| ## 5 | graph | 1 | NA | COG032 | Betweenness 0.094707521 |
| ## 6 | graph | 1 | NA | COG036 | Betweenness 0.022284123 |
| ## 7 | graph | 1 | NA | COG037 | Betweenness 0.194986072 |

|  |  |  |  |  |  |
| --- | --- | --- | --- | --- | --- |
| ## 8 | graph 1 | NA | COG043 | Betweenness | 0.066852368 |
| ## 9 | graph 1 | NA | COG054 | Betweenness | 0.030640669 |
| ## 10 | graph 1 | NA | COG055 | Betweenness | 0.089136490 |
| ## 11 | graph 1 | NA | COG061 | Betweenness | 0.044568245 |
| ## 12 | graph 1 | NA | COG062 | Betweenness | 0.167130919 |
| ## 13 | graph 1 | NA | COG073 | Betweenness | 0.221448468 |
| ## 14 | graph 1 | NA | COG075 | Betweenness | 1.000000000 |
| ## 15 | graph 1 | NA | COG081 | Betweenness | 0.437325905 |
| ## 16 | graph 1 | NA | COG085 | Betweenness | 0.002785515 |
| ## 17 | graph 1 | NA | COG095 | Betweenness | 0.041782730 |
| ## 18 | graph 1 | NA | COG101 | Betweenness | 0.068245125 |
| ## 19 | graph 1 | NA | IMP002 | Betweenness | 0.002785515 |
| ## 20 | graph 1 | NA | IMP012 | Betweenness | 0.243732591 |
| ## 21 | graph 1 | NA | IMP015 | Betweenness | 0.250696379 |
| ## 22 | graph 1 | NA | IMP019 | Betweenness | 0.038997214 |
| ## 23 | graph 1 | NA | IMP024 | Betweenness | 0.000000000 |
| ## 24 | graph 1 | NA | IMP052 | Betweenness | 0.000000000 |
| ## 25 | graph 1 | NA | IMP053 | Betweenness | 0.045961003 |
| ## 26 | graph 1 | NA | IMP056 | Betweenness | 0.175487465 |
| ## 27 | graph 1 | NA | IMP070 | Betweenness | 0.019498607 |
| ## 28 | graph 1 | NA | IMP074 | Betweenness | 0.091922006 |
| ## 29 | graph 1 | NA | IMP084 | Betweenness | 0.000000000 |
| ## 30 | graph 1 | NA | IMP088 | Betweenness | 0.000000000 |
| ## 31 | graph 1 | NA | IMP096 | Betweenness | 0.511142061 |
| ## 32 | graph 1 | NA | IMP100 | Betweenness | 0.019498607 |
| ## 33 | graph 1 | NA | UNEX033 | Betweenness | 0.001392758 |
| ## 34 | graph 1 | NA | INT048 | Betweenness | 0.043175487 |
| ## 35 | graph 1 | NA | IMP018 | Betweenness | 0.000000000 |
| ## 36 | graph 1 | NA | INT004 | Betweenness | 0.080779944 |
| ## 37 | graph 1 | NA | INT014 | Betweenness | 0.731197772 |
| ## 38 | graph 1 | NA | INT026 | Betweenness | 0.135097493 |
| ## 39 | graph 1 | NA | INT035 | Betweenness | 0.143454039 |
| ## 40 | graph 1 | NA | INT041 | Betweenness | 0.006963788 |
| ## 41 | graph 1 | NA | INT045 | Betweenness | 0.000000000 |
| ## 42 | graph 1 | NA | INT049 | Betweenness | 0.151810585 |
| ## 43 | graph 1 | NA | INT050 | Betweenness | 0.041782730 |
| ## 44 | graph 1 | NA | INT059 | Betweenness | 0.000000000 |
| ## 45 | graph 1 | NA | INT064 | Betweenness | 0.236768802 |
| ## 46 | graph 1 | NA | INT077 | Betweenness | 0.000000000 |
| ## 47 | graph 1 | NA | INT080 | Betweenness | 0.179665738 |
| ## 48 | graph 1 | NA | INT086 | Betweenness | 0.105849582 |
| ## 49 | graph 1 | NA | INT097 | Betweenness | 0.100278552 |
| ## 50 | graph 1 | NA | INT098 | Betweenness | 0.140668524 |
| ## 51 | graph 1 | NA | INT102 | Betweenness | 0.147632312 |
| ## 52 | graph 1 | NA | INT104 | Betweenness | 0.065459610 |
| ## 53 | graph 1 | NA | UNEX003 | Betweenness | 0.000000000 |
| ## 54 | graph 1 | NA | UNEX005 | Betweenness | 0.128133705 |
| ## 55 | graph 1 | NA | UNEX006 | Betweenness | 0.059888579 |
| ## 56 | graph 1 | NA | UNEX017 | Betweenness | 0.094707521 |
| ## 57 | graph 1 | NA | UNEX020 | Betweenness | 0.091922006 |
| ## 58 | graph 1 | NA | UNEX025 | Betweenness | 0.125348189 |
| ## 59 | graph 1 | NA | UNEX027 | Betweenness | 0.140668524 |
| ## 60 | graph 1 | NA | UNEX029 | Betweenness | 0.000000000 |
| ## 61 | graph 1 | NA | UNEX030 | Betweenness | 0.334261838 |

|  |  |  |  |  |
| --- | --- | --- | --- | --- |
| ## 62 | graph 1 | NA UNEX040 | Betweenness | 0.107242340 |
| ## 63 | graph 1 | NA UNEX044 | Betweenness | 0.105849582 |
| ## 64 | graph 1 | NA UNEX047 | Betweenness | 0.240947075 |
| ## 65 | graph 1 | NA UNEX058 | Betweenness | 0.133704735 |
| ## 66 | graph 1 | NA UNEX060 | Betweenness | 0.197771588 |
| ## 67 | graph 1 | NA UNEX063 | Betweenness | 0.155988858 |
| ## 68 | graph 1 | NA UNEX065 | Betweenness | 0.004178273 |
| ## 69 | graph 1 | NA UNEX071 | Betweenness | 0.000000000 |
| ## 70 | graph 1 | NA UNEX072 | Betweenness | 0.110027855 |
| ## 71 | graph 1 | NA UNEX078 | Betweenness | 0.136490251 |
| ## 72 | graph 1 | NA UNEX079 | Betweenness | 0.025069638 |
| ## 73 | graph 1 | NA UNEX082 | Betweenness | 0.005571031 |
| ## 74 | graph 1 | NA UNEX083 | Betweenness | 0.185236769 |
| ## 75 | graph 1 | NA UNEX087 | Betweenness | 0.139275766 |
| ## 76 | graph 1 | NA UNEX089 | Betweenness | 0.048746518 |
| ## 77 | graph 1 | NA UNEX090 | Betweenness | 0.260445682 |
| ## 78 | graph 1 | NA UNEX093 | Betweenness | 0.000000000 |
| ## 79 | graph 1 | NA UNEX099 | Betweenness | 0.353760446 |
| ## 80 | graph 1 | NA UNEX103 | Betweenness | 0.016713092 |
| ## 81 | graph 1 | NA COG001 | Betweenness | 0.189415042 |
| ## 82 | graph 1 | NA COG016 | Betweenness | 0.207520891 |
| ## 83 | graph 1 | NA COG023 | Betweenness | 0.012534819 |
| ## 84 | graph 1 | NA COG038 | Betweenness | 0.201949861 |
| ## 85 | graph 1 | NA COG066 | Betweenness | 0.005571031 |
| ## 86 | graph 1 | NA COG091 | Betweenness | 0.033426184 |
| ## 87 | graph 1 | NA IMP057 | Betweenness | 0.000000000 |
| ## 88 | graph 1 | NA COG008 | Closeness | 0.770017453 |
| ## 89 | graph 1 | NA COG021 | Closeness | 0.760248443 |
| ## 90 | graph 1 | NA COG022 | Closeness | 0.872047410 |
| ## 91 | graph 1 | NA COG028 | Closeness | 0.833619132 |
| ## 92 | graph 1 | NA COG032 | Closeness | 0.828399860 |
| ## 93 | graph 1 | NA COG036 | Closeness | 0.708718429 |
| ## 94 | graph 1 | NA COG037 | Closeness | 0.829979379 |
| ## 95 | graph 1 | NA COG043 | Closeness | 0.856087265 |
| ## 96 | graph 1 | NA COG054 | Closeness | 0.865310196 |
| ## 97 | graph 1 | NA COG055 | Closeness | 0.729095424 |
| ## 98 | graph 1 | NA COG061 | Closeness | 0.803175160 |
| ## 99 | graph 1 | NA COG062 | Closeness | 0.797616474 |
| ## 100 | graph 1 | NA COG073 | Closeness | 0.936796311 |
| ## 101 | graph 1 | NA COG075 | Closeness | 1.000000000 |
| ## 102 | graph 1 | NA COG081 | Closeness | 0.933472657 |
| ## 103 | graph 1 | NA COG085 | Closeness | 0.678429035 |
| ## 104 | graph 1 | NA COG095 | Closeness | 0.782918479 |
| ## 105 | graph 1 | NA COG101 | Closeness | 0.784592754 |
| ## 106 | graph 1 | NA IMP002 | Closeness | 0.688323153 |
| ## 107 | graph 1 | NA IMP012 | Closeness | 0.921413265 |
| ## 108 | graph 1 | NA IMP015 | Closeness | 0.836021064 |
| ## 109 | graph 1 | NA IMP019 | Closeness | 0.683134245 |
| ## 110 | graph 1 | NA IMP024 | Closeness | 0.372155030 |
| ## 111 | graph 1 | NA IMP052 | Closeness | 0.503325504 |
| ## 112 | graph 1 | NA IMP053 | Closeness | 0.760759996 |
| ## 113 | graph 1 | NA IMP056 | Closeness | 0.865288682 |
| ## 114 | graph 1 | NA IMP070 | Closeness | 0.692436534 |
| ## 115 | graph 1 | NA IMP074 | Closeness | 0.715294030 |

|  |  |  |  |  |  |
| --- | --- | --- | --- | --- | --- |
| ## 116 | graph 1 | NA | IMP084 | Closeness | 0.449674244 |
| ## 117 | graph 1 | NA | IMP088 | Closeness | 0.482141507 |
| ## 118 | graph 1 | NA | IMP096 | Closeness | 0.891014651 |
| ## 119 | graph 1 | NA | IMP100 | Closeness | 0.800514153 |
| ## 120 | graph 1 | NA | UNEX033 | Closeness | 0.751722572 |
| ## 121 | graph 1 | NA | INT048 | Closeness | 0.768989175 |
| ## 122 | graph 1 | NA | IMP018 | Closeness | 0.462014089 |
| ## 123 | graph 1 | NA | INT004 | Closeness | 0.811411748 |
| ## 124 | graph 1 | NA | INT014 | Closeness | 0.896665097 |
| ## 125 | graph 1 | NA | INT026 | Closeness | 0.752296706 |
| ## 126 | graph 1 | NA | INT035 | Closeness | 0.686696248 |
| ## 127 | graph 1 | NA | INT041 | Closeness | 0.687878528 |
| ## 128 | graph 1 | NA | INT045 | Closeness | 0.597914542 |
| ## 129 | graph 1 | NA | INT049 | Closeness | 0.715775663 |
| ## 130 | graph 1 | NA | INT050 | Closeness | 0.568673609 |
| ## 131 | graph 1 | NA | INT059 | Closeness | 0.689395216 |
| ## 132 | graph 1 | NA | INT064 | Closeness | 0.757543722 |
| ## 133 | graph 1 | NA | INT077 | Closeness | 0.489980162 |
| ## 134 | graph 1 | NA | INT080 | Closeness | 0.795070639 |
| ## 135 | graph 1 | NA | INT086 | Closeness | 0.579423519 |
| ## 136 | graph 1 | NA | INT097 | Closeness | 0.582700217 |
| ## 137 | graph 1 | NA | INT098 | Closeness | 0.692159357 |
| ## 138 | graph 1 | NA | INT102 | Closeness | 0.739339578 |
| ## 139 | graph 1 | NA | INT104 | Closeness | 0.599339932 |
| ## 140 | graph 1 | NA | UNEX003 | Closeness | 0.642397543 |
| ## 141 | graph 1 | NA | UNEX005 | Closeness | 0.756516157 |
| ## 142 | graph 1 | NA | UNEX006 | Closeness | 0.722822714 |
| ## 143 | graph 1 | NA | UNEX017 | Closeness | 0.786871230 |
| ## 144 | graph 1 | NA | UNEX020 | Closeness | 0.689472906 |
| ## 145 | graph 1 | NA | UNEX025 | Closeness | 0.768437841 |
| ## 146 | graph 1 | NA | UNEX027 | Closeness | 0.779681575 |
| ## 147 | graph 1 | NA | UNEX029 | Closeness | 0.538636482 |
| ## 148 | graph 1 | NA | UNEX030 | Closeness | 0.854801649 |
| ## 149 | graph 1 | NA | UNEX040 | Closeness | 0.719120102 |
| ## 150 | graph 1 | NA | UNEX044 | Closeness | 0.783200446 |
| ## 151 | graph 1 | NA | UNEX047 | Closeness | 0.782527795 |
| ## 152 | graph 1 | NA | UNEX058 | Closeness | 0.769598277 |
| ## 153 | graph 1 | NA | UNEX060 | Closeness | 0.810053426 |
| ## 154 | graph 1 | NA | UNEX063 | Closeness | 0.807172670 |
| ## 155 | graph 1 | NA | UNEX065 | Closeness | 0.636470472 |
| ## 156 | graph 1 | NA | UNEX071 | Closeness | 0.486080703 |
| ## 157 | graph 1 | NA | UNEX072 | Closeness | 0.725302666 |
| ## 158 | graph 1 | NA | UNEX078 | Closeness | 0.791390907 |
| ## 159 | graph 1 | NA | UNEX079 | Closeness | 0.653331237 |
| ## 160 | graph 1 | NA | UNEX082 | Closeness | 0.662516133 |
| ## 161 | graph 1 | NA | UNEX083 | Closeness | 0.833846991 |
| ## 162 | graph 1 | NA | UNEX087 | Closeness | 0.806736663 |
| ## 163 | graph 1 | NA | UNEX089 | Closeness | 0.717453905 |
| ## 164 | graph 1 | NA | UNEX090 | Closeness | 0.763652317 |
| ## 165 | graph 1 | NA | UNEX093 | Closeness | 0.608365074 |
| ## 166 | graph 1 | NA | UNEX099 | Closeness | 0.848553881 |
| ## 167 | graph 1 | NA | UNEX103 | Closeness | 0.635666442 |
| ## 168 | graph 1 | NA | COG001 | Closeness | 0.670256777 |
| ## 169 | graph 1 | NA | COG016 | Closeness | 0.745607507 |

|  |  |  |  |  |
| --- | --- | --- | --- | --- |
| ## 170 graph 1 | NA | COG023 | Closeness | 0.766976087 |
| ## 171 graph 1 | NA | COG038 | Closeness | 0.681957266 |
| ## 172 graph 1 | NA | COG066 | Closeness | 0.706449081 |
| ## 173 graph 1 | NA | COG091 | Closeness | 0.672202998 |
| ## 174 graph 1 | NA | IMP057 | Closeness | 0.557441114 |
| ## 175 graph 1 | NA | COG008 | Strength | 0.457774887 |
| ## 176 graph 1 | NA | COG021 | Strength | 0.321744452 |
| ## 177 graph 1 | NA | COG022 | Strength | 0.735142221 |
| ## 178 graph 1 | NA | COG028 | Strength | 0.787161175 |
| ## 179 graph 1 | NA | COG032 | Strength | 0.467752723 |
| ## 180 graph 1 | NA | COG036 | Strength | 0.429987675 |
| ## 181 graph 1 | NA | COG037 | Strength | 0.570021949 |
| ## 182 graph 1 | NA | COG043 | Strength | 0.537273563 |
| ## 183 graph 1 | NA | COG054 | Strength | 0.566118523 |
| ## 184 graph 1 | NA | COG055 | Strength | 0.336417187 |
| ## 185 graph 1 | NA | COG061 | Strength | 0.486917155 |
| ## 186 graph 1 | NA | COG062 | Strength | 0.630463981 |
| ## 187 graph 1 | NA | COG073 | Strength | 0.465761125 |
| ## 188 graph 1 | NA | COG075 | Strength | 1.000000000 |
| ## 189 graph 1 | NA | COG081 | Strength | 0.747029597 |
| ## 190 graph 1 | NA | COG085 | Strength | 0.283689117 |
| ## 191 graph 1 | NA | COG095 | Strength | 0.375427099 |
| ## 192 graph 1 | NA | COG101 | Strength | 0.451131672 |
| ## 193 graph 1 | NA | IMP002 | Strength | 0.293911774 |
| ## 194 graph 1 | NA | IMP012 | Strength | 0.693728450 |
| ## 195 graph 1 | NA | IMP015 | Strength | 0.547583480 |
| ## 196 graph 1 | NA | IMP019 | Strength | 0.210638027 |
| ## 197 graph 1 | NA | IMP024 | Strength | 0.028477701 |
| ## 198 graph 1 | NA | IMP052 | Strength | 0.059562341 |
| ## 199 graph 1 | NA | IMP053 | Strength | 0.395992510 |
| ## 200 graph 1 | NA | IMP056 | Strength | 0.379638763 |
| ## 201 graph 1 | NA | IMP070 | Strength | 0.187354602 |
| ## 202 graph 1 | NA | IMP074 | Strength | 0.313875125 |
| ## 203 graph 1 | NA | IMP084 | Strength | 0.023551992 |
| ## 204 graph 1 | NA | IMP088 | Strength | 0.048121181 |
| ## 205 graph 1 | NA | IMP096 | Strength | 0.652833355 |
| ## 206 graph 1 | NA | IMP100 | Strength | 0.300167955 |
| ## 207 graph 1 | NA | UNEX033 | Strength | 0.231917405 |
| ## 208 graph 1 | NA | INT048 | Strength | 0.407807558 |
| ## 209 graph 1 | NA | IMP018 | Strength | 0.045926250 |
| ## 210 graph 1 | NA | INT004 | Strength | 0.435872320 |
| ## 211 graph 1 | NA | INT014 | Strength | 0.721280537 |
| ## 212 graph 1 | NA | INT026 | Strength | 0.445189308 |
| ## 213 graph 1 | NA | INT035 | Strength | 0.292560547 |
| ## 214 graph 1 | NA | INT041 | Strength | 0.272292994 |
| ## 215 graph 1 | NA | INT045 | Strength | 0.178671601 |
| ## 216 graph 1 | NA | INT049 | Strength | 0.403556734 |
| ## 217 graph 1 | NA | INT050 | Strength | 0.304761583 |
| ## 218 graph 1 | NA | INT059 | Strength | 0.219830409 |
| ## 219 graph 1 | NA | INT064 | Strength | 0.476572108 |
| ## 220 graph 1 | NA | INT077 | Strength | 0.074920936 |
| ## 221 graph 1 | NA | INT080 | Strength | 0.483775732 |
| ## 222 graph 1 | NA | INT086 | Strength | 0.356652107 |
| ## 223 graph 1 | NA | INT097 | Strength | 0.425150134 |

|  |  |  |  |  |  |  |  |
| --- | --- | --- | --- | --- | --- | --- | --- |
| ## | 224 | graph | 1 | NA | INT098 | Strength | 0.465034633 |
| ## | 225 | graph | 1 | NA | INT102 | Strength | 0.539966839 |
| ## | 226 | graph | 1 | NA | INT104 | Strength | 0.346720628 |
| ## | 227 | graph | 1 | NA | UNEX003 | Strength | 0.138598528 |
| ## | 228 | graph | 1 | NA | UNEX005 | Strength | 0.498257976 |
| ## | 229 | graph | 1 | NA | UNEX006 | Strength | 0.310694891 |
| ## | 230 | graph | 1 | NA | UNEX017 | Strength | 0.618207447 |
| ## | 231 | graph | 1 | NA | UNEX020 | Strength | 0.435640392 |
| ## | 232 | graph | 1 | NA | UNEX025 | Strength | 0.568443003 |
| ## | 233 | graph | 1 | NA | UNEX027 | Strength | 0.519108644 |
| ## | 234 | graph | 1 | NA | UNEX029 | Strength | 0.141552396 |
| ## | 235 | graph | 1 | NA | UNEX030 | Strength | 0.683567190 |
| ## | 236 | graph | 1 | NA | UNEX040 | Strength | 0.410705011 |
| ## | 237 | graph | 1 | NA | UNEX044 | Strength | 0.361154996 |
| ## | 238 | graph | 1 | NA | UNEX047 | Strength | 0.456768216 |
| ## | 239 | graph | 1 | NA | UNEX058 | Strength | 0.404139039 |
| ## | 240 | graph | 1 | NA | UNEX060 | Strength | 0.489620035 |
| ## | 241 | graph | 1 | NA | UNEX063 | Strength | 0.392422257 |
| ## | 242 | graph | 1 | NA | UNEX065 | Strength | 0.280738372 |
| ## | 243 | graph | 1 | NA | UNEX071 | Strength | 0.078189237 |
| ## | 244 | graph | 1 | NA | UNEX072 | Strength | 0.449594276 |
| ## | 245 | graph | 1 | NA | UNEX078 | Strength | 0.491005311 |
| ## | 246 | graph | 1 | NA | UNEX079 | Strength | 0.338315712 |
| ## | 247 | graph | 1 | NA | UNEX082 | Strength | 0.234428450 |
| ## | 248 | graph | 1 | NA | UNEX083 | Strength | 0.637412758 |
| ## | 249 | graph | 1 | NA | UNEX087 | Strength | 0.447581364 |
| ## | 250 | graph | 1 | NA | UNEX089 | Strength | 0.361418072 |
| ## | 251 | graph | 1 | NA | UNEX090 | Strength | 0.618253676 |
| ## | 252 | graph | 1 | NA | UNEX093 | Strength | 0.099344617 |
| ## | 253 | graph | 1 | NA | UNEX099 | Strength | 0.574381468 |
| ## | 254 | graph | 1 | NA | UNEX103 | Strength | 0.217343908 |
| ## | 255 | graph | 1 | NA | COG001 | Strength | 0.453301919 |
| ## | 256 | graph | 1 | NA | COG016 | Strength | 0.574419311 |
| ## | 257 | graph | 1 | NA | COG023 | Strength | 0.385430395 |
| ## | 258 | graph | 1 | NA | COG038 | Strength | 0.345310655 |
| ## | 259 | graph | 1 | NA | COG066 | Strength | 0.367287406 |
| ## | 260 | graph | 1 | NA | COG091 | Strength | 0.373486156 |
| ## | 261 | graph | 1 | NA | IMP057 | Strength | 0.094575446 |
| ## | 262 | graph | 1 | NA | COG008 | ExpectedInfluence | 0.457774887 |
| ## | 263 | graph | 1 | NA | COG021 | ExpectedInfluence | 0.321744452 |
| ## | 264 | graph | 1 | NA | COG022 | ExpectedInfluence | 0.735142221 |
| ## | 265 | graph | 1 | NA | COG028 | ExpectedInfluence | 0.787161175 |
| ## | 266 | graph | 1 | NA | COG032 | ExpectedInfluence | 0.467752723 |
| ## | 267 | graph | 1 | NA | COG036 | ExpectedInfluence | 0.429987675 |
| ## | 268 | graph | 1 | NA | COG037 | ExpectedInfluence | 0.570021949 |
| ## | 269 | graph | 1 | NA | COG043 | ExpectedInfluence | 0.537273563 |
| ## | 270 | graph | 1 | NA | COG054 | ExpectedInfluence | 0.566118523 |
| ## | 271 | graph | 1 | NA | COG055 | ExpectedInfluence | 0.336417187 |
| ## | 272 | graph | 1 | NA | COG061 | ExpectedInfluence | 0.486917155 |
| ## | 273 | graph | 1 | NA | COG062 | ExpectedInfluence | 0.630463981 |
| ## | 274 | graph | 1 | NA | COG073 | ExpectedInfluence | 0.465761125 |
| ## | 275 | graph | 1 | NA | COG075 | ExpectedInfluence | 1.000000000 |
| ## | 276 | graph | 1 | NA | COG081 | ExpectedInfluence | 0.747029597 |
| ## | 277 | graph | 1 | NA | COG085 | ExpectedInfluence | 0.283689117 |

|  |  |  |  |  |  |  |  |
| --- | --- | --- | --- | --- | --- | --- | --- |
| ## | 278 | graph | 1 | NA | COG095 | ExpectedInfluence | 0.375427099 |
| ## | 279 | graph | 1 | NA | COG101 | ExpectedInfluence | 0.451131672 |
| ## | 280 | graph | 1 | NA | IMP002 | ExpectedInfluence | 0.293911774 |
| ## | 281 | graph | 1 | NA | IMP012 | ExpectedInfluence | 0.693728450 |
| ## | 282 | graph | 1 | NA | IMP015 | ExpectedInfluence | 0.547583480 |
| ## | 283 | graph | 1 | NA | IMP019 | ExpectedInfluence | 0.210638027 |
| ## | 284 | graph | 1 | NA | IMP024 | ExpectedInfluence | 0.028477701 |
| ## | 285 | graph | 1 | NA | IMP052 | ExpectedInfluence | 0.059562341 |
| ## | 286 | graph | 1 | NA | IMP053 | ExpectedInfluence | 0.395992510 |
| ## | 287 | graph | 1 | NA | IMP056 | ExpectedInfluence | 0.379638763 |
| ## | 288 | graph | 1 | NA | IMP070 | ExpectedInfluence | 0.187354602 |
| ## | 289 | graph | 1 | NA | IMP074 | ExpectedInfluence | 0.313875125 |
| ## | 290 | graph | 1 | NA | IMP084 | ExpectedInfluence | 0.023551992 |
| ## | 291 | graph | 1 | NA | IMP088 | ExpectedInfluence | 0.048121181 |
| ## | 292 | graph | 1 | NA | IMP096 | ExpectedInfluence | 0.652833355 |
| ## | 293 | graph | 1 | NA | IMP100 | ExpectedInfluence | 0.300167955 |
| ## | 294 | graph | 1 | NA | UNEX033 | ExpectedInfluence | 0.231917405 |
| ## | 295 | graph | 1 | NA | INT048 | ExpectedInfluence | 0.407807558 |
| ## | 296 | graph | 1 | NA | IMP018 | ExpectedInfluence | -0.045926250 |
| ## | 297 | graph | 1 | NA | INT004 | ExpectedInfluence | 0.435872320 |
| ## | 298 | graph | 1 | NA | INT014 | ExpectedInfluence | 0.721280537 |
| ## | 299 | graph | 1 | NA | INT026 | ExpectedInfluence | 0.445189308 |
| ## | 300 | graph | 1 | NA | INT035 | ExpectedInfluence | 0.235718110 |
| ## | 301 | graph | 1 | NA | INT041 | ExpectedInfluence | 0.272292994 |
| ## | 302 | graph | 1 | NA | INT045 | ExpectedInfluence | 0.178671601 |
| ## | 303 | graph | 1 | NA | INT049 | ExpectedInfluence | 0.403239754 |
| ## | 304 | graph | 1 | NA | INT050 | ExpectedInfluence | 0.287609825 |
| ## | 305 | graph | 1 | NA | INT059 | ExpectedInfluence | 0.219830409 |
| ## | 306 | graph | 1 | NA | INT064 | ExpectedInfluence | 0.476572108 |
| ## | 307 | graph | 1 | NA | INT077 | ExpectedInfluence | 0.074920936 |
| ## | 308 | graph | 1 | NA | INT080 | ExpectedInfluence | 0.483775732 |
| ## | 309 | graph | 1 | NA | INT086 | ExpectedInfluence | 0.345629419 |
| ## | 310 | graph | 1 | NA | INT097 | ExpectedInfluence | 0.425150134 |
| ## | 311 | graph | 1 | NA | INT098 | ExpectedInfluence | 0.447493307 |
| ## | 312 | graph | 1 | NA | INT102 | ExpectedInfluence | 0.539966839 |
| ## | 313 | graph | 1 | NA | INT104 | ExpectedInfluence | 0.346720628 |
| ## | 314 | graph | 1 | NA | UNEX003 | ExpectedInfluence | 0.138598528 |
| ## | 315 | graph | 1 | NA | UNEX005 | ExpectedInfluence | 0.498257976 |
| ## | 316 | graph | 1 | NA | UNEX006 | ExpectedInfluence | 0.310694891 |
| ## | 317 | graph | 1 | NA | UNEX017 | ExpectedInfluence | 0.618207447 |
| ## | 318 | graph | 1 | NA | UNEX020 | ExpectedInfluence | 0.435640392 |
| ## | 319 | graph | 1 | NA | UNEX025 | ExpectedInfluence | 0.568443003 |
| ## | 320 | graph | 1 | NA | UNEX027 | ExpectedInfluence | 0.519108644 |
| ## | 321 | graph | 1 | NA | UNEX029 | ExpectedInfluence | 0.141552396 |
| ## | 322 | graph | 1 | NA | UNEX030 | ExpectedInfluence | 0.683567190 |
| ## | 323 | graph | 1 | NA | UNEX040 | ExpectedInfluence | 0.410705011 |
| ## | 324 | graph | 1 | NA | UNEX044 | ExpectedInfluence | 0.361154996 |
| ## | 325 | graph | 1 | NA | UNEX047 | ExpectedInfluence | 0.456768216 |
| ## | 326 | graph | 1 | NA | UNEX058 | ExpectedInfluence | 0.404139039 |
| ## | 327 | graph | 1 | NA | UNEX060 | ExpectedInfluence | 0.489620035 |
| ## | 328 | graph | 1 | NA | UNEX063 | ExpectedInfluence | 0.392422257 |
| ## | 329 | graph | 1 | NA | UNEX065 | ExpectedInfluence | 0.280738372 |
| ## | 330 | graph | 1 | NA | UNEX071 | ExpectedInfluence | 0.078189237 |
| ## | 331 | graph | 1 | NA | UNEX072 | ExpectedInfluence | 0.449594276 |

```
## 332 graph 1 NA UNEX078 ExpectedInfluence 0.491005311
## 333 graph 1 NA UNEX079 ExpectedInfluence 0.338315712
## 334 graph 1 NA UNEX082 ExpectedInfluence 0.234428450
## 335 graph 1 NA UNEX083 ExpectedInfluence 0.637412758
## 336 graph 1 NA UNEX087 ExpectedInfluence 0.447581364
## 337 graph 1 NA UNEX089 ExpectedInfluence 0.361418072
## 338 graph 1 NA UNEX090 ExpectedInfluence 0.618253676
## 339 graph 1 NA UNEX093 ExpectedInfluence 0.099344617
## 340 graph 1 NA UNEX099 ExpectedInfluence 0.574381468
## 341 graph 1 NA UNEX103 ExpectedInfluence 0.217343908
## 342 graph 1 NA COG001 ExpectedInfluence 0.320326692
## 343 graph 1 NA COG016 ExpectedInfluence 0.574419311
## 344 graph 1 NA COG023 ExpectedInfluence 0.385430395
## 345 graph 1 NA COG038 ExpectedInfluence 0.341377988
## 346 graph 1 NA COG066 ExpectedInfluence 0.367287406
## 347 graph 1 NA COG091 ExpectedInfluence 0.332265848
## 348 graph 1 NA IMP057 ExpectedInfluence -0.094575446
```

```
centralityPlot(A_OLIFE_17, include = c("Strength", "Closeness", "Betweenness",
                                       "ExpectedInfluence"),
              scale = "relative", orderBy = "ExpectedInfluence")
```

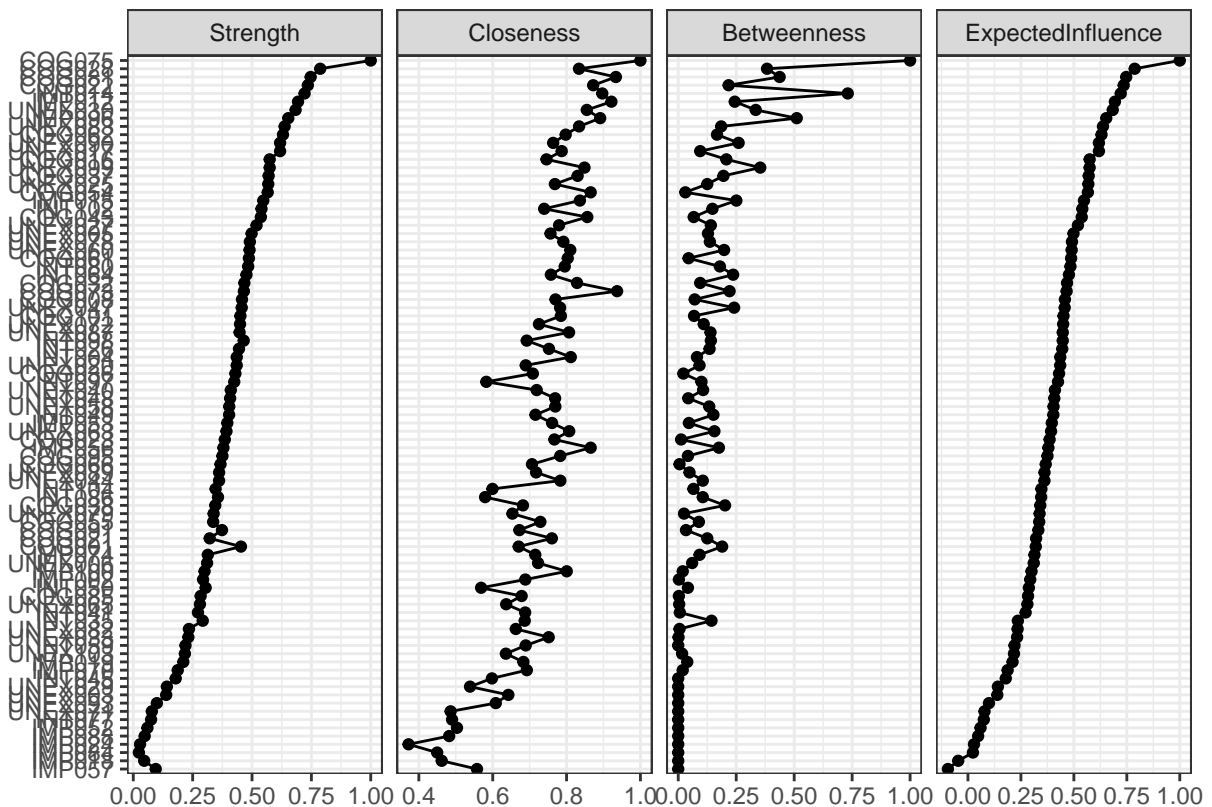

```
A_OLIFE_17 <- qgraph(graph_OLIFE_17, groups = dim.OLIFE_17$dimension,
                    label.prop = 0.6 ,
                    color = c("#66C2A5", "#fcdbf", "#8DA0CB", "#f1605d"),
```

```
layout = "spring",
vsize = centralityTableMSS[1:87,5]*3+3, # BETWEENNESS
labels = 1:87, legend.cex = 0.3,
title = "EGA model of the O-LIFE - new 4 factors")
```

#### EGA model of the O-LIFE – new 4 factors

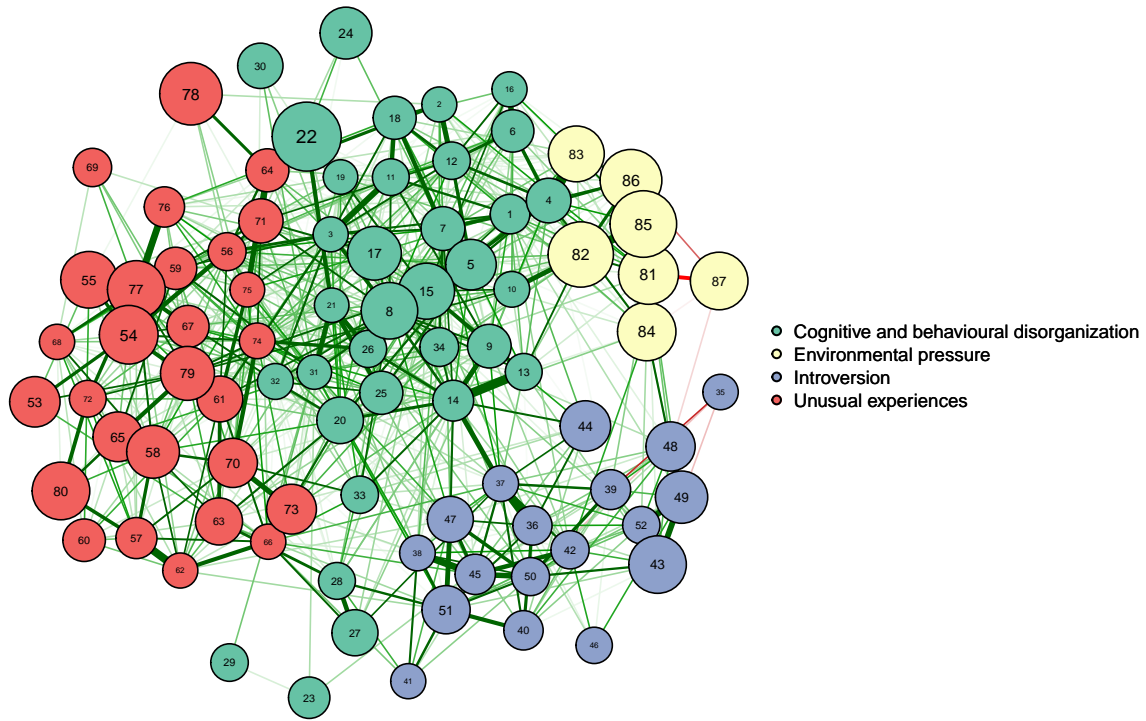

```
# pdf("EGA_OLIFE_new4dim.pdf", width = 12)
# par(mfrow = c(1,1))
# dev.off()
```

```
# Confirmatory Factor Analysis (CFA) FOR EGA MODEL, THE EFA MODEL and
# ORIGINAL MODEL
```

```
# Original model of the O-LIFE (Mason and Claridge, 2006)
```

```
model_OLIFE_original <- '
Factor1 =~ `COG001` + `COG008` + `COG016` + `COG021` + `COG022` + `COG023` +
           `COG028` + `COG032` + `COG036` + `COG037` + `COG038` + `COG043` +
           `COG054` + `COG055` + `COG061` + `COG062` + `COG066` + `COG073` +
           `COG075` + `COG081` + `COG085` + `COG091` + `COG095` + `COG101`
Factor2 =~ `IMP007` + `IMP010` + `IMP012` + `IMP013` + `IMP015` + `IMP018` +
           `IMP019` + `IMP024` + `IMP031` + `IMP051` + `IMP052` + `IMP053` +
           `IMP056` + `IMP057` + `IMP068` + `IMP069` + `IMP070` + `IMP074` +
           `IMP084` + `IMP088` + `IMP096` + `IMP100`
Factor3 =~ `UNEX003` + `UNEX005` + `UNEX006` + `UNEX017` + `UNEX020` +
           `UNEX025` + `UNEX027` + `UNEX029` + `UNEX030` + `UNEX033` +
           `UNEX040` + `UNEX044` + `UNEX047` + `UNEX058` + `UNEX060` +
```

```
        `UNEX063` + `UNEX065` + `UNEX071` + `UNEX072` + `UNEX076` +
        `UNEX078` + `UNEX079` + `UNEX082` + `UNEX083` + `UNEX087` +
        `UNEX089` + `UNEX090` + `UNEX093` + `UNEX099` + `UNEX103`
Factor4 =~ `INT004` + `INT009` + `INT011` + `INT014` + `INT026` + `INT034` +
        `INT035` + `INT039` + `INT041` + `INT042` + `INT045` + `INT046` +
        `INT048` + `INT049` + `INT050` + `INT059` + `INT064` + `INT067` +
        `INT077` + `INT080` + `INT086` + `INT092` + `INT094` + `INT097` +
        `INT098` + `INT102` + `INT104`
,

# EGA model with 87 items and 4 dimensions
model_OLIFE_ega <- '
Factor1 =~ `COG008` + `COG021` + `COG022` + `COG028` + `COG032` + `COG036` +
        `COG037` + `COG043` + `COG054` + `COG055` + `COG061` + `COG062` +
        `COG073` + `COG075` + `COG081` + `COG085` + `COG095` + `COG101` +
        `IMP002` + `IMP012` + `IMP015` + `IMP019` + `IMP024` + `IMP052` +
        `IMP053` + `IMP056` + `IMP070` + `IMP074` + `IMP084` + `IMP088` +
        `IMP096` + `IMP100` + `UNEX033` + `INT048`
Factor2 =~ `IMP018` + `INT004` + `INT014` + `INT026` + `INT035` + `INT041` +
        `INT045` + `INT049` + `INT050` + `INT059` + `INT064` + `INT077` +
        `INT080` + `INT086` + `INT097` + `INT098` + `INT102` + `INT104`
Factor3 =~ `UNEX003` + `UNEX005` + `UNEX006` + `UNEX017` + `UNEX020` +
        `UNEX025` + `UNEX027` + `UNEX029` + `UNEX030` + `UNEX040` +
        `UNEX044` + `UNEX047` + `UNEX058` + `UNEX060` + `UNEX063` +
        `UNEX065` + `UNEX071` + `UNEX072` + `UNEX078` + `UNEX079` +
        `UNEX082` + `UNEX083` + `UNEX087` + `UNEX089` + `UNEX090` +
        `UNEX093` + `UNEX099` + `UNEX103`
Factor4 =~ `COG001` + `COG016` + `COG023` + `COG038` + `COG066` + `COG091` +
        `IMP057`
,
```

*# These are not computed in the Markdown because they would take too much time.*

```
# fit.OLIFE.original <- cfa(model_OLIFE_original, data = OLIFE,
#                           estimator= "wlsmv", meanstructure = TRUE,
#                           ordered = TRUE)
#
# fit.OLIFE.efa <- cfa(model_OLIFE_efa, data = OLIFE, estimator = "wlsmv",
#                      meanstructure = TRUE, ordered = TRUE)
#
# fit.OLIFE.ega <- CFA(ega_OLIFE_17, data = OLIFE_17, estimator = "WLSMW",
#                     meanstructure = TRUE, ordered=TRUE)
#

# Results

# ORIGINAL
fitMeasures(fit.OLIFE.original, c("chisq","df","pvalue","pvalue.scaled","srmr",
    "cfi","cfi.scaled", "rmsea",
    "rmsea.scaled", "tli", "tli.scaled"))
```

| ## | chisq | df | pvalue | pvalue.scaled | srmr |
| --- | --- | --- | --- | --- | --- |
| ## | 11486.318 | 5144.000 | 0.000 | 0.000 | 0.098 |

```
##          cfi    cfi.scaled      rmsea  rmsea.scaled      tli
##          0.934          0.854      0.034      0.022      0.933
##    tli.scaled
##          0.851
```

###### # EXPLORATORY FACTOR ANALYSIS

```
fitMeasures(fit.OLIFE.efa, c("chisq","df","pvalue","pvalue.scaled","srmr","cfi",
                             "cfi.scaled", "rmsea",
                             "rmsea.scaled", "tli", "tli.scaled"))
```

```
##          chisq          df      pvalue  pvalue.scaled      srmr
##    11543.169    5246.000      0.000      0.000      0.097
##          cfi    cfi.scaled      rmsea  rmsea.scaled      tli
##          0.936          0.858      0.034      0.021      0.935
##    tli.scaled
##          0.855
```

###### # EXPLORATORY GRAPH ANALYSIS

```
fitMeasures(fit.OLIFE.ega$fit, c("chisq","df","pvalue","pvalue.scaled","srmr",
                                   "cfi","cfi.scaled", "rmsea",
                                   "rmsea.scaled", "tli", "tli.scaled"))
```

```
##          chisq          df      pvalue  pvalue.scaled      srmr
##    6789.411    3648.000      0.000      0.000      0.089
##          cfi    cfi.scaled      rmsea  rmsea.scaled      tli
##          0.965          0.903      0.029      0.021      0.965
##    tli.scaled
##          0.900
```

```
lavTestLRT(fit.OLIFE.original, fit.OLIFE.ega$fit, method="satorra.bentler.2010")
```

```
##
## Scaled Chi-Squared Difference Test (method = "satorra.bentler.2010")
##
## lavaan NOTE:
##   The "Chisq" column contains standard test statistics, not the
##   robust test that should be reported per model. A robust difference
##   test is a function of two standard (not robust) statistics.
##
##           Df AIC BIC   Chisq Chisq diff Df diff Pr(>Chisq)
## fit.OLIFE.ega$fit 3648      6789.4
## fit.OLIFE.original 5144      11486.3      2637.6      1496 < 2.2e-16 ***
## ---
## Signif. codes:  0 '***' 0.001 '**' 0.01 '*' 0.05 '.' 0.1 ' ' 1
```

```
lavTestLRT(fit.OLIFE.efa, fit.OLIFE.ega$fit, method="satorra.bentler.2010")
```

```
##
## Scaled Chi-Squared Difference Test (method = "satorra.bentler.2010")
##
## lavaan NOTE:
```

```
## The "Chisq" column contains standard test statistics, not the
## robust test that should be reported per model. A robust difference
## test is a function of two standard (not robust) statistics.
##
##           Df AIC BIC   Chisq Chisq diff Df diff Pr(>Chisq)
## fit.OLIFE.ega$fit 3648      6789.4
## fit.OLIFE.efa    5246    11543.2    2454.9    1598 < 2.2e-16 ***
## ---
## Signif. codes:  0 '***' 0.001 '**' 0.01 '*' 0.05 '.' 0.1 ' ' 1
```

```
# McDonald's Omega and Chronbach Alpha
# The first might provide a better estimate of reliability for factors with
# complex structures or fewer items.
```

```
OLIFE_disregulation <- OLIFE_17[, c("COG008", "COG021", "COG022", "COG028",
                                   "COG032", "COG036", "COG037", "COG043",
                                   "COG054", "COG055", "COG061", "COG062",
                                   "COG073", "COG075", "COG081", "COG085",
                                   "COG095", "COG101", "IMP002", "IMP012",
                                   "IMP015", "IMP019", "IMP024", "IMP052", "IMP053",
                                   "IMP056", "IMP070", "IMP074", "IMP084", "IMP088",
                                   "IMP096", "IMP100", "UNEX033", "INT048")]
OLIFE_introversion <- OLIFE_17[, c("IMP018", "INT004", "INT014", "INT026",
                                   "INT035", "INT041", "INT045", "INT049",
                                   "INT050", "INT059", "INT064", "INT077",
                                   "INT080", "INT086", "INT097", "INT098",
                                   "INT102", "INT104")]
OLIFE_unusual_exp <- OLIFE_17[, c("UNEX003", "UNEX005", "UNEX006", "UNEX017",
                                   "UNEX020", "UNEX025", "UNEX027", "UNEX029",
                                   "UNEX030", "UNEX040", "UNEX044", "UNEX047",
                                   "UNEX058", "UNEX060", "UNEX063", "UNEX065",
                                   "UNEX071", "UNEX072", "UNEX078", "UNEX079",
                                   "UNEX082", "UNEX083", "UNEX087", "UNEX089",
                                   "UNEX090", "UNEX093", "UNEX099", "UNEX103")]
OLIFE_environmental_pressure <- OLIFE_17[, c("COG001", "COG016", "COG023",
                                              "COG038", "COG066", "COG091",
                                              "IMP057")]

omega_disregulation <- omega(OLIFE_disregulation, plot = FALSE)
print(omega_disregulation)
```

```
## Omega
## Call: omegah(m = m, nfactors = nfactors, fm = fm, key = key, flip = flip,
## digits = digits, title = title, sl = sl, labels = labels,
## plot = plot, n.obs = n.obs, rotate = rotate, Phi = Phi, option = option,
## covar = covar)
## Alpha:          0.88
## G.6:            0.89
## Omega Hierarchical: 0.66
## Omega H asymptotic: 0.75
## Omega Total      0.89
##
## Schmid Leiman Factor loadings greater than 0.2
```

```
##          g    F1*   F2*   F3*   h2   u2   p2
## COG008   0.40  0.26                0.23 0.77 0.68
## COG021   0.29  0.35                0.21 0.79 0.40
## COG022   0.46  0.35                0.37 0.63 0.58
## COG028   0.48  0.27                0.34 0.66 0.68
## COG032   0.45                0.27 0.73 0.74
## COG036   0.39  0.27                0.25 0.75 0.62
## COG037   0.44  0.27                0.30 0.70 0.64
## COG043   0.45  0.21                0.27 0.73 0.75
## COG054   0.49                0.25 0.31 0.69 0.76
## COG055   0.34                0.17 0.83 0.67
## COG061   0.41  0.33                0.29 0.71 0.60
## COG062   0.43  0.45                0.40 0.60 0.47
## COG073   0.49                0.35 0.36 0.64 0.65
## COG075   0.58                0.40 0.50 0.50 0.67
## COG081   0.51                0.22 0.33 0.67 0.77
## COG085   0.33  0.25                0.18 0.82 0.63
## COG095   0.36                0.18 0.82 0.73
## COG101   0.39  0.41                0.33 0.67 0.46
## IMP002   0.35                0.18 0.82 0.68
## IMP012   0.45                0.20 0.28 0.72 0.73
## IMP015   0.39                0.35 0.29 0.71 0.51
## IMP019   0.28                0.34 0.20 0.80 0.39
## IMP024                0.20 0.08 0.92 0.47
## IMP052                0.26 0.10 0.90 0.15
## IMP053   0.37                0.27 0.23 0.77 0.61
## IMP056   0.37                0.21 0.19 0.81 0.72
## IMP070   0.25                0.11 0.89 0.55
## IMP074   0.30                0.14 0.86 0.66
## IMP084                0.32 0.12 0.88 0.14
## IMP088                0.28 0.11 0.89 0.20
## IMP096   0.41                0.33 0.29 0.71 0.58
## IMP100   0.35                0.22 0.19 0.81 0.65
## UNEX033   0.31                0.14 0.86 0.69
## INT048   0.36                0.18 0.82 0.75
##
## With Sums of squares of:
##   g  F1*  F2*  F3*
## 4.96 1.31 0.81 1.04
##
## general/max 3.78  max/min = 1.62
## mean percent general = 0.59  with sd = 0.17 and cv of 0.29
## Explained Common Variance of the general factor = 0.61
##
## The degrees of freedom are 462  and the fit is 1.04
## The number of observations was 1059  with Chi Square = 1081.72  with prob < 1.2e-51
## The root mean square of the residuals is 0.03
## The df corrected root mean square of the residuals is 0.04
## RMSEA index = 0.036  and the 10 % confidence intervals are 0.033 0.038
## BIC = -2136.14
##
## Compare this with the adequacy of just a general factor and no group factors
## The degrees of freedom for just the general factor are 527  and the fit is 1.75
## The number of observations was 1059  with Chi Square = 1824.54  with prob < 2.3e-142
```

```
## The root mean square of the residuals is 0.07
## The df corrected root mean square of the residuals is 0.07
##
## RMSEA index = 0.048 and the 10 % confidence intervals are 0.046 0.051
## BIC = -1846.06
##
## Measures of factor score adequacy
##
## Correlation of scores with factors      g   F1*   F2*   F3*
## Multiple R square of scores with factors 0.69 0.46 0.30 0.49
## Minimum correlation of factor score estimates 0.38 -0.08 -0.39 -0.02
##
## Total, General and Subset omega for each subset
##
## Omega total for total scores and subscales      g   F1*   F2*   F3*
## Omega general for total scores and subscales 0.66 0.53 0.57 0.33
## Omega group for total scores and subscales 0.12 0.27 0.17 0.30

omega_introversion <- omega(OLIFE_introversion, plot = FALSE)
print(omega_introversion)

## Omega
## Call: omegah(m = m, nfactors = nfactors, fm = fm, key = key, flip = flip,
## digits = digits, title = title, sl = sl, labels = labels,
## plot = plot, n.obs = n.obs, rotate = rotate, Phi = Phi, option = option,
## covar = covar)
## Alpha:      0.82
## G.6:      0.82
## Omega Hierarchical: 0.61
## Omega H asymptotic: 0.73
## Omega Total 0.83
##
## Schmid Leiman Factor loadings greater than 0.2
##      g   F1*   F2*   F3*   h2   u2   p2
## IMP018- 0.20      0.09 0.91 0.41
## INT004 0.51      0.35 0.38 0.62 0.68
## INT014 0.54      0.29 0.39 0.61 0.75
## INT026 0.39 0.38      0.30 0.70 0.51
## INT035 0.36      0.23      0.21 0.79 0.64
## INT041 0.42      0.31 0.28 0.72 0.64
## INT045 0.30 0.22      0.15 0.85 0.62
## INT049 0.41 0.26      0.26 0.74 0.66
## INT050 0.35      0.37      0.27 0.73 0.46
## INT059 0.34      0.26 0.18 0.82 0.63
## INT064 0.41 0.52      0.45 0.55 0.39
## INT077 0.24      0.20      0.10 0.90 0.55
## INT080 0.46 0.27      0.32 0.68 0.64
## INT086 0.37      0.34      0.26 0.74 0.52
## INT097 0.34      0.62      0.50 0.50 0.23
## INT098 0.41 0.41      0.35 0.65 0.47
## INT102 0.47      0.22 0.29 0.71 0.75
## INT104 0.38      0.36      0.29 0.71 0.51
##
## With Sums of squares of:
```

```
##      g  F1*  F2*  F3*
## 2.77 0.87 0.94 0.49
##
## general/max 2.95    max/min =    1.91
## mean percent general = 0.56    with sd = 0.13 and cv of 0.24
## Explained Common Variance of the general factor = 0.55
##
## The degrees of freedom are 102 and the fit is 0.21
## The number of observations was 1059 with Chi Square = 224.99 with prob < 2.9e-11
## The root mean square of the residuals is 0.03
## The df corrected root mean square of the residuals is 0.03
## RMSEA index = 0.034 and the 10 % confidence intervals are 0.028 0.04
## BIC = -485.45
##
## Compare this with the adequacy of just a general factor and no group factors
## The degrees of freedom for just the general factor are 135 and the fit is 0.73
## The number of observations was 1059 with Chi Square = 761.7 with prob < 4.5e-88
## The root mean square of the residuals is 0.08
## The df corrected root mean square of the residuals is 0.08
##
## RMSEA index = 0.066 and the 10 % confidence intervals are 0.062 0.071
## BIC = -178.59
##
## Measures of factor score adequacy
##
##              g  F1*  F2*  F3*
## Correlation of scores with factors      0.80 0.67 0.73 0.50
## Multiple R square of scores with factors 0.63 0.45 0.53 0.25
## Minimum correlation of factor score estimates 0.27 -0.11 0.05 -0.50
##
## Total, General and Subset omega for each subset
##
##              g  F1*  F2*  F3*
## Omega total for total scores and subscales 0.83 0.69 0.66 0.67
## Omega general for total scores and subscales 0.61 0.40 0.32 0.49
## Omega group for total scores and subscales 0.15 0.30 0.34 0.19

omega_unusual_exp <- omega(OLIFE_unusual_exp, plot = FALSE)
print(omega_unusual_exp)

## Omega
## Call: omegah(m = m, nfactors = nfactors, fm = fm, key = key, flip = flip,
##      digits = digits, title = title, sl = sl, labels = labels,
##      plot = plot, n.obs = n.obs, rotate = rotate, Phi = Phi, option = option,
##      covar = covar)
## Alpha:          0.86
## G.6:            0.87
## Omega Hierarchical: 0.62
## Omega H asymptotic: 0.71
## Omega Total      0.87
##
## Schmid Leiman Factor loadings greater than 0.2
##              g  F1*  F2*  F3*  h2  u2  p2
## UNEX003 0.22          0.27 0.13 0.87 0.38
## UNEX005 0.43 0.21      0.24 0.30 0.70 0.64
## UNEX006 0.33          0.37 0.26 0.74 0.42
```

```
## UNEX017 0.46 0.32 0.32 0.68 0.67
## UNEX020 0.34 0.49 0.37 0.63 0.31
## UNEX025 0.42 0.36 0.32 0.68 0.55
## UNEX027 0.40 0.23 0.25 0.75 0.63
## UNEX029 0.27 0.22 0.14 0.86 0.52
## UNEX030 0.46 0.21 0.30 0.70 0.71
## UNEX040 0.34 0.48 0.37 0.63 0.32
## UNEX044 0.33 0.25 0.18 0.82 0.60
## UNEX047 0.38 0.33 0.28 0.72 0.52
## UNEX058 0.38 0.25 0.23 0.26 0.74 0.55
## UNEX060 0.34 0.29 0.20 0.80 0.56
## UNEX063 0.39 0.21 0.79 0.73
## UNEX065 0.37 0.20 0.80 0.69
## UNEX071 0.24 0.08 0.92 0.72
## UNEX072 0.40 0.20 0.22 -0.21 0.29 0.71 0.54
## UNEX078 0.43 0.31 0.28 0.72 0.65
## UNEX079 0.35 0.20 0.20 0.80 0.59
## UNEX082 0.29 0.22 -0.30 0.23 0.77 0.37
## UNEX083 0.45 0.29 0.30 0.70 0.67
## UNEX087 0.40 0.23 0.22 0.78 0.73
## UNEX089 0.38 0.25 0.20 0.80 0.70
## UNEX090 0.44 0.20 0.20 0.27 0.73 0.71
## UNEX093 0.24 0.14 0.86 0.43
## UNEX099 0.43 0.20 0.29 0.71 0.66
## UNEX103 0.26 0.26 0.21 0.18 0.82 0.37
##
## With Sums of squares of:
## g F1* F2* F3*
## 3.82 0.93 1.26 0.74
##
## general/max 3.02 max/min = 1.7
## mean percent general = 0.57 with sd = 0.13 and cv of 0.24
## Explained Common Variance of the general factor = 0.57
##
## The degrees of freedom are 297 and the fit is 0.85
## The number of observations was 1059 with Chi Square = 892.73 with prob < 6.7e-61
## The root mean square of the residuals is 0.04
## The df corrected root mean square of the residuals is 0.04
## RMSEA index = 0.044 and the 10 % confidence intervals are 0.04 0.047
## BIC = -1175.9
##
## Compare this with the adequacy of just a general factor and no group factors
## The degrees of freedom for just the general factor are 350 and the fit is 1.45
## The number of observations was 1059 with Chi Square = 1523.46 with prob < 8.4e-146
## The root mean square of the residuals is 0.07
## The df corrected root mean square of the residuals is 0.08
##
## RMSEA index = 0.056 and the 10 % confidence intervals are 0.053 0.059
## BIC = -914.32
##
## Measures of factor score adequacy
##
## Correlation of scores with factors g F1* F2* F3*
## Multiple R square of scores with factors 0.80 0.53 0.70 0.67
## 0.64 0.29 0.49 0.46
```

```
## Minimum correlation of factor score estimates 0.28 -0.43 -0.02 -0.09
##
## Total, General and Subset omega for each subset
##
##           g  F1*  F2*  F3*
## Omega total for total scores and subscales    0.87 0.71 0.73 0.52
## Omega general for total scores and subscales  0.62 0.51 0.43 0.41
## Omega group for total scores and subscales    0.11 0.20 0.30 0.11

omega_environmental_pressure <- omega(OLIFE_environmental_pressure,
                                     plot = FALSE)
print(omega_environmental_pressure)

## Omega
## Call: omegah(m = m, nfactors = nfactors, fm = fm, key = key, flip = flip,
##   digits = digits, title = title, sl = sl, labels = labels,
##   plot = plot, n.obs = n.obs, rotate = rotate, Phi = Phi, option = option,
##   covar = covar)
## Alpha:          0.7
## G.6:            0.67
## Omega Hierarchical: 0.61
## Omega H asymptotic: 0.81
## Omega Total      0.75
##
## Schmid Leiman Factor loadings greater than 0.2
##           g  F1*  F2*  F3*  h2  u2  p2
## COG001  0.48          0.76      0.81 0.19 0.29
## COG016  0.54 0.55          0.60 0.40 0.49
## COG023  0.45          0.24 0.76 0.85
## COG038  0.41          0.22 0.78 0.78
## COG066  0.54 0.25          0.36 0.64 0.79
## COG091  0.53          0.23 0.34 0.66 0.84
## IMP057- 0.31          0.15 0.85 0.63
##
## With Sums of squares of:
##   g  F1*  F2*  F3*
## 1.57 0.38 0.63 0.14
##
## general/max 2.48  max/min = 4.41
## mean percent general = 0.67  with sd = 0.21 and cv of 0.32
## Explained Common Variance of the general factor = 0.58
##
## The degrees of freedom are 3  and the fit is 0
## The number of observations was 1059  with Chi Square = 1.38  with prob < 0.71
## The root mean square of the residuals is 0.01
## The df corrected root mean square of the residuals is 0.01
## RMSEA index = 0  and the 10 % confidence intervals are 0 0.038
## BIC = -19.52
##
## Compare this with the adequacy of just a general factor and no group factors
## The degrees of freedom for just the general factor are 14  and the fit is 0.08
## The number of observations was 1059  with Chi Square = 88.44  with prob < 7.5e-13
## The root mean square of the residuals is 0.06
## The df corrected root mean square of the residuals is 0.07
##
```

```
## RMSEA index = 0.071 and the 10 % confidence intervals are 0.057 0.085
## BIC = -9.07
##
## Measures of factor score adequacy
##
##           g    F1*  F2*  F3*
## Correlation of scores with factors    0.79  0.61 0.82  0.33
## Multiple R square of scores with factors    0.63  0.38 0.67  0.11
## Minimum correlation of factor score estimates 0.26 -0.25 0.33 -0.79
##
## Total, General and Subset omega for each subset
##
##           g    F1*  F2*  F3*
## Omega total for total scores and subscales    0.75 0.63 0.58 0.44
## Omega general for total scores and subscales    0.61 0.41 0.32 0.38
## Omega group for total scores and subscales    0.11 0.22 0.26 0.07
```
